# COVID-19 among migrants, refugees, and internally displaced persons: systematic review, meta-analysis and qualitative synthesis of the global empirical literature

**DOI:** 10.1101/2023.08.03.23293586

**Authors:** Maren Hintermeier, Nora Gottlieb, Sven Rohleder, Jan Oppenberg, Mazen Baroudi, Sweetmavourneen Pernitez-Agan, Janice Lopez, Sergio Flores, Amir Mohsenpour, Kolitha Wickramage, Kayvan Bozorgmehr

**Affiliations:** Section Health Equity Studies & Migration, Department of General Practice and Health Services Research, University Hospital Heidelberg, Im Neuenheimer Feld 130.3, 69120 Heidelberg, Germany; Department of Population Medicine and Health Services Research, School of Public Health, Bielefeld University, Universitätsstraße 25, 33615 Bielefeld, Bielefeld, Germany; Department of Epidemiology and Global Health, Umeå University, Sweden; Migration Health Division, International Organization for Migration, Manila, Philippines; Department of Public Health and Caring Sciences, Uppsala University, Sweden; UN Migration Agency Global Data Institute, Migration Health Division, International Organization for Migration, Berlin, Germany

**Keywords:** COVID-19, refugees, asylum seekers, IDP, migration, health inequality

## Abstract

**Background:** Pandemic response and preparedness plans aim at mitigating the spread of infectious diseases and protecting public health, but migrants are often side-lined. Evidence amounted early that migrants are disproportionately affected by the COVID-19 pandemic and its consequences. However, synthesised evidence is lacking that quantifies the inequalities in infection risk and disease outcomes, or contextualises the consequences of pandemic measures and their underlying mechanisms.

**Methods:** Systematic review searching 25 databases and grey literature (12/2019 to 11/2021). We considered empirical articles covering migrants, refugees, asylum-seekers, and internally displaced persons reporting SARS-CoV-2 cases, hospitalisation, ICU admission, mortality, COVID-19 vaccination rates or health consequences of pandemic measures. Random-effects meta-analysis of observational studies and qualitative analysis were performed for evidence synthesis. A Protocol was registered with PROSPERO (CRD42021296952).

**Findings:** Out of 6956 studies, we included 241 in the review. For the quantitative studies (n=46), meta-analysis with over 40 million study participants showed that compared to non-migrants, migrants have an elevated risk of infection (RR = 2·33; 95%-CI: 1·88-2·89) but similar risk for hospitalisation (RR = 1·05; 0·80-1·37), while the likelihood of ICU admission was higher (RR = 1·36; 1·04-1·78). Among those hospitalised, migrants had a lower risk of mortality (RR = 0·47; 0·30-0·73), while their population-based excess mortality tended to be higher (RR = 1·31; 0·95-1·80). The qualitative synthesis (n=44) highlighted the complex interplay of social and COVID-19-related factors at different levels. This involved increased exposure, risk, and impact of pandemic measures that compromised the health of migrants.

**Interpretation:** Even in the advanced stages of the pandemic, migrants faced higher infection risks and disproportionately suffered from the consequences of COVID-19 disease, including deaths. Population-level interventions in future health emergencies must better consider socio-economic, structural and community-level exposures to mitigate risks among migrants and enhance health information systems, to close coverage gaps in migrant groups.

**Funding:** None.

## INTRODUCTION

Global health emergencies like the COVID-19 pandemic occur unpredictably, and effective responses require well elaborated and actionable preparedness strategies in line with the International Health Regulations.^1^ National pandemic preparedness and response plans are part of such strategies and are supported by the WHO, e.g., through a strategic plan published on February 4, 2020, with subsequent ongoing elaboration.^2, 3^ However, migrants have been side-lined in such plans, prompting a call for urgent global action to consider migrants in pandemic responses from the Lancet Migration early in the pandemic (April 10,2020).^4^

### Panel 1: Research in context

#### Evidence before this study

Reviews on inequalities in COVID-19-related health risks published in the early phase of the pandemic found that migrants were disproportionally affected by the pandemic. High infection risk and all-cause mortality, as well as severe mental burdens among migrants, were identified. However, the *magnitude* of inequalities in the risk of infection, hospitalisation, admission to intensive care units (ICU), mortality, or vaccination coverage between migrants and non-migrants has not been quantified. Studies conducted during the early phase of the pandemic found that poor working conditions, crowded housing, language barriers, and legal aspects are among the social determinants that intersect with migration, resulting in increased COVID-19-related risks. Evidence summaries published since then (covering studies until October 2021) focused on individual health effects of the pandemic, e.g., mental health, risk of infection, or severity of SARS-CoV-2 infections among migrants, and were limited in their geographic scope (e.g., focus on single countries, or high-income countries), the outcomes considered, or covered only single or specific migrant groups.

#### Added value of this study

We mapped the global empirical literature and synthesised the available qualitative *and* quantitative literature (published in English, German and Spanish) on multiple COVID-19-related outcomes among diverse categories of migrants, as well as the impacts of the COVID-19 pandemic on migrant populations *worldwide* by the end of 2021. We quantified absolute and relative inequalities among over 40 million study participants (including migrant and resident populations) using random-effects meta-analyses for several outcomes such as risk of SARS-CoV-2 infection, hospitalisation, ICU admission and mortality. Our results showed higher infection risk among migrants compared to non-migrants, and hinted at different patterns by geographical or contextual exposures. As opposed to previous (narrative) systematic reviews, we found no evidence for increased hospitalisation risk, but higher risk for ICU admission and death as far as population-level estimates are considered.

The comparability of the data was hampered by the heterogeneity of studies and poor reporting quality, while the lack of disaggregated reporting for some outcomes (e.g., vaccination coverage) made it impossible to synthesise evidence on important dimensions of inequality between migrant and resident populations. Only seven studies (2·9% of all studies) addressed vaccination coverage. Three of those studies were conducted in the European context and reported lower vaccination rates among migrants (i.e., foreign-born individuals) compared with the non-foreign-born population. The opposite was found in a study from China. Two modelling studies recommended including migrants in vaccination strategies to prevent cases and increase cost-effectiveness, while one study focused on the effectiveness of a COVID-19 vaccine (rather than vaccination coverage). Findings from the qualitative synthesis uncover not only the ways in which interrelated social risks and inequalities (lack of social protection, working conditions, housing, legal uncertainties) engender severe and unique impacts on migrants; but they also pinpoint potential sources of resilience at individual, community and societal level (e.g., mindset, financial resources, mutual support, access to health information, trust in authorities, state assistance).

#### Implications of all available evidence

Not only in the pandemic’s early phases, but even by the end of 2021, migrants were at higher risk of SARS-CoV- 2 infection. Once infected, migrants seemed to have more severe courses of disease requiring admission to ICU. Deaths among clinical populations were lower (likely due to different age-structures), but population-based excess mortality was higher in migrants. The venue for reducing such inequalities appears to be through population-level rather than clinical interventions. Socio-economic structures, risks in communities and specific contexts (worksites, accommodation centres) appeared as major drivers for higher infection risk and are likely to be part of the underlying causes of higher mortality. To address the challenges posed by the impact of COVID-19 on migrant health, and to better prepare for future health emergencies, it is urgently required to improve health information systems and ensure the inclusion of migrant populations in national pandemic response plans. Social and health equity policies and measures are needed to avoid future pandemic measures from unfolding (unintended) negative consequences for migrants.

Evidence from the early phase of the pandemic suggests that migrants were disproportionally affected by the COVID-19 pandemic. Reviews of the early literature found an increased risk of SARS-CoV-2 infection in migrant populations and elevated all-cause mortality compared to non-migrants.^5, 6^ Migrants living in crowded housing conditions, but also undocumented migrants and migrant healthcare workers, were found to be at higher infection risk.^5, 6^ A study examining outbreaks in German accommodation centres for asylum seekers and refugees found a significantly higher attack rate if indiscriminate mass-quarantine was applied to all camp inhabitants compared with targeted contact tracing.^7^ Several studies identified risk factors such as precarious working conditions, crowded housing, language barriers, or legal restrictions among migrants and negative effects of pandemic control measures on mental health.^5, 6, 8–10^

Despite these early efforts to compile evidence on differential risks and exposures between migrant and resident populations, there is a lack of synthesised evidence quantifying the *magnitude* of inequalities in infection risk, consequences of disease, or vaccination rates. Furthermore, there is still a dearth of consolidated knowledge on the impact of pandemic response strategies on migrant health beyond studies published at the onset of the pandemic.

We conducted a systematic review covering the literature from 12/2019 to 11/2021 to map the global landscape of the empirical literature and synthesise the evidence in this field. We investigated the risk of SARS-CoV-2 infections and the consequences of disease (measured by hospitalisation, intensive care unit (ICU) admission, and mortality rates) among asylum seekers, refugees, migrants and internally displaced persons (IDP) compared to non-migrants; vaccination coverage among migrants and non-migrants; and the impact of lockdown and pandemic control measures on migrant health.

## METHODOLOGY

### Search strategy and selection criteria

We conducted a systematic literature review in line with the Preferred Reporting for Systematic Reviews and Meta-Analyses (PRISMA) guideline. Studies were eligible for inclusion if migrants, IDP, refugees or asylum seekers were studied (following definitions of the International Organisation for Migration (IOM)^11^ and the United Nations High Commissioner (UNHCR)^12^); health effects of SARS-CoV-2 (cases, hospitalisation, ICU admission, mortality, vaccination) or corresponding policy measures among refugees and migrants were assessed; and if written in English, German or Spanish. We performed a search of the Cochrane Library and the WHO COVID-19 Research Database (representing a comprehensive source of 24 bibliographic databases), thereby updating and complementing the search of a previously published review covering all studies from 12/2019 to 11/2021.^6^ All study designs were considered except case series, theoretical research papers, or policy analyses without empirical data. We included peer-reviewed articles, but also preprints and official reports from the IOM and European Public Health Association websites, as well as studies from previous reviews known to the authors. For systematic reviews, only the primary studies were considered. The review protocol was registered with PROSPERO (CRD42021296952).

Search terms were developed, refined and validated among the review team, and databases searched by experienced reviewers (SR, MH) of the Rapid Response Review Unit (RRRUN).^13^ Website searches were performed by three reviewers (JL, SA, JO). Identified records were uploaded to a management tool for systematic reviews (Covidence), where two reviewers each screened title and abstracts independently. The same applied for full-texts. Discrepancies were resolved by a third reviewer (KB) and decisions were consented in constant discussion among the review team.

The volume of included articles required adjustments to the protocol regarding data extraction and quality assessment. Data extraction was done individually and cross-checked by MH. The following items were extracted: Generic bibliographic information (author, title, year published, journal); study objectives, hypothesis and research questions; study characteristics (research method, sample size, geography); population and context characteristics; findings (e.g., main outcomes in quantitative studies, major themes/minor themes in qualitative studies) and conclusions as reported.

The Joanna Briggs Institute’s critical appraisal tools were used for the quality assessment, which was performed by pairs of two reviewers independently.^14^ A quality score (range 0-100%, from lowest to highest quality) was constructed based on the answers of applied checklists (questions indicated as “not applicable” were not counted into the overall quality score in order to avoid artificial downrating of studies). For each study, the scores obtained from two independent ratings were averaged, and studies were grouped based on their scores and classified as high (100-75% of possible score), moderate (74-50%), and low (<50%) quality studies (Supplementary File, Table S5a-f). Rating discrepancies in case of considerably different scores were resolved by discussion among the team. The quality of modelling studies was assessed using an adapted instrument derived from existing tools as used in previous reviews.^6, 15–18^

### Evidence Synthesis

We *descriptively* synthesised and mapped all studies (Panel 3), but applied different strategies and criteria for the *analytical* synthesis of qualitative and quantitative data. For the quantitative synthesis, we included only moderate- or high-quality studies that reported SARS-CoV-2 cases, hospitalisations, ICU admissions, and mortality among migrant *and* non-migrant populations, in order to draw reliable conclusions from the literature. We checked 169 quantitative studies with medium- or high-quality for their eligibility; studies reporting outcomes among migrants *without* comparison groups were excluded from the quantitative synthesis.

We performed meta-analyses for the binary outcomes (SARS-CoV-2 cases, hospitalisation, ICU admission and mortality) using the effect sizes relative risk (RR) and risk difference (RD), which allow estimation and interpretation of pooled proportional and absolute risk, respectively, associated with migratory status compared to non-migrants. Given that the denominators for mortality differed (i.e., deaths based on hospitalised cases or all deaths) we performed separate analyses. As we assume that studies are likely to have functional differences and that the true effect size θ varies between studies, we fitted random-effects models using inverse variance weighting.^19^ We carefully assessed the heterogeneity between studies, used sensitivity analyses to check for robustness of models, and performed explorative subgroup analyses (Panel 2).

#### Panel 2: Measure of heterogeneity, sensitivity and subgroup analyses

The measure of between-study heterogeneity τ^2^ estimates the variations in the true effect size *θ*. It was calculated using the restricted maximum likelihood (REML) method and the Q-Profile method to compute corresponding 95% confidence intervals (95%-CI).^19, 20^ The measure *I*^2^ was not suitable for assessing heterogeneity, as it tends toward 100% when studies have large sample sizes, which is the case for many of the included studies. The measure τ^2^, however, is insensitive to the precision of the included studies. Based on τ^2^, we calculated a prediction interval (PI) that allows to quantify the range into which a future study might fall based on the evidence considered. The PI provides a meaningful interpretation of τ^2^.^18^ We used the Kenward-Roger method to calculate the 95%-CI for both the pooled estimate *θ̂* and PI.^21^ In addition, we performed subgroup analyses for study regions and migration indicators where appropriate, to identify possible explanations for heterogeneity. We used sensitivity analysis to carefully check the robustness of the models for each outcome and effect size and, as a result, excluded some studies from the meta-analysis. Therefore, we calculated influence diagnostics to detect extremely influential studies that have a high impact on *θ̂* and τ^2^. We excluded studies based on extreme or implausible values of standardised residuals, Cook’s distance, hat value, and leave-one-out cross-validation (LOO-CV) to measure direct impact on τ^2^and *θ̂*.^22^ See supplementary file pp. 102-117 for more information on the sensitivity analysis. Analysis and visualisations were performed using R 4.2.0 statistical programming language. Meta-analyses and sensitivity analyses were performed using the *meta* and *dmetar* packages.^22, 23^

#### Panel 3: Overview of identified global empirical literature on SARS-CoV-2 health outcomes and effects of pandemic control measures (e.g., lockdown) among migrants (12/2019 - 11/2021), N = 241 studies

##### Study designs

Out of 241 included studies, 186 had a quantitative design (e.g., cross-sectional, longitudinal, cohort), including 18 modelling studies, 42 were qualitative research, and 13 studies had a mixed method design.

##### Geographic scope

Most studies were conducted in high-income countries (n=167), 35 studies took place in (upper) middle-income countries, 35 in lower middle-income countries, and only five studies were conducted in low-income countries. Some studies didn’t have a specific country context (e.g. modelling studies) or looked at multiple countries.

##### Migrant groups

The migrant groups studied varied widely. Study populations consisted mainly of *a)* international migrants (n=128) as defined by (a combination of) various indicators such as region of origin, country of birth, parents’ country of birth, ethnicity, nationality, language-proficiency, or migrants/immigrants without further specification; *b)* international migrants with a specific legal status that we know of, including international students (n=10), refugees (n=36), asylum seekers (n=14), labour migrants (n=40), undocumented migrants (n=9), ICE detainees (n=3); *c)* IDP (n=5); *d)* internal migrants (mainly in India and China) (n=14); *e)* returnees (n=8); *f)* ecological / net migration flows (n=19).

##### Health outcomes

50 studies investigated infection risk, 28 transmission risk, 84 health outcomes of disease (i.e., SARS-CoV-2 cases, hospitalisation, ICU admission, mortality, or pneumonia), 16 health services access, 49 mental health, 69 looked at the effects of pandemic control measures (e.g., lockdown, mask wearing, etc.), and only seven studies addressed vaccination.

##### Study quality

Less than half of the studies (n=112, 46·5%) were classified as high quality, 92 (38·2%) as medium quality, and 37 (15·3%) as low quality.

See supplementary file Table S6, p.35ff. for study characteristics of all included studies.

For qualitative synthesis, a coding scheme following the model of Diderichsen et al. was developed and the extracted data (i.e., main findings as reported by the included qualitative studies) was coded accordingly, using ATLAS.ti software.^20^ The development of the coding scheme involved both inductive and deductive coding strategies. During the first round, codes were developed inductively from the data. They were then compared with and integrated in the existing model, which describes COVID-19-related inequalities for migrants in three categories: *exposure, risk*, and *impact*. Through iterative meetings (first within the qualitative analysis-team, then the entire review-team), we elaborated the original model as follows: 1) through inductive coding, we added further codes to the categories; 2) the original model was linear (from exposure to risk to impact), while our data showed many interrelations and feedback loops within and among the categories; 3) within each category, we distinguished between factors on the micro-, meso- and macro-level; 4) we added the category “*sources of resilience*”, which interacts with the other three categories.

## RESULTS

The search yielded 7045 records from databases and 75 from websites. After removal of 164 duplicates, 6956 records were screened based on their title and abstract; 508 reports were sought for retrieval and 480 assessed for their eligibility whereof 255 did not meet the criteria (Fig. 1). Another 16 records were included based on previous reviews (n=8) or snowball sampling (i.e., primary studies from reviews) (n=8). A total of 241 studies were included to map the global empirical landscape (Panel 3). Of these, 44 qualified for qualitative synthesis (Table 1A) and 46 for meta-analysis (Table 1B).

**Figure 1:**
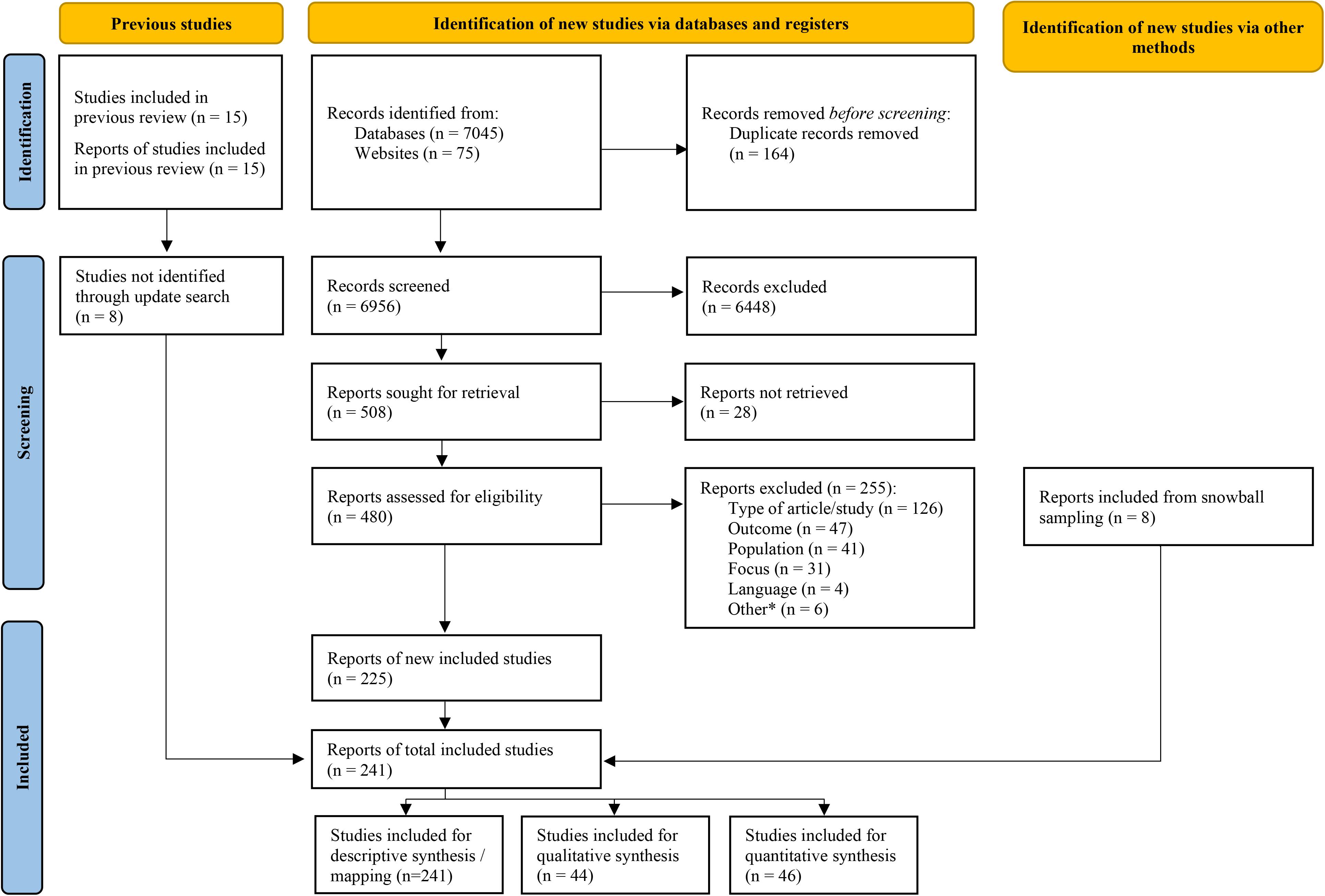
PRISMA flow-chart.

**Table 1A:** Characteristics of studies included in the qualitative synthesis.

| Author & year of publication | Country of study | Socio-economic Development of Country* | Period of Study | migrant population | Health outcomes | Study Design | Main results (as reported) | Conclusions (as reported) | Quality appraisal |
| --- | --- | --- | --- | --- | --- | --- | --- | --- | --- |
| Gele et al. 2022 | Norway | HIC | April 21, 2020 to May 15, 2020 | International migrants | effect of lockdown measures mental health | Qualitative research | Mild depression (9.1%; 40/441) and anxiety (16.1%; 71/441) was common among respondents followed by moderate depression and anxiety {depression (3.4%; 15/441), anxiety (4.1%; 18/441)} and severe depression and anxiety {depression (1.1%; 5/441), anxiety (0.7%; 3/441)}. Anxiety and depression were independent of their socio-demographic characteristics. Perceived fear of contracting COVID-19, severity and death were prominent among the respondents. Respondents experienced stigma and discrimination in addition to being at the risk of disease and possible loss of employment and financial responsibilities. In addition, poor (quality and access to) health services, and poor living condition at the quarantine centres adversely affected respondents' mental health. | Although no statistically significant association of anxiety and depression with socio-demographic variables was observed, the qualitative approach identified various factors to contribute for their [returnee labor migrants] poor mental health. Institutional quarantine centers were in poor conditions in relation to basic supplies, health, hygiene and recreational support which adversely impacted mental health of the respondents. Measures towards alleviating fear and stigma and ensuring financial securities of the population during the time of health crisis are important for preparedness against epidemics such as COVID-19. | medium |
| Babuc 2021 | Turkey | UMIC | July 2020 | Refugees | effect of lockdown measures | Qualitative research | According to the findings of the study, the suspension or slowdown of economic activities due to the pandemic has caused not only an uncertainty about the future, but also a change in familial positions and roles. Also the accepted norms of social relations have been inevitably suspended resulting in increased ambiguity. Strengthening the network of solidarity within relatives and building stronger relationships within the family are common relational strategies of the participants to mitigate the social and economic impacts of the COVID-19 outbreak (...). | Considering the fact that the lockdown measures can be tightened again at any time with respect to the increase in the spread of the disease, social and economic policies should be expanded to include the Syrian migrants. Also educational policies during the COVID-19 pandemic should include specific measures such as technological support and guidance to mitigate the risks and disadvantages that Syrian children experience in terms of distance learning. | medium |
| Biddle et al. 2021 | Germany | HIC | May 1, 2020 to July 31, 2020 | Asylum Seekers and Refugees | mental health | Qualitative research | We found substantial heterogeneity of measures taken to prevent infection, inform refugees, maintain social and health services, test for SARS-CoV-2 and quarantine positive cases. Effective inter-sectoral cooperation proved to be particularly important for coordination and implementation of measures. Need for support was expressed with regard to the improvement of infrastructure, opportunities to work with language interpreters and stronger involvement of local health experts. | Amidst multiple actors and the complexity of structures and processes, the admission authorities have been taking on essential responsibilities related to infection control on an ad hoc basis, without being sufficiently positioned to do so. In order to further contain the pandemic, a strengthening of centralised, setting-specific recommendations and information as well as their translation through the pro-active involvement of the public health authorities at the local level are essential. | medium |
| Bojorquez et al. 2021 | Mexico | UMIC | April 2020 to May 2020 | International migrants | mental health | Qualitative research | In addition to fear of contagion and economic insecurity, migrants experienced emotional distress associated with hardening migration policies, and the difficulties of having to find shelter in place in non- | To conclude, our results suggest that, as other authors anticipated, the COVID-19 pandemic and lockdown measures have important mental health consequences for migrants. (...) As recommended by inter-national | high |
|  |  |  |  |  | health services access |  | private spaces. Some CSOs continued or adapted previous psychosocial support activities, helping migrants navigate these issues, but other activities stopped amidst physical distancing measures and because of limited resources. Migrants themselves implemented some group activities. There was a surge of civil society initiatives of online support, but some shelters leaked the techno-logical and other resources to benefit from them. | organizations, instead of being limited to clinical interventions, this response should prioritize the psychosocial aspects that are the root of mental health problems in these types of populations, covering basic needs such as decent work, appropriate places to live, and access to education and health services. |  |
| Da Mosto et al. 2021 | Italy | HIC | April 2020 to September 2020 | Asylum Seekers and Refugees | mental health | Qualitative research | Even though various measures were implemented in reception centres (i.e. mass quarantine, supply of personal protective equipment, risk communication campaigns and specific governance tools) they often had a discriminatory approach towards migrants and only considered the biomedical aspects of COVID-19, excluding its social roots and repercussions. This factor, together with the lack of an effective governance system at both the local and the national level, was the most relevant issue associated with the management of the syndemic in reception facilities and affected all the social determinants that shape the health profile of RAS. | The study revealed the importance of social factors in the management of the syndemic in reception centres. It also highlighted how the underlying causes of the impact of COVID-19 are tightly correlated to the political and social approaches of local and national institutions to migration. In order to guarantee the well-being of society as a whole and successfully control the epidemic, it is necessary to consider migration as a human reality rather than an emergency, and demolish all the policies and bureaucratic systems that act as structural violence on RAS. (...) This way it will be possible to develop more inclusive approaches to public health and guarantee the conditions for RAS' empowerment. | medium |
| de Diego-Cordero et al. 2021 | Spain | HIC | April 2019 to September 2020 | Migrant workers | mental health | Qualitative research | The results show the moral debt accrued by the caregivers with the family who employ them, while worsening the physical and psychological health of many of the care-givers, due to both work overload and fear of the global pandemic. | The emotional debt and the relationship as 'quasi-family members' have contributed to increasing the work overload, since as well as the usual care, they have been responsible for all the domestic work, resulting in a non-stop working day during the lockdown, without any personal free time and being separated from the social group of their friends who live in Spain. | high |
| Mookerjee et al. 2021 | India | LMIC | 3 months into the lockdown (presumably: May 2020 to July 2020) | Internal migrants | effect of lockdown measures | Qualitative research | Financial worries were found to be endemic, with rising debt a major source of stress, and educational qualifications becoming an obstacle to earning. Returning migrants were suspected of bringing the virus from the city, and so stigmatized in their home towns and villages. However, the pandemic lockdown also showed some un-expected healthful consequences. It provided these marginalized, and always busy workers the time and space to stop working for a while, to stay home, eat home food, and take walks in the comparatively green and clean spaces of their home environments. | Any future policy about migrants must prioritize addressing the extreme financial precarity of internal migrants. This study shows the need for a kind of social security or insurance to get workers through low-or-no-income periods, but further research focused on the economic structural constraints experienced by migrant workers is necessary to shape such policy. (...) The pandemic lockdown as an enabler of health is one of three inversions that emerged in this culture-centered study (...) In this, the pandemic lockdown may be seen to have enabled a measure of agency and health in the lives of these workers, an oasis albeit temporary, and ultimately subject to the demands of the globalized cities of India. | high |
| Duggal et al. 2021 | India | LMIC | April 2020 to June 2020 | Internal migrants | effect of lockdown measures | Qualitative research | Participants reported that they were afraid of dying alone in the city without their families around so they decided to go back home. At the same time the stress of being back home without any way of financially supporting themselves and their families generated great amounts of uncertainty and helplessness in the participants. They experienced feelings of guilt and frustration at not being able to find a job and worried constantly about the future. Participants reported experiencing worries about the future, helplessness and a fear that the world and 'normal life' was about to change indefinitely. Their sense of personal agency and certainty was taken away almost overnight which generated distress in them. | The research used an intersectional frame to understand how different social identities of migrant status, gender and class coalesce together to generate unique forms of marginalization. It calls for future policy and practice work to acknowledge and integrate the lived experiences of those who remain on the edges, invisible yet critical backbones of a well-functioning and harmonious society. Finally, it emphasises that the pandemic is just a mirror being held up to all of us- and the 'reflections' and 'lessons' need to be carried forward, remembered, documented and transformed into action that is meaningful to the lives of people. | high |
| Elci et al. 2021 | Turkey | UMIC | April 5, 2020 to August 2020 | Refugees | effect of lockdown measures | Mixed-Methods: Cross-Sectional Study; Qualitative research | Our findings indicate that the initial Covid-19 measures had a higher financial impact on Syrians than on citizens when controlled for employment, wealth, and education, among other variables. In line with the literature, our research confirms that disasters' socio-economic effects disproportionately burden minority communities. | Our findings reveal that being a refugee in Turkey accelerates and magnifies the pandemic's perceived economic impact, even after controlling for social class, demographic factors, and geographical location. (...) While signalling the unequal outcomes of the Covid-19 pandemic, our research also addresses the underlying causes of these outcomes, as well as the implications of the pandemic for the future. As the Syrian displacement reached its 10th year, our results show that the livelihood of Syrians in Turkey is very much tied to the functioning of the labour market, characterised by widespread informality. | medium |
| Enriquez et al. 2021 | USA | HIC | March 2020 to June 2020 | Undocumented migrants | effect of lockdown measures | Mixed-Methods: Cross-Sectional Study; Qualitative research | Qualitative findings showed that immigration status exacerbated the negative economic impact of the pandemic, leading to severe individual and family financial strains that had cascading negative effects on undocumented students' academics and health. Quantitative findings focused on students' own prepandemic economic insecurity to show that it was associated with worse academic, financial, and health impacts during the initial months of the pandemic. | Qualitative findings highlighted how campus resources such as study space and technology were no longer available, harming students' academics. Thus, our findings suggest that the benefits of campus-wide resources, (...), depend on a continuous rather than cumulative provision of services. The quantitative results also demonstrated that being a student in the UC system, versus the CSU, was positively associated with increased negative academic, financial, and health impacts. Differences may emerge because UC campuses host a much larger number of on-campus residential students and offer more support services, thus inflicting a greater burden on UC undocumented students who were forced to relocate to their permanent homes and cut off from campus support. | high |
| Knights et al. 2021 | United Kingdom | HIC | June 18, 2020 to November 30, 2020 | Asylum Seekers and Refugees | effect of lockdown measures | Qualitative research | It was found that digitalisation had exacerbated existing inequalities regarding access for some migrant groups through a lack of access to, or knowledge of, technology; concerns about language barriers, difficulties building trust, and the risk of missing safeguarding cues in virtual consultations were also expressed. The physical closure of some surgeries was reported to have led to challenges in migrants registering with, and accessing, primary care. Communication barriers, feeling left behind in receiving support and health interventions in comparison to the general population, and a lack of access to information were issues widely raised by migrants. Additionally, they reported views of COVID-19 and COVID-19 vaccinations that ranged from acceptance to misinformation, often originating from social media or word of mouth. Some migrants experienced increased risk factors to their health and severe illness from COVID-19, partially resulting from their economic and social situations. | Pandemic-related changes to primary care delivery may become permanent; some migrant groups are at risk of digital exclusion and may need targeted additional support to access services. Solutions are needed to address vaccine hesitancy in marginalised groups to ensure equitable COVID-19 vaccine uptake. | high |
| Filippi & Giliberti 2021 | Italy | HIC | February 2020 to May 2020 | Asylum Seekers and Refugees | effect of lockdown measures | Qualitative research | By analyzing the transformation of Italian reception policies in the last years, the article shows the relationship between these changes and the condition of refugees and asylum seekers in these centers during the COVID-19 pandemic. Overcrowded housing, the absence of institutional guidance on managing the situation, and the interruption of many migrants' migratory projects are the main findings that emerged. | Our analysis encourages us to reflect upon two points. The first reflection is that the pandemic, as a new "total social event," adds another layer of complexity to the Italian reception system, which was already under duress due to the Salvini Decree that introduced changes that did not benefit refugees and asylum seekers. The second reflection is that the pandemic has amplified and brought to the fore those pre-existing dynamics of marginalization and exclusion, which often feature in the stories of migrants who enter the Italian reception system. | medium |
| Geuijen et al. 2021 | Netherlands | HIC | May 2020 to June 2020 | International migrants | effect of lockdown measures | Qualitative research | Through taking into account experiences of support workers, this study reported on the impact of the COVID-19 pandemic upon Dutch migrant families who have children with ID. Two main themes related to the COVID-19 pandemic were identified: Work of support workers during the COVID-19 pandemic and Impact of the COVID-19 pandemic upon migrant families who have children with ID. Given that video calling often proved to be an unsuitable alternative (children did not understand) through which to stay in touch, parents missed their children very much. | The current study makes us cognisant of the fact that migrant families who have children with ID have encountered unique challenges (language barriers and emergency care plan) during the COVID-19 pandemic, whereas other challenges (additional burden and missing children who live at residential facilities) are in accordance with the challenges experienced by families not from a migrant background. Consequently, in light of the potential future waves of COVID-19 and expected further lockdowns, it is important that support workers are aware of the importance of language barriers, cultural background, and the increased burden associated with staying in touch with migrant families who have children with ID. | medium |
| Guruge et al. 2020 | Canada | HIC | First six months of the pandemic (summer and fall 2020) (not further specified) | International migrants | effect of lockdown measures | Qualitative research | Results revealed that parenting experiences during the pandemic entailed dealing with changing relationships, coping with added burdens and pressures, living in persistent fear and anxiety, and rethinking lifestyles and habits. Amid these changes and challenges, some parents managed to create opportunities for their children to improve their diet, take a break from their rushed lives, get in touch with their cultural and linguistic backgrounds, and spend more quality time with their family. | While immigrant parents exhibit remarkable resilience in dealing with the pandemic-related meso and macro-levels restrictions, funding and programs are urgently needed to support them in addressing the impact of these at the microlevel. | high |
| Hari et al. 2021 | Canada | HIC | April 2020 to June 2020 | International students | effect of lockdown measures | Qualitative research | We found that international students experienced the pandemic transnationally and faced increased challenges, which heightened their reliance on support from transnational families, and generated anxieties about their future career and mobilities. | We know from this study that IS are drawing on their transnational resources, it is, therefore, imperative that we rethink locally based supports. (...) There is a need to provide more effective locally emplaced, pragmatic, emotional and social support. These recommendations require multi-stakeholder engagement and support, including universities (as key anchors for this group), governmental bodies (Immigration, Refugees and Citizenship Canada, Employment and Social Development Canada, as well as provinces and municipalities) and civil society organizations. | high |
| Im & George 2021 | USA | HIC | April 2020 to October 2020 | Refugees | effects of lockdown measures<br>health services access | Mixed-Methods: Cross-Sectional Study; Qualitative Research | Providers highlighted several preexisting health challenges that have been barriers for the refugee community before the pan-demic. The rapid assessment highlighted that underlying health concerns and limited orientation to the U.S. healthcare systems were identified as two main overarching themes that have caused challenges to the refugee community. (...) For refugees, the impact of the COVID-19 pandemic manifests through multiple factors such as health, social, and economic hardships, which were likely exacerbated by barriers to access to virtual service platforms and resources and communication breakdown with service providers. | This study proposes several implications for future research and policy in social work with refugee populations during the post-COVID time. (...) The COVID-19 pandemic uncovers cumulative issues that have obstructed healthcare and social services in the refugee community and demands a more culturally nuanced, multipronged approach to refugee community wellness and integration through building capacity and collaborative partnerships among professionals and members, and volunteers of the refugee community. | high |
| Lui et al. 2021 | Hong Kong, China | HIC | May 2020 to August 2020 | Migrant workers | effects of lockdown measures | Qualitative research | FDWs [foreign domestic workers] reported a dual-country experience of the pandemic, where they expressed concerns about local transmission risks as well as worries about their family members in their home country. Changes to their current work situation included how their employers treated them, as well as their employment status. FDWs also cited blind spots in the Hong Kong policy response that also affected their experience of the pandemic, including a lack of support from the Hong Kong government. | The current study demonstrates that the COVID-19 pandemic has exacerbated the existing power dynamics that constrain FDWs in Hong Kong. Not only is there a greater need to provide for family members back in FDWs' home countries, but there are also increased pressures from employers and a lack of support from the Hong Kong government. Through this study's findings of the current situation and challenges faced by FDWs in Hong Kong during the COVID-19 pandemic, it is clear that policy-level interventions are needed to mitigate the particularly negative effects on FDWs. More supportive policies should be adopted that not only consider the specific needs of FDWs but listens to them. | high |
| Kaur-Gill et al. 2021 | Singapore | HIC | Not reported | Migrant workers | mental health | Qualitative research | The findings in our article reveal the interplays of mental health meanings situated within a structural context of employment and a cultural environment that manifests unequal power relationships and indebtedness. During the Circuit Breaker, domestic workers in already poor employment conditions found themselves at greater precarity and limited agency. Furthermore, the analysis captures the temporalities of mental health meanings articulated within the performance of conducting migrant domestic work in confined working conditions. Precarious migrant journeys include vicious debt cycles, unethical agents, and corrupt employment practices, detailing the scripts of mental health stressors. | Our findings reveal that the COVID-19 pandemic lockdowns magnified how these practices of soft violence (...) manifested, disabling migrant worker mental health and well-being. As precarity is structured in the power imbalance to hire, fire, and deport domestic workers at will by employers, practical implications for mental health interventions must discuss labor and immigration policies of MDWs [migrant domestic workers] bound to Singapore. (...) Beyond culturally tailored programs on mental well-being in the workplace, solutions must consider the structural factors embedded in the policies of hire and the indentured nature of labor to address MDWs discussions of mental health contentions. | high |
| Kuhlmann et al. 2020 | Austria, Germany, Italy, Poland, Romania | HIC | January 2020 to May 20, 2020 | Migrant workers | effect of lockdown measures | Qualitative research | Results suggest that undersupply of carers coupled with cash benefits and a culture of family responsibility may result in high inflows of migrant carers, who are channelled in low-level positions or the informal care sector. COVID-19 made the fragile labour market arrangements of migrant carers visible, which may create new health risks for both the individual carer and the population. | The results highlight the weaknesses of existing health labour market arrangements in the LTC sector, which stretch far beyond poor workforce management. As the COVID-19 pandemic revealed, these conditions may directly impact population health and the health and wellbeing of the migrant carers, thus becoming fundamentally a public health policy issue. The research also calls for European health labour market regulation and governance models, which help to balance national interests and connect health system needs, health labour markets and the individual migrant carers. Including LTC migrant carers more systematically in health workforce governance and research, therefore, must become an issue of public health and European policy. | high |
| Kumar et al. 2021 | India | LMIC | March 24, 2020 to May 31, 2020 | Internal migrants | mental health | Qualitative research | (...), the findings suggest that the participants' plights have been exacerbated by the COVID-19 pandemic. (...) As our participants reported, they dealt with multiple hardships, including loss of jobs and income, police brutality, Othering, and differential treatment, difficulties accessing health services, food, cash transfer and other social programmes during the first COVID-19 pandemic nationwide lockdown. (...) Consequently, many of them became anxious, and were uncertain about their future. With no money and savings, many participants struggled to live and survive during the lockdown. | The findings indicate the need for policy responses to focus on addressing conditions of work, terms of employment and access to necessities for Indian MW, including ensuring conditions for a prompt job-ready recovery and mental health care after the COVID-19 pandemic. | medium |
| Li et al. 2021 | China | UMIC | January 2020 to July 2020 | Internal migrants | effect of lockdown measures<br>mental health | Mixed-Methods: Cross-Sectional Study; Qualitative Research | The research revealed three distinct long-term vulnerabilities faced by migrant worker families experiencing difficulties: physical/material, motivational/ attitudinal and social/organisational. At the same time, during our project helping migrant workers fight COVID-19, we recognised these families' capacities. | Thus, we believe that sustainable and systematic intervention is needed to establish a long-term service mechanism for migrant workers' disaster management, and improve social security policies, provision and regulations. This will promote disaster management awareness and behaviour change among migrant worker families, improve the flexibility and quality of social services, effectively reduce the vulnerabilities of migrant worker families and enhance their capacities. | medium |
| Martin-Anatias et al. 2021 | New Zealand | HIC | May 2020 to June 2020 | International migrants | effect of lockdown measures | Mixed-Methods: Cross-Sectional Study; Qualitative Research | Both our survey and our interviews indicated that many women, particularly mothers, experienced increased demands on their time and emotional energy in comparison to men, particularly in fields related to childcare and domestic labour, and that they also experienced a decline in wellbeing linked to heightened levels of exhaustion and stress. (...) Their [Indonesian migrant mothers] stresses were instead linked to feelings of anxiety, frustrations and inadequacy with homeschooling their children in English (...) our research indicates that both second-language speaking status and a prevailing gender ideology of 'state Ibuism' (as opposed to neoliberal feminism) were critical factors differentiating Indonesian migrant mothers' experiences from those of many other mothers in NZ. | Our research corroborates prevailing conclusions offered by existing work on gender relations during the 2020 coronavirus pandemic in NZ and beyond. (...) While feminist scholars are certainly correct to highlight the pervasiveness of gendered inequality during lockdown, an attention to distinct meanings of womanhood and motherhood afford a more nuanced approach, illuminating how lockdowns can offer not only gendered challenges but also gendered rewards, and cast light on measures that could be taken in future to enhance the experience of living through a lockdown in one's intersectional specificity. | medium |
| Martuscelli 2021 | Brazil | UMIC | March 27, 2020 to April 6, 2020 | International migrants | effect of lockdown measures | Qualitative research | The results indicate that refugees face three challenges connected to this pandemic: (a) same challenges as Brazilians due to their labor vulnerability social identity, (b) challenges aggravated by the pandemic due to their identity of nonnationals including access to information and services, and (c) new challenges due to their social identity of forced displaced nonnationals including closing of migration services and borders and the feeling of "living the pandemic twice." | This study reinforces the need to understand how forcibly displaced people in the Global South are affected by pandemics and responses to them adopting an intersectionality approach, especially in situations that are not refugee camps, like Brazil. Granting refugees the possibility to reflect on their experiences provides valuable insight on how to involve and engage them in responses to global health emergencies. | high |
| Nasol & Francisco-Menchavez 2021 | USA | HIC | 2017 to 2020 (not further specified) | Migrant workers | effect of lockdown measures<br>health services access | Qualitative research | Based on quantitative and qualitative data with Filipino workers before and during the COVID-19 crisis, we find that RCFEs have failed to comply with labor standards long before the pandemic where the lack of state regulation denied health and safety protections for home care workers. The racial inequities under COVID-19 via the neoliberal approach to the crisis puts home care workers at more risk. (...) Last, while the experiences of Filipino home care workers during the pandemic expose the elder care industry's exploitation, we find that they are also creating strategies to take care of one another. | Our study demonstrates how legal protections and public assistance, or the lack thereof, is a major social determinants of care workers' health. The experiences of Filipino home care workers during the COVID-19 pandemic broadly expose the elder care industry's exploitation of racialized migrant workers and the government's neglect in protecting them, endangering key essential workers during a historical public health crisis. And yet, we have found that Filipino care workers are crafting "communities of care" that circulate different forms and ethics of care among themselves. (...) Although these informal networks are not a sustainable replacement for state-sponsored health care and health resources, we uplift the innovative strategies of Filipino care workers to demonstrate their ingenuity and commitment to caring for themselves and one another. | medium |
| Qi & Ma 2021 | Australia | HIC | February 2020 to November 2020 | International students | effect of lockdown measures | Qualitative research | Our thematic analysis highlights participants' experiences and views of Australia's crisis responses in the four areas of financing, third-country transit, visas and immigration, and pandemic management. | How countries live up to their values is critical to their global reputation and soft power. Australia has a range of soft power strengths. Nevertheless, ill-considered policies have also caused much international and domestic criticism, especially in areas such as climate change, asylum seekers and foreign aid. The list has extended to include its treatment of temporary migrants including international students. Australia needs to push for a broader vision for international education that enhances its international engagement not only for economic gains, but also for effective communication of Australia's democratic and inclusive values. | high |
| Rao et al. 2020 | India | LMIC | December 1, 2019 to June 2020 | Migrant workers | effect of lockdown measures | Qualitative research | While social identity and regional location are seen as established mediators of migrant experiences, in the present context their role and impact was subdued. Social policy frameworks in the destination states, or their absence, played a key role in shaping migrant experience. Kerala, with a better social infrastructure, but also a specific legal and policy framework recognizing the role and contributions of migrant workers as 'guest workers', was able to respond more effectively to their needs and demands. Amongst the other states, Maharashtra falls in between: with some recognition of workers' rights yet divided by a strong movement of regional pride and exclusivity. Gujarat and Uttar Pradesh emerge at the bottom, offering no rights or benefits to the migrant workers, and in fact, denying any claims to citizenship that they put forth. | Constitution of India offers all citizens the freedom to live and work in any part of the country, yet the pandemic exposed the loopholes in ensuring rights in the context of population mobility, placing the spotlight on the nature of rights and their portability. While this is critical, the discursive agenda of social protection is now focused only on basic needs to food and shelter, rather than a transformational agenda that could eventually ensure equal opportunities and choices to all citizens of the country to lead fulfilling lives. | medium |
| Reynolds et al. 2021 | Mexico | UMIC | August 2020 | Asylum Seekers | effect of lockdown measures | Qualitative research | The pandemic caused significant mental health burdens but no perceived adverse physical effects, with the U.S. border closure and health care access barriers as more pressing concerns. Participants reported access to information about COVID-19 but had varied levels of know-ledge and adherence to disease reduction strategies due to camp conditions. Most participants believed that they had special protection from COVID-19, including strong immune systems or from God. The nongovernmental organizations providing health care and sanitation faced multiple challenges to implement new policies to manage COVID-19. The institution of required temperature checks and quarantine of COVID-19 positive patients led to distrust, decreased seeking of health care services among asylum seekers, and possible underreporting of COVID-19 cases. | Our findings among asylum seekers in a Matamoros camp highlight the challenges to implementing disease reduction policies in low-resource congregate camps. Policies to address disease outbreaks focusing on the social determinants of health, health care access barriers, and community engagement may be more acceptable to asylum seekers, suggesting the need for effective strategies to provide prevention information that complements such measures. | medium |
| Sabar et al. 2021 | Israel | HIC | March 2020 to May 2020 | International migrants | effect of lockdown measures | Mixed-Methods: Qualitative research; Cross-Sectional Study | During the lockdown, the challenges embedded in caregiving in Israel were amplified and new challenges emerged, such as the inability to leave the employer's premises, which some likened to being imprisoned. The major hardships voiced in all interviews were loneliness, fear of contracting the virus, worries about the safety of their families back home, and anger at restrictions imposed without consulting them by the assisted living facilities or their employers. At the same time, they expressed sincere gratitude for being employed, paid and safe. | Philanthropy for Filipinos in need in Israel and back home was found to be an effective coping mechanism. While lockdown conditions increased the fragility of the marginalized migrant workers, their ability to donate funds empowered them at both individual and communal levels. Developing a donor identity reinforced their sense of belonging to the Filipino community in Israel and back home. | medium |
| Sanna 2021 | Romania/ Italy | UMIC/ HIC | March 2020 | Returnees | effect of lockdown measures | Qualitative research | Among the reasons to return during the pandemic that my informants mentioned there were: the sudden loss of a job due to temporary or permanent closing of businesses during the lockdown in Italy and elsewhere; the closing of universities, in the case of Romanian students studying abroad; and the impossibility of affording healthcare in a foreign country due to not having a regular work contract. In other cases, of course, it was the fear of being sick in a foreign country that mattered most. | In this paper, I have shown how the image of returning migrants during the first month of the COVID-19 pandemic in Romania was constructed by the media in March 2020: by very quickly associating the migrants with the virus, by juxta-posing the noble acts of Romanians left in Italy with the criminal acts of those who came back, and by reinforcing such juxta-position by giving voice to Romanians who stayed in Italy. I have also shown that Romanians who did not come back were characterized in depth, while descriptions of returning Romanians, except for a few cases, remained vague. | medium |
| Sanò & Della Puppa 2021 | Italy | HIC | February 2020 to May 2020 | International migrants | effect of lockdown measures | Qualitative research | These two interviews show how the mobility and immobility of these subjects were differently conditioned by the intersection of several factors. (...) That is to say, the closure of the Trentino hotels and restaurants during the winter season has meant that most of the migrants employed in this sector have moved to the city, determining, among other things, the opening of services to low threshold able to accommodate all the people left without work in the valleys. On the contrary, in the Plain of Gioia Tauro, the slowdown in production was not determined by a general closure of agricultural activities, but, mainly, by the fact that, at the beginning of the pandemic, a good part of the labourers present were without regular contract and thus could not go to the workplace. | The ethnographic cases we described show the need to rethink the conceptual categories with which we usually analyse mobility, illuminating the links it has with national policies, with territorial administrative provisions, with the economic and social fabric of an area, with individual and not necessarily 'rational' choices of individuals. Observed in these terms and based on these links, one of the distinctive features of contemporaneity, namely mobility, reveals all its political nature, the disempowering character that accompanies it, as well as the economic interests connected to the productive function of those who find themselves imprisoned in the multiplicity facets of the prism of (im)mobility. | medium |
| DeCarlo Santiago et al. 2021 | USA | HIC | June 2020 to July 2020 | International migrants (including international students) | effect of lockdown measures | Qualitative research | Qualitative analyses revealed three major categories of themes. Challenges include difficulty adjusting; fear, worry, and uncertainty; financial concerns; academic challenges; lack of connection. Sources of Support include school and community supports. Coping and Adaptation includes the ways families have coped with COVID-19. | Despite limitations, the present study highlights the numerous difficulties that newcomer families are experiencing during COVID-19. Although increased distress is common (...), newcomer families may experience difficulties that are compounded by the resettlement experience or barriers based on status. In particular, schools must attend to digital literacy, accessible information (...), regular connection to teachers and school staff, in addition to challenges associated with basic necessities, health, and social-emotional distress. Professional development training focused on newcomer experiences could ensure all school staff are equipped with the knowledge and tools necessary to support newcomer youth. | medium |
| Singh et al. 2020 | India | LMIC | Not reported | Migrant workers | effect of lockdown measures; mental health | Qualitative research | Our study found that physiologic requirements, safety, and security were the predominant needs of the migrant workers during their stay in a shelter home during COVID-19-related lockdown. All of the migrants were satisfied with the need-based services provided to them in Chandigarh. The migrant workers were worried about being infected by COVID-19 and were uncertain about when they would be able to return to their native homes. | Lockdown drained migrant workers both physically and emotionally. Despite their hardship and suffering, most of the migrant workers supported the government's decision to impose lockdown. Comprehensive support should be provided to all migrant workers in all states of India to combat the migrant crisis. This support, as was provided in Chandigarh during lockdown, would safeguard the well-being of migrant workers in India. | medium |
| Sohel et al. 2021 | Bangladesh | LMIC | April 2020 to November 2020 | Migrant workers | effect of lockdown measures | Qualitative research | The results show that COVID-19 induced lockdown significantly affects informal migrants' household food security. Most participants report severe complications due to food insecurity like consuming less food, price hiking, no fish or meat, potato, and vegetable. Food insecurity leads to migrant's chronic food shortage, starvation, malnutrition of mother and children, and unhealthy food. As a result, the way of life of informal migrants has been directed to more fragility and vulnerability during the pandemic; even they are not affordable to maintain a minimal level of family affairs and necessity. | The COVID-19 led pandemic lockdown severely impacts food security among informal migrants' households. Most households confront enormous problems and challenges to meet up the daily food and its associated elements. (...) Moreover, they did not get what they needed at the time of the crisis caused by the COVID-19 pandemic at a satisfactory level from the government and other agencies. This study also uncovers the post-lockdown reality of impoverished informal migrants who are often excluded from Bangladesh's social safety net facilities. | medium |
| Srivastava et al. 2021 | India | LMIC | Not reported | Internal migrants | effect of lockdown measures; mental health | Qualitative research | The findings of the present research shows that internal migrant workers were highly affected due to the global pandemic, which bombarded multiple stressors on them causing physical stress, emotional stress, and social stress in their lives, which were created due to financial crisis, lack of social support, and fear and uncertainty related to the global pandemic. However, the workers looked after the options available to them and chose problem-oriented strategy by directly dealing with the crisis, emotion-oriented strategy by family and friends' support, as well as cognitive appraisal of the pandemic situation by bringing optimistic thoughts to deal with these stressors. | The present work strengthens the earlier studies on uncovering the stresses of migrant workers along with suggesting available options to deal with them and emphasizes the role of family and proper emotional support in the management of pandemic-related stress among internal migrant workers. | medium |
| Tang et al. 2021 | China | UMIC | April 2002 to July 2020 | Rural migrants | effect of lockdown measures | Qualitative research | The study finds that rural migrants suffered from serious social impacts due to COVID-19, especially during the associated lock-down period. Despite some similar impacts, influences of COVID-19 varied among rural migrants at different life-cycle stages, due to variations in human capital, family burdens, role in a household, and ability to find part-time work. Receiving little support from governments and employers, rural migrants tended to adopt household strategies to deal with difficulties related to COVID-19. Within a household, they assisted each other and worked as a unit to maximize resources and reduce risks. Traditional family values were highly praised by rural migrant households during the period. | Receiving little support from government and employers, rural migrants in China have to adopt a household strategy as a means of survival when they cope with disaster. If governments want to reduce the burden of COVID-19 on disadvantaged households, they need to pay more attention to rural migrants, even though they are neither local urban residents nor actual rural residents. (...) In the long-term, the social security system for rural migrants should be further improved. It is necessary to expand the accessibility of basic social services, such as medical insurance, housing funds, and pensions to rural migrants, and to include rural migrants in the housing security system, in case they experience difficult times. | medium |
| Thomas et al. 2021 | India | LMIC | March 1, 2020 to April 27, 2020 | Migrants workers, Refugees | effect of lockdown measures | Qualitative research | The outcomes of this study shed light on (1) the most urgent needs that need to be addressed per population group, (2) the variety of state-level responses as well as best practices observed to deal with mitigation issues and (3) opportunities for quick relief as well as more long-term solutions. | The COVID-19 pandemic has not only reduced people's means of maintaining a livelihood but has simultaneously revealed some of India's long-standing problems with infrastructure and resource distribution in a range of sectors, including nutrition and health, education, etc. There is an urgent need to construct effective pathways to trace and respond to those people who are desolate, and to learn from – and support – good practices at the grassroot level. | medium |
| Tosh et al. 2021 | USA | HIC | August 2020 to November 2020 | ICE detainees | risk of infection | Qualitative research | Our findings suggest that migrant detention and deportation present distinct challenges that undermine attempts to contain the spread of COVID-19. We provide testimonies from migrant detainees who speak to these challenges in unsettling personal terms. Our interviews highlight the insufficient actions by Immigration and Customs Enforcement (ICE) to contain the spread of the pandemic and a troubling lack of due process in immigration court proceedings. | Based on these findings, we argue that reducing the number of migrants detained in the United States is needed not only in the context of the COVID-19 pandemic but also as a preventative measure for future health crises. Reductions can be achieved, in part, by reforming federal immigration laws on “mandatory detention.” | medium |
| B.C. et al. 2021 | Nepal | LMIC | April 21, 2020 to | Internal and International migrants | mental health | Mixed-Methods: Cross- | Mild depression (9.1%; 40/441) and anxiety (16.1%; 71/441) was common among respondents followed by moderate depression and anxiety {depression (3.4%; 15/ | Depression and anxiety were high among quarantined population and warrants more research. Institutional quarantine centers of Karnali province of Nepal were in | high |
|  |  |  | May 15, 2020 |  |  | Sectional Study; Qualitative Research | 441), anxiety (4.1%; 18/441)} and severe depression and anxiety {depression (1.1%; 5/441), anxiety (0.7%; 3/441)}. Anxiety and depression were independent of their socio-demographic characteristics. Perceived fear of contracting COVID-19, severity and death were prominent among the respondents. Respondents experienced stigma and discrimination in addition to being at the risk of disease and possible loss of employment and financial responsibilities. In addition, poor (quality and access to) health services, and poor living condition at the quarantine centres adversely affected respondents' mental health. | poor conditions which adversely impacted mental health of the respondents. Poor resource allocation for health, hygiene and living conditions can be counter-productive to the population quarantined. |  |
| Bauza et al. 2021 | India | LMIC | May 2020 to July 2020 | Returnees | effect of lockdown measures | Mixed-Methods: Cross-Sectional Study; Qualitative Research | Respondents reported high compliance with important preventative measures, including staying home as much as possible (94%), social distancing (91%), washing hands frequently (96%), and wearing a facial mask (95%). Additionally, many respondents reported job loss (31%), financial challenges (93%), challenges related to staying home whether as a preventative measure or due to lockdowns (57%), changes in types and/or amount of food consumed (61%), and adverse emotional effects as a result of the pandemic and lockdown. (...) Although the research revealed high compliance with preventative measures, the pandemic and associated lockdowns also led to many challenges and hardships faced in daily life particularly around job loss, economic security, food security, and emotional wellbeing. | The results underscore the vulnerability of marginalized populations to the pandemic and the need for measures that increase resilience to large-scale shocks. (...) In particular, there is a need for greater government response to limit harm to livelihood and mental, social, and nutritional health during any lockdown periods required to reduce disease spread. | high |
| Vosko & Spring 2021 | Canada | HIC | 2020-2021 (not specified) | Migrant workers | effect of lockdown measures<br>Other: health outcomes of disease | Mixed-Methods: Primarily Qualitative Research | Although estimates vary by source, over 1000 migrant farmworkers in Ontario tested positive for COVID-19 between April and July 2020. Thus, while Ontario documented 36594 cases by July 2020 (i.e. 250 per 100000) (...), the rate of infection among migrant farmworkers, 20,015 of whom entered Ontario during the spring and summer growing season, was approximately 4996 cases per 100000 people. (...) The magnitude of illness among migrant farmworkers in Southern Ontario, and Canada more broadly, reflects the emphasis, deeply ingrained in programs emblematic of migration management such as the SAWP, on keeping labour costs low in the interest of protecting the national food supply. | Taking Southern Ontario as its focus, this article reveals how the federal government response to COVID-19 in agriculture perpetuated the effects of longstanding laws and policies requiring migrant farmworkers, circumscribed in their ability to politically mobilize on account of their institutionalized deportability, to shoulder disproportionate amounts of economic, social, and health risks. Centering the transnational character of migrant farmworkers' renewal, it identifies meaningful interventions to limit the structural disempowerment of migrant farmworkers and the externalization of their social reproduction. | medium |
| Yee et al. 2021 | Singapore | HIC | June 21, 2020 to July 16, 2020 | Migrant workers | mental health | Qualitative research | Three theme categories were derived from 27 interviews: migrant worker concerns during COVID-19, coping during COVID-19 and priorities after COVID-19. Major stressors in the crisis included the inability to continue providing for their families when work is disrupted, their susceptibility to infection in crowded dormitories, the shock of receiving the COVID-19 | We identified coping strategies employed by the workers in quarantine, many of which were made possible through the considered design of care and service delivery in mass quarantine facilities in Singapore. These can be adopted in the set-up of other mass quarantine facilities around the world to support the health and mental well-being of those quarantined. | medium |
|  |  |  |  |  |  |  | diagnosis while asymptomatic, as well as the isolating conditions of the quarantine environment. The workers coped by keeping in contact with their families, accessing healthcare, keeping updated with the news and continuing to practise their faith and religion. They looked forward to a return to normalcy after the crisis with keeping healthy and having access to healthcare as new priorities. | Our findings highlight the importance of targeted policy intervention for migrant workers, in areas such as housing and working environments, equitable access to healthcare, and social protection during and after this crisis. |  |
| Zambrano-Barragan et al. 2021 | Peru, Colombia | UMIC | July 2020 to September 2020 | International migrants | health services access | Qualitative research | We found that forced migrants from Venezuela in both Colombia and Peru face common obstacles along their access trajectories to healthcare, which we summarize as legal, financial, and relating to discrimination and information asymmetry. By limiting effective access to care during the pandemic, these obstacles have also affected migrants' ability to cover the costs of basic needs, particularly food and housing. Our study also found a prevalent reliance on alternative forms of care, such as telemedicine, easy-to-access pharmacies, and extra-legal care networks. | We conclude that COVID-19 has exacerbated pre-existing conditions of informality and health inequities affecting Venezuelan migrants in Colombia and Peru. (...) These findings also provide guidance for specific policy recommendations. Given legal and financial obstacles, it is important to facilitate access to public health services irrespective of migratory status. Access should give special attention to COVID-19 care in the short and medium term but should focus on creating and strengthening systems of access in the long term. | medium |
| Hargreaves et al. 2021 | EU/ EEA countries and the UK | - | June 15, 2020 to March 10, 2021 | Asylum Seekers, Refugees, International migrants | effect of lockdown measures<br>Other: health outcomes of disease vaccination | Qualitative research | Some migrants have experienced a range of direct and indirect health and social impacts including increased discrimination, more protracted lockdowns and severe restrictions on movement, and travel restrictions and border closures impacting family reunifications and the asylum process. Migrants in precarious work environments may have been at increased risk of losing jobs and sources of income. Certain migrant groups have a range of risk factors and vulnerabilities to COVID-19 that put them at higher risk of exposure, including occupational risk, overcrowded accommodation (including residing in camps, detention facilities, and homeless shelters), and/or lower levels of accessibility to public health services and messaging. There is emerging evidence of low COVID-19 vaccination rates in some migrant and ethnic minority groups in the EU/EEA. | These findings have immediate implications for national public health responses to reduce transmission and support COVID-19 vaccine rollout, with implications beyond the pandemic to promote health equity, human rights, and universal health coverage in marginalised migrant populations. | medium |
| Lusambili et al. 2020 | Kenya | LMIC | October 2020 | Refugees | Health services access | Qualitative research | Our findings suggest that within the first eight months of COVID-19, preferences for home deliveries by refugee women increased and health care workers reported having observed reduced utilisation of services and delayed care. Fear, economic challenges and lack of migrant-inclusive health system policies were key factors influencing home deliveries and delayed and low uptake of facility-based care. | Findings from this study show the need for refugee inclusive public health policies, particularly during a pandemic, and the need to tailor health care services for refugees at facilities and in communities. The findings also make clear the urgent need to mitigate and lower barriers that prevent refugee women from seeking care at health facilities. Lastly there is need for methodological design tailored towards refugees' unique needs | medium |
| *: Socioeconomic Development of Country based on the World Bank country classification by income; HIC: High income countries; UMIC: Upper-middle income countries; LMIC: Lower-middle income countries; LIC: Low income countries. |  |  |  |  |  |  |  |  |  |

**Table 1B:**
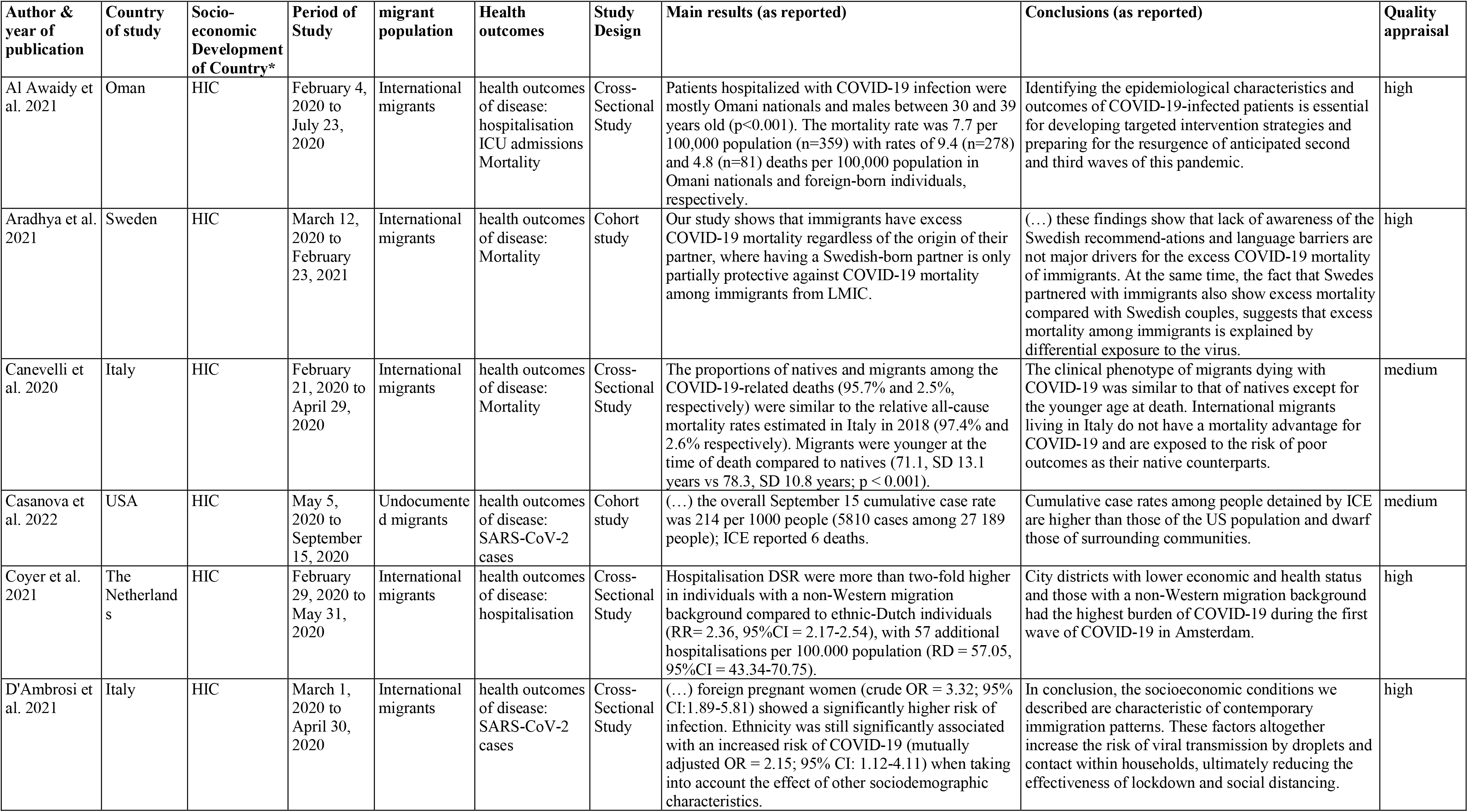
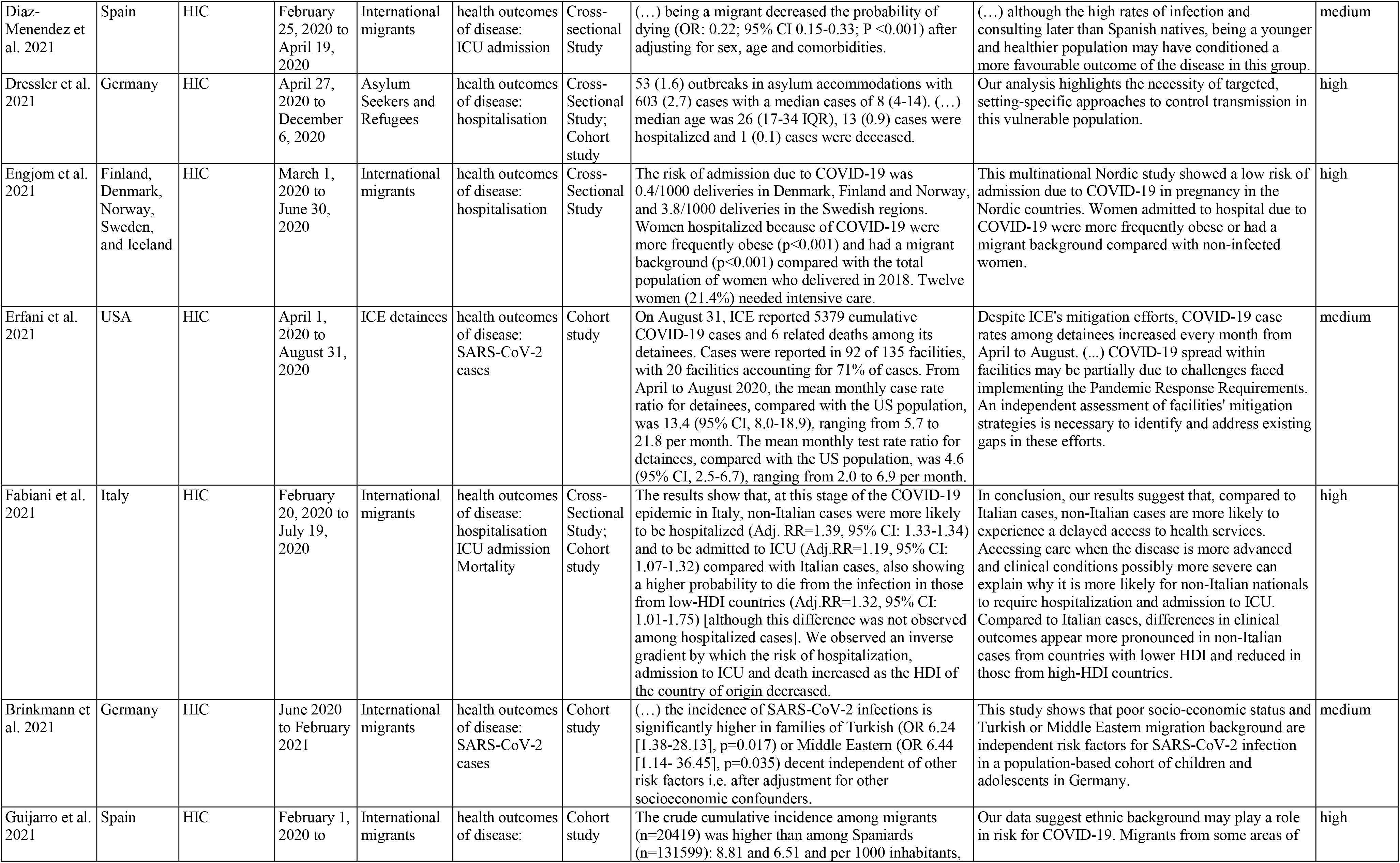
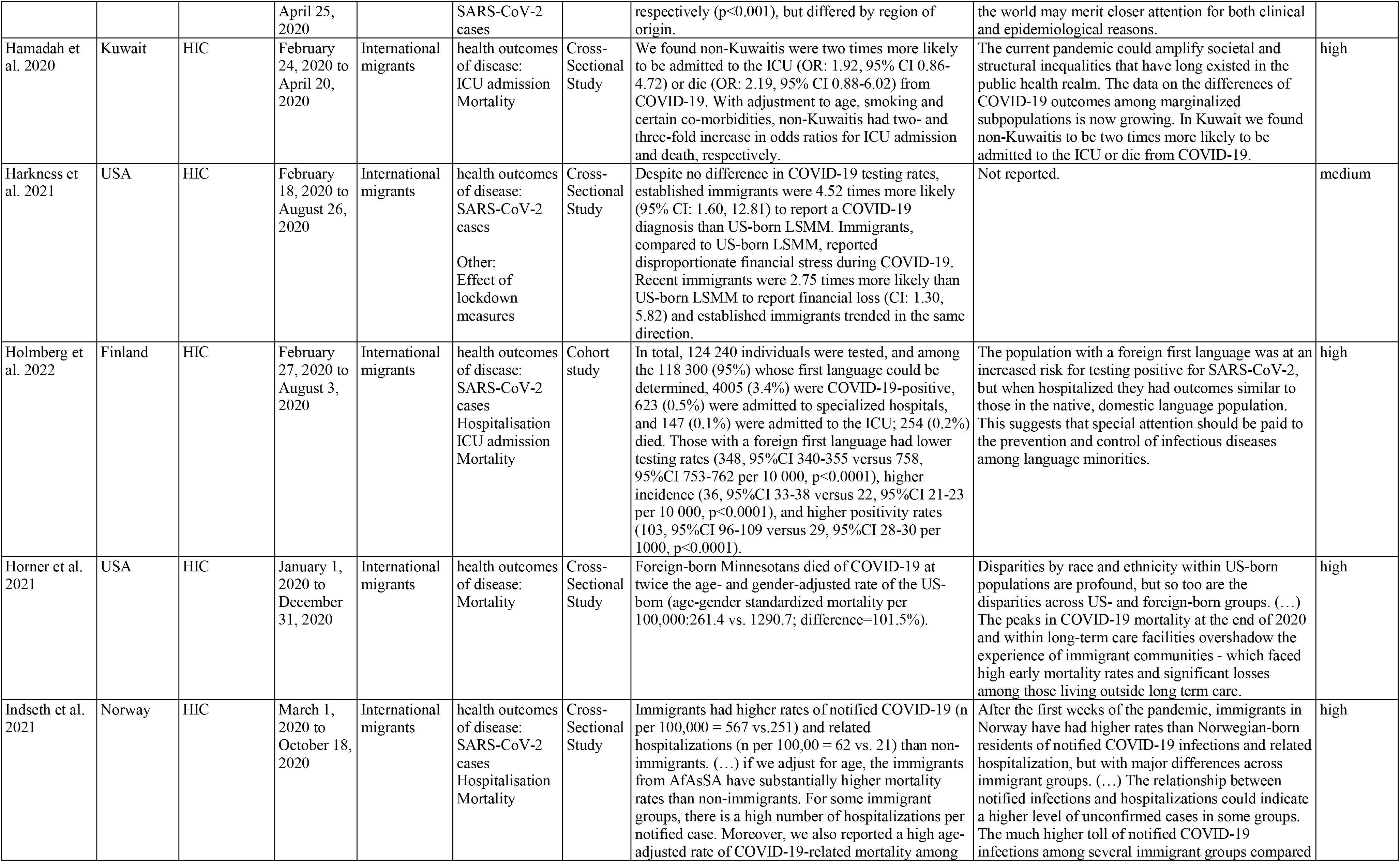
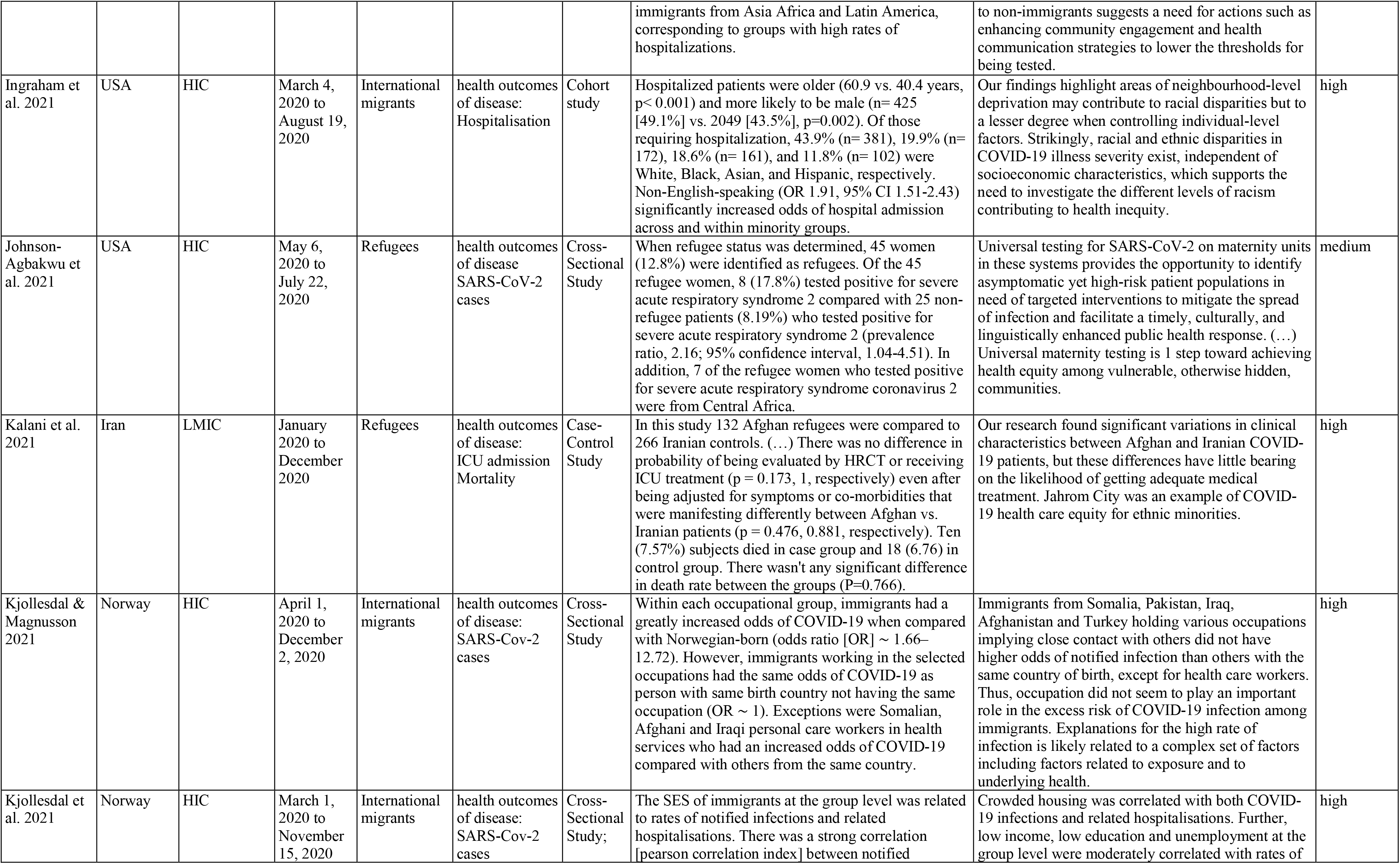
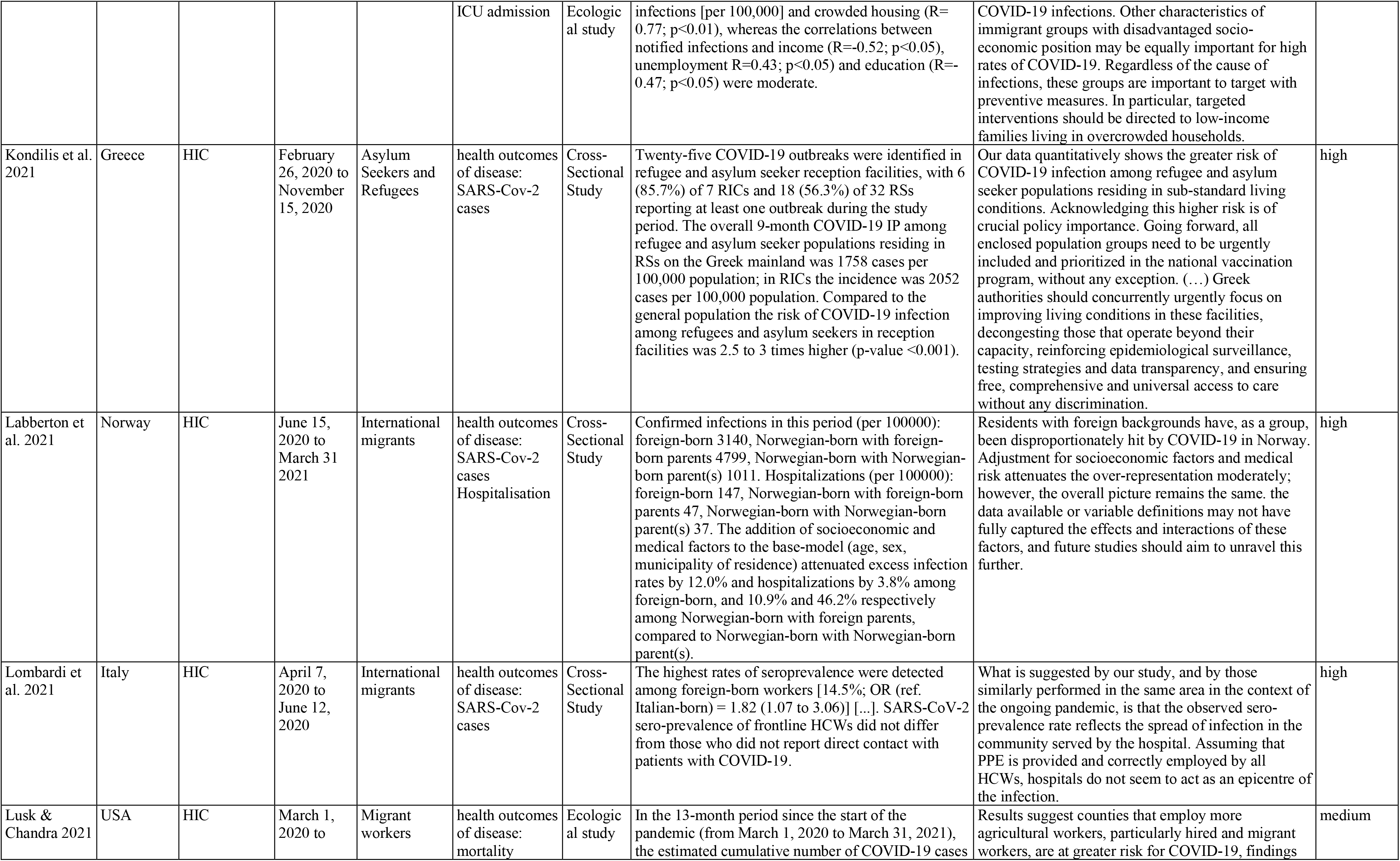
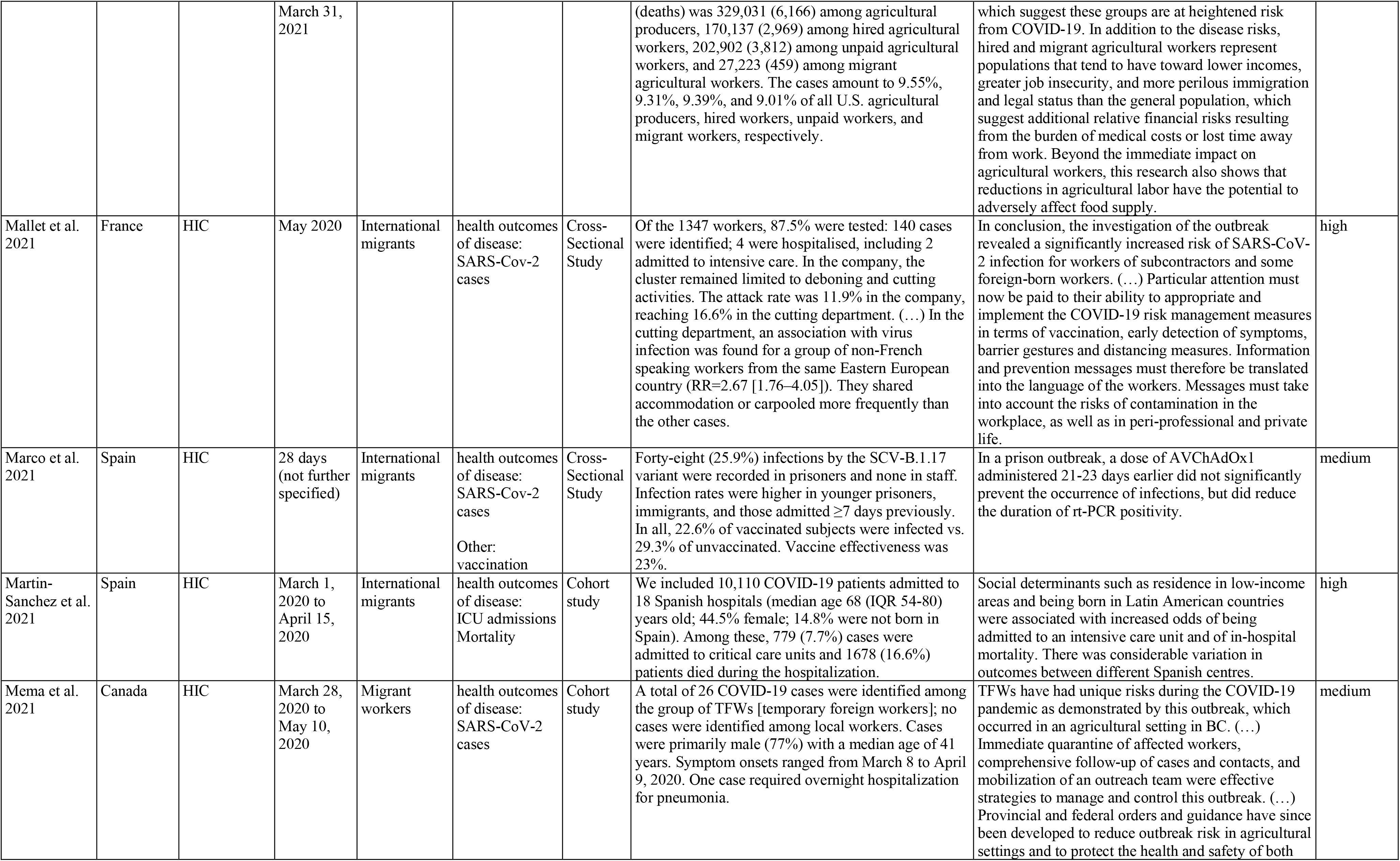
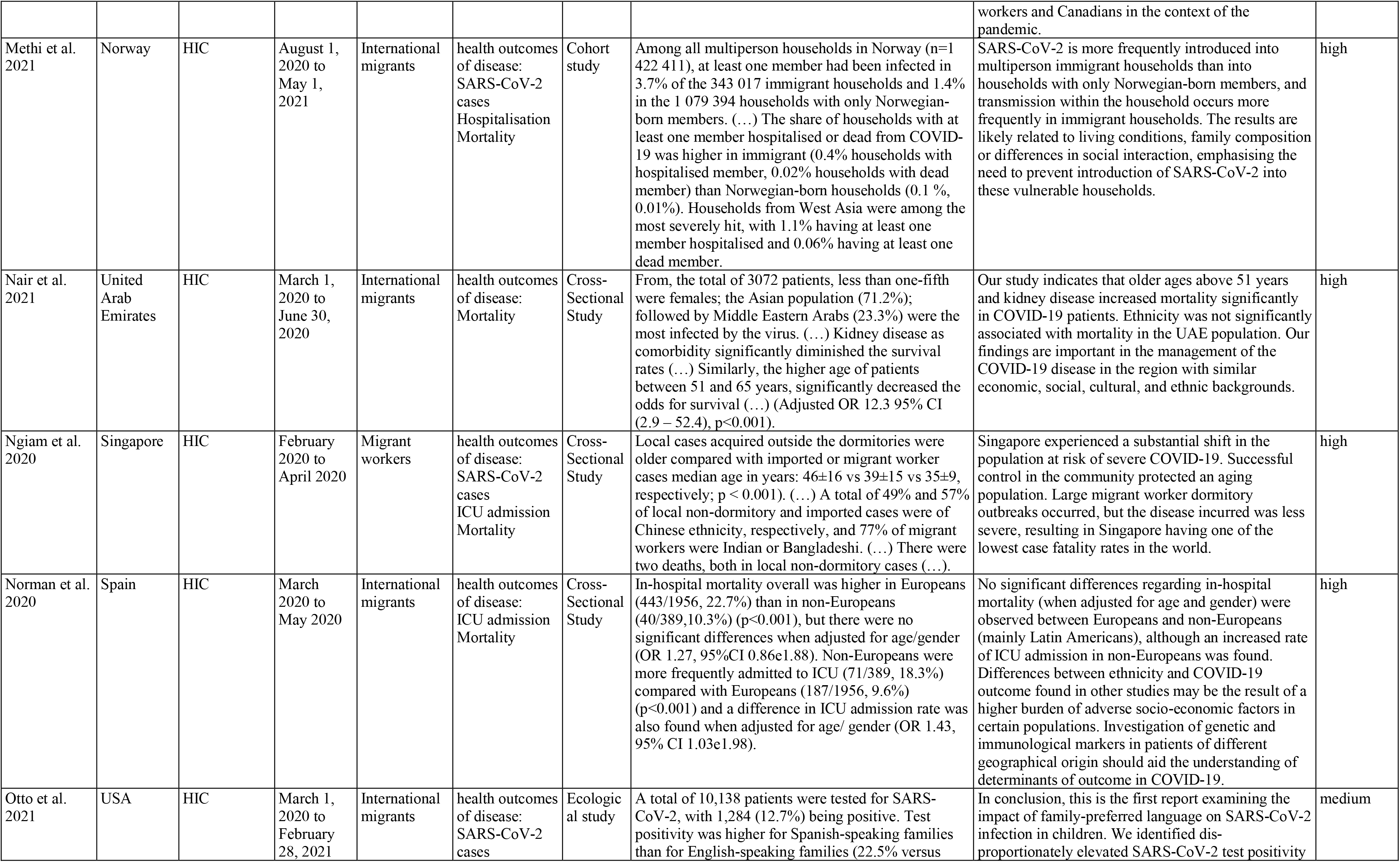
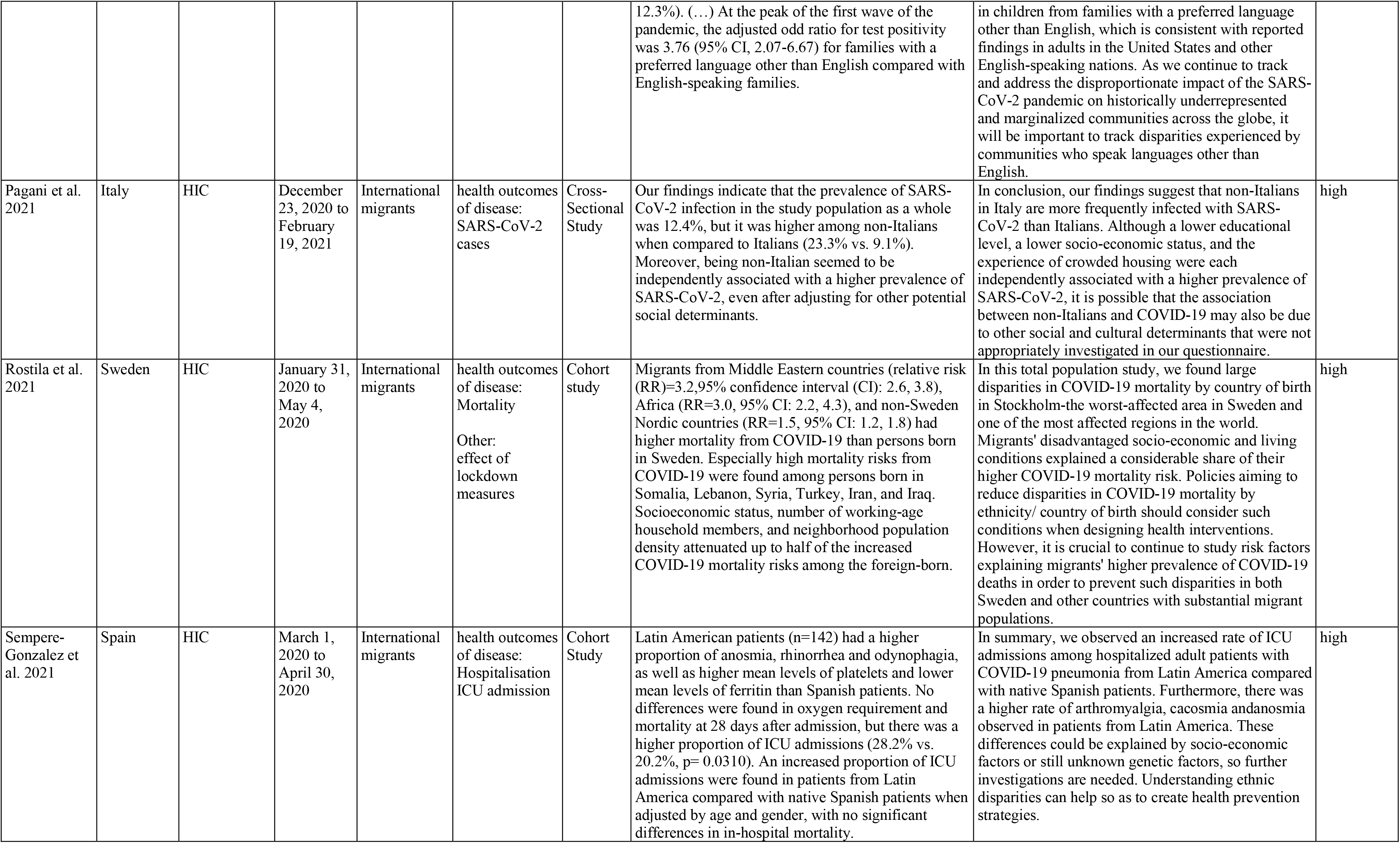
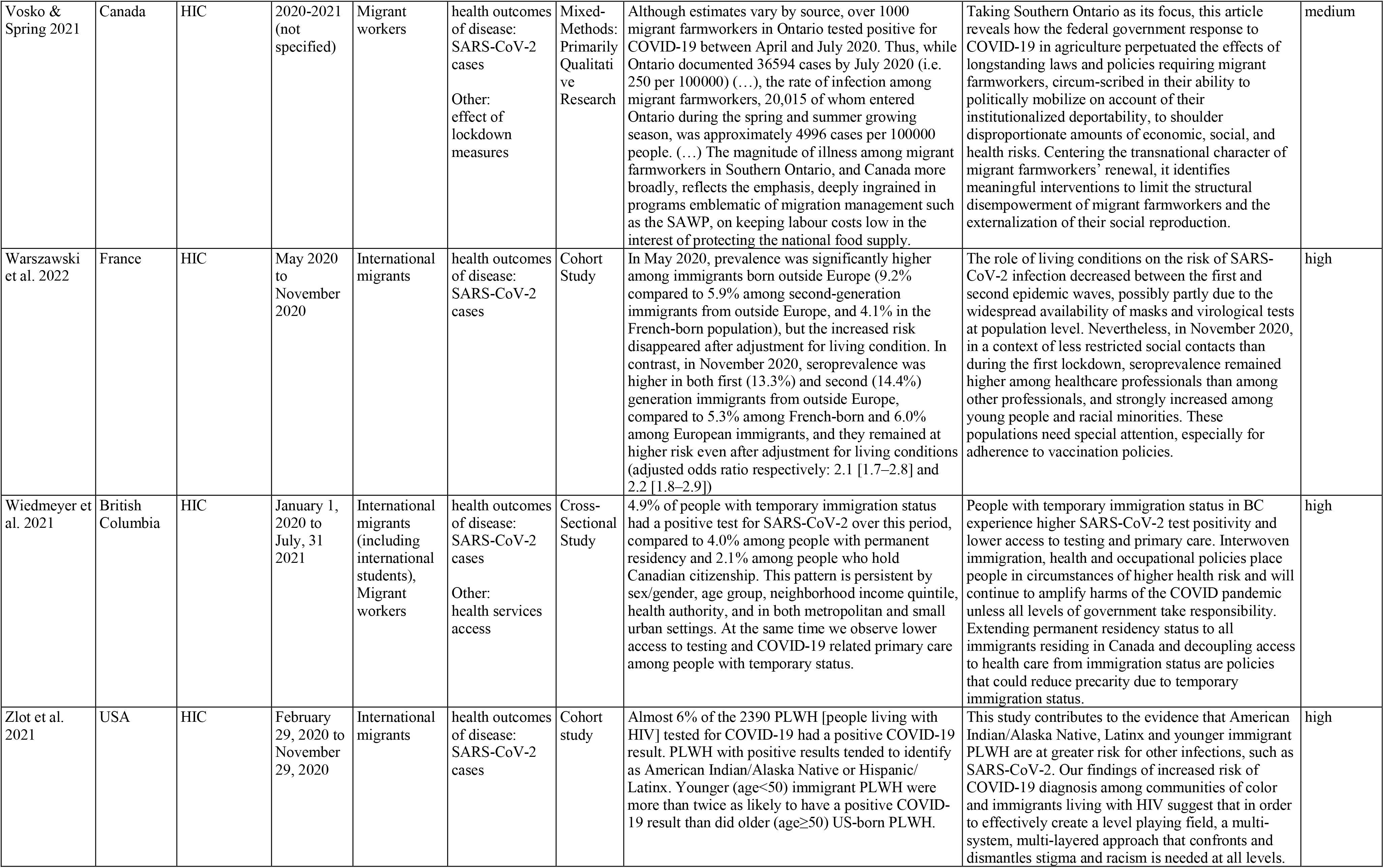
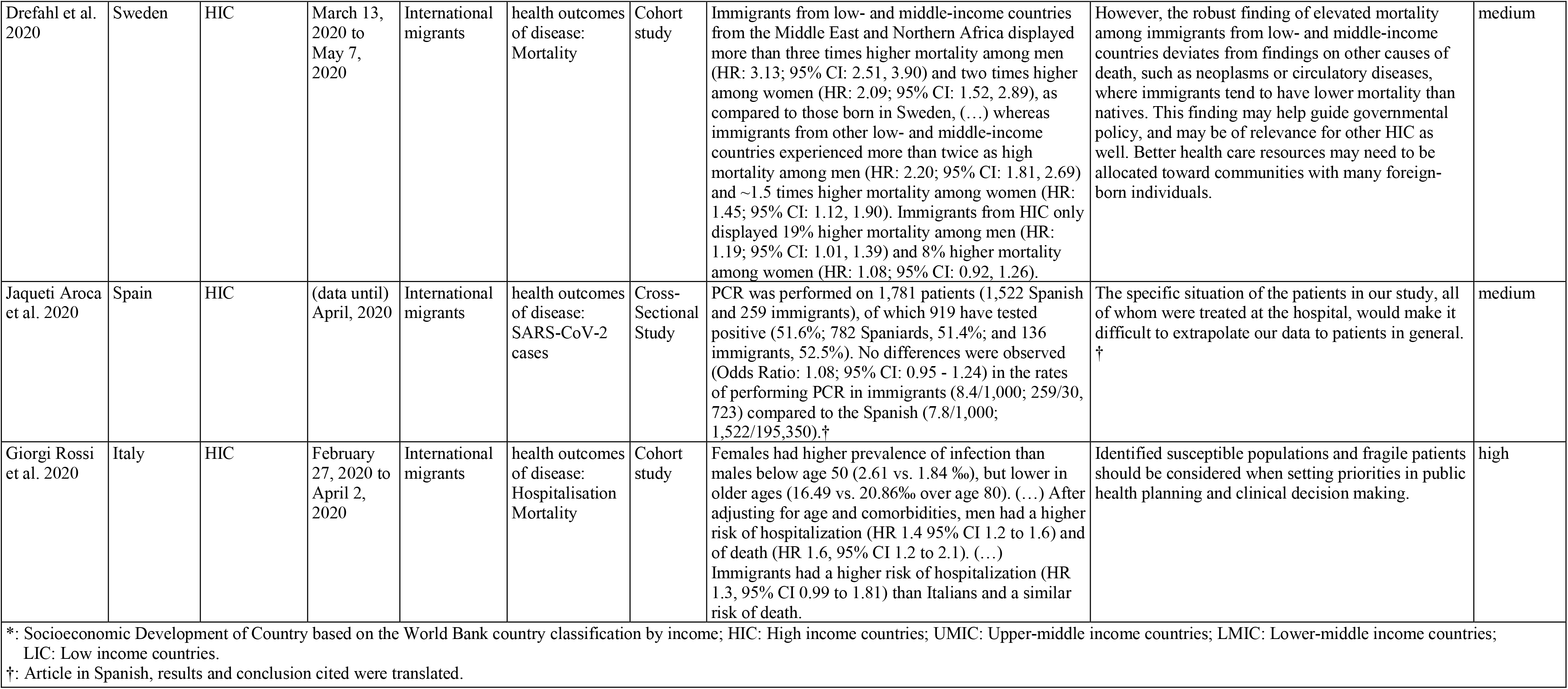
Characteristics of studies included in meta-analysis.

Based on the sensitivity analysis, few studies were not eligible for the quantitative synthesis and therefore excluded from the meta-analysis (Supplementary File, Chapter 6.1; pp.103-106). Among the 26 studies reporting on SARS-CoV-2 cases,^21–46^ 20 were eligible for quantitative synthesis.^21, 22, 24, 25, 27, 29–34, 36, 37, 39–42, 44–46^

Sample sizes comprised 40,019,567 individuals for cases, 1,310,219 for hospitalisation, 96,691 for ICU admission, and over five million for mortality. The *risk of infection* among migrants was 2·33 (95%-CI: 1·88-2·89) times the risk of non-migrants, with a RD of 7% (95%-CI: 0·02-0·13) (Fig. 2a). Inequalities in infection risk between migrants and non-migrants living in North America (RR = 2·65; 95%-CI: 1·34-5·26) or northern Europe (RR= 2·81; 95%-CI: 1·99-3·95) seemed to be higher than inequalities observed in southern Europe (RR = 1·70; 95%- CI: 1·23-2·35), whereby the actual RD compared to non-migrants was 9% (95%-CI: 0·01-0·17) in North America and 3-4% (95%-CI: 0·00-0·05; 0·01-0·08) in European countries (Fig. 2b).

**Figure 2a:**
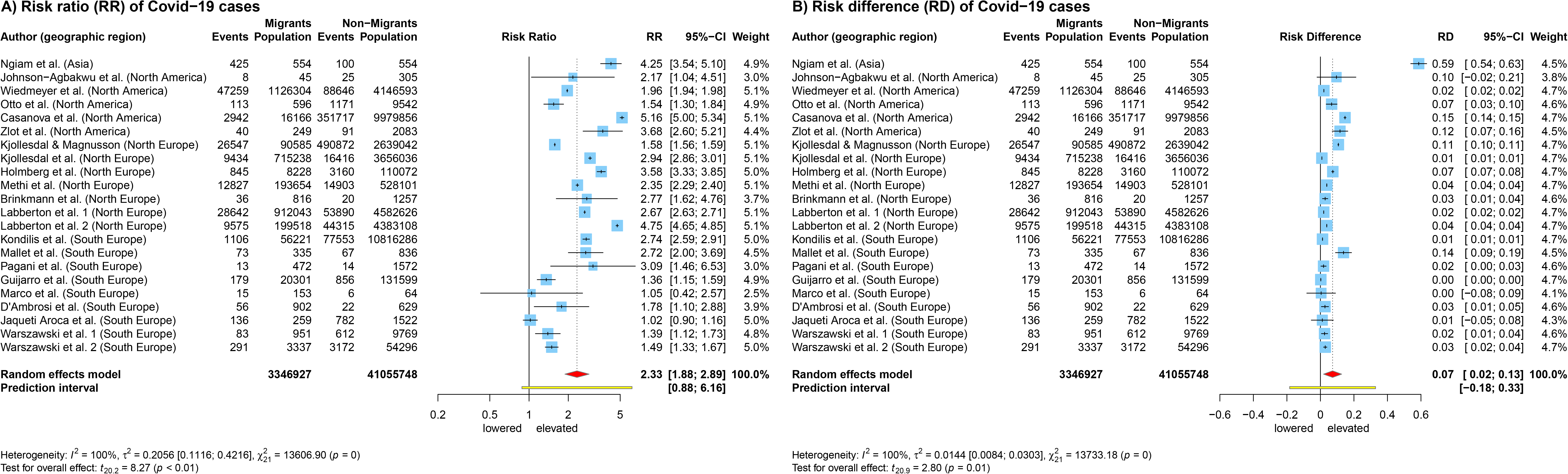
Forest Plot of Relative Risk (A) and Risk Difference (B) of SARS-CoV-2 cases between migrants and non-migrants. Legend: Events: SARS-CoV-2 cases; Population: denominators; CI: Confidence Interval; I2 and Tau2: Measures of heterogeneity. Labberton et al. grouped migrants into two subgroups (foreign-born/foreign-born parents), hence the study appears twice in the analysis.^34^ The same applies to Warszawski et al. who reported cases for May and November 2020.^46^

**Figure 2b:**
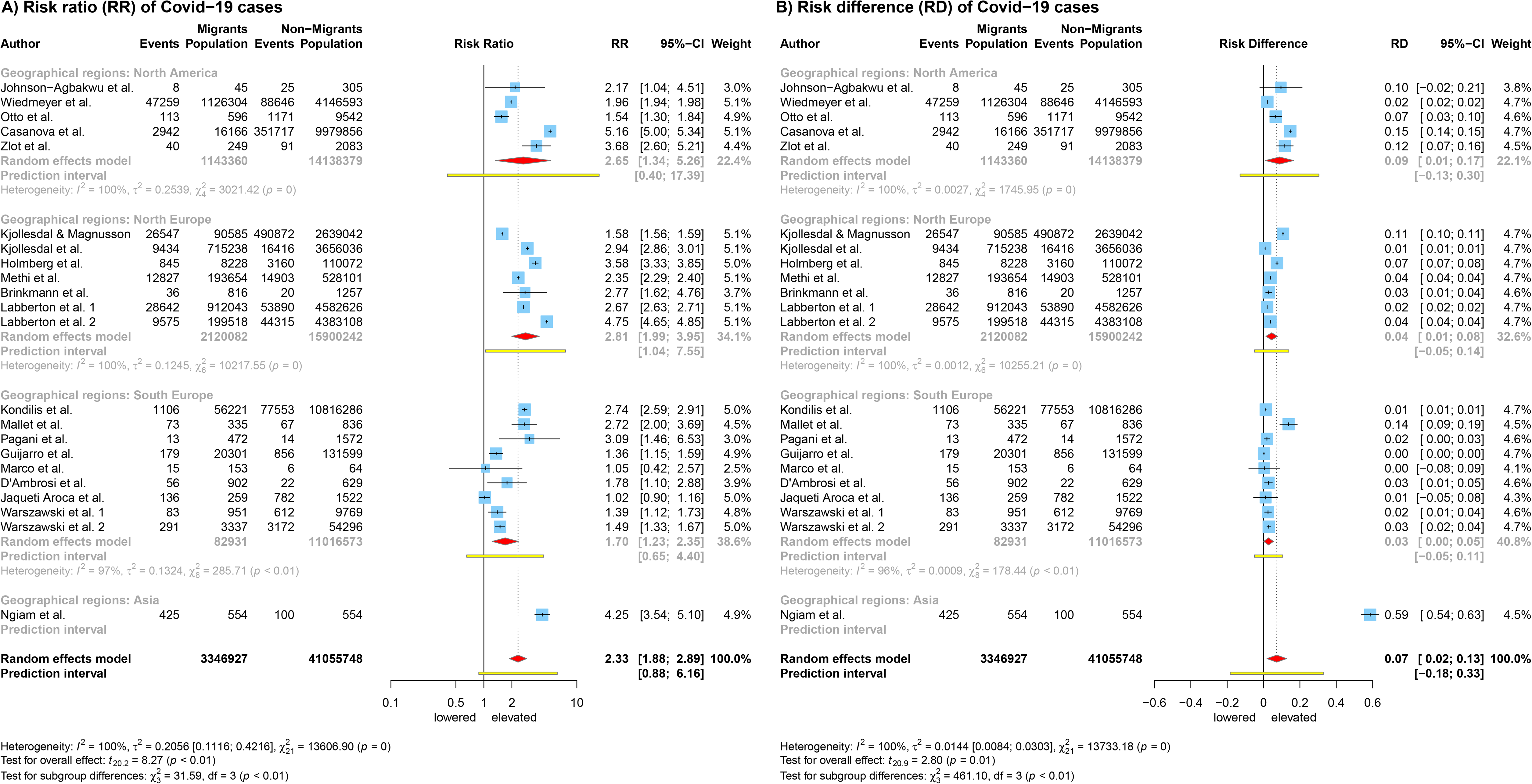
Forest Plot of Relative Risk (A) and Risk Difference (B) of SARS-CoV-2 cases between migrants and non-migrants by geographical region of study. Legend: Events: SARS-CoV-2 cases; Population: denominators; CI: Confidence Interval; I2 and Tau2: Measures of heterogeneity. Labberton et al. grouped migrants into two subgroups (foreign-born/foreign-born parents), hence the study appears twice in the analysis.^34^ The same applies to Warszawski et al. who reported cases for May and November 2020.^46^

International migrants turned out to be the main migrant category studied (i.e., 76% of studies), and were mostly defined by indicators such as region of origin, country of birth, language, or nationality. We grouped studies based on the underlying indicators of migratory status and provide an explorative subgroup analysis in the supplement (Figure S16, p.118). Among these groups, inequalities in infection risk compared to non-migrants were highest in migrants living in any kind of shared accommodation (refugee camp, dormitory, detention facility) (RR = 3·91; 95%-CI: 1·71-8·97), albeit with wide PI indicating high between study variations.^21, 33, 40^ Infection risk in migrants whose migratory status was defined by residence status, nationality of parents, country of birth, or region of origin was more than twice the risk of non-migrants (with narrow PIs), but inequalities were less pronounced among foreign workers and when migration status was defined via language (Supplementary File, Figure S16, p.118).

13 studies reported on *hospitalisation* of migrant and non-migrant SARS-CoV-2 infected individuals,^27, 28, 32, 34, 39, 47–54^ whereof nine studies were eligible for quantitative synthesis.^27, 28, 32, 34, 39, 47, 48, 51, 54^ Overall, the risk of hospitalisation appeared to be similar among both population groups (RR = 1·05; 95%-CI: 0·80-1·37) (Fig. 3). However, studies from southern Europe showed a lower risk in migrants compared to non-migrants,^51, 54^ whereas studies conducted in northern Europe reported slightly elevated risk among migrants (Fig. 3).^27, 28, 32, 34, 39, 48^

**Figure 3:**
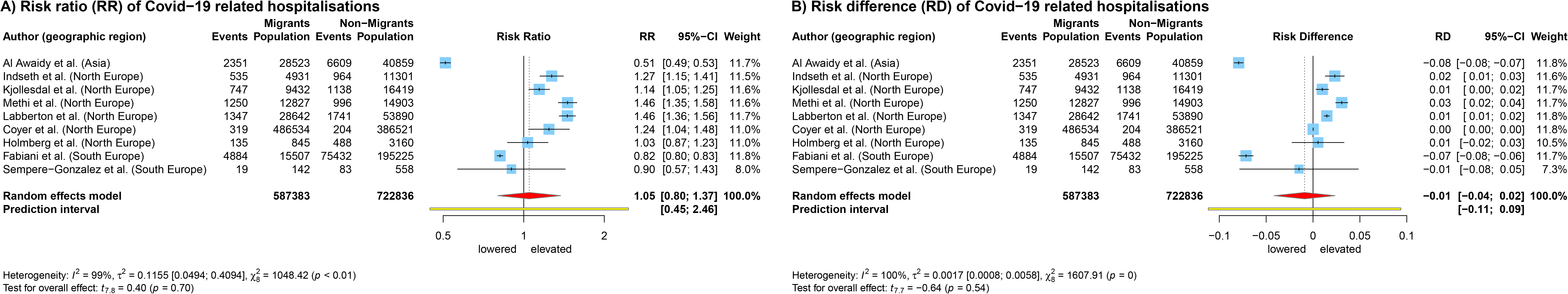
Forest Plot of Relative Risk (A) and Risk Difference (B) of hospitalised cases between migrants and non-migrants. Legend: Events: hospitalised cases; Population: denominators (SARS-CoV-2 cases); CI: Confidence Interval; I2 and Tau2: Measures of heterogeneity.

The meta-analysis of the outcome *ICU admission* included eight out of ten studies and showed 1·36 (95%-CI: 1·04-1·78) times the risk in migrants compared to non-migrants (Fig. 4). ^27, 40, 47, 51, 54–59^ The risk difference among the groups was relatively low (RD = 0·04; 95%-CI: -0·00-0·07) (Fig. 4).

**Figure 4:**
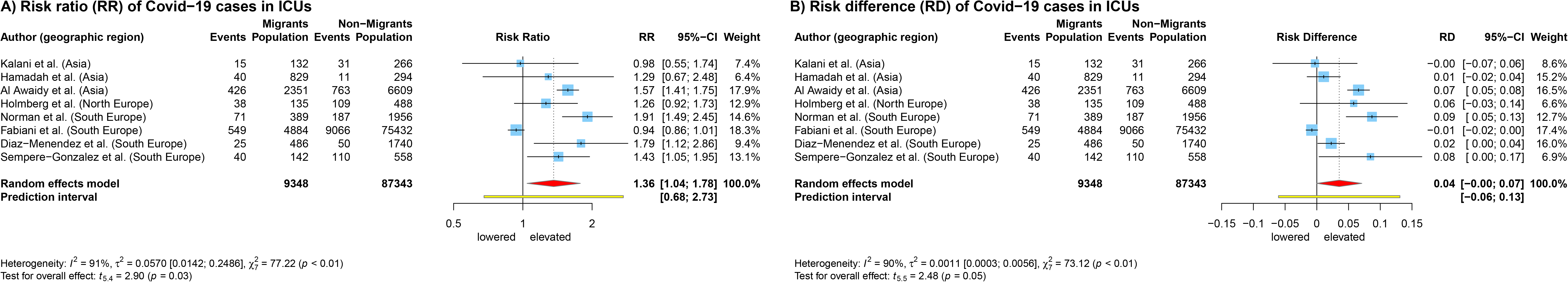
Forest Plot of Relative Risk (A) and Risk Difference (B) of ICU admissions between migrants and non-migrants. Legend: Events: ICU admissions; Population: denominators (hospitalised cases); CI: Confidence Interval; I2 and Tau2: Measures of heterogeneity.

As for *mortality*, 16 out of 18 studies were eligible for meta-analysis (Supplementary File, pp.112- 117).^27, 28, 39, 40, 47, 51, 53, 56–66^ Studies with mortality as outcome used different denominators: hospitalised cases vs. all-deaths within the respective time-period and geographic region. This resulted in different trends and patterns of inequality. With hospitalised cases as denominator for incident deaths, the mortality risk in migrants was almost half that of non-migrants (RR = 0·47; 95%-CI: 0·30-0·73) with a risk difference of -7% (95%-CI: -0·12-(-0·02)) (Fig. 5) based on a sample size of 22,561 study participants. If studies reported excess mortality (i.e. using all-deaths in the population), the risk for fatal outcomes due to or associated with SARS-CoV-2 was 1·31 (95%-CI: 0·95-1·80) the risk in migrants compared to non-migrants, however the difference between the groups was comparably low (RD = 0·03; 95%-CI: -0·02-0·09) (Fig. 5). For this analysis 5,062,317 study participants were included.

**Figure 5:**
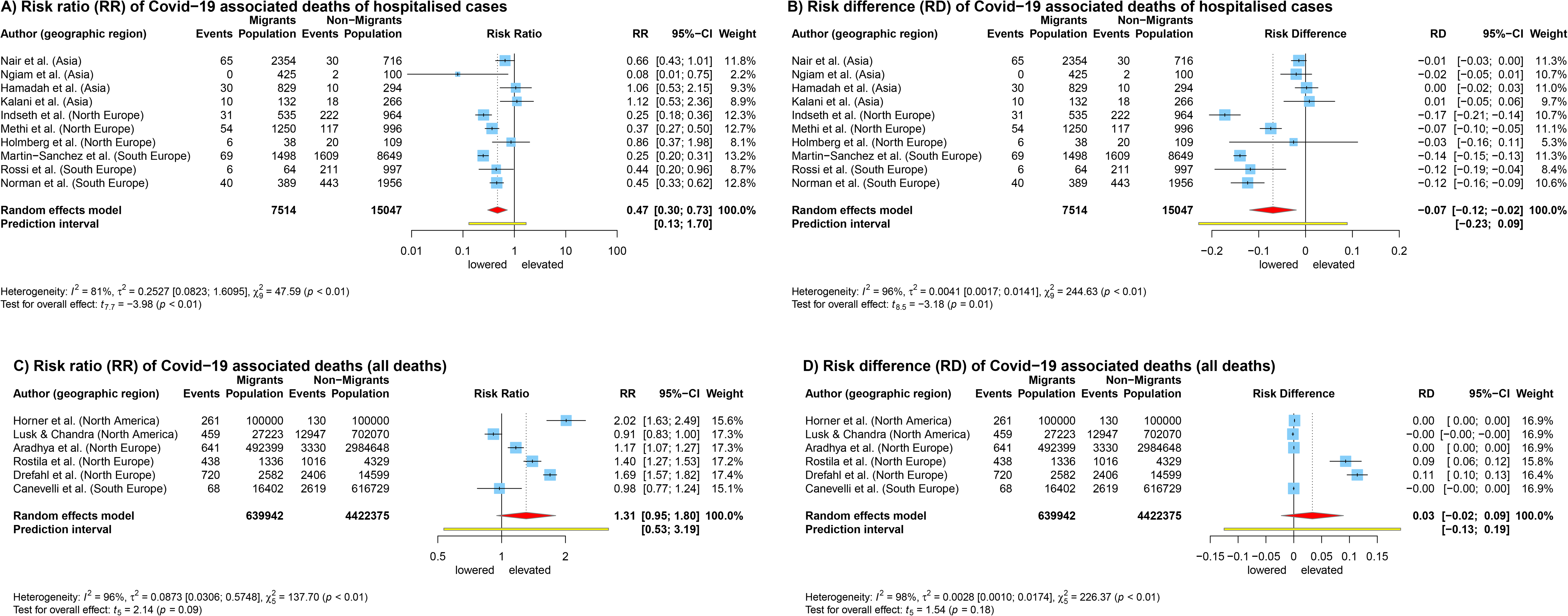
Forest Plot of Relative Risk and Risk Difference of mortality between migrants and non-migrants for hospitalised cases A) and B) and based on all deaths C) and D) Legend: Events: SARS-CoV-2-related deaths; Population: denominators (hospitalised cases A) and B); all deaths C) and D)); CI: Confidence Interval; I2 and Tau2: Measures of heterogeneity.

The *qualitative synthesis* was based on 44 high- to moderate-quality articles and provided insights into the syndemic nature of the COVID-19 pandemic by showing the complex interactions between social- and COVID- 19-related factors that have resulted in relative disadvantages for migrants.^43, 67–109^ Figure 6 illustrates exposures, risks, and impact of COVID-19 measures as well as sources of resilience for migrant populations, each at micro-, meso-, macro-levels (i.e., at the level of the individual, the family/community, and state/society, including policies and institutions such as the healthcare system or the labour market). Factors in the different categories and on the different levels interacted and compromised the physical and psychosocial health of migrants in severe and sometimes unique ways.

**Figure 6:**
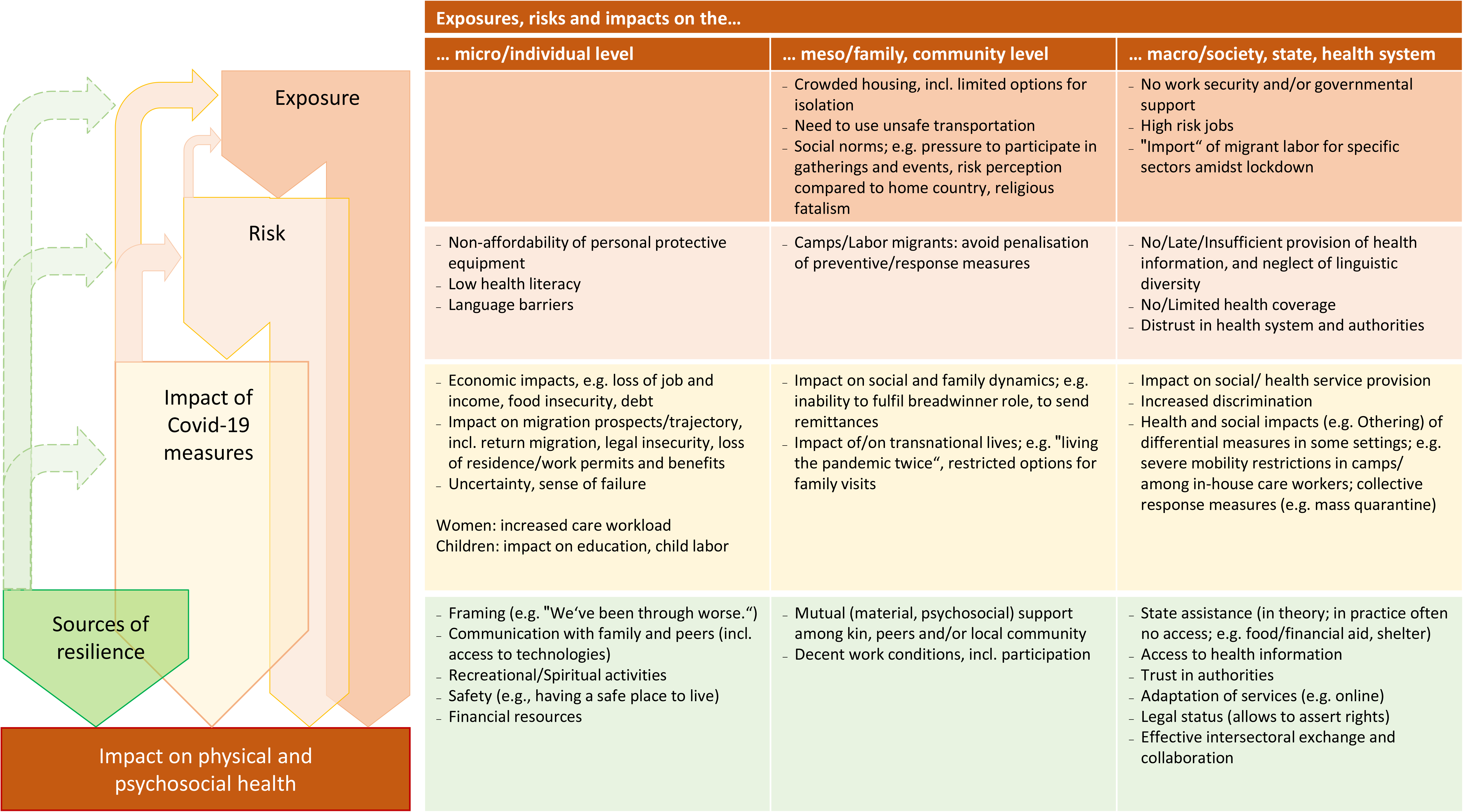
Risks and impacts related to Covid-19 and pandemic measures for migrants: A summary of qualitative research findings.

Our synthesis shows that migrants faced exposures at meso- and macro-level; among these crowded housing conditions, and lack of work security, and/or governmental support emerged as particularly critical.^43, 67, 77, 103, 108^ Some studies further pointed to the role of social norms in creating exposures, describing e.g. that some *(more communitarian)* communities may tend to put pressure on their members to participate in gatherings and events. Risks (e.g., higher risk of infection) existed at all three levels. At the individual level, the included studies identified unaffordability of personal protective equipment, low health literacy and language barriers.^67, 74, 79^ These risk factors can be cross-referenced to the macro-level, as other studies reported late, insufficient or no provision of health information in ways that accommodate linguistic diversity formal barriers to health services and distrust in institutions, including public health services.^67, 79^ Risks at the meso-level include the (threat of) penalisation of preventive and response measures by employers and camp managers, which may deter migrants, e.g., from demanding protective equipment or testing.

Among the impacts of the COVID-19 pandemic and pandemic measures, three interrelated individual-level factors figured prominently across the literature: economic consequences of lockdown measures (such as job loss), impacts on migration prospects, and trajectories (mainly related to disruption of migration related services such as visas and work permit renewal or the processing of asylum applications), which put migrants in states of legal and social insecurity, and, as a corollary, uncertainties and fears of failing one’s migration project.^68, 71, 73, 75, 77, 80, 83, 88, 90, 95, 99^ These factors are intertwined with meso-level factors: e.g., the inability to fulfil social roles (such as acting as the family’s breadwinner, sending remittances), impacted social and family dynamics, including gender and intergenerational roles).^68, 75, 80, 85, 89, 90, 102^ Further, impacts on the macro-level include increased discrimination and, in some settings, the effects of differential pandemic measures. For instance, migrants in camp-like settings (e.g. asylum-seekers, refugees, labour migrants in congregate housing) as well as in-house domestic workers were often subjected to severe mobility restrictions; also in many camp-like settings, collective response measures such as mass quarantine or labour quarantine were imposed on the residents. Our analysis thus highlights the severe, to some extent unique, and in part unintended consequences of pandemic control measures for migrants.^69, 71, 72, 80, 81, 90, 94, 104^ More than the pandemic itself, these unintended consequences of pandemic measures have contributed significantly to the severe psychosocial impact on migrants.^69, 84^

Our analysis pinpoints sources of individual, community and systemic resilience that can counteract some of the above described exposures, risks and impacts of pandemic measures. Among the main individual-level sources of resilience are optimistic framings of the crisis (e.g., “We’ve been through worse.”) and the exchange with family and peers via internet and social media. Mutual (material and/ or psychosocial) support as well as decent work conditions that allow for a sense of control and participation are important community-level sources of resilience. Macro-level sources of resilience include access to governmental assistance, the tailored provision of health information, trust in the authorities, and legal status, which is key to being able to assert one’s rights. Effective intersectoral exchange and collaboration enhance health system resilience. However, our analysis also shows that many of these sources of resilience have been inaccessible or unavailable for migrants during the COVID-19 pandemic, especially those on meso- and macro-levels.^43, 77^

Eventually, our analysis highlights the multifaceted interrelations among the various factors in the different categories and on the different levels, with some key factors triggering cascading effects and feedback loops (Panel 4).

### Panel 4: Cascading effects and feedback loops for different migrant groups deriving from pandemic control measures

#### Vignette a) “Migrants with precarious legal status”

The pandemic-related closure of offices and services obstructed the renewal of residence and work permits for migrants with precarious legal status. Informal labour markets were severely impacted by pandemic measures, leading to widespread loss of jobs and livelihoods, with no social safety nets in place. The resulting legal and economic insecurities amplified existing power differentials, put migrants at risk of exploitation, and generated major psychosocial distress due to the uncertain future prospects in the host country. The pressure to send money to families and communities in their home countries exacerbated such distress. The coping strategies of migrants and their families sometimes involved extremely hazardous employment, including child labour and survival prostitution. This, in turn, increased the exposure to and risk of a SARS-CoV-2 infection, but also other health issues such as injuries, sexually transmittable diseases, violence, unwanted pregnancies, and ill mental health. Adverse social consequences include descent into extreme poverty, food insecurity, impacts on children’s education, stigma, and tensions within families over changing gender and intergenerational roles. Access to social and health services, including tailored services such as walk-in clinics for uninsured patients, were also compromised due to the pandemic. The only available source of material and psychosocial support were often informal networks based on kinship, diaspora and/or religious communities.

*(Synthesis based on Babuc 2021, Da Mosto et al. 2021, Knights et al. 2021, Filippi & Giliberti 2021, Im & George 2021, Martuscelli 2021, Sabar et al. 2021, Sanò & Della Puppa 2021, Sohel et al. 2021, Thomas et al. 2021, Yee et al. 2021, Zambrano-Barragan et al. 2021)*

#### Vignette b): “Migrant domestic care workers”

Domestic care workers faced particularly strict pandemic measures and related changes in their working conditions: With their work permit and visa often bound to a specific client/employer, many had to accept severe mobility restrictions, namely cohabitation with their client in combination with strict curfews, and increased workloads (e.g. more cleaning and washing), alongside obligatory quarantine and testing measures, in order to keep their job. Cohabitation with the client meant a loss of personal freedom and privacy, longer working hours, constant availability, less rest and breaks, and more stress. Non-compliance with health and safety protection such as a lack of protective equipment or of paid sick leave was frequently noted. This increased health risks for domestic workers, including burnout and other mental health problems. Domestic workers were very concerned about being terminated due to a SARS-CoV-2 infection or for other health reasons, and in some studies they describe their work situation during the pandemic as a “prison” or as being trapped in a climate of constant control, abuse and fear, with no options to leave an employer without also losing their work permit and visa.

In studies which described their situation in positive terms (e.g., as having a safe job and income and a safe place to stay), such framings depended on the quality of the relationship with the employer/client, and decent employment practices (e.g., ensuring the worker’s day off and participation in familial decision-making). Being able to communicate with the family back home during the pandemic, e.g. via social media, was described as another source of resilience – and at the same time as potential stressor. Many migrant domestic care workers continued being responsible for their (left behind) families in the home country. This double responsibility of care translated into a “dual-country experience of the pandemic”; i.e. worries for the health and well-being of the family back home, increased pressure to send remittances, alongside the (often sole) responsibility for the care and health of the client in the host country.

*(Synthesis based on: de Diego-Cordero et al. 2022, Lui et al. 2021, Kaur-Gill et al. 2021, Kuhlmann et al. 2020, Nasol & Francisco-Menchavez 2021, Sabar et al. 2021, Sanna 2021)*

Among all 241 studies (Supplementary File, Table S6; p. 35ff) we identified seven studies (2·9% of all studies) that investigated vaccination coverage among migrants either empirically or anticipated by statistical models.^37, 78, 110–114^ Two modelling studies with medium and low quality recommended to include migrants in the national vaccination strategy to prevent COVID-19 incidence and ensure cost-effectiveness.^111, 113^ One medium quality study reported vaccination coverage within a Spanish prison but focussed on vaccination effectiveness of the respective vaccine.^37^ Three studies of high and medium quality from the European context reported lower vaccination rates among migrants (i.e., foreign-born) in comparison to the non-foreign-born population.^78, 110, 112^ The opposite, namely higher vaccination uptake among migrants compared to local residents was found in a medium quality study conducted in Nanjing and Chizhou, China.^114^

## DISCUSSION

Our meta-analysis quantified inequalities between migrants and non-migrants in risk of COVID-19 infection, hospitalisation, ICU-admission, and death. We found that the incidence risk was more than twice the risk among migrants than among non-migrants, and was about seven percentage-points higher on average. Inequalities tended to be more pronounced in Northern Europe and North America, and lower in Southern Europe. These differences could be related to differences in access to health systems (e.g., based on legal status), in testing policies and management of SARS-CoV-2 cases, or in environmental risks (e.g., occupation and accommodation). Migrants in camps or other forms of institutionalised/shared accommodation appeared to be at highest risk of infection, albeit PIs for the subgroup analysis by migrant categories were extremely broad and overlapped (indicating high heterogeneity between studies within subgroups), so that differences should be interpreted with care.

We found no evidence for inequalities in risk of hospitalisation, which stands in contrast to inequalities observed between ethnic minorities and majority populations, indicating that different mechanisms are at play in the pathway between health and ethnicity versus migration.^115^ However, our meta-analysis indicates that, once infected and hospitalised, migrants may have experienced more severe courses of disease: the risk of being admitted to ICU was 36% higher in relative, and 4 percentage-points higher in absolute terms among migrants compared to non-migrants (albeit with broad PIs). While the share of migrants in deaths among hospitalised cases was lower (likely due to age differences), their share among deaths at population-level measured by excess-mortality was 31% higher in relative, and three percentage-points higher in absolute terms, compared to resident populations in respective studies.

Intersections among COVID-19-related exposures, risks, and impacts of pandemic measures in migrant populations compromised their physical and mental health in severe and sometimes unique ways. Cascading effects and feedback loops became evident, highlighting the syndemic nature of the COVID-19 pandemic.^116, 117^ Yet, we also found (potential) sources of resilience, indicating entry points for measurements (e.g., support from non-governmental organisations, peers or the local community) to improve migrants’ conditions in health emergencies. Overall, the qualitative results highlight the key role of socio-economic and work-related precarity, in combination with legal precarity.^67, 86, 91^ This is further underscored by studies, which describe that different States (i.e., Austria, Canada, and Italy) approved special policies to recruit migrant workers amid lockdown to work as fruit pickers and in other agricultural sectors, often in precarious labour contexts.^43, 86, 98^ Asylum seekers and other undocumented migrants were also absorbed in “essential” sectors to meet labour shortages.^118^ These examples illustrate the reliance of global economies of production on certain migrants as a captive and cheap workforce. And at the same time the COVID-19 pandemic has also shown that the same business models systematically shift the social costs of labour, including health risks, to the migrant workers themselves.^119^ The qualitative results thus underscore the importance of a political economy-perspective to understand the structures that generate and sustain social and health inequities for migrants. Sub-regional and regional governance structures that promote trade and mobility for economic cooperation, need to ensure migrant health as a key requisite of development. There is a critical need for governments, international organisations, civil society and migrant communities to engage with private sector, employers and industries in which migrant workers are overrepresented in precarious work contexts, with low-wages and with an irregular status. The underutilisation of the domain of work, including occupational health programs, in addressing the health of migrants is indeed truly a missed opportunity for global health.^120^

We found a high degree of heterogeneity among studies regarding the design, migrant populations, quality of data, and countries of studies. Poor reporting in primary studies raised considerable challenges in meta-analysis. This was most pronounced for mortality due to the use of different denominators: hospitalised SARS-CoV-2 cases or all-cause deaths in the total population. These two ways of reporting mortality have been identified by Karanikolos and McKee as a factor which substantially limits the comparability of COVID-19-related mortality between countries.^121^ Our review underlines the need for harmonised reporting in future health emergencies to ensure comparability of mortality estimates across countries and studies. The trend to lower mortality in hospitalised cases is dependent on testing strategies, and adjustment for age and comorbidities, which was not always performed in primary studies yielding estimates prone to confounding. In contrast, excess rates of age- and sex-standardised mortality are more likely to reflect the true mortality risks. In view of this, our results show that migrants tended to be at considerably higher risk of death associated with COVID-19 than non-migrants during the pandemic (until 11/2021).

The strength of this review is the synthesis of all (high to moderate quality) evidence from qualitative and quantitative studies on health-related impacts of the COVID-19 pandemic on migrants worldwide. The analysis of the interconnectedness of exposure, risk, and impact of pandemic measures at micro-, meso-, and macro-levels benefitted from qualitative research, which captures social phenomena in their complexity and within their particular context, thereby allowing us to contextualise findings. Despite an overwhelmingly broad landscape of literature, however, our knowledge of some groups and migration phases still remains scarce and sketchy, for example, related to labour migrants, undocumented migrants, migrants with disabilities, elderly migrants, returnees, migrants in transit or in pre-migration phases. Countries of studies were mainly destination countries so that evidence is skewed toward COVID-19-related migrant health in the *post-migration* phase. Further efforts are required to better reflect health-related aspects of the complex migration trajectory.

The meta-analysis provides valuable evidence of the magnitude of COVID-19-related inequalities, despite being limited by the striking heterogeneity of underlying primary studies. Cautious interpretation is required, however, due to poor quality of reporting in primary quantitative studies. We could not adjust the RR for age and sex because the data were either not reported or even unavailable to the authors of primary studies upon request. This highlights the urgent need to improve the reporting quality and primary data in studies presenting migration-stratified outcomes. The broad search enabled to include studies from different research fields, but this posed a challenge to the applicability of the quality appraisal tools. For 15 studies, only a few questions of the JBI checklists were applicable. We included English, German and Spanish articles, so that other languages, e.g., French, Chinese or Arabic, could not be considered and thus studies from respective countries may not have been included.

Our review sheds a glaring light on the lack of evidence on important measures of pandemic response, such as vaccination coverage among migrants, not only until the end of 2021. Updating our search in the WHO COVID- 19 research database on vaccination coverage among migrants for the years 2022 and 2023 resulted in 204 hits (April 2023). Of these, only about 15 studies addressing this topic could be identified after screening titles to report vaccination coverage/uptake/rates among the respective migrant populations. As with the search for this review, the focus of the research appears to be on *vaccination hesitancy*, *knowledge* and *attitudes* toward COVID-19 vaccination, or different *ethnic* groups, rather than on *coverage rates* in migrants as defined previously. The scarcity and poor quality of reporting of studies points to the weakness of Health Information Systems (HIS) to provide reliable data on key aspects of migrant health such as vaccination coverage, by the end of 2021, and even in 2023. The lack of harmonisation across different migrant categories and outcome measures puts validity and comparability of findings at risk. Our findings hence call for urgent implementation of recommendations and WHO technical guidance to enhance the integration and availability of migrant indicators in HIS.^122, 123^ This requires urgent action in the post-COVID-19 recovery phase to bridge the divides between health and migration governance by means of effective collaboration structures at all levels of government (local, national, international).^124^ Monitoring migrant health should be considered an essential component of pandemic preparedness, and future national plans must secure an adequate inclusion of migrant groups which promote social and health equity. This includes anticipation and prevention of unequal effects or unintended (negative) consequences of the pandemic on migrant health that are exacerbated by language barriers, stigma and discrimination, and by financial, administrative and legal barriers to health systems.

## Contributors

MH: Conceptualization, Methodology, Investigation, Data curation, Formal analysis – statistical analysis, Writing – Original Draft, Visualization, Project administration

NG: Investigation, Formal analysis – qualitative analysis, Data curation, Visualization, Writing –Review & Editing

SR: Methodology, Investigation, Formal analysis – statistical analysis, Data curation, Visualization, Writing – Review & Editing

JO: Investigation, Data curation, Formal analysis, Visualization, Writing – Review & Editing MB: Investigation, Formal analysis – qualitative analysis, Writing – Review & Editing

SA: Methodology Investigation, Formal analysis – qualitative analysis, Writing – Review & Editing

JL: Methodology, Investigation, Formal analysis – qualitative analysis, Writing – Review & Editing

SF: Investigation, Writing – Review & Editing

AM: Investigation, Writing – Review & Editing

KW: Conceived the study, Conceptualization, Methodology, Writing – Review & Editing

KB: Conceived the study, Conceptualization, Methodology, Investigation, Formal analysis, Validation, Writing – Review & Editing, Supervision, Funding acquisition

All authors had full access to all data in the study and had final responsibility for the decision to submit for publication.

## Data sharing

A full list of studies identified in the search as well as the full data extracted from included studies are available for academic research projects by request to the corresponding author

## Acknowledgments

No funding received.

