## Supplementary File for "COVID-19 among migrants, refugees, and internally displaced persons: systematic review, meta-analysis and qualitative synthesis of the global empirical literature"

**Content of supplementary appendix**

6.4.2 Denominator: hospitalised cases ... 115

Figure S13: Mortality – hospitalised cases. Model diagnostics – Standardised residuals, Cook's Distance, Hat value ... 115

Figure S14: Mortality – hospitalised cases. Model diagnostics – LOO-CV ... 116

Table S10: Mortality – hospitalised cases. Modelling results ... 116

Figure S15: Mortality – hospitalised cases. Funnel plot ... 117

7 Subgroup analysis by migrant indicators for SARS-CoV-2 Cases ... 118

Figure S16: SARS-CoV-2 Cases. Subgroup analysis by migrant indicator ... 118

8 PRISMA Checklist ... 119

Table S11a: PRISMA-Checklist for Abstracts ... 119

Table S11b: PRISMA-Checklist ... 119

9 References ... 122

### 1 Search queries by database

Tables S1 and S2 contain the search strings applied to the WHO Covid-19 database and the Cochrane Library, respectively.

**Table S1: WHO COVID-19 database - 02.12.2021**

| ID | Query | Results |
|---|---|---|
| #3 | #1 AND #2 | 6928 |
| #2 | (DA:202006 OR DA:202007 OR A:202008 OR DA:202009 OR A:202010 OR DA:202011 OR DA:202012 OR DA:2021$) | 286,365 |
| #1 | (tw:(refugee OR "asylum seeker" OR asyl* OR (displaced AND (person* OR people)) OR migra* OR (forced AND migra*) OR migration OR immigra* OR "human migration" OR stateless OR "state-less" OR (irregular AND migra*) OR (undocumented AND migra*) OR "internally displaced" OR detainees OR "residence status" OR "foreign-born" OR "displaced person" OR noncitizen OR outsider OR newcomer OR "newly arrived" OR "new arrival" OR "recent entrant" OR "non national" OR "non-national" OR transient OR minoritie* OR ethnic OR "Transients and Migrants")) | 9237 |

**Table S2: The Cochrane Library - 02.12.2021**

| ID | Query | Results |
|---|---|---|
| #13 | #12 publication limit 01.06.2020 to 31.12.2021 | 117 |
| #12 | #5 AND #10 | 123 |
| #11 | #6 OR #7 OR #8 OR #9 | 28167 |
| #10 | refugee:ti,ab,kw OR “asylum seeker”:ti,ab,kw OR “asyl*”:ti,ab,kw OR (displaced:ti,ab,kw AND (person*:ti,ab,kw OR people:ti,ab,kw)) OR “migra*”:ti,ab,kw OR “forced migra*”:ti,ab,kw OR migration:ti,ab,kw OR immigra*:ti,ab,kw OR “human migration”:ti,ab,kw OR stateless:ti,ab,kw OR “state-less”:ti,ab,kw OR “irregular migra*”:ti,ab,kw OR “regular migra*”:ti,ab,kw OR “undocumented migra*”:ti,ab,kw OR “internally displaced”:ti,ab,kw OR “detainees”:ti,ab,kw OR “residence status”:ti,ab,kw OR “foreign-born”:ti,ab,kw OR “ displaced person”:ti,ab,kw OR “noncitizen”:ti,ab,kw OR “outsider”:ti,ab,kw OR “newcomer”:ti,ab,kw OR “newly arrived”:ti,ab,kw OR “new arrival”:ti,ab,kw OR “recent entrant”:ti,ab,kw OR “non national”:ti,ab,kw OR “non-national”:ti,ab,kw OR “transient”:ti,ab,kw OR “minorities”:ti,ab,kw OR “ethnic”:ti,ab,kw | 28092 |
| #9 | MeSH descriptor: [Refugees] explode all trees | 144 |
| #8 | MeSH descriptor: [Human Migration] explode all trees | 66 |
| #7 | MeSH descriptor: [Transients and Migrants] explode all trees | 73 |
| #6 | #1 OR #2 OR #3 OR #4 | 8504 |
| #5 | ("corona virus":ti,ab,kw OR "corona viruses":ti,ab,kw OR coronavir*:ti,ab,kw OR coronovirus*:ti,ab,kw OR betacoronavirus*:ti,ab,kw) AND (novel:ti,ab,kw OR 2019:ti,ab,kw OR Wuhan:ti,ab,kw OR Huanan:ti,ab,kw OR Hubei:ti,ab,kw) OR "new coronavirus":ti,ab,kw OR "COVID-19":ti,ab,kw OR COVID19:ti,ab,kw OR "SARS coronavirus 2":ti,ab,kw OR "severe acute respiratory syndrome coronavirus 2":ti,ab,kw OR nCoV:ti,ab,kw OR 2019nCoV:ti,ab,kw OR nCoV2019:ti,ab,kw OR "SARS-CoV-2":ti,ab,kw OR "SARS-CoV2":ti,ab,kw OR SARSCoV19:ti,ab,kw OR SARS-CoV19:ti,ab,kw OR SARS-CoV-19:ti,ab,kw OR HCoV-19:ti,ab,kw OR WN-CoV:ti,ab,kw | 8458 |
| #4 | MeSH descriptor: [SARS-CoV-2] explode all trees | 599 |
| #3 | MeSH descriptor: [COVID-19] explode all trees | 918 |
| #2 | MeSH descriptor: [Coronavirus] explode all trees | 612 |
| #1 | MeSH descriptor: [Coronavirus Infections] explode all trees | 1463 |

### 2 Inclusion and Exclusion criteria

Table S3 shows the a priori defined inclusion and exclusion criteria for study selection.

**Table S3: Inclusion and Exclusion criteria**

| Criteria | Inclusion | Exclusion |
|---|---|---|
| **Type of Population** | Refugees, asylum seekers, internally displaced people (IDP), and migrants as defined by the International Organisation for Migration (IOM).[1] | Unclear populations, ethnic minorities or (in full-text screening) reference to migrants, refugees or IDPs *without* stratified results (i.e., reporting migrants, refugees, or IDPs as part of a broader, undefined population including non-migrant groups). |
| **Type of studies** | • Empirical quantitative studies (cross-sectional, case-control, cohort, intervention studies, ecological studies, modelling studies, epidemiological outbreak reports).<br>• Empirical qualitative studies (interviews, focus groups, ethnography) analysing health outcomes or impact of lock-down measures and evaluation strategies that include refugees and migrants.<br>• Mixed-methods studies where qualitative and quantitative parts, where reported separately.<br>• Rapid or systematic reviews: are screened for their primary articles; meta-analyses are to be included on their own.<br>• Policy analysis with empirical data. | Case series, theoretical research work; Policy analysis without empirical data. |
| **Type of articles** | • Peer-reviewed articles<br>• Preprints,<br>• Official reports from IOM or EUPHA websites. | • Grey literature except for official reports on mentioned websites;<br>• Book sections;<br>• Commentaries and Letters to the Editor, unless reporting results of a study in a transparent scientific structure (IMRaD). This is to account for the pandemic/emergency situation in which relevant scientific findings are sometimes reported in form of Letters to the Editors.<br>• Pure opinion pieces that draw upon data (without transparent description of methods) are to be excluded.<br>• Mere conference abstracts/posters (or similar) without full-text. |
| **Focus of study** | • Studies reporting on SARS-CoV-2 epidemiology (infection, disease), course of disease (hospitalization, ICU) or outcome of disease (mortality).<br>• Studies reporting effects of lock-down measures, also on non-Covid health outcomes.<br>• Studies reporting effects of policies/strategies specifically enacted or implemented for refugees and migrants.<br>• Studies reporting on vaccination coverage or inequalities in coverage compared to a reference group. | Studies with:<br>• no focus on SARS-CoV-2 or<br>• no reference to health outcomes. |
| **Type of outcome measure** | • Measures of frequency (absolute numbers, proportions, incidence proportion, prevalence) of SARS-CoV-2 infection, disease, outcome, or vaccination.<br>• Measures of risk (prevalence ratios, incidence risk, risk ratios, rate ratios, odds ratios, hazard ratios) of SARS-CoV-2 infection, disease, outcome, or vaccination.<br>• Any measures of association between refugee and migrant health and lock-down measures or pandemic control policies/ strategies (e.g., indirect health outcomes (such as health related behavior like food insecurity, smoking, physical activity or social determinants of health like income or occupational aspects), if it is related to explicit pandemic measures e.g., specific lockdown measures like border closures, stay-at-home-orders, quarantine or specific policies)<br>• healthcare/Health service-related outcomes | • Crude cases without denominators,<br>• Proportions without denominators.<br>• Knowledge, attitude and practice related outcomes. |
| **Geographical area** | Worldwide | No studies to be excluded based on geography. |
| **Language of publication** | Studies published in English, German or Spanish will be included (but searches will be conducted only in English). | Studies in other languages than English, German or Spanish. |
| **Date of Publication** | Studies published since June 2020. | |

### 3 Excluded reports

Table S4 shows all studies that were excluded during full-text screening, with brief reasons for exclusion.

**Table S4: Excluded reports with breaf reasons for exclusion**

| Authors | Year (published) | Title | Reason for exclusion |
|---|---|---|---|
| Aambø, Arild | 2021 | Immigrants and COVID-19 infections | language |
| Aamir, Alifiya; Awan, Hashir Ali; Diwan, Mufaddal Najmuddin; Kazmi, Syeda Kanza; Hasan, Mohammad Mehedi; Douba, Zain; Ullah, Irfan | 2021 | The dilemma of treating the infected versus the wounded: Gaza's healthcare system is choked | population |
| Abbasi-Shavazi, Mohammad Jalal | 2021 | COVID-19, economic recession, and the Refugee situation | type of article |
| Aborode, Abdullahi Tunde; Tsagkaris, Christos; Oyeyemi, Ajagbe Abayomi; Akanji, Onigbinde Oluwanisola; Essar, Mohammad Yasir; Ahmad, Shoaib; Khawaja, Uzzam Ahmed; Cruz-Martins, Natália; Batiha, Gaber El-Saber | 2021 | The genes road: impact of migration on newborn screening and health amid the COVID-19 pandemic in the Eastern Mediterranean region | focus* |
| Adam, T. Schulman; Gyan, Bhanot | 2021 | Migration of households from New York City and the Second Peak in Covid-19 cases in New Jersey, Connecticut and New York Counties | population |
| Adhikari, S.; Mandal, A.; Guha, S. | 2021 | Place attachment and decisions to move: A study of indian migrants during Covid-19 pandemic and directions to future research | outcome** |
| Afzali, Mehdi; Ryazantsev, S. V.; Shakeri, Ali | 2020 | [The psychological impacts of quarantine on international students' life satisfaction in Russia during coronavirus COVID-19] | language |
| Ahmed, Randa I.; Khalil, Mahmoud A. F.; Mahmoud, Eman E.; Nakhla, Osama L.; Ali, Salah A.; Ahmed, Mona I. | 2021 | Comparison of the clinical and radiological manifestations of male patients with COVID-19 from different ethnicities | focus* |
| Akhtar, Aemal; Bawaneh, Ahmad; Awwad, Manar; Al-Hayek, Hadeel; Sijbrandij, Marit; Cuijpers, Pim; Bryant, Richard A. | 2021 | A longitudinal study of mental health before and during the COVID-19 pandemic in Syrian refugees | outcome† |
| Al-Oraibi, Amani; Martin, Christopher A.; Hassan, Osama; Wickramage, Kolitha; Nellums, Laura B.; Pareek, Manish | 2021 | Migrant health is public health: a call for equitable access to COVID-19 vaccines | type of article |
| Ala, Aftab; Edge, Chantal; Zumla, Alimuddin; Shafi, Shuja | 2021 | Specific COVID-19 messaging targeting ethnic minority communities | type of article |
| Alahmad, Barrak; Kurdi, Hussam; Colonna, Kyle; Gasana, Janvier; Agnew, Jacqueline; Fox, Mary A. | 2020 | COVID-19 stressors on migrant workers in Kuwait: cumulative risk considerations | type of study |
| Algarni, Majed A.; Alzahrani, Mohammad S.; Alatawi, Yasser; Alasmari, Raghad A.; Alsaab, Hashem O.; Almalki, Atiah H.; Alhifany, Abdullah A.; Althobaiti, Yusuf S. | 2021 | Perception of Threat and Psychological Impact of COVID-19 among Expatriates in Makkah Region, Saudi Arabia | other (unspecific pandemic measures) |
| Ali, Mohamed Abdelghafour; Al-Khani, Abdullah Murhaf; Sidahmed, Laila Awad | 2020 | Migrant health in Saudi Arabia during the COVID-19 pandemic | type of article |
| Ali, Parveen; Adam, Zulfiquar; West, John; Pareek, Manish; Raza, Muhammad; Iqbal, Javaid | 2021 | Perceptions of COVID-19-related risk and mortality among ethnically diverse healthcare professionals in the UK | population |
| Alkindi, Fahad Alabri Maather Alissaii Maryam Alkindi Shihab Alrashdi Tasneem Alharthi Salma Alfarsi Muzna Alsaidi Khalid Alghafri Adil Allawati Sultan Alasmi Shabib Alhashmi Khamis Murthui Sathiya Nadar Sunil K.; Alsabti, Hilal | 2021 | SIGNIFICANT ETHNIC DIFFERENCES IN PREVALENCE, CARDIAC MANIFESTATIONS AND MORTALITY IN PATIENTS WITH MODERATE TO SEVERE COVID-19 BETWEEN MIDDLE EASTERN ARABS AND SOUTH ASIAN ETHNICITY | type of article |
| Allan-Blitz, Lao-Tzu; Hertlein, Fred; Klausner, Jeffrey D. | 2021 | Heterogeneity in SARS-CoV-2 Positivity by Ethnicity in Los Angeles | population |

| | | | |
|---|---|---|---|
| Alsharif, Fahad | 2021 | Undocumented migrants in Saudi Arabia: COVID-19 and amnesty reforms | outcome† |
| Amoako, Joyceline; MacEachen, Ellen | 2021 | Understanding the blended impacts of COVID-19 and systemic inequalities on sub-Saharan African immigrants in Canada | focus* |
| Anderson, B., F. Poeschel and M. Ruhs | 2020 | Covid-19 and the transformation of migration and mobility globally - COVID-19 and systemic resilience: What role for migrant workers? | focus* |
| Aneja, Ranjan; Ahuja, Vaishali | 2020 | An assessment of socioeconomic impact of COVID-19 pandemic in India | outcome† |
| Angulo-Giraldo, M.; Guanipa-Ramírez, L.; Albites-Sanabria, J. | 2021 | Media, emotional impact and health recommendations on venezuelan migrant women during the covid-19 | other (unspecific pandemic measures) |
| Anis, S.; Khan, M. M.; Ali, Z.; Khan, A.; Arsalan, H. M.; Naeem, S.; Saleem, I.; Qamar, S.; Khan, M. M.; Ahmad, A.; Ahmed, N. | 2021 | Novel corona virus disease (COVID-19): An updated review on epidemiology, pathogenicity, clinical course, treatments, migrant health concerns and risk factors predictions | type of study (review without eligible included studies) |
| Anna, Deal; Sally, E. Hayward; Mashal, Huda; Felicity, Knights; Alison, F. Crawshaw; Jessica, Carter; Osama, B. Hassan; Yasmin, Farah; Yusuf, Ciftci; May, Rowland-Pomp; Kieran, Rustage; Lucy, Goldsmith; Monika, Hartmann; Sandra, Mounier-Jack; Rachel, Burns; Anna, Miller; Fatima, Wurie; Ines, Campos-Matos; Azeem, Majeed; Sally, Hargreaves | 2021 | Strategies and action points to ensure equitable uptake of COVID-19 vaccinations: A national qualitative interview study to explore the views of undocumented migrants, asylum seekers, and refugees | outcome† |
| Apostolara, P.; Tseroni, M.; Adamakidou, T. | 2021 | The covid-19 pandemic and challenges for the health care of migrants and refugees | no full-text |
| Armocida, Benedetta; Formenti, Beatrice; Missoni, Eduardo; D'Apice, Clelia; Marchese, Valentina; Calvi, Marzia; Castelli, Francesco; Ussai, Silvia | 2021 | Challenges in the equitable access to COVID-19 vaccines for migrant populations in Europe | type of article |
| Ashktorab, H.; Pizuorno, A.; Dalivand, M. M.; Adeleye, F. O.; Oskrochi, G.; Kibreab, A.; Oppong-Twene, P.; Tamanna, N.; Ibrahim, M. E.; Umeh, L. N.; Moon, E. S.; Kone, A. M.; Banson, A.; Federman, C.; Kara, A.; Gavin, W.; Shah, P.; Palaiodimos, L.; Mani, V. R.; Gayam, V.; Miller, J.; Challa, S. R.; Sherif, Z. A.; Aduli, F.; Laiyemo, A. O.; Brim, H. | 2021 | CLINICAL CHARACTERISTICS, GASTROINTESTINAL MANIFESTATIONS AND OUTCOMES OF COVID-19 PATIENTS AMONG MINORITIES IN THE USA | no full-text |
| Ashraful, Hoque; Md Marufur, Rahman; Anindita, Das | 2021 | Serial evaluation of anti-SARS-CoV-2 IgG antibody and breakthrough infections in BNT162b2 Vaccinated migrant workers from Bangladesh | outcome† |
| Asis, M. M. B. | 2021 | Migrant workers in the time of covid-19: The philippine experience | no full-text |
| B C, Udaya Bahadur; Pokharel, Sunil; Munikar, Sabika; Wagle, Chetan Nidhi; Adhikary, Pratik; Shahi, Brish Bahadur; Thapa, Chandra; Bhandari, Ram Prasad; Adhikari, Bipin; Thapa, Kanchan | 2021 | Anxiety and depression among people living in quarantine centers during COVID-19 pandemic: A mixed method study from western Nepal | duplicate |
| Baaees, Manal Salem Omar; Naiene, Jeremias D.; Al-Waleedi, Ali Ahmed; Bin-Azoon, Nasreen Salem; Khan, Muhammad Fawad; Mahmoud, Nuha; Musani, Altaf | 2021 | Community-based surveillance in internally displaced people's camps and urban settings during a complex emergency in Yemen in 2020 | focus* |
| Badrfam, Rahim; Zandifar, Atefeh | 2021 | Mental health status of Afghan immigrants in Iran during the COVID-19 pandemic: An exacerbation of a long-standing concern | type of article |
| Baggio, Stephanie; Jacquerioz, Frederique; Salamun, Julien; Spechbach, Hervé; Jackson, Yves | 2021 | Equity in access to COVID-19 testing for undocumented migrants and homeless persons during the initial phase of the pandemic | population |
| Balakrishnan, Vijay Shankar | 2021 | Impact of COVID-19 on migrants and refugees | type of study |
| Bartovic, Jozef; Datta, Siddharta Sankar; Severoni, Santino; D'Anna, Veronika | 2021 | Ensuring equitable access to vaccines for refugees and migrants during the COVID-19 pandemic | type of article |
| Bauder, H. and M. Godoy | 2020 | Covid-19 and the transformation of migration and mobility globally - Upscaling migrant sanctuary and solidarity policies and practices in times of COVID-19 | focus* |

| | | | |
|---|---|---|---|
| Bellizzi, Saverio; Aidyralieva, Chinara; Alsawhala, Lora; Al-Shaikh, Ala'a; Santoro, Alessio; Profili, Maria Cristina | 2021 | Vaccination for SARS-CoV-2 of migrants and refugees, Jordan | type of article |
| Bellizzi, Saverio; Alsawalha, Lora; Samawi, Luma; Al-Shaikh, Ala'a; Alfar, Hadeel; Muthu, Nazeema; Profili, Maria Cristina | 2021 | The Impact of the SARS-CoV-2 Pandemic on Mental Health in Vulnerable Population Settings: The Case of Jordan | type of article |
| Bhandari, Divya; Kotera, Yasuhiro; Ozaki, Akihiko; Abeysinghe, Sudeepa; Kosaka, Makoto; Tanimoto, Tetsuya | 2021 | COVID-19: challenges faced by Nepalese migrants living in Japan | focus* |
| Bingham, Rachel; Pickles, Hilary | 2021 | Prolonged solitary confinement of UK immigration detainees during the pandemic | type of article |
| Blake, Hassanatu; Brown, Nashira; Follette, Claudia; Morgan, Jessica; Yu, Hairui | 2021 | Black, Indigenous, People of Color, and International Students: Experiences and Resolutions Beyond COVID-19 | type of article |
| Bojorquez, I.; Cabieses, B.; Arosquipa, C. | 2021 | Migration and health in Latin America during the COVID-19 pandemic and beyond (vol 397, pg 1243, 2021) | type of article |
| Bojorquez, Ietza; Cabieses, Báltica; Arósquipa, Carlos; Arroyo, Juan; Novella, Andrés Cubillos; Knipper, Michael; Orcutt, Miriam; Sedas, Ana Cristina; Rojas, Karol | 2021 | Migration and health in Latin America during the COVID-19 pandemic and beyond | type of article |
| Brandariz, José A.; Fernández-Bessa, Cristina | 2021 | Coronavirus and Immigration Detention in Europe: The Short Summer of Abolitionism? | outcome† |
| Brickhill-Atkinson, Micah; Hauck, Fern R. | 2021 | Impact of COVID-19 on Resettled Refugees | type of article |
| Cauda, R.; Ondrusova, Z.; Tomanek, P.; Hardy, M. | 2021 | We shall start Health Intervention against Collateral Effect of Pandemic to Metabolic, Cardiovascular and Mental Health in Migrants, Children and Caregivers | type of article |
| Ceccarelli, Giancarlo; Lopalco, Maurizio; d'Ettorre, Gabriele; d'Ettorre, Gabriella; Ciccozzi, Massimo | 2021 | Surveillance of COVID-19 in migrant reception centres: a call for action | type of article |
| Chabbar, S.; Benmir, M.; Karkri, J. E.; Bensaid, K.; Aboulaich, R.; Nejjari, C. | 2021 | Modeling and simulation of the evolution of the corona virus pandemic in a context of Migration | population |
| Chackalackal, Dhia Joseph; Al-Aghbari, Ahmed Asa'Ad; Jang, Su Yeon; Ramirez, Tatiana Rivera; Vincent, Jose; Joshi, Anand; Banjara, Megha Raj; Asaga, Peter; Sanchez, Rocio Cardenas; Carrillo, Maria Angelica; Villa, Juan Manuel; Monsalve, Sonia Diaz; Kroeger, Axel | 2021 | The Covid-19 pandemic in low- and middle-income countries, who carries the burden? Review of mass media and publications from six countries | type of study (review without eligible included studies) |
| Charlton, Carmen L.; Nguyen, Leonard T.; Bailey, Ashley; Fenton, Jayne; Plitt, Sabrina S.; Marohn, Carol; Lau, Cheryl; Hinshaw, Deena; Lutsiak, Christie; Simmonds, Kimberley; Kanji, Jamil N.; Zelyas, Nathan; Lee, Nelson; Mengel, Michael; Tipples, Graham | 2021 | Pre-Vaccine Positivity of SARS-CoV-2 Antibodies in Alberta, Canada during the First Two Waves of the COVID-19 Pandemic | population |
| Chen, Y. Y. Brandon | 2021 | Fortress World: Refugee Protection during (and after) the COVID-19 Pandemic (preprint) | type of article |
| Chen, Yea-Hung; Glymour, M. Maria; Catalano, Ralph; Fernandez, Alicia; Nguyen, Tung; Kushel, Margot; Bibbins-Domingo, Kirsten | 2021 | Excess Mortality in California During the Coronavirus Disease 2019 Pandemic, March to August 2020 | population |
| Chung, N. N.; Chew, L. Y. | 2021 | Modelling Singapore COVID-19 pandemic with a SEIR multiplex network model | outcome† |
| Clark, Eva; Fredricks, Karla; Woc-Colburn, Laila; Bottazzi, Maria Elena; Weatherhead, Jill | 2020 | Disproportionate impact of the COVID-19 pandemic on immigrant communities in the United States | type of article |
| Cleaveland, Carol Waslin Michele | 2021 | COVID-19: Threat and Vulnerability Among Latina Immigrants | type of article |

| | | | |
|---|---|---|---|
| Corvacho, Mafalda; França, Gustavo | 2021 | The hidden damage of the COVID-19: The impact of the pandemic on migrants and ethnic minorities | type of article |
| Crawshaw, A. F.; Farah, Y.; Deal, A.; Goldsmith, L. P.; Carter, J.; Rustage, K.; Campos-Matos, I.; Vandrevala, T.; Forster, A. S.; Hargreaves, S. | 2021 | Analysing drivers of routine and COVID-19 vaccination in migrants to develop tailored interventions | type of article |
| Côté, Daniel; Durant, Steve; MacEachen, Ellen; Majowicz, Shannon; Meyer, Samantha; Huynh, Ai-Thuy; Laberge, Marie; Dubé, Jessica | 2021 | A rapid scoping review of COVID-19 and vulnerable workers: Intersecting occupational and public health issues | type of study |
| Dalexis, Rose Darly; Cénat, Jude Mary | 2020 | Asylum seekers working in Quebec (Canada) during the COVID-19 pandemic: Risk of deportation, and threats to physical and mental health | type of article |
| Donà, Giorgia | 2021 | Race, immigration and health: the Hostile Environment and public health responses to Covid-19 | type of article |
| Duh-Leong, Carol; Yin, H. Shonna; Yi, Stella S.; Chen, Sabrina L.; Mui, Angel; Perrin, Eliana M.; Zhao, Qiuqu; Gross, Rachel S. | 2021 | Material Hardship and Stress from COVID-19 in Immigrant Chinese American Families with Infants | population |
| Dut, Garang M. | 2021 | The experience of South Sudanese migrant families during the COVID-19 pandemic in Melbourne | type of article |
| Dávila Morán, R. C. | 2021 | Health of refugees and migrants during the covid-19 pandemic | type of article |
| Edmonds, Jennifer; Flahault, Antoine | 2021 | Refugees in Canada during the First Wave of the COVID-19 Pandemic | focus* |
| Ejikemel, J. N. U.; Ifeanyi, E. I.; Ejikeme, J. N. U. | 2021 | Sexual Violence and the Plights of Internally Displaced Persons During the Covid-19 Pandemic Lockdown in Nigeria | outcome† |
| Ekwonye, Angela U.; Hearst, Mary Howard Heather | 2021 | Reducing the Impact of COVID-19 Pandemic Using Meaning-Based Coping Strategies: Voices of Somali, Karen and Latinx Immigrant and Refugee Communities in the Twin Cities, Minnesota | no full-text |
| Elander, Ingemar Granberg Mikael Montin Stig | 2021 | Governance and planning in a ‘perfect storm’: Securitising climate change, migration and Covid-19 in Sweden | type of study |
| Elisabeth, Mangrio; Maneesh, Paul-Satyaseela; Michael, Strange | 2020 | Refugees in Sweden During the Covid-19 Pandemic-The Need for a New Perspective on Health and Integration | type of article |
| Endale, Tarik; St Jean, Nicole; Birman, Dina | 2020 | COVID-19 and refugee and immigrant youth: A community-based mental health perspective | type of article |
| Epicenre | 2021 | Level of Immunity, Neutralization of the COVID-19 Virus and Reinfections in Residents of 2 Migrant Workers' Residences | other (study was withdrawn) |
| Espinel, Zelde; Chaskel, Roberto; Berg, Ryan C.; Florez, Hermes Jose; Gaviria, Silvia L.; Bernal, Oscar; Berg, Kim; Muñoz, Carlos; Larkin, Marisa G.; Shultz, James M. | 2020 | Venezuelan migrants in Colombia: COVID-19 and mental health | type of article |
| Etienne, Gignoux; Frida, Athanassiadis; Ahmed, Garat Yarrow; Abdullahi, Jimale; Nicole, Mubuto; Carole DÉ, Glise; Denis, Onsongo Mosoti; Andrew, S. Azman; Matilu, Mwau; Francisco, Luquero; Iza, Ciglenecki | 2021 | Seroprevalence of SARS-CoV-2 antibodies and retrospective mortality in a refugee camp, Dagahaley, Kenya | population |
| Fabi, Rachel Cervantes Lilia | 2021 | Undocumented Immigrants and COVID-19: A Call for Federally Funded Health Care | type of article |
| Fabi, Rachel E.; Ludmir, Jack | 2021 | Pregnancy, Pandemics, and Public Health Policy: The Disparate Impact of COVID-19 on Pregnant Immigrants | type of study |
| Fallah, P. S. | 2021 | dMOBILE HEALTH CLINICS TO COMBAT THE SPREAD OF COVID-19 IN MIGRANT FARMWORKERS OF SKAGIT COUNTY, WASHINGTON | no full-text |

| | | | |
|---|---|---|---|
| Fernandez Ruiz, Maria Remedios | 2021 | Pandemic natives, pandemic immigrants: effects of COVID-19 confinement on the wellbeing of children in preschool education | no full-text |
| Fielding, Tony; Ishikawa, Yoshitaka | 2021 | COVID-19 and migration: A research note on the effects of COVID-19 on internal migration rates and patterns in Japan | outcome† |
| Finell, Eerika; Tiilikainen, Marja; Jasinskaja-Lahti, Inga; Hasan, Nasteho; Muthana, Fairuz | 2021 | Lived Experience Related to the COVID-19 Pandemic among Arabic-, Russian- and Somali-Speaking Migrants in Finland | other (knowledge attitude practice) |
| Fischer, Philip R. | 2021 | COVID, Lockdowns, and Dengue Fever | no full-text |
| Foppiano Palacios, Carlo; Travassos, Mark A. | 2021 | Vaccinating Detained Migrants against SARS-CoV-2 - Preventing Another Tragedy | type of article |
| Fuentes-Contreras, Édgar Hernán Rivas-Ramírez Daniel | 2021 | Irregular Migrants in Colombia during the COVID-19 Crisis. Last on List? (preprint) | no full-text |
| Galanaud, Pierre; Galanaud, Anne | 2020 | Recent immigrants at increased pandemic risk | type of article |
| Ghram, Amine; Bragazzi, Nicola Luigi; Briki, Walid; Jenab, Yaser; Khaled, Mehdi; Haddad, Monoem; Chamari, Karim | 2021 | COVID-19 Pandemic and Physical Exercise: Lessons Learnt for Confined Communities | focus* |
| Gjerløw, H.; Karim, S.; Østby, G. | 2021 | When Governments and International Organizations Shut Down: The Impact of COVID-19 on Refugees' and Host Community's Use of Educational Services in Cox's Bazar | outcome† |
| Gomez-Aguinaga, Barbara; Dominguez, Melanie Sayuri; Manzano, Sylvia | 2021 | Immigration and Gender as Social Determinants of Mental Health during the COVID-19 Outbreak: The Case of US Latina/os | outcome† |
| Graber, Kelsey M.; Byrne, Elizabeth M.; Goodacre, Emily J.; Kirby, Natalie; Kulkarni, Krishna; O'Farrelly, Christine; Ramchandani, Paul G. | 2021 | A rapid review of the impact of quarantine and restricted environments on children's play and the role of play in children's health | focus* |
| Greenaway, Christina; Hargreaves, Sally; Barkati, Sapha; Coyle, Christina M.; Gobbi, Federico; Veizis, Apostolos; Douglas, Paul | 2020 | COVID-19: Exposing and addressing health disparities among ethnic minorities and migrants | type of article |
| Grewal, R.; Koul, R. L. | 2021 | Refugees in the COVID-19 | type of article |
| Gunessh, H. J. | 2021 | Community-Level Factors Associated with COVID-19 Morbidity and Mortality in Queens, New York City | outcome† |
| Ha, Bui Thi Thu; Ngoc Quang, La; Quoc Thanh, Pham; Duc, Duong Minh; Mirzoev, Tolib; Bui, Thi My Anh | 2021 | Community engagement in the prevention and control of COVID-19: Insights from Vietnam | focus* |
| Hakim, A. J.; Victory, K. R.; Chevinsky, J. R.; Hast, M. A.; Weikum, D.; Kazazian, L.; Mirza, S.; Bhatkoti, R.; Schmitz, M. M.; Lynch, M.; Marston, B. J. | 2021 | Mitigation policies, community mobility, and COVID-19 case counts in Australia, Japan, Hong Kong, and Singapore | population |
| Hall, Brian J.; Zhao, Peizhen; Xiong, Ming Zhou; Latkin, Carl; Yang, Bin; Wang, Cheng | 2021 | Exploring correlates of depression, quality of life and alcohol misuse: a nationwide cross-sectional study of international migrants during the COVID-19 epidemic in China | focus* |
| Hargreaves, Sally; Hayward, Sally E.; Noori, Teymur; McKee, Martin; Kumar, Bernadette | 2021 | COVID-19: counting migrants in | type of study |
| Hayward, SE., Deal, A., Cheng, C., Crawshaw, A., Orcutt, M., Vandrevala, T. F., ... & Hargreaves, S. | 2021 | Clinical outcomes and risk factors for COVID-19 among migrant populations in high-income countries: a systematic review | type of article |
| Hayward, SE.; Deal, A.; Cheng, C.; Orcutt, M.; Norredam, M.; Veizis, A.; Campos-Matos, I.; McKee, M.; Kumar, B.; Hargreaves, S. | 2021 | Impact of COVID-19 on migrant populations in high-income countries: a systematic review | Other (duplicate) |
| Hedrick, Kyli; Borschmann, Rohan | 2021 | Addressing self-harm among detained asylum seekers in Australia during the COVID-19 pandemic | type of article |

| | | | |
|---|---|---|---|
| Hendriksen, Pauline A.; Garssen, Johan Bijlsma Elisabeth Y.; Engels, Ferdi Bruce Gillian Verster Joris C. | 2021 | COVID-19 Lockdown-Related Changes in Mood, Health and Academic Functioning | population |
| Henning-Smith, Carrie; Tuttle, Mariana; Kozhimannil, Katy B. | 2021 | Unequal Distribution of COVID-19 Risk Among Rural Residents by Race and Ethnicity | type of article |
| Herbrüggen, E. D. Cindik | 2021 | Analyzing the psychological impact of the COVID-19 pandemic among turkish immigrants treated at the neuro-psychiatric center riem Munich | type of article |
| Hintermeier, M., Gencer, H., Kajikhina, K., Rohleder, S., Hövener, C., Tallarek, M., ... & Bozorgmehr, K. | 2021 | SARS-CoV-2 among migrants and forcibly displaced populations: A rapid systematic review | type of article |
| Hunt, Lucy | 2021 | Bordered Trajectories: The Impact of Institutional Bordering Practices on Young Refugees' (Re-)Engagement with Post-15 Education in Greece | focus* |
| Ibrahim-Dwairy, Camelia Dwairy Marwan | 2021 | Anxiety and Coping among Arab Students in Israel during the 2020 Coronavirus Lockdown | population |
| International Organization for Migration (IOM) | 2021 | Kyrgyzstan: Study on The Socioeconomic Effects of Covid-19 on Returnees | focus* |
| International Organization for Migration (IOM) | 2020 | Status of Nepali Migrant Workers in Relation to Covid-19 | outcome† |
| International Organization for Migration (IOM) | 2021 | Migration Health 2020 Impact Overview | outcome† |
| International Organization for Migration (IOM) and Eurasylum Ltd | 2020 | Migration Policy Practice (Vol. X, Number 2, April–June 2020) | type of article |
| International Organization for Migration (IOM) and Eurasylum Ltd | 2020 | Migration Policy Practice (Vol. X, Number 3, July-August 2020) | type of study |
| Irene Schöfberger, Marzia Rango | 2020 | Migration in West and North Africa and across the Mediterranean - COVID-19 and migration in West and North Africa and across the Mediterranean | population |
| Irudaya Rajan, S.; Sivakumar, P.; Srinivasan, Aditya | 2020 | The COVID-19 Pandemic and Internal Labour Migration in India: A 'Crisis of Mobility' | outcome† |
| Iskander, Natsha | 2020 | Qatar, the Coronavirus, and Cordons Sanitaires: Migrant Workers and the Use of Public Health Measures to Define the Nation | type of article |
| Islam, Md Bayezid; Sultana, Tahmina; Marium, Marzia; Mahfuz, Mahzabeen; Rahman, Syed Mahbubur | 2021 | COVID-19 response during the winter season in Rohingya refugee camps in Cox's Bazar, Bangladesh | type of study |
| Islam, Mohammad Mainul; Yunus, Md Yeasir | 2020 | Rohingya refugees at high risk of COVID-19 in Bangladesh | type of study |
| Islam, Muhammad Nazrul; Inan, Toki Tahmin; Islam, A. K. M. Najmul | 2020 | COVID-19 and the Rohingya Refugees in Bangladesh: The Challenges and Recommendations | type of study |
| Jackulikova, M.; Tothova, M.; Bydzovsky, J.; Olah, M.; Bielova, M.; Kmit, I.; Mlynarcik, P.; Vlcek, R.; Krcmery, V.; Nguyen, L. | 2021 | Comparison of the Spectrum of Outpatient visits before and after Fire in the Moria Camp after Arrival of Covid-19 Positive Refugees | focus* |
| Jakab, Zsuzsanna; Severoni, Santino; Alderslade, Richard; Kaojaroen, Kanokporn; Hossain, Rifat; Melki, Simona | 2021 | Health and migration: Will migrants share the road to recovery from COVID-19? | type of article |
| James, T. | 2021 | THE COVID-19 PANDEMIC AND STRUCTURAL BARRIERS FOR MIGRANT AGRICULTURAL WORKERS IN ONTARIO | type of article |
| Jara-Labarthé, Vanessa; Cisneros Puebla, César A. | 2021 | Migrants in Chile: Social crisis and the pandemic (or sailing over troubled water…) | type of study |
| Jauhiainen, J. S.; Tedeschi, M. | 2021 | Healthcare of Undocumented Migrants | outcome† |
| Jawad, Nadine K.; Taweeleh, Lina Abu; Elharake, Jad A.; Khamis, Nicole; Alser, Osaid; Karaki, Fatima M.; Aboukhater, Layla | 2021 | Refugee access to COVID-19 vaccines in Lebanon | type of article |

| | | | |
|---|---|---|---|
| Jayaram, Nivedita; Varma, Divya | 2020 | Examining the 'Labour' in Labour Migration: Migrant Workers' Informal Work Arrangements and Access to Labour Rights in Urban Sectors | outcome† |
| Jedwab R; Khan AM; Russ J; Zaveri ED. | 2021 | Epidemics, Pandemics, and Social Conflict: Lessons from the Past and Possible Scenarios for COVID-19 | focus* |
| Jesline J; Romate J; Rajkumar E; George AJ. | 2021 | The plight of migrants during COVID-19 and the impact of circular migration in India: a systematic review | type of article (review without eligible included studies) |
| Jha, Sweety Suman; Lahiri, Arista | 2020 | Domestic migrant workers in India returning to their homes: emerging socioeconomic and health challenges during the COVID-19 pandemic | type of article |
| Jithesh, P. K. | 2021 | A model based on cellular automata for investigating the impact of lockdown, migration and vaccination on COVID-19 dynamics | population |
| Johansen, R.; Straiton, M.; Hynek, K. | 2021 | Negative change in work situation due to covid-19 pandemic and mental health problems among Norwegians and European migrants - a cross sectional study | type of article |
| John, Ekatha Ann; Kapilashrami, Anuj | 2021 | Victims, villains and the rare hero: Analysis of migrant and refugee health portrayals in the Indian print media | focus* |
| Jozaghi, Ehsan; Dahya, Azim | 2020 | Refugees, asylum seekers and COVID-19: Canada needs to do more to protect at-risk refugees during the current pandemic | type of article |
| Kamara, Mary; Bingham, Rachel | 2020 | Covid-19: unjustified risk to people held in immigration removal centres | type of article |
| Kamrath, C.; Mönkemöller, K.; Fink, K.; Biester, T.; Raile, K.; Rohrer, T.; Warncke, K.; Pappa, A.; Hammersen, J.; Holl, R. | 2021 | Ketoacidosis in children and adolescents with newly diagnosed type 1 diabetes during the COVID-19 pandemic in Germany-results from a nationwide registry | type of article |
| Khondker, H. | 2021 | Covid-19 and Migrant Workers: The Gulf and Singapore | type of article |
| Kondilis, Elias; Puchner, Karl; Veizis, Apostolos; Papatheodorou, Christos; Benos, Alexis | 2020 | Covid-19 and refugees, asylum seekers, and migrants in Greece | type of article |
| Kowalewska, Dorota; Adamczyk, Anita; Trojanowska-Strzeboszewska, Monika | 2021 | Dataset of Ukrainian migrant workers opinions on their stay in Poland during COVID-19 lockdown | other (knowledge attitude practice) |
| Krist, Lilian; Dornquast, Christina; Reinhold, Thomas; Icke, Katja; Danquah, Ina; Willich, Stefan N.; Becher, Heiko; Keil, Thomas | 2021 | Predictors of Changes in Physical Activity and Sedentary Behavior during the COVID-19 Pandemic in a Turkish Migrant Cohort in Germany | other (unspecific pandemic measures) |
| Kurt, G.; Ilkkursun, Z.; Javanbakht, A.; Uygun, E.; Karaoglan-Kahilogullari, A.; Acarturk, C. | 2021 | The psychological impacts of COVID-19 related stressors on Syrian refugees in Turkey: The role of resource loss, discrimination, and social support | outcome† |
| Lakhan, Ram; Sharma, Manoj | 2021 | Depression in Women Migrants Living in Slums in North India | type of study |
| Lam, Elene | 2020 | Migrant sex workers left behind during COVID-19 pandemic | type of article |
| Lam, Elene Shih Elena Chin Katherine Zen Kate | 2021 | The Double-Edged Sword of Health and Safety: COVID-19 and the Policing and Exclusion of Migrant Asian Massage Workers in North America | focus* |
| Lamb, Danielle Gomez Rafael Moghaddas Milad | 2021 | Unions and hazard pay for COVID-19: Evidence from the Canadian Labour Force Survey | outcome† |
| Lampo, Margarita; Hernández-Villena, Juan V.; Cascante, Jaime; Vincenti-González, María F.; Forero-Peña, David A.; Segovia, Maikell J.; Hampson, Katie; Castro, Julio; Grillet, Maria Eugenia | 2021 | Signatures of the Venezuelan Humanitarian Crisis in the First Wave of COVID-19: Fuel Shortages and Border Migration | focus* |
| Langalia, Akshay K.; Patel, Dolly; Prajapati, Dhaval; Munia, Vipul; Shah, Dhairya; Lalwani, Yash; Panchal, Drashti | 2021 | Large-Scale Rapid Antigen Testing of Migrants for Detection of SARS-CoV-2 Virus by Dental Interns at Various Checkpoints of Ahmedabad City, India: A Short Communication Report | population |

| | | | |
|---|---|---|---|
| Langellier, Brent A. | 2020 | Policy Recommendations to Address High Risk of COVID-19 Among Immigrants | outcome† |
| Lara-Cinisomo, Sandraluz; D'Anna-Hernandez, Kimberly; Non, Amy L. | 2021 | Recommendations for Clinical Practice, Research, and Policy to Address the Effects of the COVID-19 Pandemic on Anxiety Symptoms in Immigrant and U.S.-Born Latina Mothers | type of article |
| Le Bihan, C.; Faucherre, V.; Le Moing, V.; Mehenni, A.; Nantes, D.; Da Silva, A.; Jaume, C.; Lassalle, F.; Makinson, A. | 2021 | COVID-19: The forgotten cases of hidden exiles | population |
| Leiblfinger, Michael Prieler Veronika Rogoz Mădălina Sekulová Martina | 2021 | Confronted with COVID-19: Migrant live-in care during the pandemic | type of article |
| Leiner, Andrea; Sammon, Maura; Perry, Helen; Dunavant, Scott | 2020 | Facing COVID-19 and Refugee Camps on the U.S. Border | type of article |
| Lemon, E. D.; Chacon, L. D.; Santiago, C.; Sainz, M.; Renfro, T.; Crookes, D. M.; Urbina, B.; Woods-Jaeger, B. | 2021 | ADDRESSING COVID-RELATED PSYCHOSOCIAL OUTCOMES AMONG LATINO IMMIGRANT FAMILIES: LESSONS FROM A CBPR STUDY IN THE SOUTHEAST | type of article |
| Liang, Chen; Shen, Li | 2021 | An analysis of coronavirus disease 2019 with spline regression at province level during first-level response to major public health emergency out of Hubei, China | population |
| Lim, Jue Tao; Dickens, Borame Lee; Ong, Janet; Aik, Joel; Lee, Vernon J.; Cook, Alex R.; Ng, Lee Ching | 2021 | Decreased dengue transmission in migrant worker populations in Singapore attributable to SARS-CoV-2 quarantine measures | outcome† |
| Llop-Gironés, Alba; Vracar, Ana; Llop-Gironés, Gisela; Benach, Joan; Angeli-Silva, Livia; Jaimez, Lucero; Thapa, Pramila; Bhatta, Ramesh; Mahindrakar, Santosh; Bontempo Scavo, Sara; Nar Devi, Sonia; Barria, Susana; Marcos Alonso, Susana; Julià, Mireia | 2021 | Employment and working conditions of nurses: where and how health inequalities have increased during the COVID-19 pandemic? | population |
| Logie, Carmen H.; Okumu, Moses; Berry, Isha; Hakiza, Robert; Kibuuka Musoke, Daniel; Kyambadde, Peter; Mwima, Simon; Lester, Richard T.; Perez-Brumer, Amaya G.; Baral, Stefan; Mbuagbaw, Lawrence | 2021 | Kukaa Salama (Staying Safe): study protocol for a pre/post-trial of an interactive mHealth intervention for increasing COVID-19 prevention practices with urban refugee youth in Kampala, Uganda | type of study |
| Logie, Carmen H.; Okumu, Moses; Latif, Maya; Musoke, Daniel Kibuuka; Odong Lukone, Simon; Mwima, Simon; Kyambadde, Peter | 2021 | Exploring resource scarcity and contextual influences on wellbeing among young refugees in Bidi Bidi refugee settlement, Uganda: findings from a qualitative study | focus* |
| Logie, Carmen; Okumu, Moses; Hakiza, Robert; Kibuuka Musoke, Daniel; Berry, Isha; Mwima, Simon; Kyambadde, Peter; Kiera, Uwase Mimy; Loutet, Miranda; Neema, Stella; Newby, Katie; McNamee, Clara; Baral, Stefan D.; Lester, Richard; Musinguzi, Joshua; Mbuagbaw, Lawrence | 2021 | Mobile Health-Supported HIV Self-Testing Strategy Among Urban Refugee and Displaced Youth in Kampala, Uganda: Protocol for a Cluster Randomized Trial (Tushirikiane, Supporting Each Other) | type of study |
| Lokhande, Nitin Gundimeda Haripriya | 2021 | MGNREGA: The Guaranteed Refuge for Returning Migrants During COVID-19 Lockdown in India | outcome† |
| Lokken, Erica M.; Taylor, G. Gray; Huebner, Emily M.; Vanderhoeven, Jeroen; Hendrickson, Sarah; Coler, Brahm; Sheng, Jessica S.; Walker, Christie L.; McCartney, Stephen A.; Kretzer, Nicole M.; Resnick, Rebecca; Kachikis, Alisa; Barnhart, Nena; Schulte, Vera; Bergam, Brittany; Ma, Kimberly K.; Albright, Catherine; Larios, Valerie; Kelley, Lori; Larios, Victoria; Emhoff, Sharilyn; Rah, Jasmine; Retzlaff, Kristin; Thomas, Chad; Paek, Bettina W.; Hsu, Rita J.; Erickson, Anne; Chang, Andrew; Mitchell, Timothy; Hwang, Joseph K.; Gourley, Rebecca; Erickson, Stephen; Delaney, Shani; Kline, Carolyn R.; Archabald, Karen; Blain, Michela; LaCourse, Sylvia M.; Adams Waldorf, Kristina M. | 2021 | Higher severe acute respiratory syndrome coronavirus 2 infection rate in pregnant patients | population |
| Lopez, Leo; Hart, Louis H.; Katz, Mitchell H. | 2021 | Racial and Ethnic Health Disparities Related to COVID-19 | type of article |
| Lopez, William D.; Kline, Nolan; LeBrón, Alana M. W.; Novak, Nicole L.; De Trinidad Young, Maria-Elena; Gonsalves, Gregg; Mishori, Ranit; Safi, Basil A.; Kysel, Ian M. | 2021 | Preventing the Spread of COVID-19 in Immigration Detention Centers Requires the Release of Detainees | type of study |

| | | | |
|---|---|---|---|
| Luo, Yang; Chua, Cher Rui; Xiong, Zhonghui; Ho, Roger C.; Ho, Cyrus S. H. | 2020 | A Systematic Review of the Impact of Viral Respiratory Epidemics on Mental Health: An Implication on the Coronavirus Disease 2019 Pandemic | population |
| López-Jaramillo, A. M.; Rangel-Gómez, M. G.; Cruz-Piñeiro, R. | 2021 | Response to COVID-19 for the care of the Mexican migrant population in the southern United States | outcome† |
| Madge, Sara; Barber, Tristan J.; Hunter, Alan; Bhagani, Sanjay; Lipman, Marc; Burns, Fiona | 2021 | Descriptive account of 18 adults with known HIV infection hospitalised with SARS-CoV-2 infection | population |
| Mallet-Garcia, M. L.; Delvino, N. | 2021 | Rethinking exclusionary policies: The case of irregular migrants during the COVID-19 pandemic in Europe | type of study |
| Mamgain, Rajendra P. | 2021 | Understanding labour market disruptions and job losses amidst COVID-19 | population |
| Manirambona, Emery; Uwizeyimana, Theogene; Uwiringiyimana, Emmanuel; Reddy, Henna | 2021 | Impact of the COVID-19 pandemic on the food rations of refugees in Rwanda | type of article |
| Mares, Sarah; Jenkins, Kym; Lutton, Susan; Newman Am, Louise | 2021 | Impact of Covid-19 on the mental health needs of asylum seekers in Australia | type of article |
| Margineanu, I.; Mor, Z.; Garcia, D.; Gilpin, C.; Dhawan, S.; Ritz, N.; Zenner, D. | 2021 | TB and COVID-19 in migrants - the need to focus on both conditions | type of article |
| Martínez, Genaro Tapia Magali Cárdenas Núñez Ricardo Antonio Tena Moreno Adriana de la Paz Sánchez | 2021 | Following the spacial dynamics of COVID-19 in Mexico and some notes (preprint) | population |
| Maru, Sheela; Patil, Uday; Carroll-Bennett, Rachel; Baum, Aaron; Bohn-Hemmerdinger, Tracy; Ditchik, Andrew; Scanlon, Michael L.; Krishnan, Parvathy; Bogaert, Kelly; Woodbury, Carson; Maru, Duncan; Noble, Lawrence; Wasserman, Randi; Brown, Barry; Vreeman, Rachel; Masci, Joseph | 2020 | Universal screening for SARS-CoV-2 infection among pregnant women at Elmhurst Hospital Center, Queens, New York | focus* |
| Matlin, Stephen A.; Orcutt, Miriam; Bojorquez, Ietza; Caman, Ozge Karadag; Severoni, Santino; Spiegel, Paul; Veizis, Apostolos; Saso, Luciano | 2021 | COVID-19 and migrant and refugee health: A pointer to system competence in future pandemic preparedness | type of article |
| Melchior, Maria; Desgrées du Loû, Annabel; Gosselin, Anne; Datta, Geetanjali D.; Carabali, Mabel; Merckx, Joanna; Kaufman, Jay S. | 2021 | Migrant status, ethnicity and COVID-19: more accurate European data are greatly needed | type of article |
| Mia, Md Aslam; Griffiths, Mark D. | 2020 | Letter to the Editor: The economic and mental health costs of COVID-19 to immigrants | type of article |
| Miah, M. L.; Sarker, B.; Agbogan, J. A.; Tonon, B.; Koech, M. C.; Rahman, M. S. E. | 2021 | Adaptations to smart surveys in the context of COVID-19 in Cox's Bazar, Bangladesh. (Special Section: COVID-19 nutrition programming.) | type of study |
| Mishra, Bimal Kumar; Keshri, Ajit Kumar; Rao, Yerra Shankar; Mishra, Binay Kumar; Mahato, Buddhadeo; Ayesha, Syeda; Rukhaiyyar, Bansidhar Prasad; Saini, Dinesh Kumar; Singh, Aditya Kumar | 2020 | COVID-19 created chaos across the globe: Three novel quarantine epidemic models | outcome† |
| Misra, Pooja; Gupta, Jaya | 2021 | Impact of COVID 19 on Indian Migrant Workers: Decoding Twitter Data by Text Mining | outcome† |
| Molyneux, K.; Kaur, K.; Kona, N.; Malave, G. S.; Baranowski, K. A.; Singer, E. K.; Mt Sinai Human Rights, Program | 2021 | Experiences of Detained Asylum Seekers During the COVID-19 Pandemic | type of article |
| Morisod, K.; Durand, M.; Selby, K.; Pogam, M. Le Grazioli V.; Zozaya, J. Sanchis Bodenmann P.; Plessen, C. Van | 2021 | Asylum seekers' and refugees' understanding of government restrictions during the COVID-19 pandemic | type of study |
| Morisod, K.; Grazioli, V.; Schlüter, V.; Bochud, M.; Nusslé, S. Gonseth D'Acremont V.; Felappi, A.; Bühler, N.; Bodenmann, P. | 2021 | Prevalence of SARS-CoV-2 and associated risk factors among asylum seekers living in centres | no full-text |
| Moy, Foong Ming; Ng, Yit Han | 2021 | Perception towards E-learning and COVID-19 on the mental health status of university students in Malaysia | population |

| | | | |
|---|---|---|---|
| Moyo, Khangelani; Sebba, Kalyango Ronald; Zanker, Franzisca | 2021 | Who is watching? Refugee protection during a pandemic - responses from Uganda and South Africa | type of study |
| Mude, William; Meru, Clement; Njue, Carolyne; Fanany, Rebecca | 2021 | A cross-sectional study of COVID-19 impacts in culturally and linguistically diverse communities in greater Western Sydney, Australia | population |
| Mukumbang, Ferdinand C. | 2020 | Are asylum seekers, refugees and foreign migrants considered in the COVID-19 vaccine discourse? | type of article |
| Nagatani, T.; Tainaka, K. I. | 2021 | Diffusively coupled SIQRS epidemic spreading in hierarchical small-world network | no full-text |
| Nakamura, Nobuyuki Suzuki Aya | 2021 | COVID-19 and the Intentions to Migrate from Developing Countries: Evidence from Online Search Activities in Southeast Asia | focus* |
| Neha, Limaye; Brennan, Ninesling; Frantzso, Marcelin; Cody, Nolan; Walter, Sobba; Matthew, Hing; Emily, Ptaszek; Fernet, Leandre; Daniel, Palazuelos | 2021 | COVID-19 Pandemic Response in a Migrant Farmworker Community: Excess Mortality, Testing Access and Contact Tracing in Immokalee, Florida | population |
| Ngan, Olivia M. Y.; Sanip, Suhaila | 2021 | The Vulnerability of Migrant Workers in Global COVID-19 Pandemic: Highlights from Malaysia and Hong Kong | type of article |
| Niezna, Maayan Kurlander Yahel Shamir Hila | 2021 | Underlying conditions: The Increased Vulnerability of Migrant Workers Under COVID-19 in Israel (preprint) | focus* |
| Ning Ning, Chung; Lock Yue, Chew | 2020 | Modelling Singapore COVID-19 pandemic with a SEIR multiplex network model | outcome† |
| Niranjan, Kathe; Rajvi, J. Wani | 2020 | Determinants of COVID-19 Incidence and Mortality in the US: Spatial Analysis | population |
| Otieno, M. K.; Nkenyereye, L. | 2021 | Effects of pandemics on migrant communities: Analysis of existing sources | focus* |
| Panda, S.; Roy, S.; Garg, R. K.; Hui, G.; Gorard, J.; Bhutada, M.; Sun, Y. L.; Bhatnagar, S.; Mohan, A.; Dar, L.; Liu, M. | 2021 | COVID-19 disease in hospitalized young adults in India and China: Evaluation of risk factors predicting progression across two major ethnic groups | population |
| Pandey, Kritika Parreñas Rhacel Salazar Sabio Gianne Sheena | 2021 | Essential and Expendable: Migrant Domestic Workers and the COVID-19 Pandemic: PROD | outcome† |
| Parvez, M. R. | 2021 | Social stigma and covid-19: The experiences of bangladeshi returnees from italy | outcome† |
| Penning, V.; Razum, O. | 2021 | CoviD-19: A comparison between refugee shelters and military barracks | type of article |
| Perez-Brumer, Amaya; Hill, David; Andrade-Romo, Zafiro; Solari, Karla; Adams, Ellithia; Logie, Carmen; Silva-Santisteban, Alfonso | 2021 | Vaccines for all? A rapid scoping review of COVID-19 vaccine access for Venezuelan migrants in Latin America | outcome† |
| Pilato, Tara C.; Taki, Faten A.; Kaur, Gunisha | 2021 | Safeguarding pregnant asylum-seekers and refugees during the era of COVID-19 | type of study |
| Pimentel Walker, Ana Paula; Sanga, Naganika; Benson, Odessa Gonzalez; Yoshihama, Mieko | 2021 | Risk Communication and Institutional Racism: The Protective Health Effect of Refugee-Led Community Organizations | focus* |
| Pinzón-Espinosa, Justo; Valdés-Florido, María José; Riboldi, Ilaria; Baysak, Erensu; Vieta, Eduard | 2021 | The COVID-19 Pandemic and Mental Health of Refugees, Asylum Seekers, and Migrants | type of article |
| Polisena, Julie; Ospina, Maria; Sanni, Omolara; Matenchuk, Brittany; Livergant, Rachel; Amjad, Sana; Zoric, Igor; Haddad, Nisrine; Morrison, Andra; Wilson, Kumanan; Bogoch, Isaac; Welch, Vivian A. | 2021 | Public health measures to reduce the risk of SARS-CoV-2 transmission in Canada during the early days of the COVID-19 pandemic: a scoping review | outcome† |
| Rafiqul, Abu Hayet Md; Hossen, Mohammad Shahadat; Huda, Syed Nazmul | 2021 | COVID-19 Vaccination and Undocumented Migrants | type of article |

| | | | |
|---|---|---|---|
| Raghavendra, Naveen Kumara; Gopal, Suresh; Munuswamy, Jothi | 2021 | Data set on impact of COVID-19 on mental health of internal migrant workers in India: Corona Virus Anxiety Scale (CAS) approach | outcome† |
| Rajatanavin, Nattadhanai; Tuangratananon, Titiporn; Suphanchaimat, Rapeepong; Tangcharoensathien, Viroj | 2021 | Responding to the COVID-19 second wave in Thailand by diversifying and adapting lessons from the first wave | type of study |
| Ramírez-Cervantes, K. L.; Romero-Pardo, V.; Pérez-Tovar, C.; Martínez-Alés, G.; Quintana-Diaz, M. | 2021 | A medicalized hotel as a public health resource for the containment of Covid-19: more than a place for quarantining | outcome† |
| Rast, E.; Perplies, C.; Biddle, L.; Bozorgmehr, K. | 2021 | Asylum seekerś experiences with quarantine after arriving in Germany during the COVID-19 pandemic | type of study |
| Rauta, A. K.; Rao, Y. S.; Behera, J. | 2021 | Spread of COVID-19 in Odisha (India) Due to Influx of Migrants and Stability Analysis Using Mathematical Modeling | no full-text |
| Reagu, Shuja; Wadoo, Ovais; Latoo, Javed; Nelson, Deborah; Ouanes, Sami; Masoodi, Naseer; Karim, Mustafa Abdul; Iqbal, Yousaf; Al Abdulla, Samya; Al Nuaimi, Saleem Khaldoun; Abdelmajid, Alaaeldin Abdelmajid Basheer; Al Samawi, Musaed Saad; Khoodoruth, Mohamed Adil Shah; Khoodoruth, Widaad Nuzhah Chut-Kai; Al-Maslamani, Muna A. Rahman S.; Alabdulla, Majid | 2021 | Psychological impact of the COVID-19 pandemic within institutional quarantine and isolation centres and its sociodemographic correlates in Qatar: a cross-sectional study | population |
| Rebhun, U. | 2021 | Inter-country variations in COVID-19 incidence from a social science perspective | no full-text |
| Reggio Emilia Covid-19 Working Group | 2020 | Prevalence of SARS-CoV-2 (Covid-19) in Italians and in immigrants in an area of Northern Italy (Reggio Emilia) | no full-text |
| Rinaldi, P. | 2021 | Impatto della pandemia sulle comunità di immigrati in spagna | language |
| Rodriguez, Darlene Xiomara; Hill, Jessica; McDaniel, Paul N. | 2021 | A Scoping Review of Literature About Mental Health and Well-Being Among Immigrant Communities in the United States | focus* |
| Rothman, S.; Gunturu, S.; Korenis, P. | 2020 | The mental health impact of the COVID-19 epidemic on immigrants and racial and ethnic minorities | type of article |
| Rupakula, R. M. V.; Dhananjay, Kumar | 2021 | Impact of COVID-19 lockdown on migrant labourers from Bihar in Hyderabad | no full-text |
| Rupam, Bhattacharyya; Anik, Burman; Kalpana, Singh; Sayantan, Banerjee; Subha, Maity; Arnab, Auddy; Sarit Kumar, Rout; Supriya, Lahoti; Rajmohan, Panda; Veerabhadran, Baladandayuthapani | 2021 | Role of Multi-resolution Vulnerability Indices in COVID-19 spread: A Case Study in India | population |
| Saad, Neil J. | 2020 | The Al Hol camp in Northeast Syria: health and humanitarian challenges | type of article |
| Saidin, Mohd Irwan Syazli Omar Hanana Bamadhaj | 2021 | COVID-19 policy response and the vulnerabilities of low-skilled women migrant workers in Malaysia | type of article |
| Saleem, Javeria; Ishaq, Muhammad; Zakar, Rubeena; Suddahazai, Imran Hussain Khan; Fischer, Florian | 2021 | Experiences of frontline Pakistani emigrant physicians combating COVID-19 in the United Kingdom: a qualitative phenomenological analysis | outcome† |
| Salmani, Ibrahim; Seddighi, Hamed; Nikfard, Maryam | 2020 | Access to Health Care Services for Afghan Refugees in Iran in the COVID-19 Pandemic | type of article |
| Saparamadu, Amarasinghe Arachchige Don Nalin Samandika; Sharpe, Albie; Kim, Sun; Barbosa, Bruna Ligia Ferreira Almeida; Pereira, Adrian | 2021 | Low-wage migrant workers during coronavirus disease 2019: a social determinants analysis | type of study |
| Sapra, Ipsita; Nayak, Bibhu P. | 2021 | The protracted exodus of migrants from Hyderabad in the time of COVID-19 | type of study |
| Sarkar, Sanjit | 2021 | Significance of migration to the COVID 19 outbreaks in major states in India | no full-text |

| | | | |
|---|---|---|---|
| Schnake-Mahl, Alina S.; Lazo, Mariana; Dureja, Kristina; Ehtesham, Nahian; Bilal, Usama | 2021 | Racial and ethnic inequities in occupational exposure across and between US cities | outcome† |
| Schriro, Dora | 2021 | On the Other Side of the Looking Glass: COVID-19 Care in Immigration Detention | type of article (review without eligible included studies) |
| Seiffarth, Marlene | 2021 | Crisis as Catalyst? Romanian Migrant Care Workers in Italian Home-Based Care Arrangements | focus* |
| Semé, Judith L.; Bivins, Balkys L.; Sternberg, Candice A.; Barnett, Juanita D.; Junis-Florian, Thamousca; Nicolas, Guerda; Etienne, Marie; Jean, Pepita | 2021 | The Educational, Health, and Economic Impacts of COVID-19 Among Haitians in the USA: Time for Systemic Change | type of study |
| Serafini, Randal A.; Powell, Samuel K.; Frere, Justin J.; Saali, Alexandra; Krystal, Hannah L.; Kumar, Vedika; Yashaswini, Chittampalli; Hernandez, Josimar; Moody, Kate; Aronson, Anne; Meah, Yasmin; Katz, Craig L. | 2021 | Psychological distress in the face of a pandemic: An observational study characterizing the impact of COVID-19 on immigrant outpatient mental health | focus* |
| Shaaban, A.; Abecasis, A.; Fronteira, I.; Muggli, Z.; Amado, R.; Vaz, D.; Silva, A. C.; Martins, M. R. O. | 2021 | Socioeconomic impact of COVID-19 among immigrants in Lisbon Region | no full-text |
| Shahare, V. B. | 2021 | COVID-19 lockdown: the neglected migrant workers in India (vol JAN, 10.1080/02185385.2021.1875335, 2021) | outcome† |
| Shahare, Virendra Balaji | 2021 | COVID-19 lockdown: India struggles to feed migrants left behind | type of study |
| Shalaby, Mariam A. | 2021 | A Four-Year Follow-Up to "Hosting Syrian Refugees": Challenges, Resilience, and Small Joys During COVID-19 | no full-text |
| Shen, Megan J.; Maciejewski, Paul K.; Tergas, Ana I.; Prigerson, Holly G. | 2021 | Inequities in End-of-Life Care Among Immigrant Patients Exaggerated by the COVID-19 Pandemic | type of study |
| Shields, John Alrob Zainab Abu | 2021 | The Political Economy of a Modern Pandemic: Assessing Impacts of COVID-19 on Migrants and Immigrants in Canada | type of study |
| Simic, Olivera | 2021 | Locked in and locked out: A migrant woman's reflection on life in Australia during the COVID-19 pandemic | type of article |
| Singh, G. P. | 2021 | Psychosocial and Mental Health Issues of the Migrants Amidst COVID-19 Pandemic in India: A Narrative Review | no full-text |
| Singh, Gurvinder Pal; Arun, Priti; Chavan, B. S. | 2020 | Efforts to Minimize the Impact of Lockdown on Migrant Workers in India During the COVID-19 Pandemic | no full-text |
| Singh, Sujay Vikram Singh Dr Kuldeep | 2021 | Migrants Workers in Tourism and Hospitality Industry: Effect of COVID-19 (preprint) | outcome† |
| Sisti, L. G.; Cammilli, M.; Diodati, A.; Menghini, M.; Fazioli, C.; Petrelli, A.; Napoli, A. Di Cavani A.; Mirisola, C.; Costanzo, G. | 2021 | Managing the pandemic in the reception system for migrants: the Italian experience | type of article |
| Siva Sangari Coimbatore, M.; Ida Chellam, J. A.; Rajendra Kumar, R. | 2021 | A study and analysis of government policies and support system influences migration of workers with special reference to Coimbatore city | no full-text |
| Smith, H. R.; Singer, A. | 2021 | COVID-19's effect on interior immigration enforcement and detention | no full-text |
| Sohel, M. S.; Hossain, B.; Alam, M. K.; Shi, G.; Shabbir, R.; Sifullah, M. K.; Mamy, M. M. B. | 2021 | COVID-19 induced impact on informal migrants in Bangladesh: a qualitative study | no full-text |
| Song, Suzan J. | 2021 | Protecting the Global Mental Health of Forcibly Displaced Children From the COVID-19 Pandemic | type of article |
| Spiritus-Beerden, Eva; Verelst, An; Devlieger, Ines; Langer Primdahl, Nina; Botelho Guedes, Fábio; Chiarenza, Antonio; De Maesschalck, Stephanie; Durbeej, Natalie; | 2021 | Mental Health of Refugees and Migrants during the COVID-19 Pandemic: The Role of Experienced Discrimination and Daily Stressors | outcome† |

| | | | |
|---|---|---|---|
| Garrido, Rocío; Gaspar de Matos, Margarida; Ioannidi, Elisabeth; Murphy, Rebecca; Oulahal, Rachid; Osman, Fatumo; Padilla, Beatriz; Paloma, Virginia; Shehadeh, Amer; Sturm, Gesine; van den Muijsenbergh, Maria; Vasilikou, Katerina; Watters, Charles; Willems, Sara; Skovdal, Morten; Derluyn, Ilse | | | |
| Sreejith, P. M.; Sreejith, S. | 2021 | Report on Socio-Economic Impact of COVID 19 on Migrant Workers with Reference to Kerala State | no full-text |
| Srivastava, Ravi | 2020 | Growing Precarity, Circular Migration, and the Lockdown in India | type of study |
| Sserwanja, Quraish; Adam, Mohammed Bashir; Kawuki, Joseph; Olal, Emmanuel | 2021 | COVID-19 in conflict border regions: a case of South Kordofan, Sudan | type of study |
| St-Denis, Xavier | 2020 | Sociodemographic Determinants of Occupational Risks of Exposure to COVID-19 in Canada | population |
| Stathopoulou, T. | 2021 | Depression among higher education students in Greece during the first C19 lockdown | no full-text |
| Stefanelli, J. N. | 2021 | Detained during a Pandemic: Human Rights behind Locked Doors | type of study |
| Stevens, Amy J.; Ray, Anna M.; Thirunavukarasu, Ahimza; Johnson, Ella; Jones, Lucy; Miller, Anna; Elston, James W. T. | 2021 | The experiences of socially vulnerable groups in England during the COVID-19 pandemic: A rapid health needs assessment | population |
| Suphanchaimat, Rapeepong; Nittayasoot, Natthaprang; Thammawijaya, Panithee; Teekasap, Pard; Ungchusak, Kumnuan | 2021 | Predicted Impact of Vaccination and Active Case Finding Measures to Control Epidemic of Coronavirus Disease 2019 in a Migrant-Populated Area in Thailand | population |
| Suresh, Rajani; James, Justine; R S J, Balraju | 2020 | Migrant Workers at Crossroads-The Covid-19 Pandemic and the Migrant Experience in India | type of article |
| Swargiary, P.; Swargiary, P.; Swargiary, D. | 2021 | The impact of covid-19 on migrant’s workers employment and livelihood in india | population |
| Symington, Andy | 2021 | Migrant workers and the COVID-19 crisis in Australia: an overview of governmental responses | focus* |
| Søegaard, Erik Ganesh Iyer; Kan, Zhanna | 2021 | COVID-19 infections in the districts of Oslo | language |
| Tang, Ke Xie Danxia Zhang Longtian | 2021 | Contagion, Migration and Misallocation in a Pandemic (preprint) | population |
| Tazzioli, Martina Stierl Maurice | 2021 | Europe’s unsafe environment: migrant confinement under Covid-19 | type of article |
| Teerawattananon, Yot; Teo, Yik Ying; Lim, Jeremy Fung Yen; Hsu, Li Yang; Dabak, Saudamini | 2021 | Vaccinating undocumented migrants against covid-19 | type of article |
| The, Lancet | 2021 | Protecting refugees during the COVID-19 pandemic | type of article |
| Tieri, Silvia | 2021 | Migration, Borders, and Security Discourses in the Time of COVID-19: The Case of Migration from Bangladesh to India (preprint) | outcome† |
| Tisatto, C. A.; Lopes, L. B.; Bento, J. S. | 2021 | PUBLIC POLICIES TO FIGHT THE PANDEMIC: THE DEBATE ABOUT THE IMPACT OF COVID-19 ON HUMAN RIGHTS | no full-text |
| Tutor Marcom, Robin; Freeman Lambar, Elizabeth; Rodman, Beth; Thomas, Gayle; Watson, Ann; Parrish, Bryan; Wilburn, Jessica | 2020 | Working along the Continuum: North Carolina's Collaborative Response to COVID-19 for Migrant & Seasonal Farmworkers | type of article |
| Tuyisenge, Germaine; Goldenberg, Shira M. | 2021 | COVID-19, structural racism, and migrant health in Canada | type of article |
| Ullah, Akm Ahsan; Nawaz, Faraha; Chattoraj, Diotima | 2021 | Locked up under lockdown: The COVID-19 pandemic and the migrant population | type of article (lack of sufficient empirical data) |

| | | | |
|---|---|---|---|
| Valsecchi M; Durante R. | 2021 | Internal migration networks and mortality in home communities: Evidence from Italy during the Covid-19 pandemic | population |
| Van Hout MC; Bigland C; Murray N. | 2021 | Scoping the impact of COVID-19 on the nexus of statelessness and health in Council of Europe member states | type of article (review without eligible included studies) |
| Velando, P. B.; Madar, A. A.; Czapka, E.; Herrero-Arias, R.; Haj-Younes, J.; Hasha, W.; Deeb, G.; Moen, K. A.; Ortiz-Barreda, G.; Diaz, E. | 2021 | Information, trust and adherence to recommendations among migrants in Norway during the first wave of the COVID-19 pandemic | no full-text |
| Vilar-Compte, Mireya; Gaitán-Rossi, Pablo; Félix-Beltrán, Lucía; Bustamante, Arturo V. | 2021 | Pre-COVID-19 Social Determinants of Health Among Mexican Migrants in Los Angeles and New York City and Their Increased Vulnerability to Unfavorable Health Outcomes During the COVID-19 Pandemic | outcome† |
| Vilches Hinojosa, Miguel Rivas Castillo Jaime Vidal De Haymes María | 2021 | International Migration in the Central and North American Regions in the COVID-19 Pandemic Context | outcome† |
| Vilog, R. B. T.; Piocos, C. M.; Iii, | 2021 | Undocumented in the time of pandemic: exploring legal violence, health care and human rights of irregular Filipino migrants in Italy and the UK. (Special Issue: Human rights in healthcare during COVID-19 and other pandemics.) | no full-text |
| Vonen, Hanne Dahl; Olsen, Merete Lan; Eriksen, Sara Soraya; Jervelund, Signe Smith; Eikemo, Terje Andreas | 2021 | Refugee camps and COVID-19: Can we prevent a humanitarian crisis? | type of article |
| Voola, A. P.; Ray, S.; Voola, R. | 2021 | Social inclusion of migrant workers in a pandemic: employing consumer vulnerability lens to internal Indian migrant experience | no full-text |
| Wang, Qianyun Liu Jacky Ka Kei Walsh Christine Ann | 2021 | Identities: experiences and impacts of the COVID-19 pandemic from the perspectives of older Chinese immigrants in Canada | outcome† |
| Weil, Alan R. | 2021 | Borders, Immigrants, And Health | type of study |
| Wickramage, Kolitha; Ito, Chiaki; Hilali, Mahmoud; Hargreaves, Sally; Requena-Méndez, Ana | 2021 | Migrants in the Middle East and North Africa in a time of COVID-19 | type of article |
| Wong Wai Cheong, Nicholas; Meyyur Aravamudan, Veeraraghavan; Venkatachalam, Jonathen; Kuthiah, Navin | 2021 | Gastrointestinal Symptoms in Association With Hypokalemia Can Be a Predictor of Inferior Outcomes in COVID-19 | outcome† |
| Wu, M. X.; Shelar, D.; Gopalakrishnan, R.; Amin, S.; Ieee, | 2021 | Optimal Testing Strategy for Containing COVID-19: A Case-Study on Indian Migrant Worker Population | outcome† |
| Yoon, Myeong Sook; Feyissa, Israel Fisseha; Suk, So-Won | 2021 | Panic and Trust during COVID-19: A Cross-Sectional Study on Immigrants in South Korea | outcome† |
| Yozwiak, Daniel; McGuire, Tanner; Aultman, Julie M. | 2021 | The Mental Health of Refugees during a Pandemic: Striving toward Social Justice through Social Determinants of Health and Human Rights | type of study |
| Yuen, Samson; Cheng, Edmund W.; Or, Nick H. K.; Grépin, Karen A.; Fu, King-Wa; Yung, Ka-Chun; Yue, Ricci P. H. | 2021 | A tale of two city-states: A comparison of the state-led vs civil society-led responses to COVID-19 in Singapore and Hong Kong | population |
| Zajacova, Anna; Jehn, Anthony; Stackhouse, Matthew; Denice, Patrick; Ramos, Howard | 2020 | Changes in health behaviours during early COVID-19 and socio-demographic disparities: a cross-sectional analysis | outcome† |
| Zard, Monette; Lau, Ling San; Bowser, Diana M.; Fouad, Fouad M.; Lucumí, Diego I.; Samari, Goleen; Harker, Arturo; Shepard, Donald S.; Zeng, Wu; Moresky, Rachel T.; Audi, Mhd Nour; Greene, Claire M.; Kachur, S. Patrick | 2021 | Leave no one behind: ensuring access to COVID-19 vaccines for refugee and displaced populations | outcome† |
| Zhang, Meiling; Zhou, Jienan; Jia, Senquan; Zhao, Xiaonan; Chen, Yaoyao; Sun, Yanhong; Liu, Zhaosheng; Zhou, Xiaofang; Li, Duo; Luo, Chunrui; Zhang, Yong; Onsongo, Violet Magoma; Shao, Yong; Fu, Xiaoqing | 2021 | Undocumented Migrants Reintroducing COVID-19, Yunnan Province, China | type of study |

| | | | |
|---|---|---|---|
| Zhang, Q. F.; Hu, ZhanPing | 2021 | Rural China under the COVID-19 pandemic: differentiated impacts, rural-urban inequality and agro-industrialization. (Symposium: Capitalism, crisis and COVID-19: agrarian political economy and circuits of capital and labour.) | type of article |
| Zhang, Qian Forrest Hu Zhanping | 2021 | Rural China under the COVID-19 pandemic: Differentiated impacts, rural–urban inequality and agro-industrialization | type of study |
| Zhong, Zhe-Feng; Huang, Jia; Yang, Xia; Peng, Jin-Ling; Zhang, Xiao-Yan; Hu, Yang; Fu, Nian; Lin, Hai-Lian; Jiang, Bo; Tian, Ya-Ying; Yao, Hong-Yi; Deng, Li-Pu; Tang, Xiao-Qing; Zhou, Jie-Can; Tang, Jian; Xie, Xia; Liu, Qiong; Liu, Jing; Dou, Cheng-Yun; Dai, Rong-Juan; Yan, Bo; Yang, Xue-Feng | 2020 | Epidemiological and clinical characteristics of COVID-19 patients in Hengyang, Hunan Province, China | population |
| Zhou, Feng; You, Chong; Zhang, Xiaoyu; Qian, Kaihuan; Hou, Yan; Gao, Yanhui; Zhou, Xiao-Hua | 2021 | Epidemiological Characteristics and Factors Associated with Critical Time Intervals of COVID-19 in Eighteen Provinces, China: A Retrospective Study | population |
| Zlotnick, Cheryl; Dryjanska, Laura; Suckerman, Suzanne | 2021 | Factors linked to accessing COVID-19 recommendations among working migrants | focus* |
| Zlotnick, Cheryl; Dryjanska, Laura; Suckerman, Suzanne | 2021 | Health literacy, resilience and perceived stress of migrants in Israel during the COVID-19 pandemic | focus* |
| van der Velden, Peter G.; Contino, Carlo; Das, Marcel; van Loon, Peter; Bosmans, Mark W. G. | 2020 | Anxiety and depression symptoms, and lack of emotional support among the general population before and during the COVID-19 pandemic. A prospective national study on prevalence and risk factors | population |
| *: Studies with no focus on SARS-CoV-2 or the COVID-19 pandemic.<br>†: Crude cases without denominators, proportions without denominators or no reference to health outcomes. | | | |

### 4 Quality Appraisal

Tables S5 a-f show the quality appraisal for each study by group of study design based on the JBI checklists and for modelling studies based on an adapted instrument derived from existing tools as used in previous reviews.[2–7]

**Table S5a: Quality Appraisal of Cross-Sectional Studies**

| Author & Year | 1. Clear inclusion criteria | | 2. Detailed description of subjects and setting | | 3. Valid and reliable measure of Exposure(s) | | 4. Objective measure of condition(s) | | 5. Confounder identified | | 6. Confounder addressed | | 7. Valid and reliable measure of Outcome(s) | | 8. Appropriate statistical analysis | | Overall Rating | | | | Final Rating | |
|---|---|---|---|---|---|---|---|---|---|---|---|---|---|---|---|---|---|---|---|---|---|---|
| | R1 | R2 | R1 | R2 | R1 | R2 | R1 | R2 | R1 | R2 | R1 | R2 | R1 | R2 | R1 | R2 | R1 | R2 | R1 | R2 | Mean Score | Cat. |
| Acharya & Patel 2021 | yes | unclear | yes | yes | na | na | na | yes | na | na | na | na | unclear | yes | yes | yes | 75.0 | 80.0 | high | high | 77.5 | high |
| Akkaya-Kalayci et al. 2020 | yes | yes | yes | yes | yes | yes | yes | yes | no | unclear | no | unclear | yes | yes | yes | yes | 75.0 | 75.0 | high | high | 75.0 | high |
| Al Awaidy et al. 2021 | yes | yes | yes | yes | yes | yes | yes | yes | no | no | no | no | yes | yes | no | no | 62.5 | 62.5 | medium | medium | 62.5 | medium |
| Al-Awaida et al. 2021 | yes | yes | yes | yes | yes | yes | yes | yes | yes | unclear | no | unclear | yes | yes | yes | yes | 87.5 | 75.0 | high | high | 81.2 | high |
| Alali et al. 2020 | yes | yes | yes | yes | yes | no | yes | no | yes | na | no | na | yes | yes | yes | yes | 87.5 | 66.7 | high | medium | 77.1 | high |
| Alkhamisa et al. 2020 | unclear | na | no | na | yes | na | yes | na | yes | na | yes | na | yes | na | yes | na | 75.0 | 0.0 | high | low | 62.5 | medium* |
| Amengual-Moreno et al. 2020 | yes | na | yes | yes | na | na | unclear | unclear | yes | na | yes | na | yes | unclear | yes | yes | 85.7 | 50.0 | high | medium | 67.9 | medium |
| Amodio et al. 2021 | yes | yes | yes | yes | yes | yes | yes | yes | no | yes | no | yes | yes | yes | yes | yes | 75.0 | 100.0 | high | high | 87.5 | high |
| Anom et al. 2021 | unclear | yes | no | no | yes | na | yes | no | no | yes | unclear | yes | yes | yes | yes | yes | 50.0 | 71.4 | medium | medium | 60.7 | medium |
| Aragona et al. 2022 | yes | yes | yes | yes | yes | yes | yes | yes | no | no | no | no | yes | yes | no | yes | 62.5 | 75.0 | medium | high | 68.8 | medium |
| Attal et al. 2020 | yes | yes | yes | yes | na | yes | yes | yes | yes | no | yes | no | yes | yes | yes | yes | 100.0 | 75.0 | high | high | 87.5 | high |
| Bahadur et al. 2021 | na | no | na | yes | yes | yes | yes | yes | na | na | na | no | yes | yes | yes | yes | 100.0 | 71.4 | high | medium | 85.7 | high |
| Bauza et al. 2021 | yes | na | yes | na | yes | yes | yes | yes | unclear | yes | unclear | yes | yes | yes | yes | yes | 75.0 | 100.0 | high | high | 87.5 | high |
| Bolivar 2020 | unclear | unclear | yes | no | na | no | no | no | no | no | no | no | no | no | no | yes | 14.3 | 12.5 | low | low | 13.4 | low |

| | | | | | | | | | | | | | | | | | | | | | |
|---|---|---|---|---|---|---|---|---|---|---|---|---|---|---|---|---|---|---|---|---|---|
| Boogaard et al. 2021 | na | yes | na | yes | na | na | na | yes | na | yes | na | no | na | yes | na | yes | 0.0 | 85.7 | low | high | 71.4 | medium* |
| Bruni 2021 | yes | yes | yes | yes | no | na | no | na | na | no | na | no | no | yes | no | yes | 33.3 | 66.7 | low | medium | 50.0 | medium |
| Buda et al. 2020 | yes | na | na | yes | unclear | na | na | no | na | na | na | na | yes | yes | unclear | yes | 50.0 | 75.0 | medium | high | 62.5 | medium |
| Cai et al. 2021 | yes | yes | yes | yes | yes | na | yes | yes | yes | na | yes | na | yes | yes | yes | yes | 100.0 | 100.0 | high | high | 100.0 | high |
| Calderón-Larrañaga et al. 2020 | no | unclear | unclear | yes | yes | na | na | yes | unclear | yes | unclear | yes | yes | yes | yes | yes | 42.9 | 85.7 | low | high | 66.7 | medium* |
| Canevelli et al. 2020 | unclear | yes | no | yes | yes | yes | yes | yes | no | yes | no | no | yes | yes | yes | unclear | 50.0 | 75.0 | medium | high | 62.5 | medium |
| Chen et al. 2021 | no | unclear | yes | no | yes | na | yes | na | yes | yes | yes | yes | yes | yes | yes | yes | 87.5 | 66.7 | high | medium | 77.1 | high |
| Chew et al. 2020 | yes | unclear | no | yes | yes | na | yes | no | no | no | no | no | yes | unclear | unclear | yes | 50.0 | 28.6 | medium | low | 39.3 | low |
| Coyer et al. 2021 | unclear | yes | no | yes | yes | yes | yes | yes | no | yes | na | yes | yes | yes | yes | yes | 57.1 | 100.0 | medium | high | 78.6 | high |
| Cromer et al. 2020 | yes | yes | yes | yes | na | na | yes | yes | yes | yes | yes | unclear | yes | yes | yes | yes | 100.0 | 85.7 | high | high | 92.9 | high |
| Cummings et al. 2022 | yes | yes | yes | yes | yes | yes | yes | yes | yes | yes | yes | yes | yes | yes | yes | yes | 100.0 | 100.0 | high | high | 100.0 | high |
| D'Ambrosi et al. 2021 | unclear | yes | yes | yes | na | yes | no | yes | na | yes | na | yes | yes | yes | yes | yes | 60.0 | 100.0 | medium | high | 80.0 | high |
| Dale et al. 2021 | unclear | unclear | yes | yes | yes | na | yes | yes | no | yes | na | no | yes | yes | yes | no | 71.4 | 57.1 | medium | medium | 64.3 | medium |
| Das et al. 2021 | yes | yes | yes | yes | yes | na | yes | no | yes | yes | yes | yes | yes | no | yes | yes | 100.0 | 71.4 | high | medium | 85.7 | high |
| Daw et al. 2021 | no | no | no | na | yes | yes | yes | yes | no | no | no | na | yes | yes | unclear | yes | 37.5 | 66.7 | low | medium | 52.1 | medium |
| Debopadhaya et al. 2020 | unclear | no | na | no | na | yes | no | unclear | yes | yes | yes | yes | no | yes | yes | yes | 50.0 | 62.5 | medium | medium | 56.2 | medium |
| Devillanova et al. 2020 | yes | unclear | yes | no | yes | no | yes | no | na | no | na | na | yes | yes | yes | yes | 100.0 | 28.6 | high | low | 60.0 | medium* |
| Dressler et al. 2021 | yes | yes | yes | yes | no | na | yes | na | na | yes | na | no | yes | yes | yes | unclear | 83.3 | 66.7 | high | medium | 75.0 | high |
| Ebrahim et al. 2021 | na | unclear | na | yes | na | na | na | na | na | no | na | na | na | yes | na | unclear | 0.0 | 40.0 | low | low | 20.0 | low |
| Elci et al. 2021 | unclear | yes | yes | yes | na | yes | na | yes | yes | unclear | yes | unclear | no | yes | yes | yes | 66.7 | 75.0 | medium | high | 70.8 | medium |

| | | | | | | | | | | | | | | | | | | | | | | |
|---|---|---|---|---|---|---|---|---|---|---|---|---|---|---|---|---|---|---|---|---|---|---|
| Engjom et al. 2021 | yes | yes | yes | yes | yes | na | yes | na | no | na | na | na | yes | yes | yes | yes | 85.7 | 100.0 | high | high | 92.9 | high |
| Enriquez et al. 2021 | yes | yes | yes | yes | na | yes | yes | yes | yes | yes | yes | yes | unclear | yes | yes | yes | 85.7 | 100.0 | high | high | 92.9 | high |
| Fabiani et al. 2021 | yes | yes | yes | yes | yes | yes | yes | unclear | yes | yes | yes | yes | yes | yes | yes | yes | 100.0 | 87.5 | high | high | 93.8 | high |
| Fielding-Miller et al. 2020 | no | unclear | yes | yes | yes | na | na | no | yes | yes | yes | yes | yes | unclear | yes | yes | 85.7 | 57.1 | high | medium | 71.4 | medium |
| Fiorini et al. 2020 | yes | yes | yes | yes | yes | yes | yes | yes | unclear | no | unclear | no | yes | yes | yes | no | 75.0 | 62.5 | high | medium | 68.8 | medium |
| Fiorini et al. 2021 | yes | yes | yes | yes | yes | yes | yes | yes | unclear | no | unclear | no | yes | yes | yes | yes | 75.0 | 75.0 | high | high | 75.0 | high |
| Garcia et al. 2021 | unclear | unclear | yes | yes | na | yes | yes | yes | yes | no | yes | na | yes | yes | yes | yes | 85.7 | 71.4 | high | medium | 78.6 | high |
| Goodman et al. 2020 | unclear | unclear | yes | yes | yes | no | yes | yes | no | na | na | na | yes | yes | yes | yes | 71.4 | 66.7 | medium | medium | 69.0 | medium |
| Gosselin et al. 2021 | unclear | yes | yes | no | yes | yes | yes | yes | yes | yes | yes | yes | yes | yes | yes | yes | 87.5 | 87.5 | high | high | 87.5 | high |
| Gosselin et al. 2021 | unclear | na | no | yes | na | na | yes | na | yes | yes | yes | yes | unclear | yes | yes | yes | 57.1 | 100.0 | medium | high | 78.6 | high |
| Gulati et al. 2021 | yes | yes | yes | yes | yes | na | yes | na | na | yes | na | no | yes | unclear | yes | na | 100.0 | 60.0 | high | medium | 80.0 | high |
| Guttmann et al. 2020 | yes | no | yes | unclear | na | na | yes | yes | yes | yes | no | unclear | yes | yes | yes | unclear | 85.7 | 42.9 | high | low | 85.7 | high* |
| Habtamu et al. 2021 | yes | yes | yes | yes | yes | na | yes | no | unclear | yes | unclear | yes | yes | yes | yes | yes | 75.0 | 85.7 | high | high | 80.4 | high |
| Hajjar & Abu-Sittah 2021 | yes | unclear | yes | yes | yes | na | yes | unclear | unclear | no | unclear | no | yes | no | na | yes | 71.4 | 28.6 | medium | low | 50.0 | medium |
| Hamadah et al. 2020 | yes | yes | yes | yes | na | na | yes | yes | yes | yes | yes | yes | yes | yes | yes | yes | 100.0 | 100.0 | high | high | 100.0 | high |
| Hamdan et al. 2021 | yes | na | na | yes | na | na | unclear | na | yes | yes | yes | yes | no | yes | no | yes | 50.0 | 100.0 | medium | high | 75.0 | high |
| Harkness et al. 2021 | yes | yes | yes | yes | na | na | no | na | no | na | no | na | no | na | yes | yes | 42.9 | 100.0 | low | high | 42.9 | low[1] |
| Horner et al. 2021 | yes | yes | yes | yes | na | na | na | yes | yes | yes | yes | yes | yes | yes | yes | yes | 100.0 | 100.0 | high | high | 100.0 | high |
| IOM 2020 | unclear | na | no | na | unclear | unclear | no | yes | no | na | yes | na | no | yes | na | yes | 14.3 | 75.0 | low | high | 30.0 | low* |
| IOM 2020 | no | unclear | no | no | na | yes | na | yes | na | no | na | no | unclear | unclear | no | yes | 0.0 | 37.5 | low | low | 18.8 | low |
| IOM 2021 | yes | unclear | yes | yes | na | yes | yes | yes | na | no | na | no | unclear | yes | yes | yes | 80.0 | 62.5 | high | medium | 71.2 | medium |

| | | | | | | | | | | | | | | | | | | | | | | |
|---|---|---|---|---|---|---|---|---|---|---|---|---|---|---|---|---|---|---|---|---|---|---|
| IOM 2021 | unclear | yes | no | yes | no | na | yes | na | no | no | no | no | yes | yes | no | yes | 25.0 | 66.7 | low | medium | 45.8 | low |
| Im & George 2021 | yes | yes | yes | yes | yes | na | yes | na | no | na | na | na | yes | na | yes | yes | 85.7 | 100.0 | high | high | 92.9 | high |
| Imran et al. 2021 | yes | yes | no | no | no | yes | yes | yes | na | yes | na | no | yes | yes | yes | no | 66.7 | 62.5 | medium | medium | 64.6 | medium |
| Indseth et al. 2021 | yes | yes | yes | yes | na | na | yes | na | yes | yes | yes | yes | yes | yes | yes | yes | 100.0 | 100.0 | high | high | 100.0 | high |
| Ingen et al. 2021 | yes | yes | yes | yes | yes | no | unclear | no | yes | yes | yes | yes | yes | unclear | yes | yes | 87.5 | 62.5 | high | medium | 75.0 | high |
| Jahn et al. 2022 | yes | yes | yes | yes | yes | na | yes | na | no | yes | yes | yes | yes | yes | yes | yes | 87.5 | 100.0 | high | high | 93.8 | high |
| Jaqueti et al. 2020 | no | no | no | no | na | na | na | na | yes | yes | no | no | yes | yes | yes | yes | 50.0 | 50.0 | medium | medium | 50.0 | medium |
| Jehn et al. 2021 | yes | yes | yes | yes | yes | na | yes | yes | unclear | yes | unclear | yes | yes | no | yes | yes | 75.0 | 85.7 | high | high | 80.4 | high |
| Johnson-Agbakwu et al. 2021 | yes | unclear | yes | yes | yes | na | yes | yes | unclear | no | unclear | no | yes | yes | yes | yes | 75.0 | 57.1 | high | medium | 66.1 | medium |
| Khan et al. 2021 | yes | no | no | yes | na | yes | no | yes | no | unclear | no | unclear | no | yes | no | unclear | 14.3 | 50.0 | low | medium | 32.1 | low |
| Khan et al. 2021 | na | no | na | yes | na | na | na | unclear | na | unclear | na | na | na | yes | na | unclear | 0.0 | 33.3 | low | low | 16.7 | low |
| Kim et al. 2020 | unclear | unclear | no | yes | yes | na | yes | yes | unclear | no | no | no | yes | yes | unclear | yes | 37.5 | 57.1 | low | medium | 47.3 | low |
| Kjollesdal & Magnusson 2021 | yes | yes | yes | yes | na | yes | yes | yes | yes | yes | yes | yes | yes | yes | yes | yes | 100.0 | 100.0 | high | high | 100.0 | high |
| Kjollesdal et al. 2021 | yes | yes | yes | yes | na | no | yes | yes | na | yes | na | no | yes | yes | yes | no | 100.0 | 62.5 | high | medium | 81.2 | high |
| Koh 2020 | na | yes | na | no | na | na | na | unclear | na | unclear | na | no | na | yes | na | yes | 0.0 | 42.9 | low | low | 21.4 | low |
| Kondilis et al. 2021 | yes | yes | yes | yes | yes | na | yes | na | unclear | no | unclear | na | yes | yes | yes | yes | 75.0 | 80.0 | high | high | 77.5 | high |
| Kraft et al. 2021 | yes | yes | yes | yes | yes | yes | yes | yes | yes | yes | yes | unclear | yes | yes | yes | yes | 100.0 | 87.5 | high | high | 93.8 | high |
| Kumar et al. 2020 | yes | yes | yes | yes | na | yes | na | yes | no | yes | na | yes | yes | yes | unclear | yes | 60.0 | 100.0 | medium | high | 80.0 | high |
| Labberton et al. 2021 | unclear | yes | yes | yes | yes | na | yes | yes | no | yes | no | yes | yes | yes | yes | yes | 62.5 | 100.0 | medium | high | 81.2 | high |
| Lin et al. 2020 | yes | yes | yes | yes | yes | yes | na | yes | unclear | yes | no | no | yes | yes | unclear | unclear | 57.1 | 75.0 | medium | high | 66.1 | medium |

| | | | | | | | | | | | | | | | | | | | | | | |
|---|---|---|---|---|---|---|---|---|---|---|---|---|---|---|---|---|---|---|---|---|---|---|
| Lombari et al. 2021 | yes | unclear | yes | yes | yes | no | yes | no | yes | yes | yes | yes | yes | yes | yes | yes | 100.0 | 62.5 | high | medium | 81.2 | high |
| Lusk & Chandra 2021 | yes | yes | yes | yes | no | yes | yes | na | no | unclear | no | unclear | yes | yes | yes | yes | 62.5 | 71.4 | medium | medium | 67.0 | medium |
| Ly et al. 2021 | yes | unclear | yes | yes | unclear | na | yes | no | unclear | yes | unclear | yes | yes | yes | yes | yes | 62.5 | 71.4 | medium | medium | 67.0 | medium |
| Maeland et al. 2021 | yes | unclear | yes | no | na | yes | no | yes | unclear | no | unclear | no | no | yes | yes | yes | 42.9 | 50.0 | low | medium | 46.4 | low |
| Mallet et al. 2021 | unclear | yes | yes | yes | yes | yes | yes | yes | no | na | no | na | yes | yes | yes | yes | 62.5 | 100.0 | medium | high | 81.2 | high |
| Marco et al. 2021 | yes | unclear | yes | no | yes | yes | na | yes | unclear | no | na | no | yes | yes | unclear | yes | 66.7 | 50.0 | medium | medium | 58.3 | medium |
| Masai et al. 2021 | yes | yes | yes | yes | na | yes | yes | yes | unclear | na | no | na | yes | yes | yes | yes | 71.4 | 100.0 | medium | high | 85.7 | high |
| Mendez-Dominguez et al. 2020 | yes | yes | yes | unclear | yes | yes | yes | unclear | no | yes | no | yes | yes | yes | yes | yes | 75.0 | 75.0 | high | high | 75.0 | high |
| Miconi et al. 2021 | yes | yes | yes | yes | yes | na | yes | yes | yes | yes | yes | yes | yes | yes | yes | yes | 100.0 | 100.0 | high | high | 100.0 | high |
| Miconi et al. 2021 | yes | yes | yes | yes | na | yes | yes | yes | yes | no | yes | no | yes | yes | yes | yes | 100.0 | 75.0 | high | high | 87.5 | high |
| Ming & De Jong 2021 | yes | yes | yes | yes | yes | yes | yes | yes | no | no | no | na | yes | no | yes | yes | 75.0 | 71.4 | high | medium | 73.2 | medium |
| Mishra et al. 2021 | yes | yes | yes | yes | yes | yes | yes | yes | yes | no | yes | no | yes | yes | yes | yes | 100.0 | 75.0 | high | high | 87.5 | high |
| Moran et al. 2021 | yes | unclear | yes | no | yes | yes | yes | yes | yes | no | yes | no | yes | yes | yes | yes | 100.0 | 50.0 | high | medium | 75.0 | high |
| Nair et al. 2021 | yes | yes | yes | yes | yes | yes | yes | yes | yes | yes | unclear | yes | yes | na | yes | yes | 87.5 | 100.0 | high | high | 93.8 | high |
| Ngiam et al. 2020 | yes | yes | yes | yes | unclear | yes | yes | yes | yes | yes | yes | yes | yes | yes | yes | yes | 87.5 | 100.0 | high | high | 93.8 | high |
| Norman et al. 2020 | unclear | yes | yes | yes | unclear | yes | yes | yes | yes | no | yes | na | yes | yes | yes | yes | 75.0 | 85.7 | high | high | 80.4 | high |
| Ofuoku et al. 2021 | yes | no | yes | yes | yes | yes | yes | na | na | yes | na | unclear | yes | yes | yes | unclear | 100.0 | 57.1 | high | medium | 78.6 | high |
| Oluyomi et al. 2021 | yes | yes | no | yes | na | na | na | yes | yes | yes | yes | yes | yes | yes | yes | yes | 83.3 | 100.0 | high | high | 91.7 | high |
| Ornelas et al. 2021 | yes | yes | yes | yes | na | na | unclear | yes | yes | no | na | no | unclear | yes | na | yes | 60.0 | 71.4 | medium | medium | 65.7 | medium |
| Pagani et al. 2021 | yes | yes | yes | yes | yes | na | yes | yes | yes | yes | yes | yes | yes | yes | yes | yes | 100.0 | 100.0 | high | high | 100.0 | high |

| | | | | | | | | | | | | | | | | | | | | | |
|---|---|---|---|---|---|---|---|---|---|---|---|---|---|---|---|---|---|---|---|---|---|
| Pan et al. 2021 | unclear | na | yes | yes | na | yes | no | yes | yes | na | no | na | no | yes | no | yes | 28.6 | 100.0 | low | high | 57.1 | medium* |
| Quandt et al. 2021 | yes | yes | yes | yes | yes | yes | yes | yes | na | yes | na | yes | yes | yes | yes | yes | 100.0 | 100.0 | high | high | 100.0 | high |
| Rabe et al. 2021 | yes | yes | yes | yes | yes | na | yes | no | yes | yes | yes | yes | yes | no | yes | yes | 100.0 | 71.4 | high | medium | 85.7 | high |
| Rahaman et al. 2021 | unclear | unclear | yes | no | na | na | na | no | no | na | no | na | no | no | yes | no | 33.3 | 0.0 | low | low | 16.7 | low |
| Ramirez et al. 2020 | yes | yes | yes | yes | yes | na | yes | yes | yes | yes | yes | yes | yes | yes | yes | yes | 100.0 | 100.0 | high | high | 100.0 | high |
| Ravens-Sieberer et al. 2020 | yes | yes | yes | yes | na | yes | yes | yes | yes | unclear | yes | unclear | yes | yes | yes | yes | 100.0 | 75.0 | high | high | 87.5 | high |
| Rayan et al. 2021 | yes | yes | yes | yes | na | na | unclear | yes | yes | yes | unclear | no | yes | yes | unclear | yes | 57.1 | 85.7 | medium | high | 71.4 | medium |
| Raynor & Panza 2021 | no | unclear | yes | yes | yes | na | yes | no | yes | yes | yes | yes | yes | no | yes | yes | 87.5 | 57.1 | high | medium | 72.3 | medium |
| Redditt et al. 2020 | yes | yes | yes | yes | yes | na | yes | na | no | no | no | no | yes | yes | no | yes | 62.5 | 66.7 | medium | medium | 64.6 | medium |
| Salem et al. 2021 | yes | yes | yes | yes | na | na | yes | unclear | yes | yes | yes | yes | yes | yes | yes | yes | 100.0 | 85.7 | high | high | 92.9 | high |
| Santos et al. 2021 | yes | yes | yes | yes | yes | yes | yes | yes | na | na | na | na | yes | yes | yes | yes | 100.0 | 100.0 | high | high | 100.0 | high |
| Saw et al. 2021 | yes | yes | yes | yes | yes | no | yes | no | yes | yes | yes | yes | yes | yes | yes | yes | 100.0 | 75.0 | high | high | 87.5 | high |
| Schestag et al. 2021 | na | unclear | na | yes | na | na | na | no | na | no | na | no | na | yes | na | no | 0.0 | 28.6 | low | low | 14.3 | low |
| Sharov 2021 | na | na | na | na | na | na | na | na | na | na | na | na | na | na | na | na | 0.0 | 0.0 | low | low | 0.0 | low |
| Shimizutani& Yamada 2021 | yes | unclear | yes | yes | na | na | na | no | unclear | no | no | no | yes | no | yes | yes | 66.7 | 28.6 | medium | low | 47.6 | low |
| Silva et al. 2021 | unclear | yes | no | yes | yes | unclear | yes | yes | no | yes | unclear | unclear | yes | yes | yes | no | 50.0 | 62.5 | medium | medium | 56.2 | medium |
| Song et al. 2021 | unclear | yes | na | yes | no | yes | no | yes | yes | no | yes | no | no | yes | yes | yes | 42.9 | 75.0 | low | high | 85.7 | high* |
| Sorg et al. 2022 | yes | yes | yes | yes | na | na | yes | yes | yes | yes | yes | yes | yes | yes | yes | yes | 100.0 | 100.0 | high | high | 100.0 | high |
| Soussy et al. 2021 | unclear | unclear | yes | yes | yes | no | yes | no | no | no | no | no | yes | yes | unclear | no | 50.0 | 25.0 | medium | low | 37.5 | low |
| Strully et al. 2020 | na | unclear | yes | yes | na | unclear | na | unclear | yes | yes | yes | yes | na | unclear | yes | yes | 100.0 | 50.0 | high | medium | 75.0 | high |

| | | | | | | | | | | | | | | | | | | | | | |
|---|---|---|---|---|---|---|---|---|---|---|---|---|---|---|---|---|---|---|---|---|---|
| Sundaram et al. 2021 | yes | yes | yes | yes | yes | na | yes | na | yes | yes | yes | yes | yes | yes | yes | yes | 100.0 | 100.0 | high | high | 100.0 | high |
| Sundaram et al. 2021 | yes | yes | yes | yes | no | na | no | yes | yes | yes | yes | yes | yes | yes | yes | yes | 75.0 | 100.0 | high | high | 87.5 | high |
| Tan et al. 2020 | yes | yes | no | yes | no | yes | yes | yes | unclear | no | unclear | na | yes | yes | yes | unclear | 50.0 | 71.4 | medium | medium | 60.7 | medium |
| Tramuto et al. 2021 | unclear | yes | yes | unclear | no | na | yes | na | no | unclear | no | na | yes | yes | yes | yes | 50.0 | 60.0 | medium | medium | 55.0 | medium |
| Turunen et al. 2021 | yes | na | yes | no | na | na | unclear | na | yes | na | no | na | yes | yes | unclear | yes | 57.1 | 66.7 | medium | medium | 61.9 | medium |
| Tzoraki et al. 2021 | unclear | yes | yes | yes | na | yes | no | yes | no | unclear | no | unclear | no | yes | no | yes | 14.3 | 75.0 | low | high | 33.3 | low* |
| Van de Velde et al. 2021 | yes | yes | yes | yes | yes | unclear | yes | no | yes | yes | yes | yes | yes | yes | yes | yes | 100.0 | 75.0 | high | high | 87.5 | high |
| Vanthomme et al. 2021 | yes | yes | yes | yes | yes | yes | unclear | yes | yes | yes | yes | yes | yes | yes | yes | yes | 87.5 | 100.0 | high | high | 93.8 | high |
| Varkey et al. 2021 | yes | yes | yes | yes | yes | na | yes | na | unclear | yes | unclear | unclear | yes | yes | na | yes | 71.4 | 83.3 | medium | high | 77.4 | high |
| Wankhede et al. 2021 | no | unclear | unclear | no | yes | no | na | no | no | yes | no | yes | yes | no | unclear | unclear | 43.5 | 25.0 | low | low | 34.3 | low |
| Wiedmeyer et al. 2021 | yes | yes | yes | yes | na | na | na | yes | yes | na | no | na | yes | yes | unclear | yes | 66.7 | 100.0 | medium | high | 83.3 | high |
| Xia et al. 2022 | yes | yes | yes | yes | na | yes | unclear | yes | unclear | unclear | unclear | unclear | yes | yes | yes | yes | 57.1 | 75.0 | medium | high | 66.1 | medium |
| Yang et al. 2021 | unclear | no | yes | yes | unclear | unclear | yes | unclear | yes | yes | yes | yes | yes | yes | yes | yes | 75.0 | 62.5 | high | medium | 68.8 | medium |
| Yi et al. 2021 | unclear | unclear | yes | yes | na | na | na | na | na | na | na | na | yes | yes | yes | yes | 75.0 | 75.0 | high | high | 75.0 | high |
| Yueping et al. 2021 | unclear | yes | yes | yes | no | yes | no | yes | yes | yes | yes | yes | no | yes | yes | yes | 50.0 | 100.0 | medium | high | 75.0 | high |
| Yuguero et al. 2021 | yes | yes | yes | yes | na | na | yes | yes | na | na | na | na | unclear | yes | yes | unclear | 80.0 | 80.0 | high | high | 80.0 | high |
| Zhang et al. 2020 | unclear | yes | yes | yes | yes | na | yes | yes | yes | yes | yes | yes | yes | yes | yes | yes | 87.5 | 100.0 | high | high | 93.8 | high |
| Zhang et al. 2021 | yes | yes | yes | yes | yes | na | yes | yes | no | yes | no | yes | yes | yes | yes | yes | 75.0 | 100.0 | high | high | 87.5 | high |
| Zhang et al. 2021 | unclear | yes | no | yes | yes | yes | yes | yes | no | yes | yes | yes | yes | yes | yes | yes | 62.5 | 100.0 | medium | high | 81.2 | high |

*: assessed quality based on third reviewer agreement; R1: Rater 1; R2: Rater 2

### Table S5b: Quality Appraisal of Cohort Studies

| Author & Year | 1. Two groups recruited from the same population | | 2. Exposure measured similar for two groups | | 3. Exposure measured valid reliable | | 4. Confounding factors identified | | 5. Strategies to deal with confounding factors stated | | 6. Groups were free of the outcome at the start of the study | | 7. Outcomes measured in a valid and reliable way | | 8. Follow up time reported and sufficient | | 9. Follow up complete or reasons to loss to follow up described and explored | | 10. Strategies to address incomplete follow up utilized | | 11. Appropriate statistical analysis used | | Overall Rating | | | | Final Rating | |
|---|---|---|---|---|---|---|---|---|---|---|---|---|---|---|---|---|---|---|---|---|---|---|---|---|---|---|---|---|
| | R1 | R2 | R1 | R2 | R1 | R2 | R1 | R2 | R1 | R2 | R1 | R2 | R1 | R2 | R1 | R2 | R1 | R2 | R1 | R2 | R1 | R2 | R1 | R2 | R1 | R2 | Mean Score | Cat. |
| Aradhya et al. 2021 | yes | yes | yes | no | yes | na | yes | yes | yes | yes | yes | yes | yes | yes | yes | yes | no | yes | no | na | yes | yes | 81.8 | 88.9 | high | high | 85.4 | high |
| Brinkmann et al. 2021 | yes | yes | yes | yes | na | yes | yes | yes | yes | yes | na | yes | yes | yes | no | yes | no | unclear | no | unclear | yes | no | 66.7 | 72.7 | medium | medium | 69.7 | medium |
| Casanova et al. 2022 | unclear | yes | na | yes | na | yes | unclear | no | no | no | na | yes | yes | yes | yes | yes | unclear | unclear | na | unclear | yes | yes | 42.9 | 63.6 | low | medium | 53.2 | medium |
| Debopadhaya et al. 2021 | yes | yes | yes | unclear | na | yes | yes | yes | yes | yes | yes | yes | yes | yes | na | yes | na | unclear | na | unclear | yes | yes | 100.0 | 72.7 | high | medium | 86.4 | high |
| Diaz-Menendez et al. 2021 | yes | yes | yes | unclear | yes | unclear | no | yes | no | yes | yes | yes | yes | yes | unclear | yes | unclear | unclear | unclear | unclear | yes | yes | 54.5 | 63.6 | medium | medium | 59.1 | medium |
| Donati et al. 2021 | yes | yes | yes | yes | yes | yes | no | yes | no | na | yes | yes | yes | yes | yes | yes | yes | unclear | na | unclear | yes | unclear | 80.0 | 70.0 | high | medium | 75.0 | high |
| Drefahl et al. 2020 | yes | no | na | unclear | yes | na | na | yes | yes | na | unclear | yes | yes | no | yes | yes | yes | unclear | na | no | yes | yes | 87.5 | 44.4 | high | low | 66.7 | medium* |
| Efani et al. 2021 | no | yes | na | yes | yes | no | na | yes | na | na | yes | yes | yes | yes | na | unclear | na | na | na | unclear | yes | yes | 80.0 | 66.7 | high | medium | 73.3 | medium |
| Giorgi et al. 2020 | na | yes | na | na | yes | na | yes | yes | yes | yes | yes | yes | yes | no | yes | yes | yes | yes | na | no | yes | yes | 100.0 | 77.8 | high | high | 88.9 | high |
| Gorny et al. 2021 | no | yes | yes | yes | no | no | no | yes | no | no | unclear | yes | yes | yes | yes | yes | yes | yes | na | na | yes | yes | 50.0 | 80.0 | medium | high | 65.0 | medium |
| Gosselin et al. 2021 | yes | yes | yes | yes | no | yes | yes | unclear | no | unclear | yes | yes | yes | yes | yes | yes | yes | unclear | na | unclear | yes | yes | 80.0 | 63.6 | high | medium | 71.8 | medium |
| Guijarro et al. 2021 | yes | yes | yes | yes | no | yes | yes | yes | unclear | unclear | yes | yes | yes | yes | yes | yes | yes | yes | unclear | na | yes | yes | 72.7 | 90.0 | medium | high | 81.4 | high |
| Holmberg et al. 2022 | yes | yes | yes | yes | na | na | yes | yes | yes | no | yes | na | no | yes | yes | yes | yes | yes | no | yes | yes | yes | 80.0 | 88.9 | high | high | 84.4 | high |
| Inghammar et al. 2021 | yes | yes | yes | yes | yes | yes | no | unclear | no | unclear | yes | yes | yes | yes | yes | yes | yes | yes | yes | na | yes | yes | 81.8 | 80.0 | high | high | 80.9 | high |

| | | | | | | | | | | | | | | | | | | | | | | | | | | | | |
|---|---|---|---|---|---|---|---|---|---|---|---|---|---|---|---|---|---|---|---|---|---|---|---|---|---|---|---|---|
| Ingraham et al. 2021 | yes | yes | yes | yes | na | yes | yes | yes | yes | yes | yes | yes | no | yes | yes | yes | yes | yes | na | na | yes | yes | 88.9 | 100.0 | high | high | 94.4 | high |
| Kizilhan & Noll-Hussong 2020 | na | yes | na | yes | na | no | na | yes | na | no | na | na | na | yes | na | yes | na | unclear | na | no | na | yes | 0.0 | 60.0 | low | medium | 30.0 | low |
| Lehmann et al. 2021 | yes | yes | yes | yes | yes | na | unclear | unclear | unclear | unclear | yes | na | yes | yes | yes | na | unclear | na | unclear | na | yes | unclear | 63.6 | 50.0 | medium | medium | 56.8 | medium |
| Logie et al. 2021 | yes | yes | yes | yes | unclear | yes | yes | unclear | yes | unclear | na | yes | yes | yes | yes | yes | yes | yes | na | unclear | yes | yes | 88.9 | 72.7 | high | medium | 80.8 | high |
| Lopez-Mendoza et al. 2021 | no | yes | yes | yes | na | na | unclear | yes | unclear | na | na | yes | yes | yes | na | unclear | na | yes | na | na | yes | yes | 50.0 | 87.5 | medium | high | 68.8 | medium |
| Martin-Sanchez et al. 2021 | yes | yes | yes | yes | yes | yes | yes | no | yes | no | yes | yes | yes | yes | unclear | yes | na | yes | na | yes | yes | yes | 88.9 | 81.8 | high | high | 85.4 | high |
| Mema et al. 2021 | na | yes | na | yes | na | no | yes | yes | no | na | na | yes | yes | yes | yes | yes | unclear | yes | unclear | na | unclear | yes | 42.9 | 88.9 | low | high | 42.9 | low* |
| Methi et al. 2021 | yes | yes | yes | yes | na | yes | yes | yes | yes | yes | yes | yes | yes | yes | yes | yes | yes | yes | na | na | yes | yes | 100.0 | 100.0 | high | high | 100.0 | high |
| Moenkemoeller et al. 2021 | yes | na | yes | na | yes | na | yes | na | yes | na | unclear | na | yes | na | yes | na | na | na | na | na | yes | na | 88.9 | 87.5 | high | high | 88.2 | high |
| Moya et al. 2021 | yes | yes | yes | yes | yes | yes | yes | no | yes | no | yes | yes | yes | yes | yes | yes | yes | no | na | no | yes | yes | 100.0 | 63.6 | high | medium | 81.8 | high |
| Naumann et al. 2021 | yes | yes | yes | yes | yes | yes | yes | yes | yes | yes | yes | unclear | yes | yes | yes | yes | no | unclear | no | unclear | yes | yes | 81.8 | 72.7 | high | medium | 77.3 | high |
| Ngiam et al. 2021 | yes | yes | yes | no | na | yes | yes | yes | yes | yes | yes | yes | yes | yes | yes | yes | yes | yes | no | na | yes | yes | 90.0 | 90.0 | high | high | 90.0 | high |
| Nicholas et al. 2021 | yes | yes | yes | yes | no | yes | yes | yes | yes | yes | yes | yes | yes | yes | yes | na | yes | na | unclear | na | yes | yes | 81.8 | 100.0 | high | high | 90.9 | high |
| Otto et al. 2021 | yes | no | yes | yes | na | no | yes | no | yes | no | yes | no | yes | yes | yes | yes | na | yes | na | no | yes | no | 100.0 | 36.4 | high | low | 100.0 | high* |
| Rostila et al. 2021 | yes | yes | yes | yes | yes | yes | yes | yes | yes | yes | yes | unclear | yes | yes | yes | yes | yes | yes | na | no | yes | yes | 100.0 | 81.8 | high | high | 90.9 | high |
| Tee et al. 2020 | yes | yes | yes | na | no | yes | yes | unclear | no | unclear | unclear | yes | yes | yes | no | yes | na | unclear | na | unclear | yes | yes | 55.6 | 60.0 | medium | medium | 57.8 | medium |
| Terp et al. 2021 | yes | na | yes | na | yes | no | no | yes | no | no | yes | yes | yes | yes | no | yes | no | no | no | no | yes | no | 54.5 | 44.4 | medium | low | 49.5 | low |
| Warszawski et al. 2022 | unclear | yes | yes | yes | yes | yes | yes | yes | yes | yes | yes | yes | yes | yes | yes | yes | no | no | yes | yes | yes | yes | 81.8 | 90.9 | high | high | 86.4 | high |
| Zlot et al. 2021 | yes | yes | yes | yes | yes | yes | no | yes | na | yes | yes | yes | yes | yes | yes | na | no | na | unclear | na | yes | yes | 70.0 | 100.0 | medium | high | 85.0 | high |

*: assessed quality based on third reviewer agreement; R1: Rater 1; R2: Rater 2

### Table S5c: Quality Appraisal of Qualitative Studies

| Author & Year | 1. Congruity between the stated philosophical perspective and the research methodology | | 2. Congruity between the research methodology and the research question or objectives | | 3. Congruity between the research methodology and the methods used to collect data | | 4. Congruity between the research methodology and the representation and analysis of data | | 5. Congruity between the research methodology and the interpretation of results | | 6. Transparent culturally or theoretically position stated | | 7. Influence of the researcher on the research, and vice-versa, addressed | | 8. Participants adequately represented | | 9. Evidence of ethical approval and ethical methodology | | 10. Conclusions drawn flow from the analysis, or interpretation of the data | | Overall Rating | | | | Final Rating | |
|---|---|---|---|---|---|---|---|---|---|---|---|---|---|---|---|---|---|---|---|---|---|---|---|---|---|---|
| | R1 | R2 | R1 | R2 | R1 | R2 | R1 | R2 | R1 | R2 | R1 | R2 | R1 | R2 | R1 | R2 | R1 | R2 | R1 | R2 | R1 | R2 | R1 | R2 | Mean Score | Cat. |
| Acharya & Patel 2021 | no | unclear | unclear | yes | unclear | yes | unclear | unclear | unclear | na | no | no | no | no | yes | yes | no | no | yes | yes | 20.0 | 44.4 | low | low | 32.2 | low |
| Avalos Cortez & van Blerk 2021 | no | no | yes | unclear | unclear | unclear | yes | unclear | yes | no | no | no | no | no | yes | yes | no | no | yes | unclear | 50.0 | 10.0 | medium | low | 30.0 | low |
| Babuc 2021 | yes | yes | yes | yes | yes | unclear | yes | unclear | yes | unclear | no | unclear | no | no | yes | yes | yes | yes | yes | yes | 80.0 | 50.0 | high | medium | 65.0 | medium |
| Bahadur et al. 2021 | yes | yes | yes | yes | yes | yes | yes | yes | yes | yes | unclear | yes | unclear | yes | yes | yes | yes | yes | yes | yes | 80.0 | 100.0 | high | high | 90.0 | high |
| Bauza et al. 2021 | unclear | yes | yes | yes | yes | yes | yes | yes | yes | yes | yes | unclear | no | unclear | yes | yes | yes | no | yes | yes | 80.0 | 70.0 | high | medium | 75.0 | high |
| Biddle et al. 2021 | no | no | yes | yes | yes | na | yes | yes | yes | yes | no | no | no | no | no | yes | yes | yes | unclear | yes | 50.0 | 66.7 | medium | medium | 58.3 | medium |
| Bojorquez et al. 2021 | yes | yes | no | yes | yes | yes | yes | yes | yes | yes | no | yes | no | unclear | yes | yes | yes | yes | yes | yes | 70.0 | 90.0 | medium | high | 80.0 | high |
| Bonizzoni & Dotsey 2021 | yes | no | yes | yes | yes | yes | unclear | yes | unclear | na | no | no | no | no | yes | yes | no | no | unclear | yes | 40.0 | 55.6 | low | medium | 47.8 | low |
| Boogaard et al. 2021 | no | na | unclear | na | unclear | na | unclear | na | unclear | na | no | na | no | na | yes | na | no | na | na | na | 11.1 | 0.0 | low | low | 37.5 | low |
| Chander et al. 2021 | no | no | yes | unclear | unclear | unclear | yes | yes | yes | unclear | yes | no | yes | yes | no | unclear | no | no | yes | yes | 60.0 | 30.0 | medium | low | 45.0 | low |
| Da Mosto et al. 2021 | yes | no | yes | no | yes | yes | yes | yes | yes | yes | no | no | unclear | no | yes | yes | yes | yes | yes | yes | 80.0 | 60.0 | high | medium | 70.0 | medium |
| DeCarlo et al. 2021 | yes | no | unclear | yes | yes | yes | yes | yes | yes | yes | unclear | no | unclear | unclear | yes | yes | yes | yes | yes | yes | 70.0 | 70.0 | medium | medium | 70.0 | medium |
| Duggal et al. 2021 | yes | yes | yes | yes | yes | yes | yes | yes | yes | yes | yes | yes | yes | yes | yes | yes | yes | yes | yes | yes | 100.0 | 100.0 | high | high | 100.0 | high |

| | | | | | | | | | | | | | | | | | | | | | | | | | | |
|---|---|---|---|---|---|---|---|---|---|---|---|---|---|---|---|---|---|---|---|---|---|---|---|---|---|---|
| Elci et al. 2021 | no | yes | unclear | yes | unclear | yes | unclear | yes | unclear | yes | no | unclear | no | unclear | yes | yes | no | unclear | yes | yes | 20.0 | 70.0 | low | medium | 45.0 | low |
| Enriquez et al. 2021 | yes | yes | yes | yes | unclear | yes | yes | yes | yes | yes | yes | no | no | no | yes | yes | no | yes | yes | yes | 70.0 | 80.0 | medium | high | 75.0 | high |
| Filippi & Giliberti 2021 | no | no | yes | yes | yes | yes | yes | yes | yes | yes | no | no | no | no | yes | yes | no | no | yes | yes | 60.0 | 60.0 | medium | medium | 60.0 | medium |
| Gele et al. 2022 | yes | no | yes | na | unclear | yes | yes | yes | yes | yes | unclear | no | yes | no | yes | yes | yes | yes | yes | yes | 80.0 | 66.7 | high | medium | 73.3 | medium |
| Geuijen et al. 2021 | no | yes | no | yes | yes | yes | yes | yes | yes | yes | no | unclear | no | no | yes | yes | yes | yes | no | yes | 50.0 | 80.0 | medium | high | 65.0 | medium |
| Guruge et al. 2020 | yes | yes | yes | yes | yes | yes | yes | yes | yes | yes | unclear | no | yes | no | yes | yes | yes | yes | yes | yes | 90.0 | 80.0 | high | high | 85.0 | high |
| Hargreaves et al. 2021 | na | no | na | yes | na | unclear | na | unclear | na | yes | na | na | na | na | na | na | na | na | na | yes | 0.0 | 50.0 | low | medium | 25.0 | low |
| Hari et al. 2021 | yes | yes | yes | yes | yes | yes | yes | yes | yes | yes | yes | no | na | yes | yes | yes | na | no | yes | yes | 100.0 | 80.0 | high | high | 90.0 | high |
| IOM 2020 | yes | no | yes | yes | yes | yes | unclear | yes | unclear | yes | no | unclear | no | unclear | yes | yes | no | no | no | yes | 40.0 | 60.0 | low | medium | 50.0 | medium |
| Im & George 2021 | yes | yes | yes | yes | yes | yes | yes | yes | yes | yes | no | no | no | no | yes | yes | no | no | yes | yes | 70.0 | 70.0 | medium | medium | 70.0 | medium |
| Kaur-Gill et al. 2021 | yes | yes | yes | yes | yes | yes | yes | yes | yes | yes | yes | no | no | no | yes | yes | yes | no | yes | yes | 90.0 | 70.0 | high | medium | 80.0 | high |
| Knights, et al. 2021 | no | yes | yes | yes | yes | yes | yes | yes | yes | yes | yes | yes | yes | yes | yes | yes | yes | yes | yes | yes | 90.0 | 100.0 | high | high | 95.0 | high |
| Kuhlmann et al. 2020 | yes | yes | yes | yes | yes | yes | yes | yes | yes | yes | unclear | no | unclear | no | na | na | na | na | yes | yes | 75.0 | 75.0 | high | high | 75.0 | high |
| Kukreja 2021 | no | yes | unclear | yes | yes | unclear | yes | unclear | yes | unclear | no | yes | no | no | yes | yes | no | no | yes | unclear | 50.0 | 40.0 | medium | low | 45.0 | low |
| Kumar et al. 2021 | yes | yes | yes | yes | yes | yes | yes | yes | yes | yes | unclear | no | unclear | no | yes | yes | no | no | yes | yes | 70.0 | 70.0 | medium | medium | 70.0 | medium |
| Lauzardo et al. 2021 | unclear | no | yes | yes | unclear | yes | unclear | yes | unclear | unclear | no | no | no | no | no | no | yes | yes | unclear | yes | 20.0 | 50.0 | low | medium | 35.0 | low |
| Li et al. 2021 | no | yes | unclear | yes | unclear | yes | unclear | yes | unclear | yes | yes | no | no | na | yes | yes | no | no | yes | yes | 30.0 | 77.8 | low | high | 87.5 | medium* |
| Lui et al. 2021 | no | yes | yes | yes | yes | yes | yes | yes | yes | yes | no | unclear | no | unclear | yes | yes | yes | yes | yes | yes | 70.0 | 80.0 | medium | high | 75.0 | high |
| Lusambili et al. 2020 | no | yes | yes | yes | unclear | yes | no | yes | no | yes | no | no | no | no | yes | yes | yes | yes | no | yes | 30.0 | 80.0 | low | high | 60.0 | medium* |

| | | | | | | | | | | | | | | | | | | | | | | | | | | |
|---|---|---|---|---|---|---|---|---|---|---|---|---|---|---|---|---|---|---|---|---|---|---|---|---|---|---|
| Martin-Anatias et al. 2021 | yes | yes | yes | yes | yes | yes | unclear | yes | na | yes | unclear | unclear | unclear | unclear | yes | yes | no | no | yes | yes | 55.6 | 70.0 | medium | medium | 62.8 | medium |
| Martuscelli 2021 | yes | yes | yes | yes | yes | yes | yes | yes | yes | yes | no | yes | yes | unclear | yes | yes | no | yes | yes | yes | 80.0 | 90.0 | high | high | 85.0 | high |
| Mookerjee et al. 2021 | yes | yes | yes | yes | yes | yes | yes | yes | yes | yes | no | yes | no | no | yes | yes | yes | no | yes | yes | 80.0 | 80.0 | high | high | 80.0 | high |
| Nasol & Francisco-Menchavez 2021 | yes | yes | yes | yes | yes | yes | yes | yes | yes | yes | no | yes | no | unclear | yes | yes | no | na | yes | na | 70.0 | 87.5 | medium | high | 80.0 | high* |
| Pal et al. 2020 | yes | no | yes | yes | yes | yes | yes | yes | yes | yes | no | no | no | no | yes | yes | yes | no | yes | yes | 80.0 | 60.0 | high | medium | 70.0 | medium |
| Qi et al. 2021 | yes | yes | yes | yes | yes | yes | yes | yes | yes | yes | yes | no | unclear | no | yes | yes | no | no | yes | yes | 80.0 | 70.0 | high | medium | 75.0 | high |
| Rao et al. 2020 | yes | no | yes | yes | yes | yes | yes | unclear | yes | no | no | no | no | no | yes | no | yes | yes | yes | unclear | 80.0 | 30.0 | high | low | 30.0 | low* |
| Reynolds et al. 2021 | yes | yes | yes | unclear | yes | yes | yes | yes | yes | yes | no | no | unclear | no | yes | yes | unclear | no | yes | unclear | 70.0 | 50.0 | medium | medium | 60.0 | medium |
| Rocha et al. 2021 | unclear | yes | yes | yes | yes | unclear | unclear | unclear | unclear | unclear | no | unclear | unclear | unclear | yes | yes | yes | yes | no | yes | 40.0 | 50.0 | low | medium | 45.0 | low |
| Sabar et al. 2021 | yes | yes | yes | yes | yes | yes | yes | yes | yes | yes | no | no | no | no | yes | yes | unclear | no | yes | yes | 70.0 | 70.0 | medium | medium | 70.0 | medium |
| Sanfelici 2021 | no | yes | yes | yes | no | unclear | yes | yes | unclear | yes | no | no | no | unclear | no | yes | unclear | unclear | yes | yes | 30.0 | 60.0 | low | medium | 45.0 | low |
| Sanna 2021 | yes | yes | yes | yes | yes | unclear | yes | yes | yes | unclear | no | no | no | no | yes | yes | na | unclear | yes | yes | 77.8 | 50.0 | high | medium | 63.9 | medium |
| Sano et al. 2021 | no | yes | yes | yes | unclear | unclear | yes | yes | yes | yes | no | unclear | no | no | unclear | yes | unclear | no | yes | yes | 40.0 | 60.0 | low | medium | 50.0 | medium |
| Schestag et al. 2021 | unclear | yes | unclear | yes | unclear | yes | unclear | yes | unclear | yes | no | no | no | no | yes | yes | no | no | yes | yes | 20.0 | 70.0 | low | medium | 45.0 | low |
| Schriro 2021 | no | no | yes | yes | yes | yes | yes | yes | yes | yes | yes | no | no | no | na | na | na | na | yes | yes | 75.0 | 62.5 | high | medium | 68.8 | medium |
| Sohel et al. 2021 | na | yes | na | yes | na | yes | na | yes | na | yes | na | no | na | no | na | yes | na | no | na | yes | 0.0 | 70.0 | low | medium | 35.0 | low |
| Srivastava et al. 2021 | yes | yes | yes | yes | yes | yes | yes | yes | yes | yes | no | no | no | no | yes | yes | no | yes | no | yes | 60.0 | 80.0 | medium | high | 70.0 | medium |
| Tang et al. 2021 | yes | yes | yes | yes | yes | yes | unclear | yes | unclear | yes | no | no | yes | no | yes | yes | no | no | yes | yes | 60.0 | 70.0 | medium | medium | 65.0 | medium |
| Thomas et al. 2021 | yes | no | yes | yes | yes | no | yes | yes | yes | yes | no | no | unclear | no | yes | yes | no | no | na | yes | 66.7 | 50.0 | medium | medium | 58.3 | medium |

| | | | | | | | | | | | | | | | | | | | | | | | | | | |
|---|---|---|---|---|---|---|---|---|---|---|---|---|---|---|---|---|---|---|---|---|---|---|---|---|---|---|
| Tosh et al. 2021 | no | yes | yes | yes | yes | yes | yes | yes | yes | yes | no | no | no | no | yes | yes | no | unclear | yes | yes | 60.0 | 70.0 | medium | medium | 65.0 | medium |
| Vosko & Spring 2021 | yes | no | yes | yes | yes | yes | unclear | yes | yes | yes | no | no | no | na | unclear | na | no | na | yes | yes | 50.0 | 71.4 | medium | medium | 60.7 | medium |
| Yee et al. 2021 | no | yes | yes | yes | yes | yes | unclear | yes | yes | yes | no | no | no | no | yes | yes | yes | yes | yes | yes | 60.0 | 80.0 | medium | high | 70.0 | medium |
| Zambrano-Barragan et al. 2021 | yes | no | unclear | yes | yes | unclear | yes | yes | yes | yes | yes | no | unclear | no | yes | yes | no | no | yes | yes | 70.0 | 50.0 | medium | medium | 60.0 | medium |
| de Diego et al. 2021 | yes | yes | yes | yes | yes | yes | yes | yes | yes | yes | no | no | no | no | yes | yes | unclear | yes | yes | yes | 70.0 | 80.0 | medium | high | 75.0 | high |

*: assessed quality based on third reviewer agreement; R1: Rater 1; R2: Rater 2

**Table S5d: Quality Appraisal of Modelling Studies**

| Author & Year | 1. structural assumptions transparent and justified | | 2. Structural assumptions reasonable | | 3. Input parameters transparent and justified | | 4. Input parameters reasonable | | 5. External validation process described? | | 6. Model is externally valid | | 7. Internal validation process described | | 8. Model is internally valid | | 9. Technical documentation is transparent | | 10. Adequate assessment of uncertainty | | Overall Rating | | | | Final Rating | |
|---|---|---|---|---|---|---|---|---|---|---|---|---|---|---|---|---|---|---|---|---|---|---|---|---|---|---|
| | R1 | R2 | R1 | R2 | R1 | R2 | R1 | R2 | R1 | R2 | R1 | R2 | R1 | R2 | R1 | R2 | R1 | R2 | R1 | R2 | R1 | R2 | R1 | R2 | Mean Score | Cat. |
| Aylett-Bullock et al. 2021 | no/ min | no/ min | no/ min | no/ min | no/ min | no/ min | no/ min | no/ min | no/ min | mod | no/ min | mod | no/ min | mod | no/ min | mod | no/ min | no/ min | no/ min | no/ min | 50.0 | 70.0 | medium | medium | 60.0 | medium |
| Doblhammer et al. 2021 | no/ min | no/ min | no/ min | no/ min | no/ min | no/ min | no/ min | no/ min | mod | no/ min | mod | na | no/ min | no/ min | no/ min | no/ min | no/ min | no/ min | mod | mod | 65.0 | 55.6 | medium | medium | 60.3 | medium |
| Fouad et al. 2021 | no/ min | no/ min | mod | no/ min | mod | no/ min | mod | no/ min | maj | no/ min | maj | no/ min | maj | no/ min | maj | no/ min | maj | no/ min | mod | no/ min | 45.0 | 50.0 | low | medium | 47.5 | low |
| Gilman et al. 2020 | no/ min | no/ min | no/ min | no/ min | no/ min | no/ min | no/ min | no/ min | maj | mod | na | mod | mod | mod | maj | mod | no/ min | no/ min | mod | no/ min | 50.0 | 70.0 | medium | medium | 60.0 | medium |
| Giorgi & Boertien 2021 | no/ min | mod | no/ min | na | no/ min | mod | no/ min | mod | maj | mod | maj | na | maj | no/ min | maj | no/ min | maj | maj | no/ min | maj | 25.0 | 62.5 | low | medium | 43.8 | low |
| Han & Jia 2021 | no/ min | no/ min | no/ min | no/ min | no/ min | no/ min | no/ min | no/ min | no/ min | mod | na | mod | maj | mod | na | mod | no/ min | no/ min | maj | maj | 37.5 | 65.0 | low | medium | 51.2 | medium |
| Kamrujjaman, et al. 2021 | mod | no/ min | mod | no/ min | no/ min | no/ min | no/ min | no/ min | no/ min | mod | no/ min | mod | no/ min | no/ min | no/ min | no/ min | na | mod | mod | no/ min | 66.7 | 65.0 | medium | medium | 65.8 | medium |
| Kassa et al. 2020 | no/ min | no/ min | no/ min | no/ min | no/ min | no/ min | no/ min | no/ min | maj | mod | maj | mod | no/ min | mod | no/ min | mod | mod | no/ min | no/ min | no/ min | 45.0 | 70.0 | low | medium | 57.5 | medium |
| Pal et al. 2021 | no/ min | mod | no/ min | mod | mod | mod | mod | mod | maj | mod | maj | mod | maj | mod | maj | mod | maj | mod | mod | mod | 40.0 | 100.0 | low | high | n.a. | high* |

| | | | | | | | | | | | | | | | | | | | | | | | | | | |
|---|---|---|---|---|---|---|---|---|---|---|---|---|---|---|---|---|---|---|---|---|---|---|---|---|---|---|
| Pascual-Garcia et al. 2021 | no/ min | no/ min | no/ min | no/ min | no/ min | no/ min | no/ min | no/ min | no/ min | mod | no/ min | mod | no/ min | mod | no/ min | mod | no/ min | no/ min | no/ min | no/ min | 50.0 | 70.0 | medium | medium | 60.0 | medium |
| Pawar et al. 2021 | no/ min | no/ min | no/ min | no/ min | no/ min | no/ min | no/ min | no/ min | maj | maj | maj | maj | maj | maj | maj | maj | no/ min | maj | no/ min | no/ min | 30.0 | 25.0 | low | low | 27.5 | low |
| Riley et al. 2021 | mod | no/ min | no/ min | no/ min | mod | no/ min | no/ min | no/ min | maj | mod | mod | mod | no/ min | mod | no/ min | mod | mod | no/ min | mod | no/ min | 70.0 | 70.0 | medium | medium | 70.0 | medium |
| Seidu 2020 | no/ min | no/ min | no/ min | no/ min | mod | no/ min | mod | no/ min | maj | maj | maj | maj | maj | mod | maj | mod | no/ min | no/ min | no/ min | mod | 40.0 | 55.0 | low | medium | 47.5 | low |
| Suphanchaimat et al. 2021 | no/ min | no/ min | no/ min | no/ min | no/ min | no/ min | no/ min | no/ min | mod | maj | maj | maj | maj | maj | maj | maj | mod | no/ min | no/ min | no/ min | 45.0 | 30.0 | low | low | 37.5 | low |
| Truelove et al. 2020 | no/ min | no/ min | no/ min | no/ min | no/ min | no/ min | no/ min | no/ min | mod | mod | mod | mod | mod | mod | mod | mod | no/ min | no/ min | no/ min | no/ min | 70.0 | 70.0 | medium | medium | 70.0 | medium |
| Truelove et al. 2021 | mod | no/ min | no/ min | no/ min | mod | no/ min | no/ min | no/ min | maj | maj | maj | maj | maj | maj | maj | maj | mod | mod | maj | no/ min | 40.0 | 35.0 | low | low | 37.5 | low |

*: assessed quality based on third reviewer agreement; R1: Rater 1; R2: Rater 2
No/ min: No or minor concerns; mod: moderate concerns; maj: major concerns.

#### Table S5e: Quality Appraisal of Case Control Studies

| Author & Year | 1. Groups comparable to each other | | 2. Appropriate matching | | 3. Identification of the same criteria for both groups | | 4. Exposure measured valid and reliable | | 5. Exposure measured in the same for both groups | | 6. Confounder identified | | 7. Adequate strategy for confounder | | 8. Outcomes assessed valid and reliable | | 9. Exposure was long enough | | 10. Appropriate statistical analysis | | Overall Rating | | | | Final Rating | |
|---|---|---|---|---|---|---|---|---|---|---|---|---|---|---|---|---|---|---|---|---|---|---|---|---|---|---|
| | **R1** | **R2** | **R1** | **R2** | **R1** | **R2** | **R1** | **R2** | **R1** | **R2** | **R1** | **R2** | **R1** | **R2** | **R1** | **R2** | **R1** | **R2** | **R1** | **R2** | **R1** | **R2** | **R1** | **R2** | **Mean Score** | **Cat.** |
| Islamoska et al. 2021 | unclear | no | no | no | yes | no | unclear | no | no | yes | yes | no | yes | no | yes | yes | no | yes | no | no | 40.0 | 30.0 | low | low | 35.0 | low |
| Kalani et al. 2021 | unclear | yes | yes | yes | yes | yes | yes | yes | yes | yes | no | yes | no | yes | no | yes | no | yes | yes | yes | 50.0 | 100.0 | medium | high | 75.0 | high |
| Sempere-Gonzalez et al. 2021 | yes | yes | yes | yes | yes | yes | yes | yes | yes | yes | yes | yes | yes | yes | yes | yes | yes | no | no | yes | 90.0 | 90.0 | high | high | 90.0 | high |
| Svensson et al. 2021 | yes | yes | yes | yes | yes | yes | yes | yes | yes | yes | yes | yes | yes | yes | yes | yes | yes | yes | unclear | yes | 90.0 | 100.0 | high | high | 95.0 | high |

R1: Rater 1; R2: Rater 2

**Table S5f: Quality Appraisal of Quasi-experimental Studies**

| Author & Year | 1. Clear distinction between 'cause' and 'effect'? | | 2. Similar inclusion of participants | | 3. Similar treatment/care of participants, other than the exposure or intervention of interest | | 4. Control group included | | 5. Pre and post measurements of the outcome | | 6. Complete follow up and adequately described and analyzed | | 7. Outcomes measured in a valid and reliable way | | 8. Outcome measured in a reliable way | | 9. Appropriate statistical analysis used | | Overall Rating | | | | Final Rating | |
|---|---|---|---|---|---|---|---|---|---|---|---|---|---|---|---|---|---|---|---|---|---|---|---|---|
| | R1 | R2 | R1 | R2 | R1 | R2 | R1 | R2 | R1 | R2 | R1 | R2 | R1 | R2 | R1 | R2 | R1 | R2 | R1 | R2 | R1 | R2 | Mean Score | Cat. |
| Alahmad et al. 2021 | yes | yes | yes | yes | yes | unclear | no | yes | yes | yes | unclear | unclear | yes | yes | yes | yes | yes | yes | 77.8 | 77.8 | high | high | 77.8 | high |
| Aragona et al. 2020 | yes | yes | yes | yes | yes | yes | no | yes | no | yes | no | yes | yes | yes | no | yes | no | yes | 44.4 | 100.0 | low | high | 75.0 | high |
| Valsecci & Durante 2021 | no | na | yes | na | no | na | yes | na | no | na | yes | na | yes | na | yes | na | yes | na | 66.7 | 100.0 | medium | high | 83.3 | high |
| R1: Rater 1; R2: Rater 2 | | | | | | | | | | | | | | | | | | | | | | | | |

### 5 Characteristics of all included studies

Tables S6 shows the main characteristics like country and period of study, migrant group, health outcome, study design, main results and conclusions (as reported) as well as the quality of the study for all included studies N = 241.

**Table S6: Study characteristics of all included Studies (N=241)**

| Authors | Year of Publication | Title | Country of study | Socioeconomic Development of Country* | Period of Study | Migrant Group | Health Outcomes | Study Design | Main results (as reported) | Conclusions (as reported) | Quality |
|---|---|---|---|---|---|---|---|---|---|---|---|
| Acharya A; Patel S. | 2021 | Vulnerabilities of internal returnee migrants in the context of the covid-19 pandemic in India | India | LMIC | June 2020 to July 2020 | Internal migrants; Retournees | mental health | Cross Sectional Study | More than half of the total migrants (52 percent, n = 118 and 34 percent, n = 77) perceived very high and high risk, respectively, to get infected of COVID-19 looking into their awareness level, living conditions, and absence of social protection measures. Nearly 50 percent (46.3 percent, n = 105) left the place of work as industry closed due to COVID-19 lockdown, and 36 percent (n = 82) migrants returned as they were deeply worried on the spatial spreading of infection. Many migrants also cited other reasons such as economic complications, fear of getting infected by the Coronavirus, family problems, and no jobs. | This study provides insight into the socio-economic vulnerability caused by the COVID-19 pandemic among internal returnee labor migrants in India. The lock-down disturbed both the supply chain and demand, causing labor markets to shrink. The sudden paralyzation of economic activities has resulted in deeper shocks and vulnerability among employers and workers. Many migrants who were employed in these sectors have lost their jobs and returned home with limited savings. As the current study indicates, migrants are absorbed mainly into the informal economy, such as on construction sites, in the textile industry, in the brick kiln industry, or as a taxi/auto-rickshaw driver in the city. The lockdown forced the migrant population to remain in a vulnerable position as it restricts them from stepping out of their home. We found that upon their return, migrant workers' social and economic conditions worsen in their native villages. | low |
| Akkaya-Kalayci T; Kothgassner OD; Wenzel T. et al. | 2020 | The Impact of the COVID-19 Pandemic on Mental Health and Psychological Well-Being of Young People Living in Austria and Turkey: A Multicenter Study | Turkey, Austria | HIC | 22 May to 19 June 2020 | International migrants | mental heatlh; effects of lockdown measures | Cross-Sectional Study | The native Turks indicated higher "anxiety"(ps<0.010), lower "vitality"(ps<0.011), and lower "general health"(ps<0.011) than native Austrians or Austrian migrants and increased "depression"(p=0.005) and lower "self-control"(p=0.022), than Austrian migrants. 0.9% of native Turks reported a decrease in their mental health status, compared to 31.1% of native Austrians and 23.7% of Austrian migrants. Participants with financial problems (OR=1.68) and prior mental health problems (i.e., already in treatment by the time of COVID-19, | Under conditions of a pandemic and measures to respond to a pandemic, youths and young adults may be particularly vulnerable to mental health problems, especially those with pre-existing psychological problems. Research and intervention strategies should also explore the situation of migrants and refugees in Turkey and other countries with severely challenged economic and health care situations. The COVID-19 pandemic impaired the psychological well-being and mental health of young people. Especially people in Turkey were most affected, | high |

| | | | | | | | | | | | |
|---|---|---|---|---|---|---|---|---|---|---|---|
| | | | | | | | | | OR=5.83) reported a higher probability for a worsening in their mental health status. | probably due to the stringent policies to fight COVID-19. | |
| Al Awaidy S; Khamis F; Al Rashidi B; Al Wahaibi AH; Albahri A; Mahomed O. | 2021 | Epidemiological Characteristics of 69,382 COVID-19 Patients in Oman | Oman | HIC | February 4, 2020 to July 23, 2020 | International migrants | health outcomes of disease: hospitalisation; ICU admissions; Mortality | Cross-Sectional Study | Patients hospitalized with COVID-19 infection were mostly Omani nationals and males between 30 and 39 years old (p<0.001). The mortality rate was 7.7 per 100,000 population (n=359) with rates of 9.4 (n=278) and 4.8 (n=81) deaths per 100,000 population in Omani nationals and foreign-born individuals, respectively. | Identifying the epidemiological characteristics and outcomes of COVID-19-infected patients is essential for developing targeted intervention strategies and preparing for the resurgence of anticipated second and third waves of this pandemic. | high |
| Alahmad B.; AlMekhled D.; Odeh A.; Albloushi D.; Gasana J. | 2021 | Disparities in excess deaths from the COVID-19 pandemic among migrant workers in Kuwait | Kuwait | HIC | 2005 to 2020 (data) | International migrants | health outcomes of disease | Ecological study | The pandemic year of 2020 clearly exhibited a large increase in mortality rate compared to the previous years. This was seen among non-Kuwaitis, increasing from 1.0 to 1.6 deaths per 1000 population. (...) An estimated 32.4% (95% CI: 29.3 to 35.6) increase in excess deaths was seen among Kuwaitis while 71.9% (95%CI: 67.8 to 76.0) were seen among non-Kuwaitis. | The burden of mortality brought on by the COVID-19 pandemic is substantially higher than what the official tally might suggest. Systematically disadvantaged migrant workers shouldered a larger burden of deaths in the pandemic year. Public health interventions must consider structural and societal determinants that give rise to the health disparities seen among migrant workers. | high |
| Alali WQ; Bastaki H; Longenecker JC et al. | 2020 | Seroprevalence of SARS-CoV-2 in migrant workers in Kuwait | Kuwait | HIC | May 23, 2020 to June 26, 2020 | Migrant workers | health outcomes of disease; risk of infection | Cross-Sectional Study | The overall prevalence of PCR-detected SARS-CoV-2 in 525 migrant workers was 21.0% (95% CI 17.7-24.7), and the seroprevalence was 38.1% (95% CI 34.0-42.3). | High SARS-CoV-2 infection prevalence and seroprevalence in low-skilled migrant workers who worked at supermarkets in Kuwait might have played a role in the disease spread in the community. The higher prevalence is likely attributed to workers' housing conditions where physical distancing opportunities are limited and intensive testing of this population. | high |
| Al-Awaida W; Al-Ameer HJ; Al-Turk H et al. | 2021 | Psychological effects of quarantine on Syrian refugees, compared to the Jordanian populations | Jordan | UMIC | not reported | Refugees | mental health | Cross-Sectional Study | The 2380 people who received the survey online demonstrated a high prevalence of PTSD, in which PTSD showed incidence of 82.5% and 66.5 in Syrian refugees and Jordanian populations, respectively. (...) This research also found that PTSD prevalence decreased when the household's monthly combined income increased; however, this reduction in PTSD prevalence is higher among the Syrian refugees than the Jordanian population. | Our research displays that quarantine can lead to more psychological distress in the form of more post-traumatic stress disorder for Syrian refugees than Jordanian people. Public health agencies, physicians of infectious diseases and psychologists must be alerted of this issue. They should work to reduce the psychological effects of quarantine and they should be able to provide additional support to Syrian refugees at high risk for the psychological and community deleterious effects of quarantine. | high |
| Alkhamisa M; AlYouhab S; Khajahc M. et al. | 2020 | Spatiotemporal dynamics of the COVID-19 pandemic in the State of Kuwait | Kuwait | HIC | February 23, 2020 to May 7, 2020 | Migrant workers | risk of infection | Cross-Sectional Study | The pandemic size in Kuwait continues to grow (R(t)s ≥2), indicating significant ongoing spread. Significant spreading and clustering events were detected among migrant workers, due to their densely | This study provided deeper insights into the epidemiology of COVID-19 in Kuwait and provided an important platform for rapid guidance of decisions related to intervention activities. | high |

| | | | | | | | | | | | |
|---|---|---|---|---|---|---|---|---|---|---|---|
| | | | | | | | | | populated areas and poor living conditions. However, the government's aggressive intervention measures have substantially lowered pandemic growth in migrant worker areas. However, at a later stage of the study period, active spreading and clustering events among both socioeconomic classes were found. | | |
| Amengual-Moreno M; Calafat-Caules M; Carot A et al. | 2020 | Social determinants of the incidence of COVID-19in Barcelona: A preliminary ecological study using public data | Spain | HIC | (data until) May 14, 2020 | International migrants (%) | health outcomes of disease: risk of infection | Ecological study | It was found that neighbourhoods belonging to the lowest quintile of income had a 42% more incidence than those belonging to the highest quintile (...). The Pearson correlation was statistically significative between the incidence of Covid-19 and the percentage of population over 75 (r=0.487), the percentage of immigration of the neighbourhood and the origin of the immigrants (r=-0.257), the AIFI (r=-0.462), the percentage of smokers (r=0.243) and the percentage of people with BMI over 25 (r=0.483). | Social determinants are correlated with the modification of the incidence of Covid-19 in the neighbourhoods of Barcelona, with special relevance of the prevalence of BMI over 25 and the percentage of immigrants and their origin. | medium |
| Amodio E; Battisti M; Maida C; Zarcone M; Casuccio A; Vitale F. | 2021 | Socio-Demographic Factors Involved in a Low-Incidence Phase of SARS-CoV-2 Spread in Sicily, Italy | Italy | HIC | February 1, 2020 to October 15, 2020 | International migrants | risk of infection | Cross-Sectional Study | A multivariable analysis found a reduced risk of SARS-CoV-2 cases was found in census sections with higher male prevalence (adj-OR = 0.99, 95% CI = 0.99-0.99;p< 0.001) and presence of immigrants (adj-OR = 0.89,95% CI 0.86-0.92;p< 0.001) | Routinely collected socio-demographic data can be predictors of SARS-CoV-2risk infection and they may have a role in mapping high risk micro-areas for virus transmission. | high |
| Anom N; Januraga P; Indrayathi P; Gesesew H; Ward P. | 2021 | Prevalence of Depression, Anxiety, and Stress Among Repatriated Indonesian Migrant Workers During the COVID-19 Pandemic | Indonesia | LMIC | June 4, 2020 to June 30, 2020 | Migrant workers | mental health | Cross-Sectional Study | This study found that the prevalence of anxiety, depression, and stress among migrant workers was 9.25, 10.15, and 2.39%, respectively. This was relatively lower than the mental health problem among the general population during the epidemic situation, which wasabout 30.3% for depression, 36.1% for anxiety, and 32.1% for stress (...). | The prevalence of depression, anxiety, and stress among repatriated Indonesian migrant workers was relatively low compared to the general population and healthcare workers. However, the focus should be given to particular groups of migrants including those whohad bad perceived health status, high perceived susceptibility, and perceived negative stigma. | medium |
| Aradhya S.; Branden M.; Drefahl S. et al. | 2021 | Intermarriage and COVID-19 mortality among immigrants. A population-based cohort study from Sweden | Sweden | HIC | March 12, 2020 to February 23, 2021 | International migrants | health outcomes of disease: mortality | Cohort Study | Our study shows that immigrants have excess COVID-19 mortality regardless of the origin of their partner, where having a Swedish-born partner is only partially protective against COVID-19 mortality among immigrants from LMIC. | (…) these findings show that lack of awareness of the Swedish recommend-ations and language barriers are not major drivers for the excess COVID-19 mortality of immigrants. At the same time, the fact that Swedes partnered with immigrants also show excess mortality compared with Swedish couples, suggests that excess | high |

| | | | | | | | | | | | |
|---|---|---|---|---|---|---|---|---|---|---|---|
| | | | | | | | | | | mortality among immigrants is explained by differential exposure to the virus. | |
| Aragona M; Tumiati M; Ferrari F et al. | 2022 | Psychopathological effects of the Coronavirus (Sars-CoV-2) imposed lockdown on vulnerable patients in treatment in a mental health outpatient department for migrants and individuals in poor socioeconomic conditions | Italy | HIC | not reported | Asylum Seekers, Refugees, Internatio nal migrants | mental health | Cross-Sectional Study | Patients who were globally considered to have worsened if compared to their pre-coronavirus mental health conditions had in fact higher scores on several mental symptoms but mild specific fears about coronavirus issues, similar to those of patients improved or stable. Worsening was significantly associated with unemployment, lack of VISA, and treatment discontinuation. Shifting the way of providing psychotherapy into a web-based modality was significantly more frequent in stable/improved patients | Findings suggest that concrete life problems and treatment discontinuation more than the coronavirus fear, have predominantly affected the mental health conditions of our patients. | medium |
| Aragona M; Barbato A; Cavani A; Constanzo G; Mirisola C. | 2020 | Negative impacts of COVID-19 lockdown on mental health service access and follow-up adherence for immigrants and individuals in socio-economic difficulties | Italy | HIC | February and March for each year from 2017 to 2020 | Internatio nal migrants | health services access | Repeated cross-sectional study | The number of patients who visited the outpatient service between February 2017 and February 2020 was continuously increasing. In March 2020, fewer patients visited the service for psychiatric interview, in line with the introduction of lockdown measures. In addition, the number of the patients who visited in February 2020 and returned for their follow-up visits in March 2020 declined from approximately 30% over the same months in 2017e2019 to 17.53% in March 2020. | The lockdown-related reduction in numbers of patients accessing the mental health service makes it difficult to help vulnerable populations during a period of time in which their mental health needs are expected to increase. Moreover, the reduction seen in follow-up compliance increases the risk of treatment discontinuation and possible relapse. Proactive alternative strategies need to be developed to reach these vulnerable populations. | medium |
| Attal J.; Lurie I.; Neumark Y. | 2020 | A rapid assessment of migrant careworkers' psychosocial status during Israel's COVID-19 lockdown | Israel | HIC | March 31, 2020 to May 4, 2020 | Migrant workers | mental health | Cross-Sectional Study | In multivariate regression, emotional distress was associated with household food insecurity (OR: 5.85;p< 0.001), lack of confidence to care for oneself and employer during the pandemic (OR: 3.85;p< 0.001), poorer general health (OR: 2.98;p< 0.003), non-Philippine country of origin (OR: 2.83;p< 0.01), female sex (OR: 2.34;p< 0.04),, and inversely associated with age (p< 0.03). While 87.6% of care workers reported having access to hand sanitization materials regularly, only 58.0% had regular access to a medical grade mask, and 21.5% reported household food insecurity. | Migrant care workers exhibited high levels of mental distress during the COVID-19 lockdown, associated with lack of confidence or resources to properly care for themselves and their employer. | high |
| Avalos Cortez E.; | 2021 | The impact of COVID-19 on older refugees | Uganda | LMIC | March 2020 to | Refugees | effect of lockdown measures | Qualitati ve research | Access to water remains a challenge for older refugees. Although older refugees have implemented strategies to try to meet | Access to, and use of, WASH services for older refugees partially improved during the COVID-19 pandemic in Uganda, although | low |

| | | | | | | | | | | | |
|---|---|---|---|---|---|---|---|---|---|---|---|
| van Blerk L. | | | | | December 2020 | | | | their WASH needs, these strategies have wider impacts on their lives. For instance, rationing water for drinking and bathing may have consequences on their nutrition and health; it also impinges on their hygiene. | some barriers remain. (...) Although a strategy on the provision of handwashing facilities to the refugee community has been implemented, there are other factors such as distance and time for fetching water that impinge on older refugees' effective access to water. | |
| Aylett-Bullock J; Cuesta-Lazaro C; Quera-Bofarull A. et al. | 2021 | Operational response simulation tool for epidemics within refugee and IDP settlements: A scenario-based case study of the Cox's Bazar settlement | Bangladesh | LMIC | not reported | Refugees and IDP | risk of infection | Modelling Study | Our findings suggest the encouraging self-isolation at home of mild to severe symptomatic patients, as opposed to the isolation of all positive cases in purpose-built isolation and treatment centers, does not increase the risk of secondary infection meaning the centers can be used to provide hospital support to the most intense cases of COVID-19. Secondly we find that mask wearing in all indoor communal areas can be effective at dampening viral spread, even with low mask efficacy and compliance rates. | Our analysis suggests that mask wearing in indoor locations can have a significant effect on disease spread, even when wearing reusable cotton masks, which the people in the settlement can make themselves. We also look at different ways to treat individuals who only have milder symptoms and don't yet require hospitalisation, as well as various scenarios which might allow for the safe reopening of schools in the settlement. | medium |
| B.C. UB; Pokharel S; Munikar S et al. | 2021 | Anxiety and depression among people living in quarantine centers during COVID-19 pandemic: A mixed method study from western Nepal | Nepal | LMIC | April 21, 2020 to May 15, 2020 | Internal and International migrants | mental health | Mixed-Methods: Cross-Sectional Study; Qualitative Research | Mild depression (9.1%; 40/441) and anxiety (16.1%; 71/441) was common among respondents followed by moderate depress-sion and anxiety {depression (3.4%; 15/ 441), anxiety (4.1%; 18/441)} and severe depression and anxiety {depression (1.1%; 5/441), anxiety (0.7%; 3/441)}. Anxiety and depression were independent of their socio-demographic characteristics. Perceived fear of contracting COVID-19, severity and death were prominent among the respondents. Respondents experienced stigma and discrimination in addition to being at the risk of disease and possible loss of employment and financial responsibilities. In addition, poor (quality and access to) health services, and poor living con-dition at the quarantine centres adversely affected respondents' mental health. | Depression and anxiety were high among quarantined population and warrants more research. Institutional quarantine centers of Karnali province of Nepal were in poor conditions which adversely impacted mental health of the respondents. Poor resource allocation for health, hygiene and living conditions can be counter-productive to the population quarantined. | high |
| Babuc ZT. | 2021 | A Relational Sociological Analysis on the Impact of COVID-19 Pandemic Lockdown on Syrian Migrants' | Turkey | UMIC | July 2020 | Refugees | effect of lockdown measures | Qualitative Research | According to the findings of the study, the suspension or slowdown of economic activities due to the pandemic has caused not only an uncertainty about the future, but also a change in familial positions and roles. Also the accepted norms of social relations have been inevitably suspended | Considering the fact that the lockdown measures can be tightened again at any time with respect to the increase in the spread of the disease, social and economic policies should be expanded to include the Syrian migrants. Also educational policies during the COVID-19 pandemic should include | medium |

| | | | | | | | | | | | |
|---|---|---|---|---|---|---|---|---|---|---|---|
| | | Lives in Turkey: The Case of Mersin Province | | | | | | | resulting in increased ambiguity. Strengthening the network of solidarity within relatives and building stronger relationships within the family are common relational strategies of the participants to mitigate the social and economic impacts of the COVID-19 outbreak (…). | specific measures such as technological support and guidance to mitigate the risks and disadvantages that Syrian children experience in terms of distance learning. | |
| Bauza V; Sclar G, Bisoyi A, Owens A, Ghugey A, Clasen T. | 2021 | Experience of the COVID-19 pandemic in rural Odisha, India: knowledge, preventative actions, and impacts on daily life | India | LMIC | May 2020 to July 2020 | Retournees | effect of lockdown measures | Mixed-Methods: Cross-Sectional Study; Qualitative Research | Respondents reported high compliance with important preventative measures, including staying home as much as possible (94%), social distancing (91%), washing hands frequently (96%), and wearing a facial mask (95%). Additionally, many respondents reported job loss (31%), financial challenges (93%), challenges related to staying home whether as a preventative measure or due to lockdowns (57%), changes in types and/or amount of food consumed (61%), and adverse emotional effects as a result of the pandemic and lockdown. (…) Although the research revealed high compliance with preventative measures, the pandemic and associated lockdowns also led to many challenges and hardships faced in daily life particularly around job loss, economic security, food security, and emotional wellbeing. | The results underscore the vulnerability of marginalized populations to the pandemic and the need for measures that increase resilience to large-scale shocks. (…) In particular, there is a need for greater government response to limit harm to livelihood and mental, social, and nutritional health during any lockdown periods required to reduce disease spread. | high |
| Biddle L; Jahn R; Perplies C et al. | 2021 | [COVID-19 in collective accommodation centres for refugees: assessment of pandemic control measures and priorities from the perspective of authorities] | Germany | HIC | May 1, 2020 to July 31, 2020 | Asylum Seekers and Refugees | mental health | Qualitative Research | We found substantial heterogeneity of measures taken to prevent infection, inform refugees, maintain social and health services, test for SARS-CoV-2 and quarantine positive cases. Effective inter-sectoral cooperation proved to be particularly important for coordination and implementation of measures. Need for support was expressed with regard to the improvement of infrastructure, opportunities to work with language interpreters and stronger involvement of local health experts. | Amidst multiple actors and the complexity of structures and processes, the admission authorities have been taking on essential responsibilities related to infection control on an ad hoc basis, without being sufficiently positioned to do so. In order to further contain the pandemic, a strengthening of centralised, setting-specific recommendations and information as well as their translation through the pro-active involvement of the public health authorities at the local level are essential. | medium |
| Bojorquez I; Infante C; Vieitez I; Larrea S; Santoro C. | 2020 | MIGRANTS IN TRANSIT AND ASYLUM SEEKERS IN MEXICO: AN | Mexico | UMIC | January 1, 2020 to May 3, 2020 | Asylum seekers | health outcomes of disease | Ecological Study | Migrants were on average younger, had less accompanying risk conditions, and a lower percentage of suspect cases tested positive for COVID-19. The odds of hospitalization were lower among | Migrants and asylum seekers in Mexico are at increased risk for infectious respiratory diseases, and could be disproportionately affected by COVID-19. It is important to continue monitoring the situation, with more | high |

| | | | | | | | | | | | |
|---|---|---|---|---|---|---|---|---|---|---|---|
| | | EPIDEMIOLOGICAL ANALYSIS OF THE COVID-19 PANDEMIC | | | | | | | migrants, but the difference disappeared after adjusting by age, gender and underlying risk conditions. The cumulative incidence ratios comparing migrants with nonmigrants were 6.12 (CI95% 4.75,7.77) for the first scenario, and 1.49 (CI95% 1.15,1.89) for the second scenario. | detailed information about migration status, living conditions and other determinants of migrants health. | |
| Bojorquez I; Odgers-Oritz O; Olivas-Hernandez OL. | 2021 | Psychosocial and mental health during the COVID-19 lockdown: A rapid qualitative study in migrant shelters at the Mexico-United States border | Mexico | UMIC | April 2020 to May 2020 | International migrants | mental health; health services access | Qualitative Research | In addition to fear of contagion and economic insecurity, migrants experienced emotional distress associated with hardening migration policies, and the difficulties of having to find shelter in place in non-private spaces. Some CSOs continued or adapted previous psychosocial support activities, helping migrants navigate these issues, but other activities stopped amidst physical distancing measures and because of limited resources. Migrants themselves implemented some group activities. There was a surge of civil society initiatives of online support, but some shelters leaked the techno-logical and other resources to benefit from them. | To conclude, our results suggest that, as other authors anticipated, the COVID-19 pandemic and lockdown measures have important mental health consequences for migrants. (…) As recommended by inter-national organizations, instead of being limited to clinical interventions, this response should prioritize the psychosocial aspects that are the root of mental health problems in these types of populations, covering basic needs such as decent work, appropriate places to live, and access to education and health services. | high |
| Bolivar C. | 2020 | In limbo: Survey of impact of COVID-19 on Venezuelan migrants in Trinidad and Tobago | Trinidad and Tobago | HIC | May 10, 2020 to June 10, 2020 | Refugees | effect of lockdown measures | Cross-Sectional Study | 92% felt their livelihoods disrupted; 50% lost all income, and a further 36% lost between half and three quarters. Almost 80% resorted to borrowing money to avoid eviction and meet basic needs; 83 % either skipped meals or reduced choice. The majority of respondents indicated that individuals or independent church organizations were their main source of help. | Assistance from organizations whose raison d'eË†tre is to protect refugees and migrants in Trinidad and Tobago was classed as insufficient by 39 per cent, and seriously insufficient by 33 per cent | low |
| Bonizzoni P.; Dotsey S. | 2021 | Migration and legal precarity in the time of pandemic: Qualitative research on the italian case | Italy | HIC | March 2020 to June 2020 | International migrants | effect of lockdown measures | Qualitative Research | The study showed that legal precariousness is deeply rooted in – and constantly reproduced by – the Italian immigration system, treating migrants as a temporary phenomenon and making long-term and secure status especially difficult to achieve and maintain. For those unable to renew their status, the pandemic has triggered a prolonged suspension of one’s legal status, with significant spill-over effects on several aspects of one’s personal and familial life. | Accessing rights in the time of pandemic is jeopardized by the lack of reliable information. The messages issued during the pandemic were especially difficult to decode for those who had limited knowledge of the national bureaucratic language and the local institutional landscape. The virtualization of several services made those less equipped with technological skills somehow excluded, also considering that all major intermediary services were offering support only by | low |

| | | | | | | | | | | | |
|---|---|---|---|---|---|---|---|---|---|---|---|
| | | | | | | | | | | phone or had their services severely curtailed | |
| Boogaard L; Sikkema R, vanBeek J. et al. | 2021 | A mixed-methods approach to elucidate SARS-CoV-2 transmission routes and clustering in outbreaks in native workers and labour migrants in the fruit and vegetable packaging industry in South Holland, the Netherlands, May to July 2020 | Netherlands | HIC | June 5, 2020 to August 13, 2020 | Migrant workers | risk of infection | Mixed-Methods | We detected 46 SARS-CoV-2 cases and 4 outbreaks with a proportional representation of labour migrant and native workers in 6 unrelated facilities. Complete viral genome sequences revealed at least 3 clusters of native workers and labour migrants, 2 within and 1 between facilities. On-site inspections found adequate implementation of preventative measures to which both native workers and labour migrants showed suboptimal adherence. Being a labour migrant was associated with living in shared housing, but not with more contacts or different sources. | The fruit and vegetable packaging industry gave the impression of sufficient preparedness and control. Suboptimal adherence to the facilities' preventative guidelines could have facilitated work floor transmission. Community and household transmission are likely to have contributed to outbreaks. We encourage further research into risk factors for transmission in labour migrants and application of these insights into targeted public health policy. | low |
| Brinkmann F; Diebner HH; Matenar C et al. | 2021 | Seroconversion rate and socioeconomic and ethnic risk factors for SARS-CoV-2 infection in children in a population-based cohort | Germany | HIC | June 2020 to February 2021 | International migrants | health outcomes of disease: SARS-CoV-2 cases (risk of infection) | Cohort Study | (…) the incidence of SARS-CoV-2 infections is significantly higher in families of Turkish (OR 6.24 [1.38-28.13], p=0.017) or Middle Eastern (OR 6.44 [1.14- 36.45], p=0.035) decent independent of other risk factors i.e. after adjustment for other socioeconomic confounders. | This study shows that poor socio-economic status and Turkish or Middle Eastern migration background are independent risk factors for SARS-CoV-2 infection in a population-based cohort of children and adolescents in Germany. | medium |
| Bruni V. | 2021 | Kazakhstan: Study on the Socioeconomic Effects of COVID-19 on Stranded Migrants | Kazakhstan | UMIC | December 3, 2020 to January 5, 2021 | International migrants | effect of lockdown measures | Cross-Sectional Study | The research study shows that the most common types of challenges that were faced by stranded migrants include: unemployment, insufficient income and the inability to leave Kazakhstan. (...) The data demonstrates that insufficient income (21%) and inability to leave Kazakhstan (21%) are two key challenges that were indicated by a large proportion of respondents. | not reported | medium |
| Bruni V. | 2021 | Regional Overview: Survey on the Socioeconomic Effects of COVID-19 on Returnees and Stranded Migrants in Central Asia and the Russian Federation | Russian Federation, Kazakhstan, Kyrgyzstan and Tajikistan | MICs | December 2020 | International migrants and Retournees | effect of lockdown measures | Cross-Sectional Study | In total, 8 out of 10 participants reported having debts, lost part of their income, or all of their income. The inability to migrate again and the lack of remittances from abroad add pressure to this already precautious context. (...) Unemployment doubled as compared to the period before migration, and even respondents who were working at the time of the assessment reported earning less as compared to the period before the COVID-19 outbreak. | The deterioration of the financial situation resulted in partial or total income losses, debts, and inability to meet basic needs. The majority of the sample population faced such problems only after the COVID-19 outbreak, indicating that the current pandemic is simultaneously exacerbating older problems and causing new ones. | medium |

| | | | | | | | | | | | |
|---|---|---|---|---|---|---|---|---|---|---|---|
| Buda S; an der Heiden M; Altmann D; Diercke M; Hamouda O; Rexroth U. | 2020 | Infection environment from recorded COVID-19 outbreaks in Germany | Germany | HIC | up until August 11, 2020 | International migrants | health outcomes of disease: cases, hospitalisation | Cross-Sectional Study | Outbreaks around the workplace were often associated with difficult working conditions and, in some cases, cramped living conditions. This applies in a similar way to outbreaks in refugee and asylum seeker shelters and residential homes. (translated) | However, important findings can also already be summarized in the descriptive evaluation of the information on the infection environment in outbreaks in the reporting system: As described earlier, it is confirmed that intensive and prolonged personal contact is associated with a high risk of transmission. (translated) | medium |
| Cai R; Novosad P; Tandel V; Asher S; Malani A. | 2021 | Representative Estimates of COVID-19 Infection Fatality Rates from Three Locations in India | India | LMIC | March 2020 to June 2021 | Retournees | health outcomes of disease | Cross-Sectional Study | Among males aged 50-89, measured IFR was 0.12% in Karnataka (95% CI 0.09% to 0.15%), 0.42% in Tamil Nadu (95% CI 0.39% to 0.45%), 0.53% in Mumbai (95% CI 0.52% to 0.54%) and an imprecise 5.64% (95% CI 0% to 11.16%) among migrants returning to Bihar. Estimated IFR was approximately twice as high for males as for females, heterogeneous across contexts and rose less dramatically at older ages compared with similar studies in high-income countries. | Our findings suggest that the standard estimation approach may substantially underestimate IFR in low-income settings due to underreporting of COVID-19 deaths, and that COVID-19 IFRs may be similar in low-income and high-income settings. | high |
| Calderón-Larrañaga A; Vetrano DL; Rizzuto D; Bellander T; Fratiglioni L; Dekhtyar S. | 2020 | High excess mortality in areas with young and socially vulnerable populations during the COVID-19 outbreak in Stockholm Region, Sweden | Sweden | HIC | January 1, 2015 to May 31, 2020 | International migrants (%) | health outcomes of disease: mortality | Ecological study | During the outbreak, the highest excess mortality was recorded among DeSOs in the lowest tertiles for income, educational level, share of Swedish-born and share of gainfully employed (excess of 171%, 162%, 178% and 174%, respectively, for the peak week of 6–12 April 2020; figure 2). (...) The corresponding numbers for the peak mortality week (ie, 6–12 April 2020) were (...) 198% for younger DeSOs with the lowest share of Swedish-born. | Future COVID-19–related strategies of social distancing, confinement, active surveillance, and eventually vaccination, will necessarily have to consider individuals' risk of morbidity and mortality if they were to contract the virus. This necessitates to also account for their sociodemographic context. In the Stockholm Region, vulnerable individuals living in deprived areas with a high proportion of young people will require carefully tailored prevention strategies. | medium |
| Canevelli M.; Palmieri L.; Raparelli V. et al. | 2020 | COVID-19 mortality among migrants living in Italy | Italy | HIC | February 21, 2020 to April 29, 2020 | International migrants | health outcomes of disease: mortality | Cross-Sectional Study | The proportions of natives and migrants among the COVID-19-related deaths (95.7% and 2.5%, respectively) were similar to the relative all-cause mortality rates estimated in Italy in 2018 (97.4% and 2.6% respectively). Migrants were younger at the time of death compared to natives (71.1, SD 13.1 years vs 78.3, SD 10.8 years; $p < 0.001$). | The clinical phenotype of migrants dying with COVID-19 was similar to that of natives except for the younger age at death. International migrants living in Italy do not have a mortality advantage for COVID-19 and are exposed to the risk of poor outcomes as their native counterparts. | medium |
| Casanova F; Hamblett A; Brinkley L; | 2022 | Epidemiology of Coronavirus Disease 2019 in US Immigration and Customs | USA | HIC | May 5, 2020 to September 15, 2020 | Undocumented migrants | health outcomes of disease: SARS-CoV-2 cases | Cohort Study | (…) the overall September 15 cumulative case rate was 214 per 1000 people (5810 cases among 27 189 people); ICE reported 6 deaths. | Cumulative case rates among people detained by ICE are higher than those of the US population and dwarf those of surrounding communities. | medium |

| | | | | | | | | | | | |
|---|---|---|---|---|---|---|---|---|---|---|---|
| Nowotny K. | | Enforcement Detention Facilities | | | | | | | | | |
| Chander R; Murugesan M; Ritish D et al. | 2021 | Addressing the mental health concerns of migrant workers during the COVID-19 pandemic: An experiential account | Karnataka, India | LMIC | not reported | Internal migrants | effect of lockdown measures; health services access | Qualitative research | Main concerns were not Covid19-related risks; but uncertainty about duration of lockdown, eagerness to meet family, fear of job loss, insecurity of income, fears related to lack of access to services for children and pregnant women, substance use-related concerns | (...) forming an integral system to address the overall well-being of this population will drastically reduce the 'knee-jerk' measures that need to be taken during crisis like COVID-19 in the future. (...) a couple of measures like provision of unique national registration and identity, particularly for the purpose of obtaining provisions, and political inclusions, including housing, finance, legal aid and public health, should be laid for ensuring their well-being. | low |
| Chen Y; Wang Z; Dong W et al. | 2021 | Perceived discrimination and mental health among the Chinese diaspora during COVID-19 | 35 countries | HIC | April 2020 to May 2020 | International migrants | mental health | Cross-Sectional Study | Participants in the "high" discrimination group were approximately 5 times more likely to have anxiety disorder (OR, 4.60; 95 % CI, 2.44-8.66; P<0.001) and depression (OR, 4.99; 95 % CI, 2.61-9.53; P<0.001), compared with participants in the "none" discrimination group. | The findings provides important data in support of immediate and long-term interventions to combat anti-Asian (and particularly anti-Chinese) racism, and to provide timely mental health care during and after the COVID 19 pandemic, which also constitutes an important part of public health measures in the COVID-19 pandemic response. | high |
| Chew M; Koh F; Wu J et al. | 2020 | Clinical assessment of COVID-19 outbreak among migrant workers residing in a large dormitory in Singapore | Singapore | HIC | April 11, 2020 to April 19, 2020 | Migrant workers | health outcomes of disease | Cross-Sectional Study | 1264 (69%) were found to be positive for COVID-19. COVID-19-positive and -negative groups had similar median age (33 years,P: 0.71) and heart rate (108 vs 107 bpm, P: 0.17). COVID-19-positive foreign workers had slightly higher median temperatures (37.8 vs 37.7,P<0.01) | The strategy for containment in closed-living environments therefore would be to isolate symptomatic individuals, and to establish public health measures for social distancing. | low |
| Coyer L; Wynberg E; Buster M et al. | 2021 | Hospitalisation rates differed by city district and ethnicity during the first wave of COVID-19 in Amsterdam, The Netherlands | The Netherlands | HIC | February 29, 2020 and May 31, 2020 | International migrants | health outcomes of disease: hospitalisation | Cross-Sectional Study | Hospitalisation DSR were more than two-fold higher in individuals with a non-Western migration background compared to ethnic-Dutch individuals (RR= 2.36, 95%CI = 2.17-2.54), with 57 additional hospitalisations per 100.000 population (RD = 57.05, 95%CI = 43.34-70.75). | City districts with lower economic and health status and those with a non-Western migration background had the highest burden of COVID-19 during the first wave of COVID-19 in Amsterdam. | high |
| Cromer SJ; Lakhani CM; Wexler DJ; Burnett-Bowie S-AM; Udler M; Patel CJ. | 2020 | Geospatial Analysis of Individual and Community-Level Socioeconomic Factors Impacting SARS-CoV-2 Prevalence and Outcomes | USA | HIC | February 1, 2020 to June 21, 2020 | International migrants | health outcomes of disease | Cross-Sectional Study | Among those infected, individual age, sex, race, language, and insurance, and census tract-level measures of lower education, more multi-family homes, and extreme household crowding were associated with increased likelihood of hospitalization, while higher per capita testing rates were associated with decreased likelihood. Only individual-level variables (older age, male sex, Medicare insurance) were associated | This study of the first wave of the SARS-CoV-2 pandemic in a major U.S. city presents the cascade of outcomes following SARS-CoV-2 infection within a large, multi-ethnic cohort. SARS-CoV-2 infection and hospitalization rates, but not death rates among those hospitalized, are related to census tract-level socioeconomic characteristics including lower educational attainment and higher household crowding | high |

| | | | | | | | | | | | |
|---|---|---|---|---|---|---|---|---|---|---|---|
| | | | | | | | | | with increased mortality among those hospitalized. | and occupancy, but not neighborhood measures of race, independent of individual factors. | |
| Cummings KJ; Beckman J; Frederick M. et al. | 2022 | Disparities in COVID-19 Fatalities among Working Californians | USA | HIC | January 1, 2020 to December 31, 2020 | International migrants | health outcomes of disease | Cross-Sectional Study | COVID-19 accounted for 8,050 (9.9%) of 81,468 fatalities among Californians 18–64 years old. Of these decedents, 2,486 (30.9%) were matched to state employment records and classified as "confirmed working". (...) Confirmed and likely working COVID-19 decedents were predominantly male (76.3%), Latino (68.7%), and foreign-born (59.6%), with high school or less education (67.9%); 7.8% were Black. The overall age-adjusted COVID-19 mortality rate was 30.0 per 100,000 workers (95% confidence interval [CI], 29.3–30.8). | Californians in manual labor and in-person service occupations experienced disproportionate COVID-19 mortality, with the highest rates observed among male, Latino, and Black workers; these occupational group should be prioritized for prevention. | high |
| Da Mosto D; Bodini C; Mammana L; Gherardi G; Quargnolo M; Fantini MP. | 2021 | Health equity during COVID-19: A qualitative study on the consequences of the syndemic on refugees' and asylum seekers' health in reception centres in Bologna (Italy) | Italy | HIC | April 2020 to September 2020 | Asylum Seekers and Refugees | mental health | Qualitative research | Even though various measures were implemented in reception centres (i.e. mass quarantine, supply of personal protective equipment, risk communication campaigns and specific governance tools) they often had a discriminatory approach towards migrants and only considered the biomedical aspects of COVID-19, excluding its social roots and repercussions. This factor, together with the lack of an effective governance system at both the local and the national level, was the most relevant issue associated with the management of the syndemic in reception facilities and affected all the social determinants that shape the health profile of RAS. | The study revealed the importance of social factors in the management of the syndemic in reception centres. It also highlighted how the underlying causes of the impact of COVID-19 are tightly correlated to the political and social approaches of local and national institutions to migration. In order to guarantee the well-being of society as a whole and successfully control the epidemic, it is necessary to consider migration as a human reality rather than an emergency, and demolish all the policies and bureaucratic systems that act as structural violence on RAS. (…) This way it will be possible to develop more inclusive approaches to public health and guarantee the conditions for RAS' empowerment. | medium |
| Dale R; O'Rourke T; Humer E; Jesser A; Plener P; Pieh C. | 2021 | Mental Health of Apprentices during the COVID-19 Pandemic in Austria and the Effect of Gender, Migration Background, and Work Situation | Austria | HIC | March 9, 2021 to May 18, 2021 | International migrants | mental health | Cross-Sectional Study | There was a high prevalence of clinically relevant depression (cut-offs ≥11 for adolescents, ≥10 for adults: 48.3%), anxiety (cut-offs ≥11 for adolescents, ≥10 for adults: 35.4%), insomnia (cut-off ≥15: 27%), and disordered eating (cut-offs ≥2 for men, ≥3 for women: 50.6%). Linear models revealed that apprentices with female and diverse gender, migration background, and unemployed status showed the poorest scores on all mental health measures (all p-values < 0.05) except disordered eating. | These findings emphasize the need for intersectional strategies to reduce and prevent adverse mental health consequences of the COVID-19 pandemic for apprentices. | medium |

| | | | | | | | | | | | |
|---|---|---|---|---|---|---|---|---|---|---|---|
| D'Ambrosi F; Iurlaro E; Tassis B et al. | 2021 | Sociodemographic characteristics of pregnant women tested positive for COVID-19 admitted to a referral center in Northern Italy during lockdown period | Italy | HIC | March 1, 2020 to April 30, 2020 | International migrants | health outcomes of disease: SARS-CoV-2 cases (risk of infection) | Cross-Sectional Study | (…) foreign pregnant women (crude OR = 3.32; 95% CI:1.89-5.81) showed a significantly higher risk of infection. Ethnicity was still significantly associated with an increased risk of COVID-19 (mutually adjusted OR = 2.15; 95% CI: 1.12-4.11) when taking into account the effect of other sociodemographic characteristics. | In conclusion, the socioeconomic conditions we described are characteristic of contemporary immigration patterns. These factors altogether increase the risk of viral transmission by droplets and contact within households, ultimately reducing the effectiveness of lockdown and social distancing. | high |
| Das T; Basu Roy T; Roy R. | 2021 | Reintegration with family and intimate partner violence (IPV) against women among the returnee migrant worker's family during COVID-19 induced lockdown: A Block-level analysis using multinomial logistic regression model | India | LMIC | September 6, 2020 to October 3, 2020 | Retournees | effect of lockdown measures | Cross-Sectional Study | The result of the multinomial logistic regression model revealed that, after controlling for other variables, the women who had the poorest wealth background were 37% more likely (RRR: 1.37; 95% CI [1.18, 1.47]) to experience IPV almost every day in a week than those who had a middle wealth background. (...) Furthermore, partner's loan having history in this COVID-19 crisis with weekly or monthly instalment patterns were also found to be a strong predictor for experiencing IPV inside the home. These study results also found that the likelihood of experiencing IPV almost daily is higher among those women whose partners consume alcohol daily. | In the COVID-19–induced lockdown, most of the government, private, and public services were disrupted; therefore, it would be very challenging for women to confess and seek any relief. It is our appeal to the government to organise virtual, media-based, or community-based programmes such as communication and counselling, to teach young people how to create and maintain healthy relationships. | high |
| Daw M; El-Bouzedi A; Ahmed M. | 2021 | COVID-19 and African Immigrants in North Africa: A Hidden Pandemic in a Vulnerable Setting | Lybia | UMIC | not reported | International migrants | risk of infection | Cross-Sectional Study | 11 (3.1%) tested positive for SARS-CoV-2, ranging from 0.6%, among a younger age group (20-30 years old) and reaching up to 6.0% in a middle age group. | In conclusion, the disproportionate burden of the SARS-CoV-2/COVID-19 pandemic for African immigrants is distressing, and thus, immediate action by the international community and regional governments is urgently needed to implement proper public health measures to protect immigrants as residents and when they return to their countries. | medium |
| de Diego R; Tarrino L; Lato M; Garcia M. | 2021 | COVID-19 and female immigrant caregivers in Spain: Cohabiting during lockdown | Spain | HIC | April 2019 to September 2020 | Migrant workers | mental health | Qualitative research | The results show the moral debt accrued by the caregivers with the family who employ them, while worsening the physical and psychological health of many of the care-givers, due to both work overload and fear of the global pandemic. | The emotional debt and the relationship as 'quasi-family members' have contributed to increasing the work overload, since as well as the usual care, they have been responsible for all the domestic work, resulting in a non-stop working day during the lockdown, without any personal free time and being separated from the social group of their friends who live in Spain. | high |
| Debopadhaya S; Erickson | 2021 | Temporal analysis of social determinants | USA | HIC | July 5, 2020 to Decembe | International | health outcomes of disesase | Ecological Study | The results indicate that counties with certain ethnic minorities and age groups, immigrants, prevalence of diseases like | Additionally, the temporal trends observed in this study suggest that the policies implemented during this pandemic may have | high |

| | | | | | | | | | | | |
|---|---|---|---|---|---|---|---|---|---|---|---|
| JS; Bennett KP. | | associated with COVID-19 mortality | | | r 28, 2020 | migrants (%) | | | pediatric asthma and diabetes and cardiovascular disease, socioeconomic inequalities, and higher social association are associated with increased COVID-19 mortality rates. | been effective, as the impact of several socioeconomic metrics (overcrowded housing, immigrant populations, poor health in a county) appear to significantly decrease over time. | |
| Debopadhaya S; Sprague AD; Mou H et al. | 2020 | Social Determinants Associated with COVID-19 Mortality in the United States | USA | HIC | July 5, 2020 | International migrants (%) | health outcomes of disease | Ecological study | The results describe that ethnic minorities, immigrants, socioeconomic inequalities, and early exposure to COVID-19 are associated with increased COVID-19 mortality, while the prevalence of asthma, suicide, and excessive drinking is associated with decreased mortality. | Overall, we recognize that social inequality places disadvantaged groups at risk, which must be addressed through future policies and programs. Additionally, we reveal possible relationships between lung disease, mental health, and COVID-19 that need to be explored on a clinical level. | medium |
| DeCarlo Santiago C; Bustos Y; Jolie SA et al. | 2021 | The impact of COVID-19 on immigrant and refugee families: Qualitative perspectives from newcomer students and parents | USA | HIC | June 2020 to July 2020 | International migrants (including international students) | effect of lockdown measures | Qualitative research | Qualitative analyses revealed three major categories of themes. Challenges include difficulty adjusting; fear, worry, and uncertainty; financial concerns; academic challenges; lack of connection. Sources of Support include school and community supports. Coping and Adaptation includes the ways families have coped with COVID-19. | Despite limitations, the present study highlights the numerous difficulties that newcomer families are experiencing during COVID-19. Although increased distress is common (…), newcomer families may experience difficulties that are compounded by the resettlement experience or barriers based on status. In particular, schools must attend to digital literacy, accessible information (…), regular connection to teachers and school staff, in addition to challenges associated with basic necessities, health, and social-emotional distress. Professional development training focused on newcomer experiences could ensure all school staff are equipped with the knowledge and tools necessary to support newcomer youth. | medium |
| Devillanova C; Colombo C; Garofolo P; Spada A. | 2020 | Health care for undocumented immigrants during the early phase of the Covid-19 pandemic in Lombardy, Italy | Italy | HIC | January 2, 2020 to April 23, 2020 | Undocumented migrants | health services access | Cross-Sectional Study | We document a sharp reduction in visit number after lockdown, with 16% frequency of acute respiratory infections, compatible with COVID-19. Moreover, housing conditions make it difficult to implement public health measures. | We advocate the need to foster primary care services by all the people, undocumented migrants included, as an essential measure to guarantee the universal right to health and to face emergency and challenging conditions such as those caused by the COVID-19. | medium |
| Diaz-Menendez M; Trigo E; Borobia AM et al. | 2021 | Presenting characteristics and outcomes of migrants in a cohort of hospitalized patients with COVID-19: Does the origin matter? | Spain | HIC | February 25, 2020 to April 19, 2020 | International migrants (%) | health outcomes of disesase | Cohort Study | (…) being a migrant decreased the probability of dying (OR: 0.22; 95% CI 0.15-0.33; P <0.001) after adjusting for sex, age and comorbidities. | (…) although the high rates of infection and consulting later than Spanish natives, being a younger and healthier population may have conditioned a more favorable outcome of the disease in this group. | medium |

| | | | | | | | | | | | |
|---|---|---|---|---|---|---|---|---|---|---|---|
| Doblhammer G; Kreft D; Reinke C. | 2021 | Regional Characteristics of the Second Wave of SARS-CoV-2 Infections and COVID-19 Deaths in Germany | Germany | HIC | October, 2020 to December 2020 | International migrants (%) | health outcomes of disesase | Ecological and Modelling Study | Features related to international migration background were among the top characteristics through mid-to-late November and correlated positively with incidence and death rates. In the first period, the regional "%Foreigners in the total population in 2017" (...) ranked third in incidence and tenth in mortality. In the second period, it still ranked fourth in incidence and eleventh in mortality, and in the third period, it ranked sixth in mortality. | Our study showed that a number of regional characteristics were crucial for the increase in infections in the second wave. As in the first wave, they moved from high to low SES regions. Risky working conditions with reduced opportunities for social distancing, a high burden of chronic disease, and residence in nursing homes may underlie this concentration in low-SES regions. | medium |
| Donati S; Corsi E; Maraschini A; Salvatore MA. | 2021 | SARS-CoV-2 infection among hospitalised pregnant women and impact of different viral strains on COVID-19 severity in Italy: a national prospective population-based cohort study | Italy | HIC | February 1, 2021 to June 30, 2021 | International migrants (%) | health oucomes of disease | Cohort Study | 64.3% of the cohort was asymptomatic, 12.8% developed COVID-19 pneumonia and 3.3% required ventilatory support and/or ICU admission. | Our results are consistent with a low risk of severe COVID-19 disease among pregnant women and with rare adverse perinatal outcomes. During the Alpha-variant period there was a significant increase of severe COVID-19 illness. Further research is needed to describe the impact of different SARS-CoV-2 viral strains on maternal and perinatal outcomes. | high |
| Drefahl S; Wallace M; Mussino E et al. | 2020 | A population-based cohort study of socio-demographic risk factors for COVID-19 deaths in Sweden | Sweden | HIC | March 13, 2020 to May 7, 2020 | International migrants | health outcomes of disease: mortality | Cohort Study | Immigrants from low- and middle-income countries from the Middle East and Northern Africa displayed more than three times higher mortality among men (HR: 3.13; 95% CI: 2.51, 3.90) and two times higher among women (HR: 2.09; 95% CI: 1.52, 2.89), as compared to those born in Sweden, (…) whereas immigrants from other low- and middle-income countries experienced more than twice as high mortality among men (HR: 2.20; 95% CI: 1.81, 2.69) and ~1.5 times higher mortality among women (HR: 1.45; 95% CI: 1.12, 1.90). Immigrants from HIC only displayed 19% higher mortality among men (HR: 1.19; 95% CI: 1.01, 1.39) and 8% higher mortality among women (HR: 1.08; 95% CI: 0.92, 1.26). | However, the robust finding of elevated mortality among immigrants from low- and middle-income countries deviates from findings on other causes of death, such as neoplasms or circulatory diseases, where immigrants tend to have lower mortality than natives. This finding may help guide governmental policy, and may be of relevance for other HIC as well. Better health care resources may need to be allocated toward communities with many foreign-born individuals. | medium |
| Dressler A; Finci I; Wagner-Wiening C, Echner M; | 2021 | Epidemiological analysis of 3,219 COVID-19 outbreaks in the state of Baden- | Germany | HIC | April 27, 2020 to December 6, 2020 | Asylum Seekers and Refugees (%) | health outcomes of disease: hospitalisation (risk of | Ecological Study | 53 (1.6) outbreaks in asylum accom-modations with 603 (2.7) cases with a median cases of 8 (4-14). (…) median age was 26 (17-34 IQR), 13 (0.9) cases were | Our analysis highlights the necessity of targeted, setting-specific approaches to control transmission in this vulnerable population. | high |

| | | | | | | | | | | | |
|---|---|---|---|---|---|---|---|---|---|---|---|
| Brockmann SO. | | Wuerttemberg, Germany | | | | | infection/transmission) | | hospitalized and 1 (0.1) cases were deceased. | | |
| Duggal A.; Ray A.; Konantambigi R.; Kothari A. | 2021 | The nowhere people: Lived experiences of migrant workers during Covid-19 in India | India | LMIC | April 2020 to June 2020 | Internal migrants (%) | effect of lockdown measures | Qualitative research | Participants reported that they were afraid of dying alone in the city without their families around so they decided to go back home. At the same time the stress of being back home without any way of financially supporting themselves and their families generated great amounts of uncertainty and helplessness in the participants. They experienced feelings of guilt and frustration at not being able to find a job and worried constantly about the future. Participants reported experiencing worries about the future, helplessness and a fear that the world and 'normal life' was about to change indefinitely. Their sense of personal agency and certainty was taken away almost overnight which generated distress in them. | The research used an intersectional frame to understand how different social identities of migrant status, gender and class coalesce together to generate unique forms of marginalization. It calls for future policy and practice work to acknowledge and integrate the lived experiences of those who remain on the edges, invisible yet critical backbones of a well-functioning and harmonious society. Finally, it emphasises that the pandemic is just a mirror being held up to all of us- and the 'reflections' and 'lessons' need to be carried forward, remembered, documented and transformed into action that is meaningful to the lives of people. | high |
| Ebrahim SH; Ali A; Koya SF et al. | 2021 | Focus on international and domestic travellers are equally important for successful SARS-COV-2 mitigation: ecological comparison of emigrant and migrant travel patterns and COVID-19 trends in Kerala State, India | India | LMIC | January 2020 to September 30, 2020 | International migrants (%) | risk of transmission | Cross-Sectional Study | During this period [May 7 to June 30, 2020] domestic migrant returnees as a percentage of total COVID-19 cases reached 36% (compared with 9% in earlier phase) with slightly increased average overall seropositivity rate (2.3%). (...) By August, all the districts (the lowest administrative regions) reported COVID-19 cases, suggesting that the reach of SARS-COV-2 is extensive and no rural areas are free from the virus. Daily COVID-19 cases among returning domestic migrants from other states increased and peaked during July 2020. | The natural history of COVID-19 in Kerala reaffirms that synchronized multiple layers of pandemic mitigation efforts at the onset of an outbreak can help achieve pandemic mitigation. Kerala data also indicate that premature lifting of critical components of mitigation efforts diminishes the impact of mitigation. Further, during an outbreak when extent and patterns of transmission are unclear focus exclusively on a single transmission route (international travellers in this case) may not achieve optimum outcome. | low |
| Efani P; Uppal N; Lee C; Mishori R; Peeler K. | 2021 | COVID-19 Testing and Cases in Immigration Detention Centers, April-August 2020 | USA | HIC | April 1, 2020 to August 31, 2020 | ICE detainees | health outcomes of disesase: SARS-CoV-2 cases | Cohort Study | On August 31, ICE reported 5379 cumulative COVID-19 cases and 6 related deaths among its detainees. Cases were reported in 92 of 135 facilities, with 20 facilities accounting for 71% of cases. From April to August 2020, the mean monthly case rate ratio for detainees, compared with the US population, was 13.4 (95% CI, 8.0-18.9), ranging from 5.7 to 21.8 per month. The mean monthly test rate ratio for detainees, compared with the US population, was 4.6 (95% CI, 2.5-6.7), ranging from 2.0 to 6.9 per month. | Despite ICE's mitigation efforts, COVID-19 case rates among detainees increased every month from April to August. (...) COVID-19 spread within facilities may be partially due to challenges faced implementing the Pandemic Response Requirements. An independent assessment of facilities' mitigation strategies is necessary to identify and address existing gaps in these efforts. | medium |

| | | | | | | | | | | | |
|---|---|---|---|---|---|---|---|---|---|---|---|
| Elci E; Kiriscioglu E; Uestuebici A. | 2021 | How Covid-19 financially hit urban refugees: evidence from mixed-method research with citizens and Syrian refugees in Turkey | Turkey | UMIC | April 5, 2020 to August 2020 | Refugees (%) | effect of lockdown measures | Mixed-Methods: Cross-Sectional Study; Qualitative research | Our findings indicate that the initial Covid-19 measures had a higher financial impact on Syrians than on citizens when controlled for employment, wealth, and education, among other variables. In line with the literature, our research confirms that disasters' socio-economic effects disproportionally burden minority communities. | Our findings reveal that being a refugee in Turkey accelerates and magnifies the pandemic's perceived economic impact, even after controlling for social class, demographic factors, and geographical location. (…) While signalling the unequal outcomes of the Covid-19 pandemic, our research also addresses the underlying causes of these outcomes, as well as the implications of the pandemic for the future. As the Syrian displacement reached its 10th year, our results show that the livelihood of Syrians in Turkey is very much tied to the functioning of the labour market, characterised by widespread informality. | medium |
| Engjom H; Aabakke A; Klunsoyr K et al. | 2021 | COVID-19 in pregnancy-characteristics and outcomes of pregnant women admitted to hospital because of SARS-CoV-2 infection in the Nordic countries | Finland, Denmark, Norway, Sweden, and Iceland | HIC | March 1, 2020 to June 30, 2020 | International migrants (%) | health outcomes of disease: hospitalisation | Cross-Sectional Study | The risk of admission due to COVID-19 was 0.4/1000 deliveries in Denmark, Finland and Norway, and 3.8/1000 deliveries in the Swedish regions. Women hospitalized because of COVID-19 were more frequently obese ($p<0.001$) and had a migrant background ($p<0.001$) compared with the total population of women who delivered in 2018. Twelve women (21.4%) needed intensive care. | This multinational Nordic study showed a low risk of admission due to COVID-19 in pregnancy in the Nordic countries. Women admitted to hospital due to COVID-19 were more frequently obese or had a migrant background compared with non-infected women. | high |
| Enriquez LE; Rosales WE; Chavarria K; Morales Hernandez M; Valadez M. | 2021 | COVID on Campus: Assessing the Impact of the Pandemic on Undocumented College Students | USA | HIC | March 2020 to June 2020 | Undocumented migrants | effect of lockdown measures | Mixed-Methods: Cross-Sectional Study; Qualitative research | Qualitative findings showed that immigration status exacerbated the negative economic impact of the pandemic, leading to severe individual and family financial strains that had cascading negative effects on undocumented students' academics and health. Quantitative findings focused on students' own prepandemic economic insecurity to show that it was associated with worse academic, financial, and health impacts during the initial months of the pandemic. | Qualitative findings highlighted how campus resources such as study space and technology were no longer available, harming students' academics. Thus, our findings suggest that the benefits of campus-wide resources, (…), depend on a continuous rather than cumulative provision of services. The quantitative results also demonstrated that being a student in the UC system, versus the CSU, was positively associated with increased negative academic, financial, and health impacts. Differences may emerge because UC campuses host a much larger number of on-campus residential students and offer more support services, thus inflicting a greater burden on UC undocumented students who were forced to relocate to their permanent homes and cut off from campus support. | high |
| Fabiani M; Mateo-Urdiales A; | 2021 | Epidemiological characteristics of COVID-19 cases in | Italy | HIC | February 20, 2020 | International migrants | health outcomes of disesase: | Ecological study | The results show that, at this stage of the COVID-19 epidemic in Italy, non-Italian cases were more likely to be hospitalized | In conclusion, our results suggest that, compared to Italian cases, non-Italian cases are more likely to experience a delayed | high |

| | | | | | | | | | | | |
|---|---|---|---|---|---|---|---|---|---|---|---|
| Andriano X et al. | | non-Italian nationals notified to the Italian surveillance system | | | to July 19, 2020 | | hospitalisation; ICU admission; Mortality | | (Adj. RR=1.39, 95% CI: 1.33-1.34) and to be admitted to ICU (Adj.RR=1.19, 95% CI: 1.07-1.32) compared with Italian cases, also showing a higher probability to die from the infection in those from low-HDI countries (Adj.RR=1.32, 95% CI: 1.01-1.75) [although this difference was not observed among hospitalized cases]. We observed an inverse gradient by which the risk of hospitalization, admission to ICU and death increased as the HDI of the country of origin decreased. | access to health services. Accessing care when the disease is more advanced and clinical conditions possibly more severe can explain why it is more likely for non-Italian nationals to require hospitalization and admission to ICU. Compared to Italian cases, differences in clinical outcomes appear more pronounced in non-Italian cases from countries with lower HDI and reduced in those from high-HDI countries. | |
| Fakhar-e-Alam Kulyar M; Bhutta ZA; Shabbir S; Akhtar M. | 2020 | Psychosocial impact of COVID-19 outbreak on international students living in Hubei province, China | China | UMIC | not reported | International students | effect of lockdown measures | Cross-Sectional Study | This study revealed some specific socio-psychological experiences of respondents. (...) students with longer stay in China reported more concerns and consequences than the students who stayed for a short period of time. This may be associated with the respondent's age and their marital status. | Our results are valuable for decision-makers and healthcare providers to develop effective interventions for international students. Based on the findings, we suggest to create a sense of security to eliminate the helplessness. Such integrative assessment can be achieved via modern communication platforms. | high |
| Fielding-Miller RK; Sundaram ME; Brouwer K. | 2020 | Social determinants of COVID-19 mortality at the county level | USA | HIC | 2018 to July 12, 2020 | International migrants | health outcomes of disease | Ecological study | We found that across all counties that had reported a case of COVID-19 as of July 12, 2020 (n=3024), a higher percentage of farmworkers, a higher percentage of residents living in poverty, higher density, higher population, and a higher percentage of residents over the age of 65 were all independently and significantly associated with a higher number of deaths in a county. In urban counties (n = 115), a higher percentage of farmworkers, higher density, and larger population were all associated with a higher number of deaths, while lower rates of insurance coverage in a county was independently associated with fewer deaths. | Taken together, our findings suggest that farm workers may face unique risks of contracting and dying from COVID-19, and that these risks are independent of poverty, insurance, or linguistic accessibility of COVID-19 health campaigns. | medium |
| Filippi D; Giliberti L. | 2021 | Italian reception policies and pandemic: From exclusion to abandonment | Italy | HIC | February 2020 to May 2020 | Asylum Seekers and Refugees | effect of lockdown measures | Qualitative research | By analyzing the transformation of Italian reception policies in the last years, the article shows the relationship between these changes and the condition of refugees and asylum seekers in these centers during the COVID-19 pandemic. Overcrowded housing, the absence of institutional guidance on managing the situation, and the interruption of many migrants' migratory projects are the main findings that emerged. | Our analysis encourages us to reflect upon two points. The first reflection is that the pandemic, as a new “total social event,” adds another layer of complexity to the Italian reception system, which was already under duress due to the Salvini Decree that introduced changes that did not benefit refugees and asylum seekers. The second reflection is that the pandemic has amplified and brought to the fore those pre-existing dynamics of marginalization and exclusion, | medium |

| | | | | | | | | | | | |
|---|---|---|---|---|---|---|---|---|---|---|---|
| | | | | | | | | | | which often feature in the stories of migrants who enter the Italian reception system. | |
| Fiorini G; Franchi M; Corrao G et al. | 2021 | Effects of SARS-CoV-2 pandemic on follow-up and pharmacological treatment of chronic diseases in undocumented migrants | Italy | HIC | January 2019 to to May 31, 2020 | Undocu mented migrants | health services access; effect of lockdown measures | Cross-Sectional Study | The number of consultations during the first 5 months of 2020 was much smaller than that of the same period of 2019. During the lockdown, we found 4048 consultations for 1914 patients, while they were 8051 in the same period of 2019 and 5681 in the first 2 months of 2020. The quantity of medicines dispensed by OSF showed a marked decrease in the period of the study and mainly during the lockdown. The decrease in consultations and dispensation of medicines was most evident for psychiatric patients and almost not existent for patients with diabetes. Female patients suffered a more pronounced reduction. | Western countries need strategies to better assist the very poor during epidemics. Differences among different groups of disadvantaged persons should be taken into account when designing recovery plans. | high |
| Fiorini G; Rigamonti AE; Galanopoul os C et al. | 2020 | Undocumented migrants during the COVID-19 pandemic: socio-economic determinants, clinical features and pharmacological treatment | Italy | HIC | February 24, 2020 to May 24, 2020 | Undocu mented migrants | risk of infection | Cross-Sectional Study | We analysed the records of 272 patients with respiratory issues attending the outpatient clinic of a large charity in Milan, Italy: amongst them, 18 had COVID-19 confirmed by rhino-pharyngeal swab and 1 of them deceased. All the patients attending the clinic appeared to have several risk factors for COVID-19 and chronic conditions suspected to predispose to the disease and/or to worsen severity and outcomes (...). | There is a lack of specific and systematic provisions for migrant populations in context of COVID-19 and cases are managed through ad hoc solutions, mostly through spontaneous initiatives by local NPOs or quarantine ships in case of sea arrivals, thus resulting in limited effectiveness of strategies to contain COVID-19 and unnecessary burden of disease. (...) Therefore, specific and systematic measures are needed to adequately extend coverage of epidemiological surveillance and healthcare systems and services in this population. | medium |
| Fouad FM; McCall SJ; Ayoub H; Abu-Raddad LJ; Mumtaz GR. | 2021 | Vulnerability of Syrian refugees in Lebanon to COVID-19: quantitative insights | Lebanon | UMIC | not reported | Refugees | risk of transmissio n | Modellin g Study | Mathematical modeling simulations we conducted illustrated that even modest increases in transmission among Syrian refugees could result in a large increase in the incidence and cumulative total number of infections in the absence of interventions. | In conclusion, while the young age structure of the Syrian refugee population might play a protective role against the scale and disease-burden severity of a potential COVID-19 epidemic, the epidemic potential due to several vulnerability factors warrants an immediate response in this population group. Local and international actors are required to mobilize and coordinate efforts to prevent the transmission of COVID-19, and to mitigate its impact amongst the vulnerable refugee populations globally. | low |
| Garcia E; Eckel SP; Chen Z, Li | 2021 | COVID-19 Mortality in California Based on Death Certificates: | USA | HIC | March 1, 2020 to July 31, 2020 | Internatio nal migrants | health outcomes of disesase | Cross-Sectional Study | MRR indicated elevated COVID-19 morality rates among Asian/Pacific Islander, Black, and Hispanic groups compared with the White group, with | Differential COVID-19 mortality was observed in California across racial/ethnic groups and by ethnicity and nativity combined, with evidence of greater | high |

| | | | | | | | | | | | |
|---|---|---|---|---|---|---|---|---|---|---|---|
| K; Gililand FD. | | Disproportionate Impacts Across Racial/Ethnic Groups and Nativity | | | | | | | Black and Hispanic groups having the highest MRR at 2.75 (95%CI: 2.54-2.97) and 4.18 (95%CI: 3.99-4.37), respectively. Disparities were larger at younger ages. Similar results were observed with PMR [proportionate mortality rates], and patterns of age-racial/ethnic disparities remained in analyses stratified by education. | disparities among younger age groups. Patterns of disparities persisted after accounting for educational attainment. Drivers of these disparate COVID-19 impacts are likely multifactorial and involve the interplay of structural, social, and individual factors that must be considered in the design and implementation for public health interventions to effectively mitigate the impacts of this disease. | |
| Gele A; Sheikh NS; Kour P; Qureshi SA. | 2022 | Uptake of Covid-19 preventive measures among 10 immigrant ethnic groups in Norway | Norway | HIC | April 21, 2020 to May 15, 2020 | International migrants | effect of lockdown measures; mental health | Qualitative Research | Mild depression (9.1%; 40/441) and anxiety (16.1%; 71/441) was common among respondents followed by moderate depression and anxiety {depression (3.4%; 15/441), anxiety (4.1%; 18/441)} and severe depression and anxiety {depression (1.1%; 5/441), anxiety (0.7%; 3/441)}. Anxiety and depression were independent of their socio-demographic characteristics. Perceived fear of contracting COVID-19, severity and death were prominent among the respondents. Respondents experienced stigma and discrimination in addition to being at the risk of disease and possible loss of employment and financial responsibilities. In addition, poor (quality and access to) health services, and poor living condition at the quarantine centres adversely affected respondents' mental health. | Although no statistically significant association of anxiety and depression with socio-demographic variables was observed, the qualitative approach identified various factors to contribute for their [returnee labor migrants] poor mental health. Institutional quarantine centers were in poor conditions in relation to basic supplies, health, hygiene and recreational support which adversely impacted mental health of the respondents. Measures towards alleviating fear and stigma and ensuring financial securities of the population during the time of health crisis are important for preparedness against epidemics such as COVID-19. | medium |
| Geuijen P; Vromans L; Embregts P. | 2021 | A qualitative investigation of support workers' experiences of the impact of the COVID-19 pandemic on Dutch migrant families who have children with intellectual disabilities | Netherlands | HIC | May 2020 to June 2020 | International migrants | effect of lockdown measures | Qualitative Research | Through taking into account experiences of support workers, this study reported on the impact of the COVID-19 pandemic upon Dutch migrant families who have children with ID. Two main themes related to the COVID-19 pandemic were identified: Work of support workers during the COVID-19 pandemic and Impact of the COVID-19 pandemic upon migrant families who have children with ID. Given that video calling often proved to be an unsuitable alternative (children did not understand) through which to stay in touch, parents missed their children very much. | The current study makes us cognisant of the fact that migrant families who have children with ID have encountered unique challenges (language barriers and emergency care plan) during the COVID-19 pandemic, whereas other challenges (additional burden and missing children who live at residential facilities) are in accordance with the challenges experienced by families not from a migrant background. Consequently, in light of the potential future waves of COVID-19 and expected further lockdowns, it is important that support workers are aware of the importance of language barriers, cultural background, and the increased burden associated with staying in touch with migrant families who have children with ID. | medium |

| | | | | | | | | | | | |
|---|---|---|---|---|---|---|---|---|---|---|---|
| Gilman R; Mahroof-Shaffi S; Harkensee C; Chamberlain A. | 2020 | Modelling interventions to control COVID-19 outbreaks in a refugee camp | Greece | HIC | not reported | Refugees | risk of transmission | Modelling Study | Spatial subdivision of the camp ('sectoring') was able to 'flatten the curve', reducing peak infection by up to 70% and delaying peak infection by up to several months. The use of face masks coupled with the efficient isolation of infected individuals reduced the overall incidence of infection, and sometimes averted epidemics altogether. These interventions must be implemented quickly in order to be maximally effective. Lockdowns had only small effects on COVID-19 dynamics. | Agent-based models are powerful tools for forecasting the spread of disease in spatially structured and heterogeneous populations. Our findings suggest that feasible interventions can slow the spread of COVID-19 in a refugee camp setting, and provide an evidence base for camp managers planning intervention strategies. Our model can be modified to study other closed populations at risk from COVID-19 or future epidemics. | medium |
| Giorgi J; Boertien D. | 2021 | The potential impact of co-residence structures on socio-demographic inequalities in COVID-19 mortality | France | HIC | May 27, 2020 to February 22, 2021 (census data 2009-2013) | International migrants | health outcomes of disease | Modelling Study | We found that education, citizenship, and place of birth of the head of household are all related to this expected number of deaths. The simulations showed that the infection of lower educated individuals, of persons who were born abroad, and of non-citizens are indeed expected to lead to more deaths related to within household transmission of the virus as compared the infection of higher educated individuals, the native born and citizens. However, an interesting element that came forward in the analysis is that these socio-economic gradients in vulnerability reverse with age. | Also, by decomposing vulnerability inequalities on the basis of two criteria—namely the age and socio-demographic category of the primarily infected person—we posit that the social gradient in COVID19 vulnerability linked to within-household transmission is subject to finer logic than the social gradient in mortality usually presented in the literature as its direction changes with the age of the initially infected person. | low |
| Giorgi Rossi P; Marino M; Formisano D; Venturelli F; Vicentini M; Grilli R. | 2020 | Characteristics and outcomes of a cohort of COVID-19 patients in the Province of Reggio Emilia, Italy | Italy | HIC | February 27, 2020 to April 2, 2020 | International migrants | health outcomes of disease: hospitalisation, mortality | Cohort Study | Females had higher prevalence of infection than males below age 50 (2.61 vs. 1.84 ‰), but lower in older ages (16.49 vs. 20.86‰ over age 80). (…) After adjusting for age and comorbidities, men had a higher risk of hospitalization (HR 1.4 95% CI 1.2 to 1.6) and of death (HR 1.6, 95% CI 1.2 to 2.1). (…) Immigrants had a higher risk of hospitalization (HR 1.3, 95% CI 0.99 to 1.81) than Italians and a similar risk of death. | Identified susceptible populations and fragile patients should be considered when setting priorities in public health planning and clinical decision making. | high |
| Goodman J; Wang S; Guadrrama R; Hernandez M. | 2020 | Mental health of undocumented college students during the COVID-19 pandemic | USA | HIC | May 15, 2020 to May 22, 2020 | Undocumented migrants | mental health | Cross-Sectional Study | We observed remarkably high clinical levels of anxiety and depression: 47% of the dreamers met the clinical cutoff for anxiety, 63% met the cutoff for depression, and 67% (2 in 3) met the cutoff for anxiety and/or depression. Rates of anxiety and depression in our sample were significantly higher than those | The present findings are consistent with recent predictions by social scientists that the COVID-19 pandemic would have a disproportionately negative impact on the mental health of undocumented immigrants. | medium |

| | | | | | | | | | | | |
|---|---|---|---|---|---|---|---|---|---|---|---|
| | | | | | | | | | recently reported for college students overall, suggesting that dreamers may be experiencing a more severe mental health impact of the COVID-19 pandemic. We also found that pandemic-induced concerns about finances, COVID-19 infection, immigration, and unemployment (among other factors) were associated with greater anxiety, stress, and depression among the dreamers in our sample. | | |
| Gorny AW; Bagdasarian N; Koh AHK et al. | 2021 | SARS-CoV-2 in migrant worker dormitories: Geospatial epidemiology supporting outbreak management | Singapore | HIC | April 3, 2020 to June 10, 2020 | Migrant workers | risk of infection | Cohort Study | Multiple transmission peaks were identified involving at least 5467 cases of SARS-CoV-2 infection across six dormitories. Our geospatial heat maps gave an early indication of outbreak severity in affected buildings. We found that the number of cases of SARS-CoV-2 infection doubled every 1.56 days (95% CI 1.29–1.96) in barracks-style buildings. The corresponding doubling time for apartment-style buildings was 2.65 days (95% CI 2.01–3.87). | Geospatial epidemiology was useful in shaping outbreak management strategies in dormitories. Our results indicate that building design plays an integral role in transmission and should be considered in the prevention of future outbreaks. | medium |
| Gosselin A; Longchamps C; Oulahal R et al. | 2021 | Social and mental health risks faced by undocumented migrants during the COVID-19 pandemic: Evidence from three surveys in France | France | HIC | April 1, 2020 to June 7, 2020 | Undocumented migrants | mental health | Cross-Sectional Study | Undocumented migrants had a higher probability of experiencing food insecurity increase (aORs=10.40 [3.59, 30.16], and 2.19 [1.39, 3.50]in APART TOGETHER and ECHO), a higher probability of depression (aOR=2.65 [1.01, 6.97]in MAKASI). In all three surveys, undocumented migrants were more likely to lose their job (aORs=6.51 [1.18, 36.00], 8.36 [1.08, 64.70] and 3.96 [1.79, 9.16]in APART TOGETHER, MAKASI and ECHO respectively). | Our results suggest that the lives of undocumented migrants have been dramatically worsened by the COVID-19 pandemic, exposing and amplifying the inequalities facing this group. There is an urgent need for action to address these inequalities. | high |
| Gosselin A; Melchior M; Carillon S et al. & MAKASI Study Group. | 2021 | Deterioration of mental health and insufficient Covid-19 information among disadvantaged immigrants in the greater Paris area | France | HIC | April 1, 2020 to June 7, 2020 | International migrants | mental health | Cohort Study | Among the 100 participants, 68% had no legal residence permit. Food insecurity was more often reported during lockdown than before (62% vs 52%). 9% of participants had a score indicative of severe depression (PHQ9) before lockdown and 17% afterwards (p = 0.17). Only 51% knew about the possibility of asymptomatic transmission of the COVID-19 virus. | This study brings original information on a hard-to-reach population group. Our results suggest that the lockdown had a detrimental impact on various economic and mental health aspects among disadvantaged migrants residing in the Greater Paris area. | medium |
| Gosselin A; Melchior | 2021 | Were immigrants on the frontline during the lockdown? | France | HIC | not reported | International migrants | risk of infection | Cross-Sectional Study | Compared to natives (Metropolitan France), being an African immigrant was associated to higher probability to work in | These results plead for taking occupational exposure into account in exploring Sars- | high |

| | | | | | | | | | | | |
|---|---|---|---|---|---|---|---|---|---|---|---|
| M; Desprat D et al. | | Evidence from France | | | | | | | a frontline job [adjusted odds ratio (aOR) 1.82 (1.23-2.71)], aswell as being born in French Overseas Departments [aOR 1.64 (1.23-2.18)], reflecting racial division of work and higher Sars-Cov-2 exposure of immigrant and minority populations. | Cov-2 mortality disparities according to sex and migratory status. | |
| Guijarro C.; Perez E.; Gonzalez B. et al. | 2021 | Differential risk for COVID-19 in the first wave of the disease among Spaniards and migrants from different areas of the world living in Spain | Spain | HIC | February 1, 2020 to April 25, 2020 | Internatio nal migrants | health outcomes of disease: SARS-CoV-2 cases | Cohort Study | The crude cumulative incidence among migrants(n=20419) was higher than among Spaniards(n=131599): 8.81 and 6.51 and per 1000 inhabitants ,respectively(p<.001),but differed by region of origin. | Our data suggest ethnic background may play a role in risk for COVID-19. Migrants from some areas of the world may merit closer attention for both clinical and epidemiological reasons. | high |
| Gulati A; Jose S; Singh BB. | 2021 | COVID-19: Emergence, Spread and Its Impact on the Indian Economy and Migrant Workers (preprint) | India | LMIC | June 2020 to April 2021 | Migrant workers | effect of lockdown measures | Cohort Study | The sudden imposition of the lockdown had a severe impact not only on employment but consequently on the earnings and savings of these migrants once they reached their villages. At their native place, with no proper employment opportunities, the household income of migrants fell by 85 percent during June-August 2020, as per the survey finding. (...) Although the migrant's household income has increased after remigration, there is still a contraction of 7.7 percent relative to the pre-lockdown level. The household income of the migrants who are still at their native place post-lockdown contracted more than 82 percent compared to pre-lockdown. | The pandemic has exposed these migrants to vulnerabilities that transcended employment and income loss to food and nutritional insecurity. The implementation of 'One nation, one ration card' to provide PDS entitlements at the destination, with an option to receive cash or grain in kind, needs to be fastened. | medium |
| Guruge S; Lamaj P; Lee C et al. | 2020 | COVID-19 restrictions: experiences of immigrant parents in Toronto | Canada | HIC | First six months of the pandemic (summer and fall 2020) (not further specified ) | Internatio nal migrants | effect of lockdown measures | Qualitati ve research | Results revealed that parenting experiences during the pandemic entailed dealing with changing relationships, coping with added burdens and pressures, living in persistent fear and anxiety, and rethinking lifestyles and habits. Amid these changes and challenges, some parents managed to create opportunities for their children to improve their diet, take a break from their rushed lives, get in touch with their cultural and linguistic backgrounds, and spend more quality time with their family. | While immigrant parents exhibit remarkable resilience in dealing with the pandemic-related meso and macro-levels restrictions, funding and programs are urgently needed to support them in addressing the impact of these at the microlevel. | high |

| | | | | | | | | | | | |
|---|---|---|---|---|---|---|---|---|---|---|---|
| Guttmann A; Gandhi S; Wanigaratne S et al. | 2020 | COVID-19 in Immigrants, Refugees and Other Newcomers in Ontario: Characteristics of Those Tested and Those Confirmed Positive, as of June 13, 2020. | Canada | HIC | January 15, 2020 to June 13, 2020 | Refugees and International migrants | health outcome of disease: risk of infection; effect of lockdown measures | Cross-Sectional Study | Although immigrants, refugees and other newcomers make up just over 25% of the Ontario population, they accounted for 43.5% of all COVID-19 cases. Rates of testing were lower for immigrants and refugees compared with Canadian-born and long-term residents. (...) Employment as a health care worker, especially among women, accounted for a disproportionate number of COVID-19 cases among immigrants and refugees. | The systemic inequities that have resulted in income disparities and related precarious employment and housing for many immigrants and refugees will need multi-faceted societal change. In the immediate term, a number of public health initiatives may help to mitigate the disproportionate burden of COVID-19 infections as Ontario prepares for the possibility of a second wave of infections in the fall. This includes initiatives such as more accessible testing options, including mobile testing to help identify and address barriers to testing. | medium |
| Habtamu K; Desie Y; Asnake M; Lera EG; Mequanint T. | 2021 | Psychological distress among Ethiopian migrant returnees who were in quarantine in the context of COVID-19: institution-based cross-sectional study | Ethiopia | LIC | May 1, 2020 to June 15, 2020 | Retournees | mental health | Cross-Sectional Study | More than half of the participants had depressive symptoms; 22.0% had severe or extremely severe symptoms of depression. Nearly half of the participants had anxiety symptoms, with 26.1% severe or extremely severe anxiety symptoms. More than a third of the participants had stress symptoms, with 13.4% sever or extremely severe stress symptoms. | We found a very high prevalence of depressive, anxiety and stress symptoms among Ethiopianmigrant returnees who were in quarantine due to the COVID-19 pandemic. Screening, integration of mental healthservices with other socioeconomic and psychosocial services, and effective and efficient referral may be useful to address the burden of psychological distress in this group. | high |
| Hajjar MS.; Abu-Sittah GS. | 2021 | The multidimensional burden of COVID-19 on Syrian refugees in Lebanon | Lebanon | UMIC | May 2020 | Refugees | mental health | Cross-Sectional Study | During the pandemic, 79% of breadwinners lost their jobs; of those who kept their jobs, 68% had their wages reduced. None of the families was capable of affording all of their basic needs with 55% only partially affording and 45% not able to afford. Thirty percent of Syrian refugee families did not receive support from organizations during the pandemic reflecting the impact of this crisis on humanitarian organizations. Education was also affected as 70% of children did not continue their education at home. Stress and anxiety were the most commonly reported behavioral changes among both children and adults. | The impact of this crisis is multidimensional affecting the economy, global health and education level of the public. Measures should be taken to lessen the detrimental effect of this crisis on the community as a whole and on vulnerable populations in particular. | medium |
| Hamadah H; Alahmad B; Behbehani M et al. | 2020 | COVID-19 clinical outcomes and nationality: results from a Nationwide registry in Kuwait | Kuwait | HIC | February 24, 2020 to April 20, 2020 | International migrants | health outcomes of disease: ICU admission, Mortality | Cross-Sectional Study | We found non-Kuwaitis were two times more likely to be admitted to the ICU (OR: 1.92, 95% CI 0.86-4.72) or die (OR: 2.19, 95% CI 0.88-6.02) from COVID-19. With adjustment to age, smoking and certain co-morbidities, non-Kuwaitis had two- and three-fold increase in odds ratios for ICU admission and death, respectively. | The current pandemic could amplify societal and structural inequalities that have long existed in the public health realm. The data on the differences of COVID-19 outcomes among marginalized subpopulations is now growing. In Kuwait we found non-Kuwaitis to be two times more likely to be admitted to the ICU or die from COVID-19. | high |

| | | | | | | | | | | | |
|---|---|---|---|---|---|---|---|---|---|---|---|
| Hamdan A; Ghanim M; Mosleh R. | 2021 | COVID-19 confinement and related well being measurement using the EQ-5D questionnaire: A survey among the Palestinian population | Palestine Territories | n.a. | November 2020 to December 2020 | Refugees | effect of lockdown measures; mental health | Cross-Sectional Study | The statistical analysis showed that factors significantly associated with more reported problems in at least one EQ-5D dimension ($P < .05$) were: females, ageing, being unmarried, low income, school studies, living in refugee camps, and villages, unemployment, having chronic diseases or pain, and obesity. | This work provides important evidence on the health status and well-being during the COVID-19 confinement in a sample of the Palestinian population, affecting almost all the aspects of the health state and wellbeing. This effect could be minimised by improving the COVID-19 preventive education and monitoring that can play an important role in all health and life aspects among the Palestinian population in facing this pandemic. | high |
| Han L.; Jia J. | 2021 | Reductions of migrant population reduces the number of COVID-19 epidemic: a case study in China | China | UMIC | not reported | International migrants | risk of transmission | Modelling Study | The predictive results showed that, if no travel restrictions are imposed on the migrant population as usual, this number of COVID-19 would have reached 27,483 (95% CI 16,074, 48,097; the actual number was 23,177). The increase in one city (Jian) would be 577.23% (95% CI 322.73%, 972.73%) compared with the real confirmed cases of COVID-19. The average increase in 73 cities was 85.53% (95% CI 19.53%, 189.81%). (...) Without restrictions on migration, the high prevalence areas in Hubei province and its surrounding areas will be further expanded. | The reduced population mobility and population density have positive effects on slowing down the spread of the epidemic. In conclusion, we strongly recommend that other countries learn from China's experience, suspend the transportation between cities, comprehensively and strictly control the population travel, and decrease the population density, so as to reduce the spread of diseases. | low |
| Hargreaves S, Hayward S, Deal A et al. | 2021 | Reducing COVID-19 transmission and strengthening vaccine uptake in migrants in the EU | EU/ EEA countries and the UK | HIC | June 15, 2020 to March 10, 2021 | Asylum Seekers, Refugees, International migrants | effect of lockdown measures; health outcomes of disease; vaccination | Qualitative research | Some migrants have experienced a range of direct and indirect health and social impacts including increased discrimination, more protracted lockdowns and severe restrictions on movement, and travel restrictions and border closures impacting family reunifications and the asylum process. Migrants in precarious work environments may have been at increased risk of losing jobs and sources of income. Certain migrant groups have a range of risk factors and vulnerabilities to COVID-19 that put them at higher risk of exposure, including occupational risk, overcrowded accommodation (including residing in camps, detention facilities, and homeless shelters), and/or lower levels of accessibility to public health services and messaging. There is emerging evidence of low COVID-19 vaccination rates in some migrant and ethnic minority groups in the EU/EEA. | These findings have immediate implications for national public health responses to reduce transmission and support COVID-19 vaccine rollout, with implications beyond the pandemic to promote health equity, human rights, and universal health coverage in marginalised migrant populations. | medium |

| | | | | | | | | | | | |
|---|---|---|---|---|---|---|---|---|---|---|---|
| Hari A; Nardon L; Zhang H. | 2021 | A transnational lens into international student experiences of the COVID-19 pandemic | Canada | HIC | April 2020 to June 2020 | International students | effect of lockdown measures | Qualitative research | We found that international students experienced the pandemic transnationally and faced increased challenges, which heightened their reliance on support from transnational families, and generated anxieties about their future career and mobilities. | We know from this study that IS are drawing on their transnational resources, it is, therefore, imperative that we rethink locally based supports. (…) There is a need to provide more effective locally emplaced, pragmatic, emotional and social support. These recommendations require multi-stakeholder engagement and support, including universities (as key anchors for this group), governmental bodies (Immigration, Refugees and Citizenship Canada, Employment and Social Development Canada, as well as provinces and municipalities) and civil society organizations. | high |
| Hariri M; Rihawi H; Safadi S; McGlasson MA; Obaid W. | 2020 | THE COVID-19 FORECAST IN NORTHWEST SYRIA The Imperative of Global Action to Avoid Catastrophe | Northwest Syria | LIC | not reported | IDP | health outcomes of disease: forecasted cases | Modelling Study | Camp-population Scenario predicts 240,000 cases (20% of the IDP population) of which 36,000 are severe and 12,000 are critical and 14,328 deaths in the first 6 weeks. With only 2,429 inpatient beds and 240 ICU beds (98 with adult ventilators, 62 with paediatric ventilators) in northwest Syria, ward and ICU bed capacities will be overwhelmed within 4-7 weeks. | Should a COVID-19 outbreak occur in NW Syria, projected cases and deaths will be particularly severe among IDPs. Health system capacity will be overwhelmed within a short period after the first case in camp settings. There is need for further research to account for additional variables that can impact projections. | high |
| Harkness A; Weinstein E; Mayo D; Rodriguez-Diaz C; Safren S. | 2021 | Latinx Sexual Minority Men's Behavioral, Psychosocial, and Medical Experiences during COVID-19: Differences across Immigration Statuses | USA | HIC | February 18, 2020 to August 26, 2020 | International migrants | health outcomes of disease: SARS-CoV-2 cases; effect of lockdown measures | Cross-Sectional Study | Despite no difference in COVID-19 testing rates, established immigrants were 4.52 times more likely (95% CI: 1.60, 12.81) to report a COVID-19 diagnosis than US-born LSMM. Immigrants, compared to US-born LSMM, reported disproportionate financial stress during COVID-19. Recent immigrants were 2.75 times more likely than US-born LSMM to report financial loss (CI: 1.30, 5.82) and established immigrants trended in the same direction. | Not reported | medium |
| Holmberg V; Salmi H; Kattainen S et al. | 2022 | Association between first language and SARS-CoV-2 infection rates, hospitalization, intensive care admissions and death in Finland: a population-based | Finland | HIC | February 27, 2020 to August 3, 2020 | International migrants | health outcomes of disease: SARS-CoV-2 Cases, Hospitalisation, ICU admission, Mortality | Cohort Study | In total, 124 240 individuals were tested, and among the 118 300 (95%) whose first language could be determined, 4005 (3.4%) were COVID-19-positive, 623 (0.5%) were admitted to specialized hospitals, and 147 (0.1%) were admitted to the ICU; 254 (0.2%) died. Those with a foreign first language had lower testing rates (348, 95%CI 340-355 versus 758, 95%CI 753-762 per 10 000, p<0.0001), | The population with a foreign first language was at an increased risk for testing positive for SARS-CoV-2, but when hospitalized they had outcomes similar to those in the native, domestic language population. This suggests that special attention should be paid to the prevention and control of infectious diseases among language minorities. | high |

| | | | | | | | | | | | |
|---|---|---|---|---|---|---|---|---|---|---|---|
| | | observational cohort study | | | | | (risk of infection) | | higher incidence (36, 95%CI 33-38 versus 22, 95%CI 21-23 per 10 000, p<0.0001), and higher positivity rates (103, 95%CI 96-109 versus 29, 95%CI 28-30 per 1000, p<0.0001). | | |
| Horner KM; Wrigley-Field E; Leider JP. | 2021 | A First Look: Disparities in COVID-19 Mortality Among US-Born and Foreign-Born Minnesota Residents | USA | HIC | January 1, 2020 to December 31, 2020 | International migrants | health outcomes of disease: Mortality | Cross-Sectional Study | Foreign-born Minnesotans died of COVID-19 at twice the age- and gender-adjusted rate of the US-born (age-gender standardized mortality per 100,000:261.4 vs. 1290.7; difference=101.5%). | Disparities by race and ethnicity within US-born populations are profound, but so too are the disparities across US- and foreign-born groups. (…) The peaks in COVID-19 mortality at the end of 2020 and within long-term care facilities overshadow the experience of immigrant communities - which faced high early mortality rates and significant losses among those living outside long term care. | high |
| Im H; George N. | 2021 | Impacts of COVID-19 on Refugee Service Provision and Community Support: A Rapid Assessment during the Pandemic | USA | HIC | April 2020 to October 2020 | Refugees | effects of lockdown measures; health services access | Mixed-Methods: Cross-Sectional Study; Qualitative Research | Providers highlighted several preexisting health challenges that have been barriers for the refugee community before the pan-demic. The rapid assessment highlighted that underlying health concerns and limited orientation to the U.S. healthcare systems were identified as two main overarching themes that have caused challenges to the refugee community. (…) For refugees, the impact of the COVID-19 pandemic manifests through multiple factors such as health, social, and economic hardships, which were likely exacerbated by barriers to access to virtual service platforms and resources and communication breakdown with service providers. | This study proposes several implications for future research and policy in social work with refugee populations during the post-COVID time. (…) The COVID-19 pandemic uncovers cumulative issues that have obstructed healthcare and social services in the refugee community and demands a more culturally nuanced, multipronged approach to refugee community wellness and integration through building capacity and collaborative partnerships among professionals and members, and volunteers of the refugee community. | high |
| Imran J; Nasa P; Alexander L; Upadhyay S; Alanduru V. | 2021 | Psychological distress among survivors of moderate-to-critical COVID-19 illness: A multicentric prospective cross-sectional study | India | LMIC | March 2020 to June 2020 | Migrant workers | mental health | Cross-Sectional Study | In 103 patients (96% were immigrant workers), the prevalence rate of clinically significant anxiety, depression, and PTSD was 21.4%, 12.7%, and 8.7% at day 30 and 9.5%, 7.1%, and 4.7% at day 60, respectively. There was significantly higher anxiety in patients of Indian nationality and depression with preexisting chronic illness. | There is a high prevalence of clinically significant anxiety, depression, and posttraumatic stress among the survivors of moderate to critically COVID-19. We recommend formal evaluation by psychiatrists and long-term mental rehabilitation for survivors of COVID-19. | medium |
| Indseth T; Grosland M; Arnesen T et al. | 2021 | COVID-19 among immigrants in Norway, notified infections, related hospitalizations and associated mortality: | Norway | HIC | March 1, 2020 to October 18, 2020 | International migrants | health outcomes of disease: SARS-CoV-2 Cases, | Cross-Sectional Study | Immigrants had higher rates of notified COVID-19 (n per 100,000 = 567 vs.251) and related hospitalizations (n per 100,00 = 62 vs. 21) than non-immigrants. (…) if we adjust for age, the immigrants from AfAsSA have substantially higher | After the first weeks of the pandemic, immigrants in Norway have had higher rates than Norwegian-born residents of notified COVID-19 infections and related hospitalization, but with major differences across immigrant groups. (…) The | high |

| | | | | | | | | | | | |
|---|---|---|---|---|---|---|---|---|---|---|---|
| | | A register-based study | | | | | Hospitalisation, Mortality | | mortality rates than non-immigrants. For some immigrant groups, there is a high number of hospitalizations per notified case. Moreover, we also reported a high age-adjusted rate of COVID-19-related mortality among immigrants from Asia Africa and Latin America, corresponding to groups with high rates of hospitalizations. | relationship between notified infections and hospitalizations could indicate a higher level of unconfirmed cases in some groups. The much higher toll of notified COVID-19 infections among several immigrant groups compared to non-immigrants suggests a need for actions such as enhancing community engagement and health communication strategies to lower the thresholds for being tested. | |
| Inghammar M; Moghaddassi M; Rasmussen M; Malmqvist U; Kahn F; Bjork J. | 2021 | COVID-19 vaccine uptake among older people in relation to sociodemographic factors: cohort results from southern Sweden | Sweden | HIC | December 27, 2020 to June 4, 2021 | International migrants | vaccination | Cohort Study | Uptake of the first dose was high (91.9%) overall, but markedly lower (75.3%) among persons born outside the Nordic countries. Vaccine uptake was generally satisfactory among native Swedes also in areas with lower socioeconomic status, but dropped substantially among non-Nordic born in those areas. | In conclusion, the identified clusters of unvaccinated older people, mainly representing ethnic minorities in disadvantaged areas, is a public health concern that warrants intensified efforts regarding tailored communication, easier vaccine access, and engagement from local communities. Continuous monitoring of the vaccine uptake and immunity of the population is essential for pandemic control and for assessing risks for local or more general disease outbreaks. | high |
| Ingraham NE; Purcell LN; Karam BS et al. | 2021 | Racial and Ethnic Disparities in Hospital Admissions from COVID-19: Determining the Impact of Neighborhood Deprivation and Primary Language | USA | HIC | March 4, 2020 to August 19, 2020 | International migrants | health outcomes of disease: Hospitalisation | Cohort Study | Hospitalized patients were older (60.9 vs. 40.4 years, p< 0.001) and more likely to be male (n= 425 [49.1%] vs. 2049 [43.5%], p=0.002). Of those requiring hospitalization, 43.9% (n= 381), 19.9% (n= 172), 18.6% (n= 161), and 11.8% (n= 102) were White, Black, Asian, and Hispanic, respectively. Non-English-speaking (OR 1.91, 95% CI 1.51-2.43) significantly increased odds of hospital admission across and within minority groups. | Our findings highlight areas of neigh-bourhood-level deprivation may contribute to racial disparities but to a lesser degree when controlling individual-level factors. Strikingly, racial and ethnic disparities in COVID-19 illness severity exist, independent of socioeconomic characteristics, which supports the need to investigate the different levels of racism contributing to health inequity. | high |
| International Organization for Migration (IOM). | 2020 | IOM Ireland Rapid Policy Survey Report: The Irish Governmentals Policy Response to COVID-19 from a Migration Governance Lens (March 2020 - August 2020) | Ireland | HIC | May 2020 to June 2020 | International and undocuments migrants | effect of lockdown measures | Cross-Sectional Study | Despite efforts to increase beds and reduce the number of residents sharing bedrooms to allow for greater social distancing in Direct Provision centres, clusters have emerged in several centres across the country. (...) Those with limited incomes find it the most difficult to social distance or self-isolate and are more likely to live in cramped accommodation with non-relatives and work in jobs where it is not possible to social distance. | While addressing the COVID-19 crisis, it is important that governments maintain a longer-term, holistic perspective on migration governance. COVID-19 has exacerbated inequalities, impacting people most affected by social exclusion. Structural inequalities limit the effectiveness of measures taken to contain the spread of the virus. | low |

| | | | | | | | | | | | |
|---|---|---|---|---|---|---|---|---|---|---|---|
| International Organization for Migration (IOM). | 2020 | Rapid Needs Assessment: COVID-19 Related Vulnerabilities among Migrants in Armenia | Armenia | UMIC | April 2020 to May 2020 | International students, Migrant workers, Asylum seekers | health services access | Mixed-Methods | interviewed migrants highlighted financial difficulties both for themselves and for their communities, housing issues, access to health care, access to information and visa and migration issues. (...) Students had issues with education, such as online classes, tuition fees, etc., together with housing, residence rights and regularization of their status, as well as access to information and health issues. (...) Many migrants were employed without being formally contracted (...). | Even though in Armenia migrants are entitled to health-care services, provided they have relevant documentation, factors such as language barriers and lack of knowledge about where and to whom to apply resulted, on occasions, in insufficient access to health care. Some respondents said that they only visited doctors when they had serious problems, so they visited hospitals directly and do not use primary health-care facilities. | low |
| International Organization for Migration (IOM). | 2021 | Regional Assessment: Key Informant Survey on the Socioeconomic Effects of COVID-19 on Returnees and Stranded Migrants in Central Asia | Kazakhstan, Kyrgyzstan, and Tajikistan | MICs | January 2021 to February 2021 | International migrants, Retournees | effect of lockdown measures | Cross-Sectional Study | Kazakhstan: Overall, the most important unmet needs of stranded migrants were linked to employment/livelihoods, shelter/housing, reunification with family members who got separated during migration, and access to and replacement of personal and other documentation. (...) Kyrgyzstan: (...) there were some returnees without documents, and this can result in the inability to access basic services (health, education), humanitarian assistance, and legal employment. (...) In terms of specific problems affecting returnee children, it was found that working without going to school, substance abuse, marriage before 18, and not having a birth certificate were issues in both cities. (...)<br>Tajikistan: Overall, the most important unmet needs of returnees were linked to food, employment/livelihoods, and access to or replacement of personal documentation. (...) Other issues concerning returnee children were working without going to school, substance abuse, and experiencing physical violence. | The findings of this study can be used as a basis to identify new migration patterns, common problems and vulnerabilities, and better inform policy makers working on international migration challenges. In addition, the population numbers and locations identified under this study represent a unique type of baseline data which can be used to run additional surveys and research studies. | low |
| Irvine M; Coombs D; Skarha J et al. | 2020 | Modeling COVID-19 and Its Impacts on U.S. Immigration and Customs Enforcement (ICE) Detention Facilities, 2020 | USA | HIC | October 1, 2019 to March 1, 2020 | ICE detainee | risk of infection; transmission | Modelling Study | We found that 72% of individuals are expected to be infected by day 90 under the optimistic scenario (R0 = 2.5), while nearly 100% of individuals are expected to be infected by day 90 under a more pessimistic (R0 = 7) scenario. Although asynchronous outbreaks are more likely, day 90 estimates provide an | As policymakers seek to rapidly implement interventions that ensure the continued availability of life-saving medical resources across the USA, they may be overlooking the pressing need to slow the spread of COVID-19 infection in ICE's detention facilities. Preventing the rapid spread necessitates intervention measures such as granting ICE detainees widespread release | low |

| | | | | | | | | | | | |
|---|---|---|---|---|---|---|---|---|---|---|---|
| | | | | | | | | | approximation of total positive cases after all ICE facility outbreaks. | from an unsafe environment by returning them to the community. | |
| Islamoska S; Petersen JH; Benfield T; Norredam M. | 2021 | Socio-economic and Demographic Risk Factors in COVID-19 Hospitalization among Migrants and Ethnic Minorities | Denmark | HIC | February 1, 2020 to June 30, 2020 | International migrants | health outcomes of disease | Cohort Study | Among 2232 COVID-19 cases, the OR of hospitalization with COVID-19 among immigrants and descendants of Non-Western origin was 2.5 times higher (95% CI: 2.23 to 2.89) than that of the majority population, with most pronounced results among individuals from Iraq, Morocco, Pakistan, and Somalia.The OR was largely attributed to comorbidity and socioeconomic factors, especially household size, occupation, and population density. | There is a significantly higher OR of hospitalization with COVID-19 among Non-Western migrants and ethnic minorities compared with ethnic Danes. This knowledge is crucial for health policymakers and practitioners in both the current and future pandemics to identify more vulnerable groups and target prevention initiatives. | low |
| Jahn R; Hintermeier M; Bozorgmehr K. | 2022 | Sars-CoV-2 attack rate in reception and accommodation centres for asylum seekers: systematic review of outbreak media reports in Germany | Germany | HIC | January 27, 2020 to June 24, 2020 | Asylum Seekers | risk of infection | Cross-Sectional Study | The pooled SARS-CoV-2 attack rate was 13.1% (95% confidence interval, CI: 9.8–16.7). Outbreak sites implementing mass quarantine (n = 76) showed higher rates (15.7; 95% CI: 11.6–20.2) compared to sites using conventional strategies (6.6; 95% CI: 3.1–11.2), yielding a rate ratio of 0.44 (95% CI: 0.27–0.72) adjusted for testing strategies, type and size of accommodation. | Mass quarantine is associated with higher attack rates, and appears to be a counter-productive containment measure in overcrowded camps, but further research with individual-level data is required to rule out residual confounding. | high |
| Jaqueti Aroca J; Molina Esteban LM; García-Arata I; García-Martínez J. | 2020 | COVID-19 in Spanish and immigrant patients in a health area of Madrid | Spain | HIC | (data until) April, 2020 | International migrants | health outcomes of disease: SARS-CoV-2 cases (risk of infection) | Cross-Sectional Study | PCR was performed on 1,781 patients (1,522 Spanish and 259 immigrants), of which 919 have tested positive (51.6%; 782 Spaniards, 51.4%; and 136 immigrants, 52.5%). No differences were observed (Odds Ratio: 1.08; 95% CI: 0.95 - 1.24) in the rates of performing PCR in immigrants (8.4/1,000; 259/30, 723) compared to the Spanish (7.8/1,000; 1,522/195,350). (translated) | The specific situation of the patients in our study, all of whom were treated at the hospital, would make it difficult to extrapolate our data to patients in general. (translated) | medium |
| Jehn A.; Stackhouse M.; Zajacova A. | 2021 | COVID-19 Health Precautions: Identifying Demographic and Socio-Economic Disparities and Changes over Time | Canada | HIC | June to July 2020 | International migrants | effects of lockdown measures | Cross-Sectional Study | We find a nearly universal increase in precautionary behaviours from June to July in mask wearing (67.3 percent to 83.6 percent), social distancing (82.4 percent to 89.2 percent), and avoiding crowds (84.1 percent to 88.9 percent); no significant change occurred in the frequency of hand washing. We observe significant disparities in adherence to precautionary behaviours, especially for mask wearing, in June; female, older, immigrant, urban, and highly educated adults were | Our analysis of socio-demographic discrepancies identified groups with particularly low adherence, especially to mask wearing: men, rural residents, adults with lower education, and those with trade certificates. These findings should inform policy initiatives and targeted public health interventions. | high |

| | | | | | | | | | | | |
|---|---|---|---|---|---|---|---|---|---|---|---|
| | | | | | | | | | significantly more likely to adhere to precautionary behaviours than male, younger, Canadian-born, rural, and low-educated adults. | | |
| Johnson-Agbakwu CE; Eakin CM; Bailey CV et al. | 2021 | Severe acute respiratory syndrome coronavirus 2: a canary in the coal mine for public safety net hospitals | USA | HIC | May 6, 2020 to July 22, 2020 | Refugees | health outcomes of disease: SARS-CoV-2 Cases | Cross-Sectional Study | When refugee status was determined, 45 women (12.8%) were identified as refugees. Of the 45 refugee women, 8 (17.8%) tested positive for severe acute respiratory syndrome 2 compared with 25 non-refugee patients (8.19%) who tested positive for severe acute respiratory syndrome 2 (prevalence ratio, 2.16; 95% confidence interval, 1.04-4.51). In addition, 7 of the refugee women who tested positive for severe acute respiratory syndrome coronavirus 2 were from Central Africa. | Universal testing for SARS-CoV-2 on maternity units in these systems provides the opportunity to identify asymptomatic yet high-risk patient populations in need of targeted interventions to mitigate the spread of infection and facilitate a timely, culturally, and linguistically enhanced public health response. (…) Universal maternity testing is 1 step toward achieving health equity among vulnerable, otherwise hidden, communities. | medium |
| Kalani N; Hatami N; Ali S; Haghbeen M; Yaqoob U; Doost ER. | 2021 | Covid-19 health care for afghan refugees as a minor ethnicity in iran;clinical differences and racial equality in health | Iran | LMIC | January 2020 to Decembe r 2020 | Refugees | health outcomes of disease: ICU admission, Mortality | Case-Control Study | In this study 132 Afghan refugees were compared to 266 Iranian controls. (…) There was no difference in probability of being evaluated by HRCT or receiving ICU treatment (p = 0.173, 1, respectively) even after being adjusted for symptoms or co-morbidities that were manifesting differently between Afghan vs. Iranian patients (p = 0.476, 0.881, respectively). Ten (7.57%) subjects died in case group and 18 (6.76) in control group. There wasn't any significant difference in death rate between the groups (P=0.766). | Our research found significant variations in clinical characteristics between Afghan and Iranian COVID-19 patients, but these differences have little bearing on the likelihood of getting adequate medical treatment. Jahrom City was an example of COVID-19 health care equity for ethnic minorities. | high |
| Kamrujjam an Md; Mahmud MdS; Ahmed S. et al. | 2021 | SARS-CoV-2 and Rohingya Refugee Camp, Bangladesh: Uncertainty and How the Government Took Over the Situation | Banglade sh | LMIC | May 14, 2020 to August 12, 2020 | Refugees | risk of transmissio n | Modellin g Study | Our best fitting model suggests that there will be reasonable control over the transmission of the COVID-19 disease. At the end of December 2020, there will be only 169 confirmed COVID-19 cases in the Rohingya refugee camps. The average basic reproduction number (R0) has been estimated to be 0.7563. | Our analysis suggests that, due to the extensive precautions from the Bangladesh government and other humanitarian organizations, the coronavirus disease will be under control if the maintenance continues like this. However, detailed and pragmatic preparedness should be adopted for the worst scenario. | medium |
| Kassa SM; Njagarah JBH; Terefe YA. | 2020 | Modelling Covid-19 mitigation and control strategies in the presence of migration and vaccination: the case of South Africa | South Africa | UMIC | not reported | Internatio nal migrants | vaccination ; risk of transmissio n | Modellin g Study | Our results suggest that in presence of the vaccine,as long as migration is in place, even a 98% vaccination coverage may not be enough to curbnew infections. We therefore suggest that if a highly effective vaccine is made available, it may be necessary to implement universal coverage if the new infections are to be completely contained. | We also note however that, when immigration is allowed into the country, even a 98% vaccination coverage may not completely stop new infections. (...) That means, unless a country is completely isolated from the remaining world, or apply a mandatory vaccination for the incoming individuals, it is not possible to eradicate the infection from the country. We therefore, | medium |

| | | | | | | | | | | | |
|---|---|---|---|---|---|---|---|---|---|---|---|
| | | | | | | | | | | recommend that, if a vaccine is available, universal vaccination will be the ideal protocol if the new infections are to be curbed. | |
| Kaur-Gill S; Qin-Liang Y; Hassan S. | 2021 | Negotiating mental health during the COVID-19 pandemic: performing migrant domestic work in contentious conditions. (Special Issue: The COVID-19 pandemic: precarious migrants and outbreak inequality.) | Singapore | HIC | not reported | Migrant workers | mental health | Qualitative research | The findings in our article reveal the interplays of mental health meanings situated within a structural context of employment and a cultural environment that manifests unequal power relationships and indebtedness. During the Circuit Breaker, domestic workers in already poor employment conditions found themselves at greater precarity and limited agency. Furthermore, the analysis captures the temporalities of mental health meanings articulated within the performance of conducting migrant domestic work in confined working conditions. Precarious migrant journeys include vicious debt cycles, unethical agents, and corrupt employment practices, detailing the scripts of mental health stressors. | Our findings reveal that the COVID-19 pandemic lockdowns magnified how these practices of soft violence (…) manifested, disabling migrant worker mental health and well-being. As precarity is structured in the power imbalance to hire, fire, and deport domestic workers at will by employers, practical implications for mental health interventions must discuss labor and immigration policies of MDWs [migrant domestic workers] bound to Singapore. (…) Beyond culturally tailored programs on mental well-being in the workplace, solutions must consider the structural factors embedded in the policies of hire and the indentured nature of labor to address MDWs discussions of mental health contentions. | high |
| Khan MA; Khan MI; Illiyan A; Khojah M. | 2021 | The Economic and Psychological Impacts of COVID-19 Pandemic on Indian Migrant Workers in the Kingdom of Saudi Arabia | Kingdom of Saudi Arabia | HIC | April 2021 to May 2021 | Migrant workers | effect of lockdown measures | Cross-Sectional Study | The study revealed that majority of the migrants do not worked during the lockdown period and 42% of the migrants worked less than 8 hours in a day. (...) Around 61% of the migrants reported an average loss in salary of below SAR 500 and 26% reported a loss between SAR 500 to 1000, however only 12% reported a loss above SAR 1000 during lockdown period of the pandemic. It has also been observed that number of migrants who were remitting below SAR 1000 had increased and remittances by middle income group were found to be decreased during lockdown period of the pandemic. | The policy implications from the findings of the research draw attention of the policymakers towards protective measures need to be implemented to save migrants during ongoing pandemic. The government should take some necessary steps such as financial benefit scheme to overcome the problems in the reduction of migrant earnings and remittances. (...) The government should not focus only on vaccination and physical fitness of the migrants but also need to find out the cure of the psychological impact arising during the pandemic. | low |
| Khan S; Akbar SMF; Kimitsuki K et al. | 2021 | Recent downhill course of COVID-19 at Rohingya refugee camps in Bangladesh: Urgent action solicited | Bangladesh | LMIC | May 2020 to June 2021 | Refugees | risk of infection | Cross-Sectional Study | Seven out of 34 camps (...) reported more than 100 cases by early-July 2021. (...) Analyzing the severity of disease at presentation, about 15% of patients had moderate to severe disease (...). However, the disease condition of 80% of patients was unknown (data missing or under reported). There have been 20 deaths in the Rohingya Refugees from COVID-19 and 60% were female. | The COVID-19 pandemic has highlighted clear discriminatory limitations on access to health care facilities particularly hospital beds, oxygen supplies, intensive care capacities and vaccination for Rohingya refugees. Preparedness for COVID-19 in the refugee camps are affected by the lack of human resources, inadequate management, and laboratory and hospital facilities for testing and treating COVID-19. | low |

| | | | | | | | | | | | |
|---|---|---|---|---|---|---|---|---|---|---|---|
| Kim H; Lan K; Nkyekyer E et al. | 2020 | Assessment of Disparities in COVID-19 Testing and Infection Across Language Groups in Seattle, Washington | USA | HIC | February 29, 2020 to May 31, 2020 | International migrants | risk of infection | Cross-Sectional Study | Non-English speakers were overall less likely to have completed testing compared with English-speakers (4.7% [95% CI, 4.5%-4.9%] vs 5.6% [95% CI, 5.6%-5.7%]). Notably, the proportion of positive cases was 4.6-fold higher among non-English speakers overall (18.6%; 95% CI, 16.8%-20.4%) compared with English speakers (4.0%; 95% CI,3.8%-4.2%). | Our study found that despite the availability of interpreter services across clinical locations, non–English-speaking patients in our health system were tested less frequently for COVID-19 and had significantly higher burden of infection. (...) Our outreach efforts, which included mobile clinics and drive-up testing, was implemented in partnership with local health coalitions representing communities of color to overcome testing barriers and should be considered where limited access and disproportionate rates of infection are a concern. | low |
| Kizilhan JI; Noll-Hussong M. | 2020 | Psychological impact of COVID-19 in a refugee camp in Iraq | Iraq | UMIC | October 2019 to April 2020 | Refugees | mental health | Cohort Study | The prevalence of PTSD increased to ~58% (95% confidence interval [CI] = 34.1%–62.5%) in females and ~47% (95% CI = 28.5%–52.4%) in males. (...) The results of the IES-R for men and women together show a significant effect in the PTSD subdomains (...) The documented differences are highly statistically significant (P< 0.001) and canbe described as 'severe' in all patients. | Altogether, our analyses in a refugee camp provide additional evidence that being unable to obtain any medical support, the experience of repeated helplessness, and the loss of control seem to play an impressive role among those who still report suffering from traumatization and other mental disorders 5 years after experiencing genocide. | medium |
| Kjollesdal M; Magnusson K. | 2021 | Occupational risk of COVID-19 by country of birth. A register-based study | Norway | HIC | April 1, 2020 to December 2, 2020 | International migrants | health outcomes of disease: SARS-Cov-2 cases (risk of infection) | Cross-Sectional Study | Within each occupational group, immigrants had a greatly increased odds of COVID-19 when compared with Norwegian-born (odds ratio [OR] ~ 1.66–12.72). However, immigrants working in the selected occupations had the same odds of COVID-19 as person with same birth country not having the same occupation (OR ~ 1). Exceptions were Somalian, Afghani and Iraqi personal care workers in health services who had an increased odds of COVID-19 compared with others from the same country. | Immigrants from Somalia, Pakistan, Iraq, Afghanistan and Turkey holding various occupations implying close contact with others did not have higher odds of notified infection than others with the same country of birth, except for health care workers. Thus, occupation did not seem to play an important role in the excess risk of COVID-19 infection among immigrants. Explanations for the high rate of infection is likely related to a complex set of factors including factors related to exposure and to underlying health. | high |
| Kjollesdal M; Skyrud K; Gele A et al. | 2021 | The correlation between socioeconomic factors and COVID-19 among immigrants in Norway: a register-based study | Norway | HIC | March 1, 2020 to November 15, 2020 | International migrants | health outcomes of disease: SARS-Cov-2 cases, ICU admission (risk of infection) | Cross-Sectional Study; Ecological Study | The SES of immigrants at the group level was related to rates of notified infections and related hospitalisations. There was a strong correlation [pearson correlation index] between notified infections [per 100,000] and crowded housing (R= 0.77; p<0.01), whereas the correlations between notified infections and income (R=-0.52; p<0.05), unemployment R=0.43; p<0.05) | Crowded housing was correlated with both COVID-19 infections and related hospitalisations. Further, low income, low education and unemployment at the group level were moderately correlated with rates of COVID-19 infections. Other characteristics of immigrant groups with disadvantaged socio-economic position may be equally important for high rates of COVID-19. Regardless of the cause of | high |

| | | | | | | | | | | | |
|---|---|---|---|---|---|---|---|---|---|---|---|
| | | | | | | | | | and education (R=-0.47; p<0.05) were moderate. | infections, these groups are important to target with preventive measures. In particular, targeted interventions should be directed to low-income families living in overcrowded households. | |
| Knights F; Carter J; Deal A. et al. | 2021 | Impact of COVID-19 on Migrants' Access to Primary Care:A National Qualitative Study | United Kingdom | HIC | June 18, 2020 to November 30, 2020 | Asylum Seekers and Refugees | effect of lockdown measures | Qualitative research | It was found that digitalisation had exacerbated existing inequalities regarding access for some migrant groups through a lack of access to, or knowledge of, technology; concerns about language barriers, difficulties building trust, and the risk of missing safeguarding cues in virtual consultations were also expressed. The physical closure of some surgeries was reported to have led to challenges in migrants registering with, and accessing, primary care. Communication barriers, feeling left behind in receiving support and health interventions in comparison to the general population, and a lack of access to information were issues widely raised by migrants. Additionally, they reported views of COVID-19 and COVID-19 vaccinations that ranged from acceptance to misinformation, often originating from social media or word of mouth. Some migrants experienced increased risk factors to their health and severe illness from COVID-19, partially resulting from their economic and social situations. | Pandemic-related changes to primary care delivery may become permanent; some migrant groups are at risk of digital exclusion and may need targeted additional support to access services. Solutions are needed to address vaccine hesitancy in marginalised groups to ensure equitable COVID-19 vaccine uptake. | high |
| Koh D. | 2020 | Migrant workers and COVID-19 | Singapore | HIC | March 2020 to May 2020 | Migrant workers | risk of infection | Ecological Study | (...) the majority of cases occurred among an estimated 295000 low-skilled migrant workers living in foreign worker dormitories. As of 6 May 2020, there were 17758 confirmed COVID-19 cases among dormitory workers (88% of 20198 nationally confirmed cases). One dormitory housing approximately 13000 workers had 19.4% of residents infected. | The COVID-19 pandemic has highlighted migrant workers as a vulnerable occupational group. Ideally, matters related to inadequate housing of vulnerable migrant workers need to be addressed before a pandemic. | low |
| Kondilis E; Papamichail D; McCann S et al. | 2021 | The impact of the COVID-19 pandemic on refugees and asylum seekers in Greece: A retrospective analysis of national | Greece | HIC | February 26, 2020 to November 15, 2020 | Asylum Seekers and Refugees | health outcomes of disease: SARS-Cov-2 cases (risk of | Cross-Sectional Study | Twenty-five COVID-19 outbreaks were identified in refugee and asylum seeker reception facilities, with 6 (85.7%) of 7 RICs and 18 (56.3%) of 32 RSs reporting at least one outbreak during the study period. The overall 9-month COVID-19 IP among refugee and asylum seeker | Our data quantitatively shows the greater risk of COVID-19 infection among refugee and asylum seeker populations residing in sub-standard living conditions. Acknowledging this higher risk is of crucial policy importance. Going forward, all enclosed population groups need to be | high |

| | | | | | | | | | | | |
|---|---|---|---|---|---|---|---|---|---|---|---|
| | | surveillance data from 2020 | | | | | infection/transmission) | | populations residing in RSs on the Greek mainland was 1758 cases per 100,000 population; in RICs the incidence was 2052 cases per 100,000 population. Compared to the general population the risk of COVID-19 infection among refugees and asylum seekers in reception facilities was 2.5 to 3 times higher (p-value <0.001). | urgently included and prioritized in the national vaccination program, without any exception. (…) Greek authorities should concurrently urgently focus on improving living conditions in these facilities, decongesting those that operate beyond their capacity, reinforcing epidemiological surveillance, testing strategies and data transparency, and ensuring free, comprehensive and universal access to care without any discrimination. | |
| Kraft KB; Elgersma I; Lyngstad TM; Elstrom P; Telle K. | 2021 | COVID-19 vaccination rates among health care workers by immigrant background. A nation-wide registry study from Norway | Norway | HIC | December 2020 to August 31, 2021 | Migrant workers | vaccination | Cross-Sectional Study | Among all HCWs in Norway, immigrants had a 9 percentage point lower vaccination rate (85%) than HCWs without an immigrant background (94%) at 31 August 2021. The overall vaccination rate varied by country of birth (…) When we adjusted for demographics and detailed occupational codes, immigrant groups that more often worked as health care assistants, such as immigrants from Eritrea and Somalia, increased their vaccination rates. | Substantial differences in vaccination rates among immigrant groups employed in the healthcare sector in Norway indicate that measures to improve vaccine uptake should focus on specific immigrant groups rather than all immigrants together. Lower vaccination rates in some immigrant groups appear to be largely driven by the occupational composition, suggesting that some of the differences in vaccine rates can be attributed to variation in vaccine access. | high |
| Kuhlmann E; Falkenbach M; Klasa K; Pavolini E; Ungureanu M. | 2020 | Migrant carers in Europe in times of COVID-19: a call to action for European health workforce governance and a public health approach | Austria, Germany, Italy, Poland, Romania | HIC | January 2020 to May 20, 2020 | Migrant workers | effect of lockdown measures | Qualitative research | Results suggest that undersupply of carers coupled with cash benefits and a culture of family responsibility may result in high inflows of migrant carers, who are channelled in low-level positions or the informal care sector. COVID-19 made the fragile labour market arrangements of migrant carers visible, which may create new health risks for both the individual carer and the population. | The results highlight the weaknesses of existing health labour market arrange-ments in the LTC sector, which stretch far beyond poor workforce management. As the COVID-19 pandemic revealed, these conditions may directly impact population health and the health and wellbeing of the migrant carers, thus becoming fundamentally a public health policy issue. The research also calls for European health labour market regulation and governance models, which help to balance national interests and connect health system needs, health labour markets and the individual migrant carers. Including LTC migrant carers more systematically in health work-force governance and research, therefore, must become an issue of public health and European policy. | high |
| Kukreja R. | 2021 | Covid-19, racial capitalism, and undocumented bangladeshi | Greece | HIC | July 2019 to July 2020 | Undocumented migrants | effect of lockdown measures | Qualitative research | Bangladeshi men, emasculated by their illegality, "willingly" accept exploitative work arrangements (...) Greek farmers do not provide housing to their 10,000 odd farmhands, nor do the locals easily rent | Race-based devaluation of workers intersects with migrant illegality and culturally-specific masculine norms to normalize a discourse of healthcare “undeservingness” for undocumented | low |

| | | | | | | | | | | |
|---|---|---|---|---|---|---|---|---|---|---|
| | | agricultural workers in manolada, greece | | | | | | | accommodations. The workers are forced to rent unused farmland and build makeshift shacks colloquially called Barangas (slang for the Greek word, faranga, or tent) out of salvaged plastic sheets, cardboard, and reeds. (...) Reluctance to access health care due to past discriminations makes the workers choose to become invisible to the health system, thus creating a falsity about their health status. (...) Those who were not hired for the day - they had to stay inside. | racialized migrants. Unfree labor is legislated through restrictive migrant labor laws and selective detention and deportation of "illegal" migrants. Structural and systemic discriminations increase health precarities for undocumented agricultural workers. | |
| Kumar K; Mehra A; Sahoo S; Nehra R. | 2020 | The psychological impact of COVID-19 pandemic and lockdown on the migrant workers: A cross-sectional survey | India | LMIC | during the 5th week of lockdown (not further specified) | Migrant workers | mental health | Cross-Sectional Study | About three fourth of the participants (73.5 %) were found to be screen positive for depression on the PHQ-2 and about half of the participant (50%) were found to be screen positive for anxiety on the GAD-2. | the present study suggests that the current COVID-19 pandemic is causing severe anxiety and depressive symptoms in migrants' workers. | high |
| Kumar N; Udah H; Francis A; Singh S; Wilson A. | 2021 | Indian Migrant Workers' Experience During the COVID-19 Pandemic Nationwide Lockdown | India | LMIC | March 24, 2020 to May 31, 2020 | Internal migrants | mental health | Qualitative research | (…), the findings suggest that the participants' plights have been exacerbated by the COVID-19 pandemic. (…) As our participants reported, they dealt with multiple hardships, including loss of jobs and income, police brutality, Othering, and differential treatment, difficulties accessing health services, food, cash transfer and other social programmes during the first COVID-19 pandemic nationwide lockdown. (…) Consequently, many of them became anxious, and were uncertain about their future. With no money and savings, many participants struggled to live and survive during the lockdown. | The findings indicate the need for policy responses to focus on addressing conditions of work, terms of employment and access to necessities for Indian MW, including ensuring conditions for a prompt job-ready recovery and mental health care after the COVID-19 pandemic. | medium |
| Labberton A; Godoy A; Elgersma I. et al. | 2021 | SARS-CoV-2 infections and hospitalizations among immigrants in Norway: significance of occupation, household crowding, education, household income | Norway | HIC | June 15, 2020 to March 31, 2021 | International migrants | health outcomes of disease: SARS-Cov-2 cases, Hospitalisation (risk of infection) | Cross-Sectional Study | Confirmed infections in this period (per 100000): foreign-born 3140, Norwegian-born with foreign-born parents 4799, Norwegian-born with Norwegian-born parent(s) 1011. Hospitalizations (per 100000): foreign-born 147, Norwegian-born with foreign-born parents 47, Norwegian-born with Norwegian-born parent(s) 37. The addition of socioeconomic and medical factors to the base-model (age, sex, municipality of | Residents with foreign backgrounds have, as a group, been disproportionately hit by COVID-19 in Norway. Adjustment for socioeconomic factors and medical risk attenuates the over-representation moderately; however, the overall picture remains the same. the data available or variable definitions may not have fully captured the effects and interactions of these factors, and future studies should aim to unravel this further. | high |

| | | | | | | | | | | | |
|---|---|---|---|---|---|---|---|---|---|---|---|
| | | and medical risk. A nationwide register study | | | | | | | residence) attenuated excess infection rates by 12.0% and hospitalizations by 3.8% among foreign-born, and 10.9% and 46.2% respectively among Norwegian-born with foreign parents, compared to Norwegian-born with Norwegian-born parent(s). | | |
| Lauzardo M; Kovacevich N; Dennis A; Myers P; Flocks J; Morris JG. | 2021 | An Outbreak of COVID-19 Among H-2A Temporary Agricultural Workers | USA | HIC | May 2020 to June 2020 | Migrant workers | risk of infection | Qualitative research | The COVID-19 outbreak described here highlights the ease with which SARS-CoV-2 can spread through this population when workers are housed and transported in crowded conditions with minimal medical oversight (...) In this instance, two previously positive workers were not identified until after they had exposed the larger crew, no one was responsible for ensuring that exposed workers were separated or isolated during transportation or at their housing site, and public health officials' recommendations to keep the ill and exposed crew members from working or moving to their next worksite or to notify individuals at that site were not heeded. | Serious consideration should be given at the local, state, and national levels to the implementation of public health strategies to protect H-2A and other agricultural workers, particularly in light of their essential role in maintaining the US food supply. | low |
| Lehmann S; Skogen JC; Haug E et al. | 2021 | Perceived consequences and worries among youth in Norway during the COVID-19 pandemic lockdown | Norway | HIC | April 27, 2020 to May 11, 2020 | International migrants | mental health | Cohort Study | Overall, 28% reported feeling somewhat to a lot impacted by schools closing, 63% reported learning less. In total, 62% reported improvement of everyday life. The youth’s situation in their family was worse for 13%. Regarding sleep problems, 19% reported difficulties initiating and maintaining sleep, 12% had more nightmares, while 90% reported later bedtime and rise time. Seven per cent worried about getting infected, while 53% worried about infection among family members. | The perceived consequences and degree of worries varied by age, gender, socioeconomic status and to a certain degree country of birth. Girls, older youth, youth with lower socioeconomic status and with a migrant background from developing countries seemed to experience the lockdown as more difficult, and thereby possibly accentuating the need for services in these groups. | medium |
| Li T; Li Z; Pan Y; Wang X. | 2021 | Frangibility and potentiality: migrant worker families in China during COVID-19 | China | UMIC | January 2020 to July 2020 | Internal migrants | effect of lockdown measures; mental health | Mixed-Methods: Cross-Sectional Study; Qualitative Research | The research revealed three distinct long-term vulnerabilities faced by migrant worker families experiencing difficulties: physical/material, motivational/ attitudinal and social/organisational. At the same time, during our project helping migrant workers fight COVID-19, we recognised these families' capacities. | Thus, we believe that sustainable and systematic intervention is needed to establish a long-term service mechanism for migrant workers' disaster management, and improve social security policies, provision and regulations. This will promote disaster management awareness and behaviour change among migrant worker families, improve the flexibility and quality of social services, effectively reduce the | medium |

| | | | | | | | | | | | |
|---|---|---|---|---|---|---|---|---|---|---|---|
| | | | | | | | | | | vulnerabilities of migrant worker families and enhance their capacities. | |
| Lin Y; Zhong P; Chen T. | 2020 | Association Between Socioeconomic Factors and the COVID-19 Outbreak in the 39 Well-Developed Cities of China | China | UMIC | March 2020 | Internal migrants | risk of infection; transmission | Cross-Sectional Study | Results of multiple linear regression showed a statistically significant association between the number of cases and the number of travelers from Wuhan (t = 6.746, P = 0.000) and the number of rural-to-urban migrants (t = 3.776, P = 0.001) in these cities. However, other seven potential contributors were not associated with the number of cases. | In conclusion, travelers from Wuhan and rural-to-urban migrants were independently associated with the COVID-19 outbreak in the 39 well-developed cities of China. Thus, travelers from an epicenter and rural-to-urban migrants should be paid more attention in the early stage of the COVID-19 outbreak in the well-developed cities. | medium |
| Logie C. H.; Berry I.; Okumu M. et al. | 2021 | The prevalence and correlates of depression before and after the COVID-19 pandemic declaration among urban refugee adolescents and youth in informal settlements in Kampala, Uganda: A longitudinal cohort study | Uganda | LMIC | February 2020 to December 2020 | Refugees | mental health | Cohort Study | The prevalence of depression was high, but there was no significant difference before (27.5%), and after (28.9%) the pandemic declaration (P = .583). In adjusted models, food insecurity (aOR: 2.54; 95% CI: 1.21-5.33) and experiencing violence (aOR: 2.53; 95% CI: 1.07-5.96) were associated with increased depression, and social support was associated with decreased depression (aOR: 0.85; 95% CI: 0.81-0.89). | Although depression scores and severity did not significantly change before and after the pandemic declaration, this work signals a need for urgent attention to chronic levels of depression, food insecurity, and the ongoing mental health effects of violence exposure among urban refugee adolescents and youth in Kampala's informal settlements. | high |
| Lombardi A; Mangioni D; Consonni D et al. | 2021 | Seroprevalence of anti-SARS-CoV-2 IgG among healthcare workers of a large university hospital in Milan, Lombardy, Italy: a cross-sectional study | Italy | HIC | April 7, 2020 to June 12, 2020 | International migrants | health outcomes of disease: SARS-Cov-2 cases (risk of infection) | Cross-Sectional Study | The highest rates of seroprevalence were detected among foreign-born workers [14.5%; OR (ref. Italian-born) = 1.82 (1.07 to 3.06)] [...]. SARS-CoV-2 sero-prevalence of frontline HCWs did not differ from those who did not report direct contact with patients with COVID-19. | What is suggested by our study, and by those similarly performed in the same area in the context of the ongoing pandemic, is that the observed seroprevalence rate reflects the spread of infection in the community served by the hospital. Assuming that PPE is provided and correctly employed by all HCWs, hospitals do not seem to act as an epicentre of the infection. | high |
| Lopez-Mendoza H; Montanes A; Moliner-Lahoz FJ. | 2021 | Disparities in the Evolution of the COVID-19 Pandemic between Spanish Provinces | Spain | HIC | June 2020 to November 2020 | International migrants | risk of infection | Cohort Study | Provinces with developed agricultural and industrial economic sectors, high mobility, and a high proportion of Central and South American immigrants had the highest level of CI14 [cumulative incidence for 14 days]. We show that the transmission of the virus is not homogeneous in the Spanish national territory. | Our results show that mobility, economic structure, migration and overall provincial health status are strongly associated with COVID-19 outcomes. However, we recognizethat additional studies are necessary to study in detail the causality of the variables that we have included in our model. | medium |
| Lopez-Pena P, Austin Davis C, Mushfiq Mobarak | 2020 | Prevalence of COVID-19 symptoms, risk factors, and health behaviors in host | Bangladesh | LIC | April 2020 | Refugees | health outcomes of disease; (risk of | Cross-Sectional Study | Camp residents report COVID-19 symptoms almost twice as frequently a s members of the host community. We also document differences in self- | COVID-19 symptoms are highly prevalent in Cox's Bazar, especially in refugee camps. Attendance at religious and social events threatens efforts to contain the spread of the disease. Pharmacies and religious leaders are | low |

| | | | | | | | | | | | |
|---|---|---|---|---|---|---|---|---|---|---|---|
| A; Raihan S. | | and refugee communities in Cox's Bazar: A representative panel study | | | | | transmission) | | reported non-COVID-19 symptoms, but these are not statistically significant. | promising outlets to disseminate life-saving information. | |
| Lui IF.; Vandan N.; Davies SE. et al. | 2021 | We also deserve help during the pandemic: The effect of the COVID-19 pandemic on foreign domestic workers in Hong Kong | Hong Kong, China | HIC | May 2020 to August 2020 | Migrant workers | effects of lockdown measures | Qualitative research | FDWs [foreign domestic workers] reported a dual-country experience of the pandemic, where they expressed concerns about local transmission risks as well as worries about their family members in their home country. Changes to their current work situation included how their employers treated them, as well as their employment status. FDWs also cited blind spots in the Hong Kong policy response that also affected their experience of the pandemic, including a lack of support from the Hong Kong government. | The current study demonstrates that the COVID-19 pandemic has exacerbated the existing power dynamics that constrain FDWs in Hong Kong. Not only is there a greater need to provide for family members back in FDWs' home countries, but there are also increased pressures from employers and a lack of support from the Hong Kong government. Through this study's findings of the current situation and challenges faced by FDWs in Hong Kong during the COVID-19 pandemic, it is clear that policy-level interventions are needed to mitigate the particularly negative effects on FDWs. More supportive policies should be adopted that not only consider the specific needs of FDWs but listens to them. | high |
| Lusambili A; Martini M; Abdirahman F; Asante A; Ochieng S; N Guni; Maina R; Luchters S. | 2020 | "We have a lot of home deliveries:" A qualitative study on the impact of COVID-19 on access to and utilization of reproductive, maternal, newborn and child health care among refugee women in urban Eastleigh, Kenya | Kenya | LMIC | October 2020 | Refugees | Health services access | Qualitative research | Our findings suggest that within the first eight months of COVID-19, preferences for home deliveries by refugee women increased and health care workers reported having observed reduced utilisation of services and delayed care. Fear, economic challenges and lack of migrant-inclusive health system policies were key factors influencing home deliveries and delayed and low uptake of facility-based care. | Findings from this study show the need for refugee inclusive public health policies, particularly during a pandemic, and the need to tailor health care services for refugees at facilities and in communities. The findings also make clear the urgent need to mitigate and lower barriers that prevent refugee women from seeking care at health facilities. Lastly there is need for methodological design tailored towards refugees' unique needs. | medium |
| Lusk JL; Chandra R. | 2021 | Farmer and farm worker illnesses and deaths from COVID-19 and impacts on agricultural output | USA | HIC | March 1, 2020 to March 31, 2021 | Migrant workers | health outcomes of disease: Mortality (risk of infection) | Ecological study | In the 13-month period since the start of the pandemic (from March 1, 2020 to March 31, 2021), the estimated cumulative number of COVID-19 cases (deaths) was 329,031 (6,166) among agricultural producers, 170,137 (2,969) among hired agricultural workers, 202,902 (3,812) among unpaid agricultural workers, and 27,223 (459) among migrant agricultural workers. The cases amount to 9.55%, 9.31%, 9.39%, and 9.01% of all U.S. agricultural producers, hired workers, | Results suggest counties that employ more agricultural workers, particularly hired and migrant workers, are at greater risk for COVID-19, findings which suggest these groups are at heightened risk from COVID-19. In addition to the disease risks, hired and migrant agricultural workers represent populations that tend to have toward lower incomes, greater job insecurity, and more perilous immigration and legal status than the general population, which suggest additional relative financial risks resulting | medium |

| | | | | | | | | | | | |
|---|---|---|---|---|---|---|---|---|---|---|---|
| | | | | | | | | | unpaid workers, and migrant workers, respectively. | from the burden of medical costs or lost time away from work. Beyond the immediate impact on agricultural workers, this research also shows that reductions in agricultural labor have the potential to adversely affect food supply. | |
| Ly TDA; Nguyen NN; Hoang VT et al. | 2021 | Screening of SARS-CoV-2 among homeless people, asylum-seekers and other people living in precarious conditions in Marseille, France, March-April 2020 | France | HIC | March 26, 2020 to April 17, 2020 | Asylum Seekers | risk of infection | Cross-Sectional Study | Overall, 49 people (7.0%) were positive for SARS-CoV-2, including 37 homeless individuals (of 411, 9.0%) and 12 employees (of 152, 7.9%). SARS-CoV-2 positivity correlated with symptoms, although 51% of patients who tested positive did not report respiratory symptoms or fever. Among homeless people, being young (18-34 years) (odds ratio 3.83, 95% confidence interval 1.47-10.0, p = 0.006) and being housed in one specific shelter (odds ratio 9.13, 95% confidence interval 4.09-20.37, p < 0.001) were independent factors associated with SARS-CoV-2 positivity (rates of 11.4% and 20.6%, respectively). | Symptom screening alone is insufficient to prevent SARS-CoV-2 transmission in vulnerable sheltered people. Systematic testing should be promoted. | medium |
| Maeland S; Bjorknes R; Lehmahn S et al. | 2021 | How the Norwegian population was affected by non-pharmaceutical interventions during the first six weeks of the COVID-19 lockdown | Norway | HIC | April 2020 | International migrants | health services access (?) effect of lockdown measures (?) | Cross-Sectional Study | An important finding is that immigrants were more prone to have a reduction in follow-up, particularly from psychology and physiotherapy services. | The non-pharmaceutical interventions were perceived as manageable by the majority of the adult general population in Norway at the beginning of the COVID-19 pandemic. A substantial proportion of adults <30 years old experienced difficulties with social distancing, and those >70 years old lacked the digital tools and knowledge. Further, immigrant access to health services needs monitoring and future attention. | low |
| Mallet Y; Pivette M; Revest M et al. | 2021 | Identification of Workers at Increased Risk of Infection During a COVID-19 Outbreak in a Meat Processing Plant, France, May 2020 | France | HIC | May 2020 | International migrants | health outcomes of disease: SARS-CoV-2 Cases (risk of infection/transmission) | Cross-Sectional Study | Of the 1347 workers, 87.5% were tested: 140 cases were identified; 4 were hospitalised, including 2 admitted to intensive care. In the company, the cluster remained limited to deboning and cutting activities. The attack rate was 11.9% in the company, reaching 16.6% in the cutting department. (…) In the cutting department, an association with virus infection was found for a group of non-French speaking workers from the same Eastern European country (RR=2.67 [1.76–4.05]). They shared accommodation or carpooled more frequently than the other cases. | In conclusion, the investigation of the outbreak revealed a significantly increased risk of SARS-CoV-2 infection for workers of subcontractors and some foreign-born workers. (…) Particular attention must now be paid to their ability to appropriate and implement the COVID-19 risk manage-ment measures in terms of vaccination, early detection of symptoms, barrier gestures and distancing measures. Information and prevention messages must therefore be translated into the language of the workers. Messages must take into account the risks of contamination in the workplace, as well as in peri-professional and private life. | high |

| | | | | | | | | | | | |
|---|---|---|---|---|---|---|---|---|---|---|---|
| Marco A; Teixido N; Guerrero RA et al. | 2021 | Outbreak of SARS-CoV-2 in a prison: Low effectiveness of a single dose of the adenovirus vector ChAdOx1 vaccine in recently vaccinated inmates | Spain | HIC | 28 days (not further specified ) | Internatio nal migrants | health outcomes of disease: SARS-CoV-2 Cases; vaccination | Cross-Sectional Study | Forty-eight (25.9%) infections by the SCV-B.1.17 variant were recorded in prisoners and none in staff. Infection rates were higher in younger prisoners, immigrants, and those admitted ≥7 days previously. In all, 22.6% of vaccinated subjects were infected vs. 29.3% of unvaccinated. Vaccine effectiveness was 23%. | In a prison outbreak, a dose of AVChAdOx1 administered 21-23 days earlier did not significantly prevent the occurrence of infections, but did reduce the duration of rt-PCR positivity. | medium |
| Martin-Anatias N; Long N; Davies SG et al. | 2021 | Lockdown ibuism: Experiences of Indonesian migrant mothers during the COVID-19 pandemic in aotearoa New Zealand | New Zealand | HIC | May 2020 to June 2020 | Internatio nal migrants | effect of lockdown measures | Mixed-Methods: Cross-Sectional Study; Qualitati ve Research | Both our survey and our interviews indicated that many women, particularly mothers, experienced increased demands on their time and emotional energy in comparison to men, particularly in fields related to childcare and domestic labour, and that they also experienced a decline in wellbeing linked to heightened levels of exhaustion and stress. (…) Their [Indonesian migrant mothers] stresses were instead linked to feelings of anxiety, frustrations and inadequacy with homeschooling their children in English (…) our research indicates that both second-language speaking status and a prevailing gender ideology of 'state Ibuism' (as opposed to neoliberal feminism) were critical factors differentiating Indonesian migrant mothers' experiences from those of many other mothers in NZ. | Our research corroborates prevailing conclusions offered by existing work on gender relations during the 2020 coronavirus pandemic in NZ and beyond. (…) While feminist scholars are certainly correct to highlight the pervasiveness of gendered inequality during lockdown, an attention to distinct meanings of womanhood and motherhood afford a more nuanced approach, illuminating how lockdowns can offer not only gendered challenges but also gendered rewards, and cast light on measures that could be taken in future to enhance the experience of living through a lockdown in one's intersectional specificity. | medium |
| Martin-Sanchez FJ; Carbo AV; Miro O et al. | 2021 | Socio-Demographic Health Determinants Are Associated with Poor Prognosis in Spanish Patients Hospitalized with COVID-19 | Spain | HIC | March 1, 2020 to April 15, 2020 | Internatio nal migrants | health outcomes of disease: ICU admission, Mortality | Cohort Study | We included 10,110 COVID-19 patients admitted to 18 Spanish hospitals (median age 68 (IQR 54-80) years old; 44.5% female; 14.8% were not born in Spain). Among these, 779 (7.7%) cases were admitted to critical care units and 1678 (16.6%) patients died during the hospitalization. | Social determinants such as residence in low-income areas and being born in Latin American countries were associated with increased odds of being admitted to an intensive care unit and of in-hospital mortality. There was considerable variation in outcomes between different Spanish centers. | high |
| Martuscelli PN. | 2021 | How are forcibly displaced people affected by the COVID-19 pandemic outbreak? Evidence from Brazil | Brazil | UMIC | March 27, 2020 to April 06, 2020 | Internatio nal migrants | effect of lockdown measures | Qualitati ve research | The results indicate that refugees face three challenges connected to this pandemic: (a) same challenges as Brazilians due to their labor vulnerability social identity, (b) challenges aggravated by the pandemic due to their identity of nonnationals including access to information and services, and (c) new challenges due to their social identity of | This study reinforces the need to understand how forcibly displaced people in the Global South are affected by pandemics and responses to them adopting an intersectionality approach, especially in situations that are not refugee camps, like Brazil. Granting refugees the possibility to reflect on their experiences provides valuable insight on how to involve and | high |

| | | | | | | | | | | | |
|---|---|---|---|---|---|---|---|---|---|---|---|
| | | | | | | | | | forced displaced nonnationals including closing of migration services and borders and the feeling of “living the pandemic twice.” | engage them in responses to global health emergencies. | |
| Masai A. N.; Gueciz-Dogan B.; Ouma P. N.; Nyadera I. N.; Ruto V. K. | 2021 | Healthcare services utilization among international students in Ankara, Turkey: a cross-sectional study | Turkey | UMIC | September 2020 to October 2020 | International students | health services access | Cross-Sectional Study | Of the study population, 80.6% accessed the general practitioner (GP), 40% accessed the student health centres, and 11.4% were admitted to the hospital at least once. About 80% of international students reported changing their views to access healthcare more because of the COVID-19 pandemic. | According to the survey results, the lack of awareness, language barrier, and fear caused by perceived stigma hinder these students from fully accessing health services. This may be disastrous in case a student with an infectious disease does not seek medical attention and ends up infecting other people. It can also be catastrophic on the patient if at all the disease could have been treatable during the early stages but they end up not seeking treatment. Therefore, the data obtained show the need for and will inform the provision of targeted health services to international students in Turkey. | high |
| Mema S; Frosst G; Hanson K et al. | 2021 | COVID-19 outbreak among temporary foreign workers in British Columbia, March to May 2020 | Canada | HIC | March 28, 2020 to May 10, 2020 | Migrant workers | health outcomes of disease: SARS-CoV-2 cases | Cohort Study | A total of 26 COVID-19 cases were identified among the group of TFWs [temporary foreign workers]; no cases were identified among local workers. Cases were primarily male (77%) with a median age of 41 years. Symptom onsets ranged from March 8 to April 9, 2020. One case required overnight hospitalization for pneumonia. | TFWs have had unique risks during the COVID-19 pandemic as demonstrated by this outbreak, which occurred in an agricultural setting in BC. (…) Immediate quarantine of affected workers, comprehensive follow-up of cases and contacts, and mobilization of an outreach team were effective strategies to manage and control this outbreak. (…) Provincial and federal orders and guidance have since been developed to reduce outbreak risk in agricultural settings and to protect the health and safety of both workers and Canadians in the context of the pandemic. | medium |
| Mendez-Dominguez N; Alvarez-Baeza A; Carrillo G. | 2020 | Demographic and Health Indicators in Correlation to Interstate Variability of Incidence, Confirmation, Hospitalization, and Lethality in Mexico: Preliminary Analysis from Imported and Community Acquired Cases during COVID-19 Outbreak | Mexico | UMIC | Februrary 28, 2020 to April 21, 2020 | Internal and International migrants | risk of infection; health outcomes of disease | Cross-Sectional Study | The national incidence was 13.89/100,000 inhabitants with a 6.52% overall lethality and a confirmed-case mortality of 11.1%. The incidence variation significantly correlated with migration, but not urbanization. (…) State lethality positively correlated with the proportion of the population assisted at public hospitals and correlated inversely to the number of hospitals and clinics in the state. | Migration strongly correlated with incidence; elderly patients had lower odds of being hospitalized but were likely to die. Patients aged <15 were less prone to be laboratory-confirmed. Case confirmation was not performed in all hospitalized patients, but 72.15% of hospitalized patients had favorable outcomes to date. | high |

| | | | | | | | | | | | |
|---|---|---|---|---|---|---|---|---|---|---|---|
| Methi F; Hart RK; Godoy AA; Jorgensen SB; Kacelnik O; Telle KE. | 2021 | Transmission of SARS-CoV-2 into and within immigrant households. Nation-wide registry-study from Norway | Norway | HIC | August 1, 2020 to May 1, 2021 | International migrants | health outcomes of disease: SARS-CoV-2 cases, Hospitalisation, Mortality (risk of transmission) | Cohort Study | Among all multiperson households in Norway (n=1 422 411), at least one member had been infected in 3.7% of the 343 017 immigrant households and 1.4% in the 1 079 394 households with only Norwegian-born members. (…) The share of households with at least one member hospitalised or dead from COVID-19 was higher in immigrant (0.4% households with hospitalised member, 0.02% households with dead member) than Norwegian-born households (0.1 %, 0.01%). Households from West Asia were among the most severely hit, with 1.1% having at least one member hospitalised and 0.06% having at least one dead member. | SARS-CoV-2 is more frequently introduced into multiperson immigrant households than into households with only Norwegian-born members, and transmission within the household occurs more frequently in immigrant households. The results are likely related to living conditions, family composition or differences in social interaction, emphasising the need to prevent introduction of SARS-CoV-2 into these vulnerable households. | high |
| Miconi D; Li ZY; Frounfelker RL et al. | 2021 | Ethno-cultural disparities in mental health during the COVID-19 pandemic: a cross-sectional study on the impact of exposure to the virus and COVID-19-related discrimination and stigma on mental health across ethno-cultural groups in Quebec (Canada) | Canada | HIC | June 1, 2020 to June 23, 2010 | International migrants | mental health | Cross-Sectional Study | Mental health varied significantly based on socioeconomic status and ethno-cultural group, with those with lower incomes and Arab participants reporting higher psychological distress. Exposure to the virus, COVID-19-related discrimination, and stigma were associated with poorer mental health. Associations with mental health varied across ethno-cultural groups, with exposed and discriminated Black participants reporting higher mental distress. | Findings indicate sociocultural inequalities in mental health related to COVID-19 in the Canadian context. COVID-19-related risk factors, including exposure, discrimination and stigma, jeopardise mental health. This burden is most noteworthy for the Black community. There is an urgent need for public health authorities and health professionals to advocate against the discrimination of racialised minorities, and ensure that mental health services are accessible and culturally sensitive during and,in the aftermath of the pandemic. | high |
| Miconi D; Li ZY; Frounfelker RL; Venkatesh V; Rousseau C. | 2021 | Socio-cultural correlates of self-reported experiences of discrimination related to COVID-19 in a culturally diverse sample of Canadian adults | Canada | HIC | June 1, 2020 to June 23, 2020 | International migrants | effect of lockdown measures | Cross-Sectional Study | COVID-related discrimination was reported by 16.58 % of participants. Non-white participants, health-care workers and younger participants were more likely to experience discrimination than white, unemployed and older participants, respectively. Discrimination was reported primarily in association with participants' ethno-cultural group, age, occupation and physical health and in the context of public spaces. | Our findings indicate that health communication actions informed by a social pedagogy approach should target public beliefs related to the association of COVID-19 with ethnicity, age and occupation, to minimize pandemic-related discrimination. Visible minorities, health-care workers and seniors should be protected and supported, especially in public spaces. | high |
| Ming X.; De Jong M. D. T. | 2021 | Mental Well-Being of Chinese Immigrants in the Netherlands during | Netherlands | HIC | November 24, 2020 to December | International migrants | mental health effects of | Cross-Sectional Study | Results show that participants judged the Chinese COVID-19 situation significantly more positively than the Dutch situation. Five antecedents of decreased mental | The results call for a broad perspective on factors associated with mental well-being and for special attention for minority groups | medium |

| | | | | | | | | | | | |
|---|---|---|---|---|---|---|---|---|---|---|---|
| | | the COVID-19 Pandemic: A Survey Investigating Personal and Societal Antecedents | | | r 12, 2020 | | lockdown measures | | well-being were found: financial concerns, social isolation, feelings of lost time, experienced racism, and distrust of Dutch COVID-19 information and figures. | in the societal dynamics. | |
| Mishra S; Ma H; Moloney G et al. | 2021 | Increasing concentration of COVID-19 by socioeconomic determinants and geography in Toronto, Canada: an observational study | Canada | HIC | January 2020 to November 2020 | International migrants (%) | risk of transmission | Cross-Sectional Study | Gini coefficient of cumulative cases by population size was 0.41 (95% confidence interval [CI]:0.36-0.47) and estimated for: household income (0.20, 95%CI: 0.14-0.28); visible minority (0.21, 95%CI:0.16-0.28); recent immigration (0.12, 95%CI:0.09-0.16); suitable housing (0.21, 95%CI:0.14-0.30); multigenerational households (0.19, 95%CI:0.15-0.23); and essential workers (0.28, 95%CI:0.23-0.34). | In sum, there was a rapid epidemiologic transition in COVID-19 from an epidemic concentrated among higher-income communities in Toronto related to travel to local transmission in lower-income communities secondary to structural risks including structural racism often defining where people work and how they live. Similar to other infectious diseases, the findings of marked inequities in COVID-19 burden in Toronto presented here suggests the existence of core-groups concentrated by SDOH. | high |
| Moenkemoeller K; Kamrath C; Hammersen J et al. | 2021 | [Is it possible to prevent diabetic ketoacidosis at diagnosis of pediatric type-1 diabetes? Lessons from the COVID-19 pandemic] | Germany | HIC | March 3, 2020 to May 13, 2020 | International migrants | health outcomes of disease | Cross-Sectional Study | Children <6 years had the highest risk with an 141.6% increase for DKA and 97.0% for severe DKA compared tothe previous years. Migration background was a risk factor independent of COVID-19.Of the patients 31% had either a delayed presentation or a missed diagnosis. | During the COVID-19 lockdownthe frequency of DKA and severe DKA at DM1 onset was significantly increased for childrenand adolescents in Germany. Age <6 years, migration background and delayed diagnosis were the main risk factors. | high |
| Mookerjee D.; Chakravarty S.; Roy S.; Tagat A.; Mukherjee S. | 2021 | A culture-centered approach to experiences of the coronavirus pandemic lockdown among internal migrants in India. (Special Issue: The COVID-19 pandemic: precarious migrants and outbreak inequality.) | India | LMIC | 3 months into the lockdown (presumably: May 2020 to July 2020) | Internal migrants (%) | effect of lockdown measures | Qualitative Research | Financial worries were found to be endemic, with rising debt a major source of stress, and educational qualifications becoming an obstacle to earning. Returning migrants were suspected of bringing the virus from the city, and so stigmatized in their home towns and villages. However, the pandemic lockdown also showed some unexpected healthful consequences. It provided these marginalized, and always busy workers the time and space to stop working for a while, to stay home, eat home food, and take walks in the comparatively green and clean spaces of their home environments. | Any future policy about migrants must prioritize addressing the extreme financial precarity of internal migrants. This study shows the need for a kind of social security or insurance to get workers though low-or-no-income periods, but further research focused on the economic structural constraints experienced by migrant workers is necessary to shape such policy. (…) The pandemic lockdown as an enabler of health is one of three inversions that emerged in this culture-centered study (…) In this, the pandemic lockdown may be seen to have enabled a measure of agency and health in the lives of these workers, an oasis albeit temporary, and ultimately subject to the demands of the globalized cities of India. | high |
| Moran JK.; Bretz J; Winkler J; | 2021 | The Differential Impact of Lockdown | Germany | HIC | April 1, 2020 to | International migrants | mental health | Cross-Sectional Study | In the migration background group, we found that women had more lockdown-related psychological distress than men. | Our results support earlier findings on differential vulnerability of diagnostic groups to these stressors. Thus, clinicians | high |

| | | | | | | | | | | | |
|---|---|---|---|---|---|---|---|---|---|---|---|
| Gutwinski S; Brandl EJ; Schouler-Ocak M. | | Measures Upon Migrant and Female Psychiatric Patients - A Cross-Sectional Survey in a Psychiatric Hospital in Berlin, Germany | | | June 30, 2020 | | | | This was not apparent in those patients with a German background. We found that females were more strongly affected by family-related distress, particularly those with a migration background. | can optimize treatment by taking family-related stressors into account particularly for females and people with a migrant background. | |
| Motta I; Centis R; D'Ambrosio et al. | 2020 | Tuberculosis, COVID-19 and migrants: preliminary analysis of deaths occurring in 69 patients from two cohorts | Belgium, Brazil, France, Italy, Russia, Singapore, Spain, Switzerland | HIC | March 12, 2020 to May 5, 2020 | International migrants | health outcomes of disease | Cohort Study | To the best of our knowledge this is the first report of patients dying with TB and COVID-19, including 69 patients from the two largest cohorts of co-infected patients available so far. Although the case-fatality rate was rather high (overall 10.6%, but 14.3% in the first cohort) and still preliminary (it can increase over time within both cohorts), the results seem consistent with those observed in other cohorts of COVID-19 patients. In general, all patients (except one) were aged >65 years, and were affected by >2 comorbidities. In all cases COVID-19 contributed to worsen the prognosis of TB patients and/or to cause death. | he study findings show that: (1) mortality is likely to occur in elderly patients with comorbidities; (2) TB might not be a major determinant of mortality and (3) migrants had lower mortality, probably because of their younger age and lower number of co-morbidities. However, in settings where advanced forms of TB frequently occur and are caused by drug-resistant strains of M. tuberculosis, higher mortality rates can be expected in young individuals. | high |
| Moya A; Serneels P; Desrosiers A; Reyes V; Torres MJ; Lieberman A. | 2021 | The COVID-19 pandemic and maternal mental health in a fragile and conflict-affected setting in Tumaco, Colombia: a cohort study | Colombia | UMIC | 2018 to 2020 | IDP | mental health | Cohort Study | Results showed that the likelihood of reporting symptoms above the risk threshold increased by 14 percentage points for anxiety (95% CI 10-17), 5 percentage points for depression (0.5-9), and 10 percentage points for parental stress (5-15). The deterioration in mental health was stronger for IDP, participants with lower education or pre-existing mental health conditions, and for those reporting a higher number of stressors, including food insecurity and job loss. | Maternal mental health significantly worsened during the early stages of the pandemic. Considering the vulnerability and pre-existing mental health conditions of this population, the estimated effects are substantial. Policies in fragile and conflict-affected settings targeting IDP and other vulnerable people will be important to mitigate further mental health and socioeconomic problems. | high |
| Nair SC; Gasmelseed HI; Khan AA et al. | 2021 | Assessment of mortality from COVID-19 in a multicultural multi-ethnic patient population | United Arab Emirates | HIC | March 1, 2020 to June 30, 2020 | International migrants | health outcomes of disease: Mortality | Cross-Sectional Study | From, the total of 3072 patients, less than one-fifth were females; the Asian population (71.2%); followed by Middle Eastern Arabs (23.3%) were the most infected by the virus. (…) Kidney disease as comorbidity significantly diminished the survival rates (…) Similarly, the higher age of patients between 51 and 65 years, significantly decreased the odds for survival (…) (Adjusted OR 12.3 95% CI (2.9 – 52.4), p<0.001). | Our study indicates that older ages above 51 years and kidney disease increased mortality significantly in COVID-19 patients. Ethnicity was not significantly associated with mortality in the UAE population. Our findings are important in the management of the COVID-19 disease in the region with similar economic, social, cultural, and ethnic backgrounds. | high |

| | | | | | | | | | | | |
|---|---|---|---|---|---|---|---|---|---|---|---|
| Nasol K; Francisco-Menchavez V. | 2021 | Filipino home care workers: invisible frontline workers in the COVID-19 crisis in the United States | USA | HIC | 2017 to 2020 (not further specified ) | Migrant workers | effect of lockdown measures; health services access | Qualitative research | Based on quantitative and qualitative data with Filipino workers before and during the COVID-19 crisis, we find that RCFEs have failed to comply with labor standards long before the pandemic where the lack of state regulation denied health and safety protections for home care workers. The racial inequities under COVID-19 via the neoliberal approach to the crisis puts home care workers at more risk. (…) Last, while the experiences of Filipino home care workers during the pandemic expose the elder care industry's exploitation, we find that they are also creating strategies to take care of one another. | Our study demonstrates how legal protections and public assistance, or the lack thereof, is a major social determinants of care workers' health. The experiences of Filipino home care workers during the COVID-19 pandemic broadly expose the elder care industry's exploitation of racialized migrant workers and the government's neglect in protecting them, endangering key essential workers during a historical public health crisis. And yet, we have found that Filipino care workers are crafting "communities of care" that circulate different forms and ethics of care among themselves. (…) Although these informal networks are not a sustainable replacement for state-sponsored health care and health resources, we uplift the innovative strategies of Filipino care workers to demonstrate their ingenuity and commitment to caring for themselves and one another. | medium |
| Naumann E; von den Driesch E; Schumann A; Thönnissen C. | 2021 | [Increase of depressive symptoms among adolescents during the first COVID-19 lockdown in Germany : Results from the German family panel pairfam] | Germany | HIC | May 2020 to July 2020 | International migrants | mental health | Cohort Study | During the first lockdown, adolescents show a significant increase indepressive symptoms. Prior to the lockdown, 10.4% had clinically relevant depressive symptoms [95% CI: 8.4; 12.5]. In spring 2020, the prevalence increased to 25.3%[95% CI: 22.4; 28.2]. Young women havea significantly higher risk of developing depressive symptoms than men of the sameage. Immigrant background is an equally strong risk factor. The prevalence of depressive symptoms among adolescents with an immigrant background increased from 11% to 33%. | To address this increased mental health risk and the inequalities, policy makers and society should ensure access and availability of target-group-specific and low-threshold prevention and counselling. | high |
| Ngiam JN; Chew N; Tham SM et al. | 2021 | Elevated liver enzymes in hospitalized patients with COVID-19 in Singapore | Singapore | HIC | February 2020 to April 2020 | Migrant workers | health outcomes of disease | Cohort Study | Multivariable analysis showed that being a migrant worker from a dormitory was an independent risk factor (adjusted OR 2.11, 95% CI 1.10-4.00,P=.024) for liver enzyme elevated levels on admission. This young and otherwise well cohort (largely asymptomatic or withmild COVID-19 illness) were also surprisingly more likely to have persistently abnormal LFTs at time of discharge (88.9% vs16.6%,P=.009). | Despite relatively mild COVID-19 disease, there was a significant prevalence of liver dysfunction, particularly amongst migrant workers. Elevated liver enzymes were associated with more severe disease, despite similar haemodynamic characteristics. Future studies should explore whether pre-existing liver disease may predispose to more severe COVID-19 disease. | high |

| | | | | | | | | | | | |
|---|---|---|---|---|---|---|---|---|---|---|---|
| Ngiam JN; Chew N; Tham SM et al. | 2020 | Demographic shift in COVID-19 patients in Singapore from an aged, at-risk population to young migrant workers with reduced risk of severe disease | Singapore | HIC | February 2020 to April 2020 | Migrant workers | health outcomes of disease: SARS-CoV-2 Cases, ICU-admission, Mortality | Cross-Sectional Study | Local cases acquired outside the dormitories were older compared with imported or migrant worker cases median age in years: 46±16 vs 39±15 vs 35±9, respectively; p < 0.001). (…) A total of 49% and 57% of local non-dormitory and imported cases were of Chinese ethnicity, respectively, and 77% of migrant workers were Indian or Bangladeshi. (…) There were two deaths, both in local non-dormitory cases (…). | Singapore experienced a substantial shift in the population at risk of severe COVID-19. Successful control in the community protected an aging population. Large migrant worker dormitory outbreaks occurred, but the disease incurred was less severe, resulting in Singapore having one of the lowest case fatality rates in the world. | high |
| Norman F; Crespillo C; Perez J et al. | 2020 | COVID-19 and geographical area of origin | Spain | HIC | March 2020 to May 2020 | International migrants | health outcomes of disease: ICU-admission, Mortality | Cross-Sectional Study | In-hospital mortality overall was higher in Europeans (443/1956, 22.7%) than in non-Europeans (40/389,10.3%) (p<0.001), but there were no significant differences when adjusted for age/gender (OR 1.27, 95%CI 0.86e1.88). Non-Europeans were more frequently admitted to ICU (71/389, 18.3%) compared with Europeans (187/1956, 9.6%) (p<0.001) and a difference in ICU admission rate was also found when adjusted for age/ gender (OR 1.43, 95% CI 1.03e1.98). | No significant differences regarding in-hospital mortality (when adjusted for age and gender) were observed between Europeans and non-Europeans (mainly Latin Americans), although an increased rate of ICU admission in non-Europeans was found. Differences between ethnicity and COVID-19 outcome found in other studies may be the result of a higher burden of adverse socio-economic factors in certain populations. Investigation of genetic and immunological markers in patients of different geographical origin should aid the understanding of determinants of outcome in COVID-19. | high |
| Ofuoku AU; Opia BOP; Ikpoza EA. | 2021 | Impact of COVID-19-induced rural-rural migration on agricultural productivity in Delta State, Nigeria | Nigeria | LMIC | not reported | International migrants (%) | effect of lockdown measures; risk of transmission | Cross-Sectional Study | In sending communities, 41% and 65% of the migrant and indigenous populations, respectively, contacted the disease, while there was no recorded case in receiving communities. The mean yield in receiving communities was higher than that in the sending communities. COVID-19 incidents and other factors influenced immigration to receiving communities. Migration increased agricultural productivity in receiving communities, while productivity was reduced in sending communities. | These findings have some important implications. For instance, the use of state and local security formations in large rural communities to enforce the COVID-19 lockdown did not put into consideration the fact that farm families naturally observe social distance while carrying out farm operations. The government ought to have considered this and relax it for farm families going to attend to their farms. | high |
| Oluyomi AO; Gunter SM; Leining LM; Murray KO; Amos C. | 2021 | COVID-19 Community Incidence and Associated Neighborhood-Level Characteristics in | USA | HIC | June 23, 2020 to August 3, 2020 | International migrants (%) | risk of transmission | Ecological study | We assessed the associations between 29 neighborhood-level characteristics and COVID-19 incidence using a series of aspatial and spatial models. The variables that maintained significant and positive associations with COVID-19 incidence in our final aspatial model and later represented in a geographically weighted | The current findings could enhance decision making by local public health officials in responding to the COVID-19 pandemic. By understanding factors that drive community transmission, we can better target disease control measures. | high |

| | | | | | | | | | | | |
|---|---|---|---|---|---|---|---|---|---|---|---|
| | | Houston, Texas, USA | | | | | | | regression model were the percentage of the Black/African American population, percentage of the foreign-born population, area derivation index (ADI), percentage of households with no vehicle, and percentage of people over 65 years old inside each census tract. | | |
| Ornelas I; Tornberg-Belanger S; Balkus J et al. | 2021 | Coping With COVID-19: The Impact of the Pandemic on Latina Immigrant Women's Mental Health and Well-being | USA | HIC | June 2020 to Septemb er 2020 | Internatio nal migrants | mental health; effect of lockdown measures | Cross-Sectional Study | Very few women had been infected with COVID-19, and 23% reported having been tested. Most frequent reasons for not being tested were not knowing where to go (14%), concerns over the cost (15%), and not wanting to know if they were infected (12%). Most participants had concerns about paying for housing (76%) and food (73%). Depression and anxiety symptoms were in the moderate range. Almost all participants were practicing recommended preventive behaviors. | Our findings suggest the need for more affordable and accessible mental health services in Spanish to help Latina immigrant women cope with stress in healthy ways. There is also a need for policies to reduce the social and economic stressors that lead to mental health disparities. | medium |
| Otto WR; Grundmeie r RW; Montoya-Williams D et al. | 2021 | Association between Preferred Language and Risk of Severe Acute Respiratory Syndrome Coronavirus 2 Infection in Children in the United States | USA | HIC | March 1, 2020 to February 28, 2021 | Internatio nal migrants | health outcomes of disease: SARS-CoV-2 Cases (risk of infection) | Ecologic al Study | A total of 10,138 patients were tested for SARS-CoV-2, with 1,284 (12.7%) being positive. Test positivity was higher for Spanish-speaking families than for English-speaking families (22.5% versus 12.3%). (…) At the peak of the first wave of the pandemic, the adjusted odd ratio for test positivity was 3.76 (95% CI, 2.07-6.67) for families with a preferred language other than English compared with English-speaking families. | In conclusion, this is the first report examining the impact of family-preferred language on SARS-CoV-2 infection in children. We identified dis-proportionately elevated SARS-CoV-2 test positivity in children from families with a preferred language other than English, which is consistent with reported findings in adults in the United States and other English-speaking nations. As we continue to track and address the disproportionate impact of the SARS-CoV-2 pandemic on historically underrepresented and mar-ginalized communities across the globe, it will be important to track disparities experienced by communities who speak languages other than English. | medium |
| Pagani G; Conti F; Giacomelli A et al. | 2021 | Differences in the Prevalence of SARS-CoV-2 Infection and Access to Care between Italians and Non-Italians in a Social-Housing Neighbourhood of Milan, Italy | Italy | HIC | Decembe r 23, 2020 to February 19, 2021 | Internatio nal migrants | health outcomes of disease: SARS-CoV-2 Cases | Cross-Sectional Study | Our findings indicate that the prevalence of SARS-CoV-2 infection in the study population as a whole was 12.4%, but it was higher among non-Italians when compared to Italians (23.3% vs. 9.1%). Moreover, being non-Italian seemed to be independently associated with a higher prevalence of SARS-CoV-2, even after adjusting for other potential social determinants. | In conclusion, our findings suggest that non-Italians in Italy are more frequently infected with SARS-CoV-2 than Italians. Although a lower educational level, a lower socio-economic status, and the experience of crowded housing were each independently associated with a higher prevalence of SARS-CoV-2, it is possible that the association between non-Italians and COVID-19 may also be due to other social and cultural determinants that were not | high |

| | | | | | | | | | | | |
|---|---|---|---|---|---|---|---|---|---|---|---|
| | | | | | | | | | | appropriately investigated in our questionnaire. | |
| Pal SC; Saha A; Chowdhuri I; Roy P; Chakrabortty R; Shit M. | 2021 | Threats of unplanned movement of migrant workers for sudden spurt of COVID-19 pandemic in India | India | LMIC | July 3, 2020 to December 31, 2020 | Migrant workers | risk of transmission; health outcomes of disease | Modelling Study | Total confirmed cases and deaths were reported in the last week of April 2020 at 33,050 and 1074. Total confirmed cases and deaths increased to 182,143 and 5164, respectively. Approximately 451.15% (5.5 times) of this increase was observed earlier in the overall confirmed case. The increase in the total death rate was 380.46% (4.8 times) in the last week of May 2020. This was due to the unplanned movement of migrant workers in India. | The government should therefore implement region wise policy strategy in the various sectors to ensure that every human being has proper shelter, food, medicine and digital contact surveillance technology (Aragyau Shetu) so that the rate of decline in these states will differ in the coming days. | medium |
| Pan H; Fokkema T; Switsers L; Dury S; Hoens S; De Donder L. | 2021 | Older Chinese migrants in coronavirus pandemic: exploring risk and protective factors to increased loneliness | Belgium; Netherlands | HIC | March 2020 to April 2020 | International migrants | mental health | Cross-Sectional Study | The fndings first indicate that the coronavirus pandemic has a significant impact on older Chinese migrants' lives. One in five experienced more loneliness. Second, reduced social participation (...) and financial insecurity (...) lead to higher than pre-pandemic loneliness levels. Problem-focused coping strategies (...) and emotion-focused coping (...) were not found to protect against increased loneliness in the pandemic. | Two practical implications for loneliness interventions for older Chinese migrants are put forward. Organizing COVID-19-safe social participation activities and paying more attention to older Chinese migrants' fnancial situation can be benefcial when addressing higher levels of loneliness due to the coronavirus pandemic. | medium |
| Pascual-Garcia A; Klein JD; Villers J; Campillo-Funollet E; Sarkis C. | 2021 | Empowering the crowd: feasible strategies for epidemic management in high-density informal settlements. The case of COVID-19 in Northwest Syria | Syria | LIC | not reported | IDP | risk of transmission; health outcomes of disease | Modelling Study | All interventions significantly reduce outbreak probability and some of them reduce mortality when an outbreak does occur. Self-distancing reduces mortality by up to 35% if contacts are reduced by 50%. A reduction in mortality by up to 18% can be achieved by providing one self-isolation tent per eight people. (...). Our model predicts that a combination of all simulated interventions may reduce mortality by more than 90% and delay an outbreak's peak by almost 2 months. | Our results highlight the potential for non-medical interventions to mitigate the effects of the pandemic. Similar measures may be applicable to controlling COVID-19 in other informal settlements, particularly IDP camps in conflict regions, around the world. | medium |
| Pawar DD; Patil WD; Raut DK. | 2021 | Fractional-order mathematical model for analysing impact of quarantine on transmission of COVID-19 in india | India | LMIC | not reported | International migrants | risk of infection | Modelling Study | The estimated data of Fig. 4 gives reproduction number as $R0 = 0.6124 < 1$. The disease free equilibrium is globally asymptomatically stable. The figures also demonstrate that there is considerable downfall in the number of infected population over the period of time. In this case, we have proposed the set of parameters to decide the control strategy. Since, the reproduction number is less than a unit, here, the disease will be under | Finally, we may conclude that the present fractional-order model has given wide platform to decide the future strategy for the control of COVID-19 in India. | low |

| | | | | | | | | | | | |
|---|---|---|---|---|---|---|---|---|---|---|---|
| | | | | | | | | | control after some period. The scenario will be some what suitable to avoid the pandemic situation. | | |
| Qi J; Ma C. | 2021 | Australia's crisis responses during COVID-19: The case of international students | Australia | HIC | February 2020 to November 2020 | International students | effect of lockdown measures | Qualitative research | Our thematic analysis highlights participants' experiences and views of Australia's crisis responses in the four areas of financing, third-country transit, visas and immigration, and pandemic management. | How countries live up to their values is critical to their global reputation and soft power. Australia has a range of soft power strengths. Nevertheless, ill-considered policies have also caused much international and domestic criticism, especially in areas such as climate change, asylum seekers and foreign aid. The list has extended to include its treatment of temporary migrants including international students. Australia needs to push for a broader vision for international education that enhances its international engagement not only for economic gains, but also for effective communication of Australia's democratic and inclusive values. | high |
| Qiu J; Shen B; Zhao M; Wang Z; Xie B; Xu Y. | 2020 | A nationwide survey of psychological distress among Chinese people in the COVID-19 epidemic: implications and policy recommendations | China | UMIC | January 31, 2020 to February 10, 20202 | Migrant workers | mental health | Cross-Sectional Study | Almost 35% of the respondents experienced psychological distress (29.29% of the respondents' scores were between 28 and 51, and 5.14% of the respondents' scores were ≥52). Multinomial logistic regression analyses showed that one's CPDI score was associated with their gender, age, education, occupation and region. | Findings of this study suggest the following recommendations for future interventions: (1) more attention needs to be paid to vulnerable groups (...); (2) accessibility to medical resources and the public health service system should be further strengthened and improved (...); (3) nationwide strategic planning and coordination for psychological first aid during major disasters (...) should be established and (4) a comprehensive crisis prevention and intervention system (...). | medium |
| Quandt S; LaMonto N; Mora D; Talton J; Laurienti P; Arcury T. | 2021 | COVID-19 Pandemic Among Immigrant Latinx Farmworker and Non-farmworker Families: A Rural-Urban Comparison of Economic, Educational, Healthcare, and Immigration Concerns | USA | HIC | May 1, 2020 to June 5, 2020 | Migrant workers | mental health effect of lockdown measures | Cross-Sectional Study | Analyses show that, although rural and urban groups experienced substantial economic effects, impacts were more acute for urban families. Rural workers reported fewer workplace protective measures for COVID-19. For both groups, fear and worry, particularly about finances and children, dominated reports of their situations with numerous reports of experiencing stress and anxiety. | The results of this study illustrate the contextual vulnerability of Latinx immigrants in North Carolina. Several policy reforms have been identified that can help to alleviate it. Labor and immigration policies are needed so that those immigrants doing essential work can have increased wages and access benefits available to other workers, including sick leave, family leave, nutrition benefits, and healthcare. Policies related to worker safety and health that are specific to COVID-19 are needed to provide protections in the workplace. | high |
| Rabe JE; Schillok H; | 2021 | [Burden in parents of school-aged children during | Germany | HIC | March 2020 to | International migrants | effect of lockdown measures | Repeated Cross- | During the first COVID-19 wave, parents of school-aged children were significantly more burdened compared to the general | School measures for infection control have to be weighed carefully against the psychological impact on parental burden | high |

| | | | | | | | | | | | |
|---|---|---|---|---|---|---|---|---|---|---|---|
| Merkel C et al. | | different phases of the COVID-19 pandemic in Germany: an analysis within the COVID-19 snapshot monitoring (COSMO) study] | | | January 2021 | | | Sectional Study | study population. However, burden decreased significantly from March/April to June 2020. During the second COVID-19 wave in January 2021, burden was homogeneously high across all groups. Single parenthood, a low household income, having a chronic health condition, a COVID-19 infection and a migration background were associated with higher burden, although none of these factors was consistently significant across the survey waves. | with subsequent negative impact on the family system. | |
| Rahaman M; Roy A; Chouhan P; Das KC; Rana MJ. | 2021 | Risk of COVID-19 Transmission and Livelihood Challenges of Stranded Migrant Labourers during Lockdown in India | India | LMIC | not reported | Internal migrants | effect of lockdown measures | Cross-Sectional Study | The factors of COVID-19 transmission such as poor housing, co-morbidities, poor practice of WASH and COVID-19 precautions were signifcantly high among the migrant labourers. The lockdown created livelihood crisis among them. For instance, ration shortage (86%), fnancial distress (82%), reduction of wages (13%), job loss (86%) and anxiety for COVID-19 infection (81%) were often seen. Many of the labourers did not receive any ration kits (30%) and fnancial assistance (86%) during lockdown. | The governmental assistance to overcome the stranded migrant labourers' challenges during lockdown was less than desirable. India needs to frame a sustainable and efective policy for social security for labourers, particularly in emergency situations. | low |
| Ramirez-Aldana R; Gomez-Verjan JC; Bello-Chavolla OY; Garcia-Pena C. | 2020 | Modelling the impact of health-related variables, age, migration, and socio-economic factors in the geographical distribution of early tested case-fatality risks associated with COVID-19 in Mexico | Mexico | UMIC | not reported | Internatio nal migrants | risk of transmissio n | Cross-Sectional Study | As results, considering health related variables; diabetes and obesity were highly associated with COVID-19 fatality. We identified that both external and internal migration had an important impact over early COVID-19 risks in Mexico, with external migration having the second highest impact when analyzing Mexico as a whole. | In conclusion, metabolic diseases, internal and external migration, physicians-to-population ratio, GDP per capita in states without the biggest cities, and age group between 10 to 39 years old were significantly associated with early COVID-19 fatality risks in Mexico. These predictors likely influence the growth of the pandemic moving forward, but variables as prevalence of metabolic diseases cannot be easily modified in the short-term | high |
| Rao N; Narain N; Chakraborty S; Bhanjdeo A; Pattnaik A. | 2020 | Destinations Matter: Social Policy and Migrant Workers in the Times of Covid | India | LMIC | December 1, 2019 to June 2020 | Migrant workers | effect of lockdown measures | Qualitative research | While social identity and regional location are seen as established mediators of migrant experiences, in the present context their role and impact was subdued. Social policy frameworks in the destination states, or their absence, played a key role in shaping migrant experience. Kerala, with a better social infrastructure, but also a specific legal and policy framework recognizing the role and contributions of | Constitution of India offers all citizens the freedom to live and work in any part of the country, yet the pandemic exposed the loopholes in ensuring rights in the context of population mobility, placing the spotlight on the nature of rights and their portability. While this is critical, the discursive agenda of social protection is now focused only on basic needs to food and shelter, rather than a transformational agenda that could | medium |

| | | | | | | | | | | |
|---|---|---|---|---|---|---|---|---|---|---|
| | | | | | | | | | migrant workers as 'guest workers', was able to respond more effectively to their needs and demands. Amongst the other states, Maharashtra falls in between: with some recognition of workers' rights yet divided by a strong movement of regional pride and exclusivity. Gujarat and Uttar Pradesh emerge at the bottom, offering no rights or benefits to the migrant workers, and in fact, denying any claims to citizenship that they put forth. | eventually ensure equal opportunities and choices to all citizens of the country to lead fulfilling lives. | |
| Ravens-Sieberer U.; Kaman A.; Erhart M.; Devine J.; Schlack R.; Otto C. | 2020 | Impact of the COVID-19 pandemic on quality of life and mental health in children and adolescents in Germany | Germany | HIC | May 2020 to June 2020 | Internatio nal migrants | mental health effect of lockdown measures | Cross-Sectional Study | Two-thirds of the children and adolescents reported being highly burdened by the COVID-19 pandemic. They experienced signifcantly lower HRQoL (40.2% vs. 15.3%), more mental health problems (17.8% vs. 9.9%) and higher anxiety levels (24.1% vs. 14.9%) than before the pandemic. Children with low socioeconomic status, migration background and limited living space were afected signifcantly more. | Health promotion and prevention strategies need to be implemented to maintain children's and adolescents' mental health, improve their HRQoL, and mitigate the burden caused by COVID-19, particularly for children who are most at risk. | high |
| Rayan I; Qaddomi SE; Najjar O et al. | 2021 | Sero Prevalence of COVID-19 in Palestine in 2020 | Palestine | n.a. | Decembe r 2020 | Refugees and IDP | risk of infection | Cross-Sectional Study | The results show clearly that a large percentage of the Palestinian population in the West Bank and Gaza had been infected with COVID-19 before the study was conducted. While rt-PCR tests indicated that 4% of the population had been infected by the end of 2020, sero-t testing indicated that nearly 40% of the population had been detected within the same time frame, meaning that rt-PCR testing had detected only 10% of cases. (...) The type of residence was a major factor in seroprevalence. Refugee camps in the West Bank were at much higher risk than rural areas. | The Sero Survey revealed that Palestine has 10 times more cases than previously thought, meaning that the epidemic reached far more people than previously anticipated. (...) Certain groups are at more risk than others: females, diabetics, older people, and low-income families. Specific behaviors have serious repercussions on the likelihood of infection: travel, type of employment, awareness of contact, and suspicion of infection. This means that individuals can significantly reduce the likelihood of infection if they are made aware of these factors. | medium |
| Raynor K; Panza L. | 2021 | Tracking the impact of COVID-19 in Victoria, Australia: Shocks, vulnerability and insurances among residents of share houses | Australia | HIC | June 2020 | Internatio nal migrants | mental health | Cross-Sectional Study | We found dramatic results, with 74% losing their job or having their hours reduced, 47% experiencing a reduction in their financial situation and 50% reporting that their mental health had deteriorated. These outcomes were worse for young people, casual employees or immigrants. | Our research highlights the positive influence of social support for low-income individuals. We find that government social welfare payments are the most impactful form of insurance, calling for a greater appreciation of the role of social welfare in supporting resilience following a disaster. | medium |

| | | | | | | | | | | |
|---|---|---|---|---|---|---|---|---|---|---|
| Redditt V; Wright V; Rashid M; Male R; Bogosch I. | 2020 | Outbreak of SARS-CoV-2 infection at a large refugee shelter in Toronto, April 2020: a clinical and epidemiologic descriptive analysis | Canada | HIC | April 2020 | Refugees | risk of transmission | Cross-Sectional Study | We found a high incidence of SARS-CoV-2 infection (41.7%, 25/60) with universal testing of on-site shelter residents. The overall estimated positivity rate among all 87 residents is likely higher, accounting for individuals who tested positive before Apr. 20, 2020, and thereafter. | We found a high rate of SARS-CoV-2 infection in this shelter population. Our study underscores the high risk of SARS-CoV-2 transmission in congregate living settings and the importance of mobilizing timely testing and management of symptomatic, paucisymptomatic and asymptomatic residents in shelters. | medium |
| Reynolds CW; Ramanathan V; Lorenzana E et al. | 2021 | Challenges and Effects of the COVID-19 Pandemic on Asylum Seeker Health at the U.S.-Mexico Border | Mexico | UMIC | August 2020 | Asylum Seekers | effect of lockdown measures | Qualitative research | The pandemic caused significant mental health burdens but no perceived adverse physical effects, with the U.S. border closure and health care access barriers as more pressing concerns. Participants reported access to information about COVID-19 but had varied levels of know-ledge and adherence to disease reduction strategies due to camp conditions. Most participants believed that they had special protection from COVID-19, including strong immune systems or from God. The nongovernmental organizations providing health care and sanitation faced multiple challenges to implement new policies to manage COVID-19. The institution of required temperature checks and quarantine of COVID-19 positive patients led to distrust, decreased seeking of health care services among asylum seekers, and possible underreporting of COVID-19 cases. | Our findings among asylum seekers in a Matamoros camp highlight the challenges to implementing disease reduction policies in low-resource congregate camps. Policies to address disease outbreaks focusing on the social determinants of health, health care access barriers, and community engagement may be more acceptable to asylum seekers, suggesting the need for effective strategies to provide prevention information that complements such measures. | medium |
| Riley AR; Chen YH; Matthay EC et al. | 2021 | Excess deaths among Latino people in California during the COVID-19 pandemic | USA | HIC | March 1, 2020 to October 3, 2020 | International migrants | health outcomes of disease | Modelling Study | Our results indicate that during the first seven months of the pandemic, Latino deaths in California exceeded expected deaths by 10,316, a 31% increase. Excess death rates were greatest for individuals born in Mexico (RR 1.44; 95% PI, 1.41, 1.48) or a Central American country (RR 1.49; 95% PI, 1.37, 1.64), with less than a high school degree (RR 1.41; 95% PI, 1.35, 1.46), or in food-andagriculture (RR 1.60; 95% PI, 1.48, 1.74) or manufacturing occupations (RR 1.59; 95% PI, 1.50, 1.69). | In sum, the COVID-19 pandemic has disproportionately impacted mortality among Latino immigrants, especially those in unprotected essential jobs. Interventions to reduce these inequities should include targeted vaccination, work-place safety enforcement, and expanded access to medical care and economic support. | medium |
| Rocha L; Rose R; Hoch A et al. | 2021 | The Impact of the COVID-19 Pandemic on the Brazilian Immigrant | USA | HIC | June 2020 to July 2020 | International migrants | mental health health services | Qualitative research | Key informants perceived that community members faced higher risk of COVID-19 infection due to overcrowded living conditions and over-representation in | Our findings indicate that the Brazilian community has endured a variety of challenges such as financial insecurity, fears about documentation status, and language | low |

| | | | | | | | | | | | |
|---|---|---|---|---|---|---|---|---|---|---|---|
| | | Community in the U.S: Results from a Qualitative Study | | | | | access effect of lockdown measures | | public-facing and informal (e.g., housecleaning) jobs. They reported barriers to COVID-19-related healthcare services including language, immigration status, and fear of deportation. | barriers, which have left them vulnerable to the impacts of the COVID-19 pandemic. (…) Given that our findings indicate a shortage of adequate government support, capturing the number of COVID-19 infections and deaths among immigrant communities will help demonstrate the need for more resources. | |
| Rostila M; Cederström A; Wallace M; Branden M; Malmberg B; Andersson G. | 2021 | Disparities in Coronavirus Disease 2019 Mortality by Country of Birth in Stockholm, Sweden: A Total-Population-Based Cohort Study | Sweden | HIC | January 31, 2020 to May 4, 2020 | International migrants | health outcomes of disease: Mortality; effect of lockdown measures | Cohort Study | Migrants from Middle Eastern countries (relative risk (RR)=3.2,95% confidence interval (CI): 2.6, 3.8), Africa (RR=3.0, 95% CI: 2.2, 4.3), and non-Sweden Nordic countries (RR=1.5, 95% CI: 1.2, 1.8) had higher mortality from COVID-19 than persons born in Sweden. Especially high mortality risks from COVID-19 were found among persons born in Somalia, Lebanon, Syria, Turkey, Iran, and Iraq. Socioeconomic status, number of working-age household members, and neighborhood population density attenuated up to half of the increased COVID-19 mortality risks among the foreign-born. | In this total population study, we found large disparities in COVID-19 mortality by country of birth in Stockholm-the worst-affected area in Sweden and one of the most affected regions in the world. Migrants' disadvantaged socio-economic and living conditions explained a considerable share of their higher COVID-19 mortality risk. Policies aiming to reduce disparities in COVID-19 mortality by ethnicity/ country of birth should consider such conditions when designing health interventions. However, it is crucial to continue to study risk factors explaining migrants' higher prevalence of COVID-19 deaths in order to prevent such disparities in both Sweden and other countries with substantial migrant populations. | high |
| Rzymski P; Nowicki M. | 2020 | COVID-19-related prejudice toward Asian medical students | Poland | HIC | February 2020 to March 4, 2020 | International students | effect of lockdown measures | Cross-Sectional Study | We have found that 61.2% of the surveyed students have experienced prejudice in Poland related to the current coronavirus epidemic, and it was more frequently witnessed by those wearing face masks than those who do not (71.2% vs 28.2%). The prejudice most commonly encountered, by 47.1%, in public transportation and on the street. | In conclusion, the reactions presented here clearly show that Asian students in regions yet unaffected by SARS-CoV-2 could have already experienced an uncomfortable level of prejudice inthe public spaces encompassing transport, gastronomy, shopping, health services and university. Such behaviors can particularly affect those individuals of Asian origin who are tending to wear face masks. | high |
| Sabar G; Babis D; Sabar Ben-Yehoshua N. | 2021 | From Fragility to Empowerment through Philanthropy: The Filipino Labor Migrant Community in Israel During COVID-19 | Israel | HIC | March 2020 to May 2020 | International migrants | effect of lockdown measures | Mixed-Methods: Qualitative research; Cross-Sectional Study | During the lockdown, the challenges embedded in caregiving in Israel were amplified and new challenges emerged, such as the inability to leave the employer's premises, which some likened to being imprisoned. The major hardships voiced in all interviews were loneliness, fear of contracting the virus, worries about the safety of their families back home, and anger at restrictions imposed without consulting them by the assisted living | Philanthropy for Filipinos in need in Israel and back home was found to be an effective coping mechanism. While lockdown conditions increased the fragility of the marginalized migrant workers, their ability to donate funds empowered them at both individual and communal levels. Developing a donor identity reinforced their sense of belonging to the Filipino community in Israel and back home. | medium |

| | | | | | | | | | | |
|---|---|---|---|---|---|---|---|---|---|---|
| | | | | | | | | | facilities or their employers. At the same time, they expressed sincere gratitude for being employed, paid and safe. | | |
| Salem Al-Abri S; Al-Wahaibi A; Al-Kindi H et al. | 2021 | Seroprevalence of SARS-CoV-2 antibodies in the general population of Oman: results from four successive nationwide sero-epidemiological surveys | Oman | HIC | July 2020 to November 2020 | International migrants | risk of infection | Cross-Sectional Study | The seropositivity rate was higher in migrants during Cycles 1 and 2, with a subsequent increase in Omanis in Cycles 3 and 4. In addition, the risk of infection was higher in labourers compared with office workers, as supported by the logistic regression model. | The substantially higher seroprevalence estimates in migrants in Cycles 1 and 2 of the study reflects the high efficiency of SARS-CoV-2 spread in congregate living facilities and work settings, and the effectiveness of occupational health measures put into place to reduce transmission in these settings. | high |
| Sanfelici M. | 2021 | The impact of the COVID-19 crisis on marginal migrant populations in Italy | Italy | HIC | April 2020 | International migrants | effect of lockdown measures health services access | Qualitative research | Our analysis of the response stage showed the serious challenges faced by migrants and professionals, and a joint effort of several actors in civil society in tackling these circumstances, showing vulnerability and resilience both at the individual and at the community level. | Nevertheless, even if the COVID-19 crisis has created the possibility to make structural problems more visible, and indicated the direction to “build back better,” the cultural and structural variables that create a condition of hyperprecarity for more marginalized migrants seem to be unchanged. | low |
| Sanna J. | 2021 | The othering of returning migrants in Romania during the first wave of the COVID-19 pandemic: Event analysis | Romania/ Italy | UMIC/HIC | March 2020 | Retournees | effect of lockdown measures | Qualitative research | Among the reasons to return during the pandemic that my informants mentioned there were: the sudden loss of a job due to temporary or permanent closing of businesses during the lockdown in Italy and elsewhere; the closing of universities, in the case of Romanian students studying abroad; and the impossibility of affording healthcare in a foreign country due to not having a regular work contract. In other cases, of course, it was the fear of being sick in a foreign country that mattered most. | In this paper, I have shown how the image of returning migrants during the first month of the COVID-19 pandemic in Romania was constructed by the media in March 2020: by very quickly associating the migrants with the virus, by juxta-posing the noble acts of Romanians left in Italy with the criminal acts of those who came back, and by reinforcing such juxta-position by giving voice to Romanians who stayed in Italy. I have also shown that Romanians who did not come back were characterized in depth, while descriptions of returning Romanians, except for a few cases, remained vague. | medium |
| Sano G; Della Puppa F. | 2021 | The multiple facets of (im)mobility. A multisited ethnography on territorialisation experiences and mobility trajectories of asylum seekers and refugees outside the Italian reception system | Italy | HIC | February 2020 to May 2020 | International migrants | effect of lockdown measures | Qualitative research | These two interviews show how the mobility and immobility of these subjects were differently conditioned by the intersection of several factors. (…) That is to say, the closure of the Trentino hotels and restaurants during the winter season has meant that most of the migrants employed in this sector have moved to the city, determining, among other things, the opening of services to low threshold able to accommodate all the people left without work in the valleys. On the contrary, in the Plain of Gioia Tauro, the slowdown in | The ethnographic cases we described show the need to rethink the conceptual categories with which we usually analyse mobility, illuminating the links it has with national policies, with territorial administrative provisions, with the economic and social fabric of an area, with individual and not necessarily ‘rational’ choices of individuals. Observed in these terms and based on these links, one of the distinctive features of contemporaneity, namely mobility, reveals all its political nature, the disempowering character that accompanies it, as well as the | medium |

| | | | | | | | | | | | |
|---|---|---|---|---|---|---|---|---|---|---|---|
| | | | | | | | | | production was not determined by a general closure of agricultural activities, but, mainly, by the fact that, at the beginning of the pandemic, a good part of the labourers present were without regular contract and thus could not go to the workplace. | economic interests connected to the productive function of those who find themselves imprisoned in the multiplicity facets of the prism of (im)mobility. | |
| Santos GM; Ackerman B; Rao A et al. | 2021 | Economic, Mental Health, HIV Prevention and HIV Treatment Impacts of COVID-19 and the COVID-19 Response on a Global Sample of Cisgender Gay Men and Other Men Who Have Sex with Men | Brazil; France; Mexico; Taiwan; Russia | HIC; UMIC | April 16, 2020 to May 4, 2020 | Internatio nal migrants | health services access | Cross-Sectional Study | Many gay men and other MSM not only reported economic and mental health consequences, but also interruptions to HIV prevention and testing, and HIV care and treatment services. These consequences were significantly greater among people living with HIV, racial/ethnic minorities, immigrants, sex workers, and socio-economically disadvantaged groups. | These fndings highlight the urgent need to mitigate the negative impacts of COVID-19 among gay men and other MSM. | high |
| Saw YE; Tan EYQ; Buvanaswa ri P; Doshi K; Liu JCJ. | 2021 | Mental health of international migrant workers amidst large-scale dormitory outbreaks of COVID-19: A population survey in Singapore | Singapor e | HIC | June 22, 2020 to October 11, 2020 | Migrant workers | mental health | Cross-Sectional Study | Complete movement restrictions were associated with increased depression and stress symptoms, while being diagnosed with COVID-19 was associated with increased anxiety. (...) Across the cohort, rates of severe or extremely severe depression (3.1%, 95% CI: 2.1-4.3%), anxiety (4.1%, 95% CI: 2.9-5.5%), and stress (1.3%, 95% CI: 0.7-2.2%) were similar to those observed in the general population for the host country (Singapore). | As the first mental health survey of low-waged migrant workers during the pandemic, our findings provide a basis to identify and support at-risk individuals. This is critical as migrant workers often have limited access to healthcare services. At the same time, we documented resilience within the cohort as a whole. | high |
| Schestag L; Mehner-Gentner J; Stein L et al. | 2021 | Ghosts in the nursery in exile-supporting parenting in exile during the covid-19 pandemic | Germany | HIC | 2018 to 2021 | Refugees | mental health | Mixed-Methods | For refugee families who have only recently arrived in Germany, the COVID-19 pandemic poses a particularly great challenge. (...) Many expressed great insecurity, heightened levels of anxiety, re-experiencing of traumatic scenes, even attraction to fundamentalist ideologies. | We are only at the beginning of understanding the effects of the COVID-19 pandemic. However, (...), it is evident that those severely traumatized are particularly endangered by the reactivation of their traumatic experiences as their traumatic histories have not been sufficiently internally integrated. | low |
| Seidu B. | 2020 | Optimal Strategies for Control of COVID-19: A Mathematical Perspective | not related to a specific country or context | - | not reported | Internatio nal migrants | effect of lockdown measures | Modellin g Study | It is shown that unless the immigration of infected persons is restricted, the disease cannot be eradicated, and that if border closure is strictly adhered to, the disease can be eradicated if a threshold parameter R0 is kept below unity. | The model is modified into an optimal control problem by seeking to minimize an objective function that measures total infected persons and Surface viral concentration and the total cost associated with implementing the various controls. It is shown that the best strategy in controlling the spread of COVID-19 is social distancing and use of nose masks. | low |

| | | | | | | | | | | | |
|---|---|---|---|---|---|---|---|---|---|---|---|
| Sempere-Gonzalez A; Salvador F; Monforte A et al. | 2021 | COVID-19 Clinical Profile in Latin American Migrants Living in Spain: Does the Geographical Origin Matter? | Spain | HIC | March 1, 2020 to April 30, 2020 | International migrants | health outcomes of disease: Hospitalisation, ICU admission | Cohort Study | Latin American patients (n=142) had a higher proportion of anosmia, rhinorrhea and odynophagia, as well as higher mean levels of platelets and lower mean levels of ferritin than Spanish patients. No differences were found in oxygen requirement and mortality at 28 days after admission, but there was a higher proportion of ICU admissions (28.2% vs. 20.2%, p= 0.0310). An increased proportion of ICU admissions were found in patients from Latin America compared with native Spanish patients when adjusted by age and gender, with no significant differences in in-hospital mortality. | In summary, we observed an increased rate of ICU admissions among hospitalized adult patients with COVID-19 pneumonia from Latin America compared with native Spanish patients. Furthermore, there was a higher rate of arthromyalgia, cacosmia andanosmia observed in patients from Latin America. These differences could be explained by socio-economic factors or still unknown genetic factors, so further investigations are needed. Understanding ethnic disparities can help so as to create health prevention strategies. | high |
| Sharov KS. | 2021 | SARS-CoV-2 spread in different biosocial strata in Russia in 2020: Groups of risk and victimised groups | Russian Federation | MIC | May 2020 to December 2020 | Migrant workers | risk of infection | Longitudinal Study | Point-by-point analysis of the data in the antibody testing set collected in ten Russian regions studied (...), shows that there is almost no correlation between the refugee/migrant status of a person and probability of his/her being infected with SARS-CoV-2 (average Spearman correlation coefficient C = 0.2716 at P less than 0.4431). | The major conclusion deduced (…), is that migrants and refugees do not contribute to spreading SARS-CoV-2 in Russia significantly. | n.a. |
| Shimizutani S; Yamada E. | 2021 | Resilience against the pandemic: The impact of COVID-19 on migration and household welfare in Tajikistan | Tajikistan | LMIC | January 2018 to December 2020 | International migrants | effect of lockdown measures | Cross-Sectional Study | First, the adverse effect of the pandemic was severe in the second quarter of the 2020s. Second, contrasting to expectation, the pandemic had a sharp but only transitory effect on the number of migrants temporarily staying abroad. (...) Third, despite a sharp decline in employment and the remittances of migrants in April and May, both factors quickly recovered. Regression analyses confirmed migration and remittances and employment at the destination had eased the economic shock of the COVID-19 pandemic in terms of not only material well-being such as food, utilities, consumption, and health but also their views on their financial and economic situations. | Overall, the unfavorable effects of the COVID-19 pandemic were severe and temporary right after the outbreak, but households with migrants were more resilient against the pandemic. | low |
| Silva H; Nogueira I; Cordeiro C et al. | 2021 | Migration in times of pandemic: SARS-CoV-2 infection among the Warao indigenous refugees | Brazil | UMIC | September 2020 to January 2021 | Refugees | risk of transmission | Cross-Sectional Study | positive serum prevalence of 83.2% (84), of which 77.6% (45/58) were females and90.7% (39/43) were males. | The finding of the high seroprevalence of IgG anti-SARS-CoV-2 antibodies reveals a high exposure of the Warao population in Belem to infection with the new coronavirus. | medium |

| | | | | | | | | | | | |
|---|---|---|---|---|---|---|---|---|---|---|---|
| | | in Belem, Para, Amazonia, Brazil | | | | | | | | | |
| Singh GP; Arun P; Chavan B. | 2020 | Migrant Workers' Needs and Perceptions While Lodged in a Shelter Home in India During the COVID-19 Pandemic | India | LMIC | not reported | Migrant workers | effect of lockdown measures; mental health | Qualitative research | Our study found that physiologic requirements, safety, and security were the predominant needs of the migrant workers during their stay in a shelter home during COVID-19-related lockdown. All of the migrants were satisfied with the need-based services provided to them in Chandigarh. The migrant workers were worried about being infected by COVID-19 and were uncertain about when they would be able to return to their native homes. | Lockdown drained migrant workers both physically and emotionally. Despite their hardship and suffering, most of the migrant workers supported the government's decision to impose lockdown. Comprehensive support should be provided to all migrant workers in all states of India to combat the migrant crisis. This support, as was provided in Chandigarh during lockdown, would safeguard the well-being of migrant workers in India. | medium |
| Sohel MS; Hossain B; Sarker MNI; Horaira GA; Sifullah MK; Rahman MA. | 2021 | Impacts of COVID-19 induced food insecurity among informal migrants: Insight from Dhaka, Bangladesh | Bangladesh | LMIC | April 2020 to November 2020 | Migrant workers | effect of lockdown measures | Qualitative research | The results show that COVID-19 induced lockdown significantly affects informal migrants' household food security. Most participants report severe complications due to food insecurity like consuming less food, price hiking, no fish or meat, potato, and vegetable. Food insecurity leads to migrant's chronic food shortage, starvation, malnutrition of mother and children, and unhealthy food. As a result, the way of life of informal migrants has been directed to more fragility and vulnerability during the pandemic; even they are not affordable to maintain a minimal level of family affairs and necessity. | The COVID-19 led pandemic lockdown severely impacts food security among informal migrants' households. Most households confront enormous problems and challenges to meet up the daily food and its associated elements. (…) Moreover, they did not get what they needed at the time of the crisis caused by the COVID-19 pandemic at a satisfactory level from the government and other agencies. This study also uncovers the post-lockdown reality of impoverished informal migrants who are often excluded from Bangladesh's social safety net facilities. | medium |
| Song S; Zang S; Gong L et al. | 2021 | Willingness and Uptake of the COVID-19 Testing and Vaccination in Urban China: An Online Cross-Sectional Survey at Early 2021 (preprint) | China | UMIC | January 29, 2021 to February 4, 2021 | International migrants | vaccination | Cross-Sectional Study | Willingness and uptake of COVID-19 testing were not associated with socio-demographic characteristics, except for occupation. Being of older age, migrants, having higher educational attainment and secure employment were associated with a higher uptake of COVID-19 vaccination among the surveyed respondents, while willingness to vaccinate was consistent across socio-demographic haracteristics among those who had not been vaccinated. | Chinese adults expressed almost universal willingness of COVID-19 testing and relatively low willingness of COVID-19 vaccination at the low-risk period of the COVID-19 pandemic. The willingness and uptake of COVID-19 testing and vaccination are independent from their socio-demographic characteristics at most cases. | medium |
| Sorg A.; Bergfeld L; Jank M et al. | 2022 | Cross-sectional seroprevalence surveys of SARS-CoV-2 antibodies in children in | Germany | HIC | June 2020 to May 2021 | International migrants | risk of infection | Cross-Sectional Study | Clear population differences in SARS-CoV-2 infection have been reported since the pandemic outbreak. The differences occurred in terms of occupational or age groups, and when stratified by ethnicity... The higher prevalence of SARS-CoV-2 | The increase in seroprevalence varied by age group, with a higher prevalence in young children during June to September 2020 and by migrant background. The impact of measures to limit virus spread might have | high |

| | | | | | | | | | | | |
|---|---|---|---|---|---|---|---|---|---|---|---|
| | | Germany, June2020 to May 2021 | | | | | | | antibodies in children with migrant background in the present study resembles these findings. Causes for differences in seroprevalence according to migrant background might be related to temporal and regional variation in incidence, structural and systemic differences such as higher risk occupations, and differences in mobility patterns or households [sic] sizes. The volatile interplay of these mechanisms may explain the observed changes of the effect of migrant background on seropositivity over time. | been improved by approaches taking these factors into account. | |
| Soussy N; Dufayet L; Ravault E; Viron S; Gorgiard C; Fournier L. | 2021 | Management of the COVID-19 epidemic in a French immigration detention center | france | HIC | April 2020 | International migrants | risk of transmission; health outcomes of disease | Cross-Sectional Study | The test was done for all the present individuals (48), except two who refused. Eight tests (17.4%) were positive and only one was symptomatic. Individuals testing positive for COVID-19 were transferred into COVID-centers specially created during this health crisis. | Good cooperation between the judicial authorities and the medical team in charge of the IDC resulted in a limited number of infections and no severe infection or death was recorded in the Paris IDC population. | low |
| Srivastava A; Arya YK; Joshi S et al. | 2021 | Major Stressors and Coping Strategies of Internal Migrant Workers During the COVID-19 Pandemic: A Qualitative Exploration | India | LMIC | not reported | Internal migrants | effect of lockdown measures; mental health | Qualitative research | The findings of the present research shows that internal migrant workers were highly affected due to the global pandemic, which bombarded multiple stressors on them causing physical stress, emotional stress, and social stress in their lives, which were created due to financial crisis, lack of social support, and fear and uncertainty related to the global pandemic. However, the workers looked after the options available to them and chose problem-oriented strategy by directly dealing with the crisis, emotion-oriented strategy by family and friends' support, as well as cognitive appraisal of the pandemic situation by bringing optimistic thoughts to deal with these stressors. | The present work strengthens the earlier studies on uncovering the stresses of migrant workers along with suggesting available options to deal with them and emphasizes the role of family and proper emotional support in the management of pandemic-related stress among internal migrant workers. | medium |
| Strully K; Yang TC; Liu H. | 2020 | Regional variation in COVID-19 disparities: connections with immigrant and Latinx communities in U.S. counties | USA | HIC | (data until) May 28, 2020 | International migrants (%) | risk of infection | Cross-Sectional Study | Percentage of foreign-born residents is positively associated with COVID-19 case rate (IRR: 1.106; 95% CI: 1.074e1.139). Adjusted associations for percentage Latinx are non significant for all counties combined, but this obscures heterogeneity. Counties with more Central Americans have higher case rates(IRR: 1.130; 95% CI: 1.067e1.197). And, in the Northeast and Midwest, counties with more Puerto | Counties with more immigrants, as well as more Central American or Black residents, have more COVID-19 cases. In the Northeast and Midwest, counties with more Puerto Rican residents also have more COVID-19 cases. | high |

| | | | | | | | | | | | |
|---|---|---|---|---|---|---|---|---|---|---|---|
| | | | | | | | | | Ricans have higher case rates. Associations with percentage Asians are non significant after adjusting for percentage foreign-born. With the confirmation of prior evidence, the percentage of Black residents is positively and robustly associated with COVID-19 case rate (IRR: 1.031; 95% CI: 1.025e1.036). | | |
| Sundaram ME.; Calzavara A; Mishra S et al. | 2021 | The Individual and Social Determinants of COVID-19 in Ontario, Canada: A Population-Wide Study | Canada | HIC | March 1, 2020 to June 20, 2020 | Internatio nal migrants | risk of infection | Cross-Sectional Study | We found that there was less variability in testing by social determinants across analytic designs. Residing in areas with the highest household density (adjusted odds ratio [OR] 1.86, 95% confidence interval [CI] 1.75-1.98), highest proportion of essential workers (adjusted OR 1.58, 95% CI 1.48-1.69), lowest educational attainment (adjusted OR 1.33, 95% CI 1.26-1.41) and highest proportion of recent immigrants (adjusted OR 1.10, 95% CI 1.05-1.15) were consistently related to increased odds of SARS-CoV-2 diagnosis regardless of analytic design. | Optimizing COVID-19 responses necessitates investment in and sufficient coverage of structural interventions tailored to heterogeneity in social determinants of risk, including household crowding, occupation and structural racism. | high |
| Suphanchai mat R; Tuangratan anon T; Rajatanavin N; Phaiyarom M; Jaruwanno W; Uansri S. | 2021 | Prioritization of the Target Population for Coronavirus Disease 2019 (COVID-19) Vaccination Program in Thailand | Thailand | UMIC | not reported | Internatio nal migrants | vaccination ; health outcomes of disease | Modellin g Study | The migrant-centric vaccination policy scenario received the lowest incremental cost per one case or one death averted compared with the other scenarios. The Thai-centric policy scenario yielded an incremental cost of 27,191 Baht per one life saved, while the migrant-centric policy scenario produced a comparable incremental costof 3782 Baht. | A migrant-centric policy yielded the smallest volume of cumulative infections and deaths and was the most cost-effective scenario, independent of R and VE values. Further studies should address political feasibility and social acceptability of migrant vaccine prioritization. | low |
| Svensson P; Hofmann R; Haebel H; Jernberg T; Nordberg P. | 2021 | Association between cardiometabolic disease and severe COVID-19: a nationwide case-control study of patients requiring invasive mechanical ventilation | Sweden | HIC | March 1, 2020 and May 11, 2020 | Internatio nal migrants | health outcomes of disease | Case-Control Study | In this nationwide case-control study, 1.086 patients admitted to intensive care with COVID-19 requiring mechanical ventilation (cases), and 10.860 population-based controls matched for age, sex and district of residency were included from mandatory national registries. (...) The risk associated with type 2 diabetes was higher in women (p=0.001) and in patients with a region of birth outside European Union(EU) (p=0.004). | Diabetes, obesity and hypertension were all independently associated with severe COVID-19 with stronger associations in the younger population. Type 2 diabetes implied a greater risk among women and in non-EU immigrants. These findings, originating from high-quality Swedish registries, may be important to direct preventive measures such as vaccination to susceptible patient groups. | high |

| | | | | | | | | | | | |
|---|---|---|---|---|---|---|---|---|---|---|---|
| Tan IB; Tan C; Hsu LY et al. | 2020 | Prevalence and Outcomes of SARS-CoV-2 Infection Among Migrant Workers in Singapore | Singapore | HIC | March 25, 2020 to July 25, 2020 | Migrant workers | risk of infection; health outcomes of disease | Cross-Sectional Study | The COVID-19 outbreak among Singapore's migrant workers was characterized by a high prevalence of infection, low morbidity with few ICU admissions, and low mor-tality. The low case fatality rate compares favorably with ratesreported in other national registries | Although migrant workers are younger and generally healthy, the nationally coordinated public health response and clinical care for all cases likely contributed to favorable health outcomes. The infection prevalence would have been severely underestimated if only persons presenting for medical consultation were tested because most infections were subclinical and were identified through mass screening. | medium |
| Tang S; Li X. | 2021 | Responding to the pandemic as a family unit: social impacts of COVID-19 on rural migrants in China and their coping strategies | China | UMIC | April 2020 to July 2020 | Rural migrants | effect of lockdown measures | Qualitative research | The study finds that rural migrants suffered from serious social impacts due to COVID-19, especially during the associated lock-down period. Despite some similar impacts, influences of COVID-19 varied among rural migrants at different life-cycle stages, due to variations in human capital, family burdens, role in a household, and ability to find part-time work. Receiving little support from governments and employers, rural migrants tended to adopt household strategies to deal with difficulties related to COVID-19. Within a household, they assisted each other and worked as a unit to maximize resources and reduce risks. Traditional family values were highly praised by rural migrant households during the period. | Receiving little support from government and employers, rural migrants in China have to adopt a household strategy as a means of survival when they cope with disaster. If governments want to reduce the burden of COVID-19 on disadvantaged households, they need to pay more attention to rural migrants, even though they are neither local urban residents nor actual rural residents. (...) In the long-term, the social security system for rural migrants should be further improved. It is necessary to expand the accessibility of basic social services, such as medical insurance, housing funds, and pensions to rural migrants, and to include rural migrants in the housing security system, in case they experience difficult times. | medium |
| Tee LY; Alhamid SM; Tan JL et al. | 2020 | COVID-19 and Undiagnosed Pre-diabetes or Diabetes Mellitus Among International Migrant Workers in Singapore | Singapore | HIC | April 21, 2020 to June 1, 2020 | Migrant workers | risk of infection; health outcomes of disease | Cohort Study | Two hundred and fouty male migrant workers, (...), were included. Twenty one patients (8.8%) were diagnosed with pre-diabetes, and 19 (7.9%) with DM [diabetes mellitus]. (...) Moreover, patients with DM or pre-diabetes developed COVID-19 infection with lower viral RNA levels. | The high prevalence of undiagnosed pre-diabetes among international migrant workers increases their risk of pneumonia and electrolyte abnormalities from COVID-19. | medium |
| Terp S; Ahmed S; Burner E et al. | 2021 | Deaths in Immigration and Customs Enforcement (ICE) detention: FY2018-2020 | USA | HIC | 2018 to 2020 | International migrants | health outcomes of disease | Cohort Study | Since April 2018, 35 individuals have died in ICE detention. (...) Among 26 deaths attributable to medical causes, 8 (30.8%) were attributed to COVID-19, representing 72.7% of 11 deaths occurring since April 2020. | Findings suggest that individuals detained by ICE may benefit from improved psychiatric care and prevention measures to combat suicide, as well as increased infection control efforts to reduce mortality associated with COVID-19. | low |
| Thomas J; de Wit E; Radhakrishnan RK; Kulkarni | 2021 | Mitigating the COVID-19 pandemic in India: an in-depth exploration of | India | LMIC | March 1, 2020 to April 27, 2020 | Migrants workers, Refugees | effect of lockdown measures | Qualitative research | The outcomes of this study shed light on (1) the most urgent needs that need to be addressed per population group, (2) the variety of state-level responses as well as best practices observed to deal with | The COVID-19 pandemic has not only reduced people's means of maintaining a livelihood but has simultaneously revealed some of India's long-standing problems with infrastructure and resource distribution in a | medium |

| | | | | | | | | | | | |
|---|---|---|---|---|---|---|---|---|---|---|---|
| N; Bunders-Aelen J. | | challenges and opportunities for three vulnerable population groups | | | | | | | mitigation issues and (3) opportunities for quick relief as well as more long-term solutions. | range of sectors, including nutrition and health, education, etc. There is an urgent need to construct effective pathways to trace and respond to those people who are desolate, and to learn from – and support – good practices at the grassroot level. | |
| Tosh SR; Berg UD; Leon KS. | 2021 | Migrant Detention and COVID-19: Pandemic Responses in Four New Jersey Detention Centers | USA | HIC | August 2020 to Novembe r 2020 | ICE detainees | risk of infection | Qualitati ve research | Our findings suggest that migrant detention and deportation present distinct challenges that undermine attempts to contain the spread of COVID-19. We provide testimonies from migrant detainees who speak to these challenges in unsettling personal terms. Our interviews highlight the insufficient actions by Immigration and Customs Enforcement (ICE) to contain the spread of the pandemic and a troubling lack of due process in immigration court proceedings. | Based on these findings, we argue that reducing the number of migrants detained in the United States is needed not only in the context of the COVID-19 pandemic but also as a preventative measure for future health crises. Reductions can be achieved, in part, by reforming federal immigration laws on "mandatory detention." | medium |
| Tramuto F; Reale S; Lo Presti A et al. | 2021 | Genomic Analysis and Lineage Identification of SARS-CoV-2 Strains in Migrants Accessing Europe Through the Libyan Route | Italy | HIC | not reported | Internatio nal migrants | risk of transmissio n | Cross-Sectional Study | 28 (13 men, nine women, and six children) out of the tested 209 migrants were positive (13.4%). Our analysis identified some genomic lineages previously detected in different low-income countries. | Our findings documented that most of the investigated migrants acquired SARS-CoV-2 infection before landing in Sicily. However, SARS-CoV-2 transmission during travel or in overcrowded Libyan immigrant camps and/or illegal transport boats could not be ruled out. | medium |
| Truelove S; Abrahim O; Altare C et al. | 2020 | The potential impact of COVID-19 in refugee camps in Bangladesh and beyond: A modeling study | Banglade sh | LMIC | not reportd | Refugees | risk of transmissio n | Modellin g Study | Using a dynamic model of severe acute respiratory syndrome coronavirus 2 (SARS-CoV-2) transmission, we simulated how a COVID-19 outbreak could spread within the expansion site according to three possible transmission scenarios (high, moderate, and low) | A COVID-19 epidemic in a high population density refugee settlement may have profound consequences, requiring increases in healthcare capacity and infrastructure that exceed what is feasible in this setting. | medium |
| Truelove SA; Hegde S; Niehaus L et al. | 2021 | Estimating the Burden of SARS-CoV-2 among the Rohingya Refugees | Banglade sh | LMIC | March 2020 to October 2020 | Refugees | risk of transmissio n<br>health outcomes of disease | Modellin g Study | Among the 144 survey-identified deaths, 48 were consistent with suspected COVID-19. These deaths were consistent with viral exposures during Ramadan, a period of increased social contacts, and coincided with a spike in reported cases and testing positivity in June 2020. The age profile of suspected COVID-19 deaths mirrored that expected. Through the probability framework, we find that under each scenario, a substantial outbreak likely occurred, though the cumulative size and timing varyconsiderably. | With the high population density, inability to home isolate adequately, and limited personal protective equipment, infection prevention and control in the Rohingya population is extremely challenging. Despite the low reported numbers of cases and deaths, our results suggest an early large-scale outbreak is consistent with multiple sources of data, particularly when accounting for limited care seeking behavior and low infection severity among this young population. | low |

| | | | | | | | | | | | |
|---|---|---|---|---|---|---|---|---|---|---|---|
| Turunen T; Kontunen K; Sugulle K et al. | 2021 | COVID-19 outbreak at a reception centre for asylum seekers in Espoo, Finland | Finland | HIC | April 28, 2020 to May 6, 2020 | Asylum Seekers | risk of transmission | Cross-Sectional Study | 260 clients were screened. Of them, 96 (37%) were found positive for SARS-CoV-2 and isolated. (...) Of the 35 initially asymptomatic individuals, 12 developed symptoms during follow-up, while 23 (or 18% of all screened SARS-CoV-2 positive clients) remained asymptomatic. No widespread transmission of COVID-19 was detected after the quarantine was lifted. | In this large COVID-19 outbreak, voluntary mass screening provided valuable information about its extent and helped guide the public health response. Comprehensive quarantine and isolation measures were likely instrumental in containing the outbreak. | medium |
| Tzoraki O; Dimitrova S; Barzakov M et al. | 2021 | The Impact of the COVID-19 Pandemic on the Working Conditions, Employment, Career Development and Well-Being of Refugee Researchers | Germany | HIC | December 2020 to January 2021 | International migrants | effect of lockdown measures | Cross-Sectional Study | The majority of RRs working in academia seem not to have been affected at all as far as their income is concerned, while the majority of those employed in other sectors became unemployed during the pandemic (58%). | The socioeconomic and health consequences of the COVID-19 pandemic have increased refugee researchers' (RRs) vulnerability, especially those found in asylum procedures. (...) The decrease of income, the loss of employment and the difficulty with staying productive in working from home are important concerns. Recommendations are provided (...) as well as various measures to improve RRs' integration such as online advanced language courses and provision of technological hardware and software licenses. | medium |
| Van de Velde S; Buffel V; van der Heijde C et al. | 2021 | Depressive symptoms in higher education students during the first wave of the COVID-19 pandemic. An examination of the association with various social risk factors across multiple high- and middle-income countries | 26 countries | HIC; UMIC | April 27, 2020 to July 7, 2020 | International students | mental health | Cross-Sectional Study | We find substantial cross-national variation in depressive symptoms (...). Elevated risk for depressive symptoms was found in female students, students with fewer social support resources and in a more disadvantaged socioeconomic position, and students with a migrant background. | It shows that not the pandemic itself, but rather the pandemic's secondary effects relate to students' mental health. Our results enable HEIs to be better equipped to target groups that are at risk during a pandemic. | high |
| van Ingen T; Akingbola S; Brown KA et al. | 2021 | Neighbourhood-level risk factors of COVID-19 incidence and mortality | Canada | HIC | January 23, 2020 to July 28, 2020 | International migrants | risk of infection health outcomes of disease | Ecological study | There were large inequities in COVID-19 incidence and mortality across the socio-demographic variables examined. Neighbourhoods having a higher proportion immigrants, racialized populations, large households and low socio-economic status were associated with COVID-19 risk. | Socio-demographic factors account for some of the neighbourhood-level differences in COVID-19 across Ontario. Housing characteristics account for a portion, but not all, of the excess burden of COVID-19 experienced by immigrant, racialized, low income and low education populations. | high |
| Vanthomme K; Gadeyne S; | 2021 | A population-based study on mortality among Belgian | Belgium | HIC | March 2019 to | International migrants | health outcomes of disease; | Cross-Sectional Study | Among the middle-aged, ASMRs revealed increased mortality in all origin groups, with significant excess mortality for | This study underlined important inequalities in overall and excess mortality in specific migrant communities, especially in men. | high |

| | | | | | | | | | | | |
|---|---|---|---|---|---|---|---|---|---|---|---|
| Lusyne P; Vandenhee de H. | | immigrants during the first COVID-19 wave in Belgium. Can demographic and socioeconomic indicators explain differential mortality? | | | May 2020 | | risk of infection | | Belgians and Sub-Saharan African men. At old age, excess mortality up to 60% was observed for all groups. In relative terms, most male elderly migrant groups showed higher mortality than natives, as opposed to 2019 and to women. Adding the control variables decreased this excess mortality. | Tailor-made policy measures and communication strategies should be set-up taking into account the particular risks to which groups are exposed. | |
| Varkey RS; Joy J; Sarmah G; Panda PK. | 2021 | Socioeconomic determinants of COVID-19 in Asian countries: An empirical analysis | 42 countries | HIC, UMIC, LMIC, LIC | (data until) June 18, 2020 | Internatio nal Migrants (%) | risk of transmissio n | Cross-Sectional Study | A number of factors like the net migration and per capita gross national income are found to have a positive significant relationship on the occurrence of total COVID cases in Asian countries. In alternate models, countries having high incidents of poverty are also having higher cases of COVID. In addition, voter's turnover ratio as a proxy for people's participation emerged significant and negative for daily new cases. | The countries in Asia which are more poverty ridden are also affected with higher COVID rates. (...) Creating awareness, provision of sanitary kits, and making people more participatory are important in managing COVID crises. Therefore, countries in Asia will have to follow an inclusive strategy to be well equipped to manage crises. | high |
| Vosko LF; Spring C. | 2021 | COVID-19 Outbreaks in Canada and the Crisis of Migrant Farmworkers' Social Reproduction: Transnational Labour and the Need for Greater Accountability Among Receiving States | Canada | HIC | 2020 to 2021 (not specified ) | Migrant workers | effect of lockdown measures; health outcomes of disease: SARS-CoV-2 cases | Mixed-Methods: Primarily Qualitati ve Research | Although estimates vary by source, over 1000 migrant farmworkers in Ontario tested positive for COVID-19 between April and July 2020. Thus, while Ontario documented 36594 cases by July 2020 (i.e. 250 per 100000) (…), the rate of infection among migrant farmworkers, 20,015 of whom entered Ontario during the spring and summer growing season, was approximately 4996 cases per 100000 people. (…) The magnitude of illness among migrant farmworkers in Southern Ontario, and Canada more broadly, reflects the emphasis, deeply ingrained in programs emblematic of migration management such as the SAWP, on keeping labour costs low in the interest of protecting the national food supply. | Taking Southern Ontario as its focus, this article reveals how the federal government response to COVID-19 in agriculture perpetuated the effects of longstanding laws and policies requiring migrant farmworkers, circum-scribed in their ability to politically mobilize on account of their institutionalized deportability, to shoulder disproportionate amounts of economic, social, and health risks. Centering the transnational character of migrant farmworkers' renewal, it identifies meaningful interventions to limit the structural disempowerment of migrant farmworkers and the externalization of their social reproduction. | medium |
| Wankhede P; Mukherjea S; Banuana V et al. | 2021 | The Impact of COVID-19 on India's Internal Migration (preprint) | India | LMIC | not reported | Internal migrants | mental health | Mixed-Methods | 82% of the respondents reported that the COVID-19 pandemic and the subsequent lockdowns impacted their income, resulting in an equally substantial percentage of respondents being concerned about running out of money and being unable to afford basic necessities. | Having analyzed the evidence, it is clear that not only do antiquated gender norms, stigma surrounding mental illnesses, caste and socioeconomic barriers significantly impact the quality of life led by migrant workers, but that they are only made worse by the government of India's lack of adequate policies and timely actions. | low |

| | | | | | | | | | | | |
|---|---|---|---|---|---|---|---|---|---|---|---|
| Warszawski J; Meyer L; Franck JE et al. | 2022 | Trends in social exposure to SARS-Cov-2 in France. Evidence from the national socio-epidemiological cohort – EPICOV | France | HIC | May 2020 to November 2020 | International migrants | health outcomes of disease: SARS-CoV-2 cases | Cohort Study | In May 2020, prevalence was significantly higher among immigrants born outside Europe (9.2% compared to 5.9% among second-generation immigrants from outside Europe, and 4.1% in the French-born population), but the increased risk disappeared after adjustment for living condition. In contrast, in November 2020, seroprevalence was higher in both first (13.3%) and second (14.4%) generation immigrants from outside Europe, compared to 5.3% among French-born and 6.0% among European immigrants, and they remained at higher risk even after adjustment for living conditions (adjusted odds ratio respectively: 2.1 [1.7–2.8] and 2.2 [1.8–2.9]) | The role of living conditions on the risk of SARS-CoV-2 infection decreased between the first and second epidemic waves, possibly partly due to the widespread availability of masks and virological tests at population level. Nevertheless, in November 2020, in a context of less restricted social contacts than during the first lockdown, seroprevalence remained higher among healthcare professionals than among other professionals, and strongly increased among young people and racial minorities. These populations need special attention, especially for adherence to vaccination policies. | high |
| Wiedmeyer M; Goldenberg S; Peterson S et al. | 2021 | SARS-CoV-2 testing and COVID-19 related primary care use among people with citizenship, permanent residency, and temporary immigration status in British Columbia: Cross-sectional analysis of population-based administrative data | British Columbia | HIC | January 1, 2020 to July 31, 2021 | International migrants (including international students), Migrant workers | health outcomes of disease: SARS-CoV-2 cases; health services access (risk of infection) | Cross-Sectional Study | 4.9% of people with temporary immigration status had a positive test for SARS-CoV-2 over this period, compared to 4.0% among people with permanent residency and 2.1% among people who hold Canadian citizenship. This pattern is persistent by sex/gender, age group, neighborhood income quintile, health authority, and in both metropolitan and small urban settings. At the same time we observe lower access to testing and COVID-19 related primary care among people with temporary status. | People with temporary immigration status in BC experience higher SARS-CoV-2 test positivity and lower access to testing and primary care. Interwoven immigration, health and occupational policies place people in circumstances of higher health risk and will continue to amplify harms of the COVID pandemic unless all levels of government take responsibility. Extending permanent residency status to all immigrants residing in Canada and decoupling access to health care from immigration status are policies that could reduce precarity due to temporary immigration status. | high |
| Xia Y; Ma H; Moloney G et al. | 2022 | Geographic concentration of SARS-CoV-2 cases by social determinants of health in metropolitan areas in Canada: a cross-sectional study | Canada | HIC | January 23, 2020 to February 28, 2021 | International migrants (%) | risk of infection | Ecological study | Cases were disproportionately concentrated in areas with lower income and educational attainment, and in areas with a higher proportion of visible minorities, recent immigrants, high-density housing and essential workers. Although a consistent feature across cities was concentration by the proportion of visible minorities, the magnitude of concentration by social determinant varied across cities. | Geographic concentration of SARS-CoV-2 cases was observed in all of the included cities, but the pattern by social determinants varied. Geographically prioritized allocation of resources and services should be tailored to the local drivers of inequalities in transmission in response to the resurgence of SARS-CoV-2. | medium |
| Yang B; Wu P; Lau EHY et al. | 2021 | Changing Disparities in Coronavirus Disease 2019 (COVID-19) | Hong Kong SAR, China | HIC | May 2020 to August 2020 | Migrants workers, Internatio | risk of infection | Cross-Sectional Study | For every 10% increase in the proportion of population employed as executives or professionals in a given geographic region, there was an 84% (95% confidence | Consistent findings on the reversed socioeconomic patterning of COVID-19 burden between infection waves in Hong Kong in both individual- and population- | medium |

| | | | | | | | | | | | |
|---|---|---|---|---|---|---|---|---|---|---|
| | | Burden in the Ethnically Homogeneous Population of Hong Kong Through Pandemic Waves: An Observational Study | | | | nal students | | | interval [CI], 1.97%) reduction in the incidence of COVID-19 during the third wave. In contrast, in the first 2 waves, the same was associated with 3.69 times (95% CI, 1.02-13.33) higher incidence. | level analyses indicated that risks of infections may be related to occupations involving high exposure frequency and WFH [work from home] flexibility. Contextual determinants should be taken into account in policy planning aiming at mitigating such disparities. | |
| Yee K; Peh HP; Tan YP et al. | 2021 | Stressors and coping strategies of migrant workers diagnosed with COVID-19 in Singapore: a qualitative study | Singapor e | HIC | June 21, 2020 to July 16, 2020 | Migrant workers | mental health | Qualitati ve research | Three theme categories were derived from 27 interviews: migrant worker concerns during COVID-19, coping during COVID-19 and priorities after COVID-19. Major stressors in the crisis included the inability to continue providing for their families when work is disrupted, their susceptibility to infection in crowded dormitories, the shock of receiving the COVID-19 diagnosis while asymptomatic, as well as the isolating conditions of the quarantine environment. The workers coped by keeping in contact with their families, accessing healthcare, keeping updated with the news and continuing to practise their faith and religion. They looked forward to a return to normalcy after the crisis with keeping healthy and having access to healthcare as new priorities. | We identified coping strategies employed by the workers in quarantine, many of which were made possible through the considered design of care and service delivery in mass quarantine facilities in Singapore. These can be adopted in the set-up of other mass quarantine facilities around the world to support the health and mental well-being of those quarantined. Our findings highlight the importance of targeted policy intervention for migrant workers, in areas such as housing and working environments, equitable access to healthcare, and social protection during and after this crisis. | medium |
| Yi H; Ng ST; Farwin A; Pei Ting Low A; Chang CM; Lim J. | 2021 | Health equity considerations in COVID-19: geospatialnetwork analysis of the COVID-19 outbreak in themigrant population in Singapore | Singapor e | HIC | April 2020 | Migrant workers | risk of transmissio n | Cross-Sectional Study | COVID-19 risk exposure was associated with the density of dormitory, social ties and visit areas. The migrant worker hub in the city centre was the most frequently visited for essential services of grocery shopping and remittance, followed by south central areas mainly for social gathering. The hub was positioned as the core with the highest degree of centrality with a cluster of workers exposed to COVID-19. | Our study highlights the importance of distributive justice concerning the equal allocation of essential services, including healthcare and living necessities, for low-wage dormitory-dwelling migrant workers, who are disproportionately affected by COVID-19 largely due to high-density and unhygienic built environment. | high |
| Yueping S; Hantao W; Xiao-yuan D; Zhili W. | 2021 | To Return or Stay? The Gendered Impact of the COVID-19 Pandemic on Migrant Workers in China | China | UMIC | March 2020 | Internal migrants | effect of lockdown measures | Cross-Sectional Study | We find that women migrants were less likely than men migrants to return to the cities, and less likely to return to paid work after the COVID-19 disruption. We also find that having a preschool-age child had a strong negative effect on women migrants' migration and employment | These results expand the literature on the economic vulnerabilities of Chinese migrant workers. More importantly, the findings indicate that the COVID-19 pandemic has caused a setback in the progress made in pre-pandemic times in advancing Chinese rural women’s position in the labor market. | high |

| | | | | | | | | | | | |
|---|---|---|---|---|---|---|---|---|---|---|---|
| | | | | | | | | | decisions, but had no effect on male migrants' decisions. | | |
| Yuguero O; Calahorra M; Cuevas S et al. | 2021 | Cross-sectional study of SARS-CoV-2 clinical characteristics in an immigrant population attended in a Hospital Emergency Department in the Catalunya Health Region in Spain | Spain | HIC | March 15, 2020 to Novembe r 30, 2020 | Internatio nal migrants | health outcomes of disease | Cross-Sectional Study | Most patients (72.5%) were discharged to home after evaluation in the emergency department, especially European patients. One-quarter of patients required social resources to be able to comply with quarantine measures, of whom 87% were African. Forty-seven percent of patients became infected at home and 41% in the workplace. | The immigrant population is generally younger and less infected than the population at large. In addition, the use of social resources to guarantee patient isolation has often proved essential in controlling outbreaks that have arisen in these communities. | high |
| Zambrano-Barragan P; Ramirez Hernandez S; Freier LF et al. | 2021 | The impact of COVID-19 on Venezuelan migrants' access to health: A qualitative study in Colombian and Peruvian cities | Peru, Colombi a | UMIC | July 2020 to Septemb er 2020 | Internatio nal migrants | health services access | Qualitati ve research | We found that forced migrants from Venezuela in both Colombia and Peru face common obstacles along their access trajectories to healthcare, which we summarize as legal, financial, and relating to discrimination and information asymmetry. By limiting effective access to care during the pandemic, these obstacles have also affected migrants' ability to cover the costs of basic needs, particularly food and housing. Our study also found a prevalent reliance on alternative forms of care, such as telemedicine, easy-to-access pharmacies, and extralegal care networks. | We conclude that COVID-19 has exacerbated preexisting conditions of informality and health inequities affecting Venezuelan migrants in Colombia and Peru. (…) These findings also provide guidance for specific policy recommendations. Given legal and financial obstacles, it is important to facilitate access to public health services irrespective of migratory status. Access should give special attention to COVID-19 care in the short and medium term but should focus on creating and strengthening systems of access in the long term. | medium |
| Zhang M; Gurung A; Anglewicz P; Baniya K; Yun K. | 2020 | Discrimination and Stress Among Asian Refugee Populations During the COVID-19 Pandemic: Evidence from Bhutanese and Burmese Refugees in the USA | USA | HIC | April 15, 2020 to June 1, 2020 | Refugees | mental health | Cross-Sectional Study | Among 218 refugees from 23 states, nearly one third of participants reported experiencing at least one type of discrimination, and more than two-thirds experienced at least one type of pandemic-related stress. | Reducing pandemic-related discrimination should remain a priority, as should the promotion of social support and coping strategies. (...) More importantly, policies, regulations, and education are needed to address pandemic-related stigma and discrimination. | high |
| Zhang M; Gurung A; Anglewicz P; Yun K. | 2021 | COVID-19 and Immigrant Essential Workers: Bhutanese and Burmese Refugees in the United States | USA | HIC | May 15, 2020 to June 1, 2020 | Refugees , Internatio nal migrants | risk of infection | Cross-Sectional Study | Of 218 refugees in 23 states who completed the survey from May 15 through June 1, 2020, fifteen (6.9%) reported infection with COVID-19. Being an essential worker during the pandemic (odds ratio [OR] = 5.25; 95% CI, 1.21-22.78), having an infected family member (OR = 26.92; 95% CI, 5.19-139.75), and being female (OR = 5.63; 95% CI, 1.14-27.82) were risk factors for infection. | Although we had a small snowball sample, we found that working in essential industries was associated with an increased risk of COVID-19 infection among Bhutanese and Burmese refugees. We call for larger studies that include Asian immigrant subgroups, as well as immediate attention to protecting immigrant essential workers during the COVID-19 pandemic. | high |

| | | | | | | | | | | | |
|---|---|---|---|---|---|---|---|---|---|---|---|
| | | | | | | | | | Among 33 infected family members, 23 (69.7%) were essential workers. | | |
| Zhang R; Lu Y; Du H. | 2021 | Vulnerability and resilience in the wake of COVID-19: family resources and children's well-being in China | China | HIC | February 2020 | International migrants | mental health | Cross-Sectional Study | The results highlight the complex ways in which family resources intersect with the pandemic to affect these different groups of children. Family economic resources have generally declined across all groups but left-behind children have suffered the most severe economic shock. (...) Parent–child relationships have had a more pronounced positive impact on psychological outcomes for migrant and left-behind children, who are the most deprived of parental input under normal circumstances, than for other groups of children. | The results underscore the ways in which disasters intersect with familial resources to shape child well-being and resilience in the face of adversities. The impact of COVID-19 on children and their families seems to be less disruptive than the impacts of natural disasters, which often decrease both economic and social resources. Consequently, rather than reinforcing existing patterns of inequality across different groups of children, the pandemic seems to have mitigated the developmental differences among children. | high |
| Zlot AI; Capizzi J; Bush L; Menza TW. | 2021 | Impact of COVID-19 Among Immigrant and Communities of Color Living with HIV in Oregon, 2020: Two Pandemics Rooted in Racism | USA | HIC | February 29, 2020 to November 29, 2020 | International migrants | health outcomes of disease: SARS-CoV-2 cases | Cohort Study | Almost 6% of the 2390 PLWH [people living with HIV] tested for COVID-19 had a positive COVID-19 result. PLWH with positive results tended to identify as American Indian/Alaska Native or Hispanic/ Latinx. Younger (age<50) immigrant PLWH were more than twice as likely to have a positive COVID-19 result than did older (age≥50) US-born PLWH. | This study contributes to the evidence that American Indian/Alaska Native, Latinx and younger immigrant PLWH are at greater risk for other infections, such as SARS-CoV-2. Our findings of increased risk of COVID-19 diagnosis among communities of color and immi-grants living with HIV suggest that in order to effectively create a level playing field, a multi-system, multi-layered approach that confronts and dismantles stigma and racism is needed at all levels. | high |

*: Socioeconomic Development of Country based on the World Bank country classification by income; HIC: High income countries; UMIC: Upper-middle income countries; LMIC: Lower-middle income countries; LIC: Low income countries.

### 6 Sensitivity Analysis

We examined the robustness of the models for each outcome by performing influence diagnostics, on which basis we excluded studies from the meta-analysis with the aim to reduce the variance estimate regarding the between-study heterogeneity. We examined the influence of the single studies using standardised residuals, Cook's distance, hat value, and leave-one-out cross validations (LOO-CV) for measuring the impact on the estimated effect size $\hat{\theta}$ and the heterogeneity measure $\tau^2$.[8] Models for each outcome were fitted in three steps:

- model 1) including all studies;
- model 2) excluding studies with considerable outlying values for diagnostic statistics (see above) as well as LOO-CVs for $\hat{\theta}$ and $\tau^2$;
- model 3) further exclusion of studies with considerable influence on LOO-CVs for $\hat{\theta}$ and $\tau^2$.

We compared the results of each model for changes in $\tau^2$ while evaluating the models for both effect sizes – risk ratio (RR) and risk difference (RD) – and selected the respective model with comparably low *and* robust value of $\tau^2$.

For each outcome and selected model, we additionally inspected asymmetry patterns for assessing potential publication bias. Therefore, we used shaded funnel plots with contours for $p < \{0{\cdot}1,\ 0{\cdot}05, 0{\cdot}01\}$, which allows distinguishing publication bias from other sources of asymmetry (e.g. functional differences in studies),[8] and compared study's effect estimate versus study's precision (i.e. inverse standard error: $1/SE$). Using precision puts emphasis on the larger studies, while smaller studies are scattered at the bottom. The 95% confidence region is indicated by two curved lines, which do not follow a funnel shape consequently.[9] Additionally to the visual assessment of asymmetry using shaded funnel plots, we performed Peters' Regression Test for evaluating asymmetry of binary outcomes. We used Peters' Test to avoid inflated risk of false positives. The weighted regression test is based on the inverse of total sample size as predictor and the inverse variance of average event probability as weight, and follows a t-distribution with $n - 2$ degree of freedom.[8,10]

### 6.1 Cases

In model 2, studies from Mema et al. and Harkness et al. were excluded due to outlying hat values (Fig. S1). The study from Indseth et al. was also excluded due to high direct impact on $\hat{\theta}$ and $\tau^2$ (Fig. S2). The model configurations improved $\tau^2$ (Table S7) for both effect sizes RR and RD. In model 3, we further excluded studies from Erfani et al., Vosko et al., and Lombardi et al., because of outlying standardised residuals (Fig. S1) for effect size RR.[11–13] The exclusion of studies improved $\tau^2$ for the RR model. $\tau^2$ for RD changed marginally (Table S7). Due to stable estimates of $\tau^2$, for both RR and RD, model 3 was selected for presenting results.

**Figure S1: Cases. Model diagnostics - Standardised residuals, Cook's Distance, Hat value**

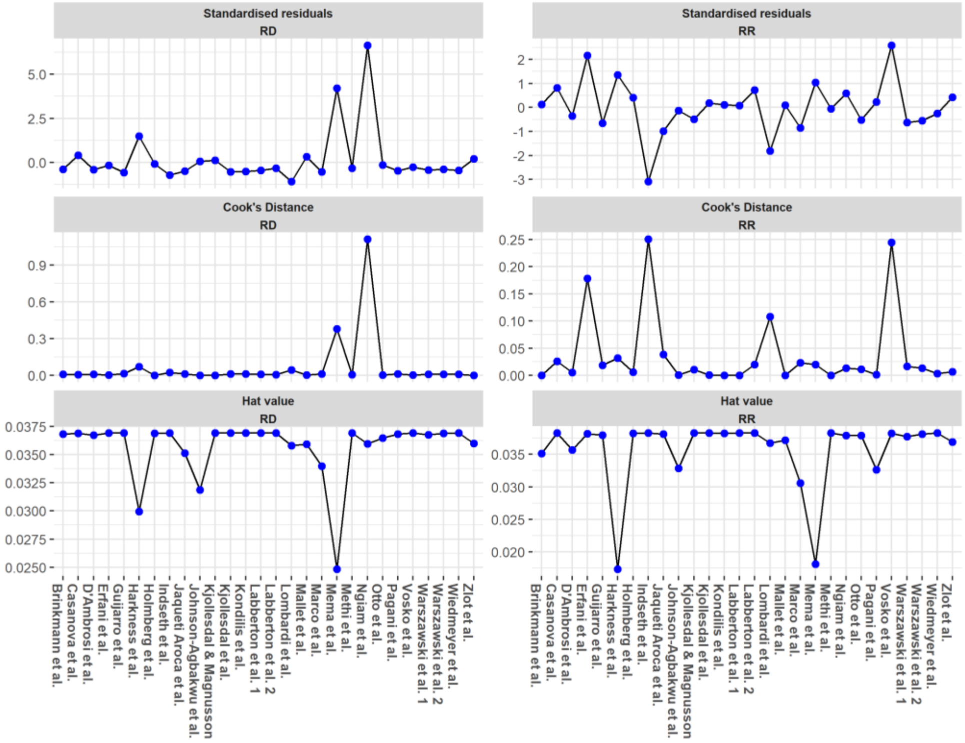

### Figure S2: Cases. Model diagnostics – LOO-CV

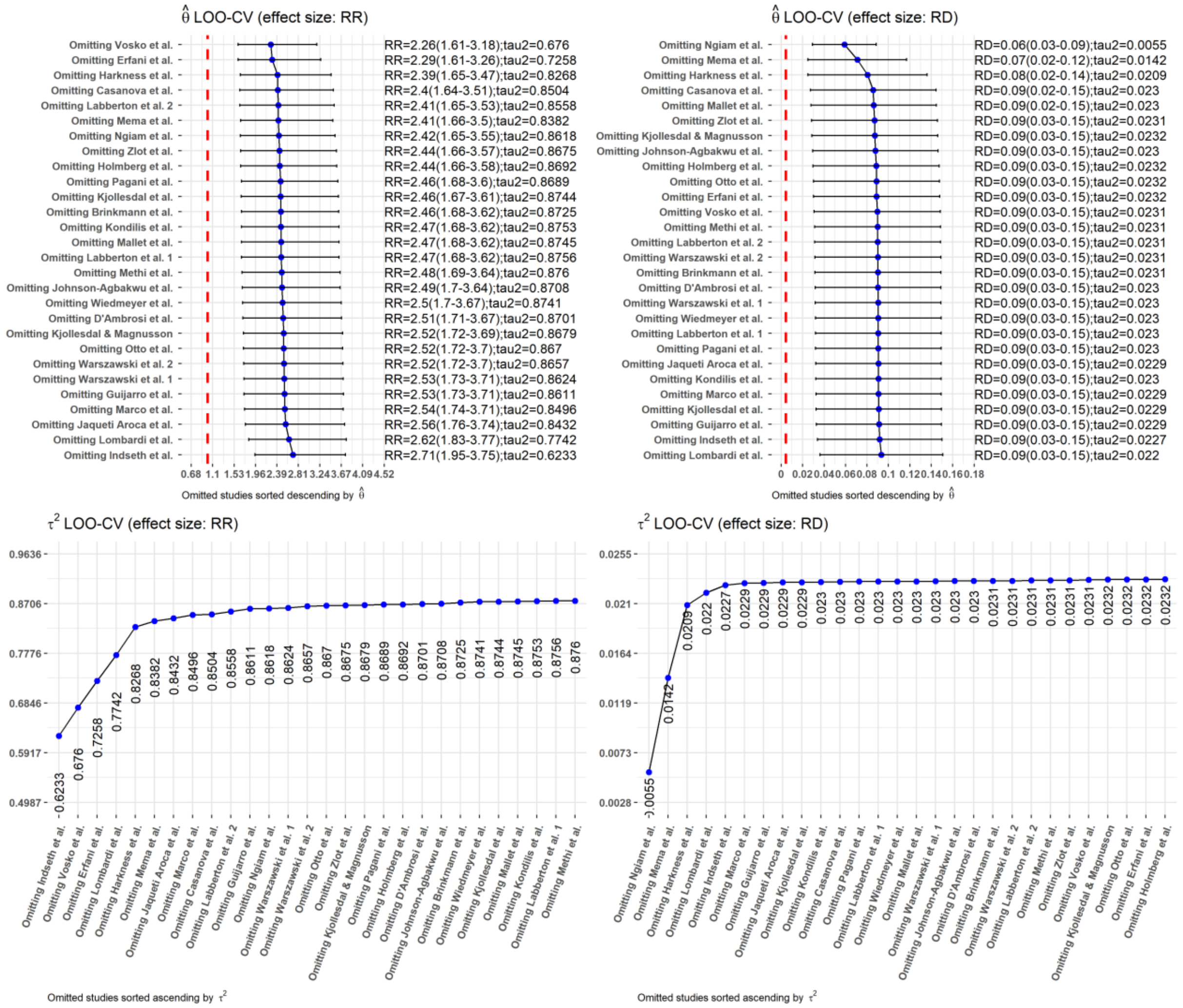

### Table S7: Cases. Modelling results

| Model-ID | Excluded studies | Risk Ratio (RR) | | | Risk Difference (RD) | | |
|---|---|---|---|---|---|---|---|
| | | $\hat{\theta}$ (95% *CI*) | $\tau^2$ (95% *CI*) | $\chi^2$ (*p*) | $\hat{\theta}$ (95% *CI*) | $\tau^2$ (95% *CI*) | $\chi^2$ (*p*) |
| 1 | | 2·47 (1·71-3·58) | 0·8402 (0·5073-1·6253) | 37646·64 (p = 0) | 0·09 (0·03-0·15) | 0·0220 (0·0148-0·0524) | 33422·01 (p = 0) |
| 2 | Mema et al. (North America), Harkness et al. (North America), Indseth et al. (North Europe) | 2·58 (1·85-3·59) | 0·6134 (0·3614-1·1815) | 19800·31 (p = 0) | 0·07 (0·02-0·11) | 0·0133 (0·0080-0·0267) | 14876·99 (p = 0) |
| 3 | Mema et al. (North America), Harkness et al. (North America), Indseth et al. (North Europe), Erfani et al. (North America), Vosko et al. (North America), Lombardi et al. (South Europe) | 2·33 (1·88-2·89) | 0·2056 (0·1116-0·4216) | 13606·90 (p = 0) | 0·07 (0·02-0·13) | 0·0144 (0·0084-0·0303) | 13733·18 (p = 0) |

The funnel plot for model 3 (Fig. 3) shows the relation of effects sizes (RR or RD) and precision of studies ($1/SE$) between migrants and non-migrants for Covid-19 cases. Smaller studies are scattered at the bottom due to low precision, and studies with larger sample size are scattered within the 0.01 significance region. Peter's Regression Test indicated no funnel plot asymmetry, i.e. based on the underlying data, publication bias is expected to be unlikely for this outcome (RR: t = 0·10, p = 0·9248; RD: t = 0·20, p = 0·8454).

**Figure S3: Cases. Funnel plot**

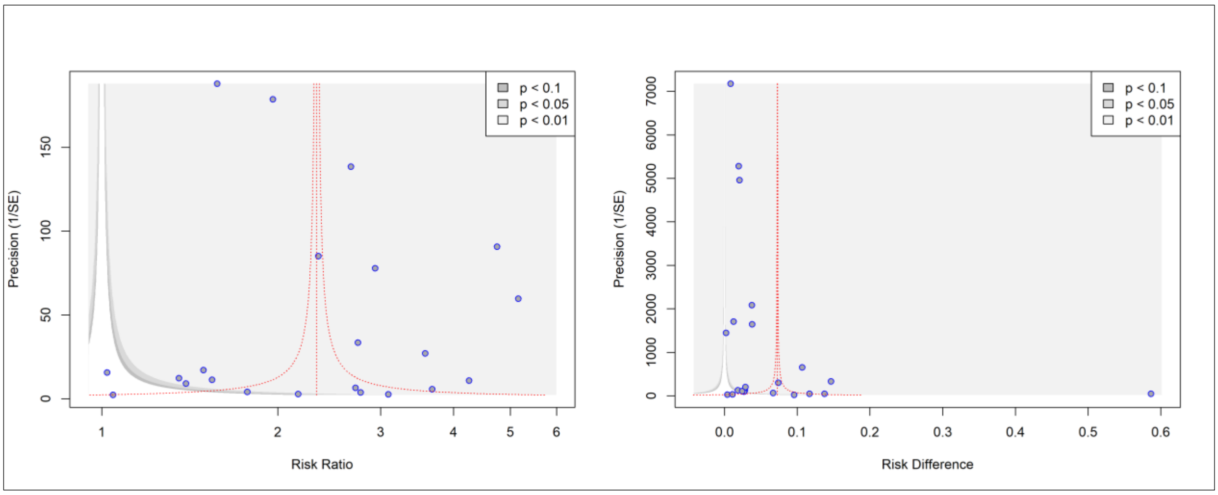

**Peter's Regression test of funnel plot asymmetry**

| | **RR** | | **RD** | |
|---|---|---|---|---|
| **Test results** | t, df | p-value | t; df | p-value |
| | *t = -0·10*<br>*df = 20* | *0·9248* | *t = 0·20*<br>*df = 20* | *0·8454* |
| **Sample test** | bias (se) | Intercept (se) | bias (se) | Intercept (se) |
| | *-338·8431 (3543·7363)* | *0·9763 (0·1132)* | *72·4952 (366·9675)* | *0·0891 (0·0117)* |

### 6.2 Hospitalisations

In model 2, studies from Dressler et al. and Engjom et al. were excluded due to a considerable impact on $\tau^2$ as well as high values of standardised residuals and Cook's Distance. The exclusion of mentioned studies improved $\tau^2$ (Table S8). In model 3, we further excluded studies from Ingraham et al. and Rossi et al. because of outlying values found in standardised residuals and Cook's Distance (Ingraham et al.)[14] as well as high direct impact on $\hat{\theta}$ and $\tau^2$ (Rossi et al.)[15] for the effect size risk difference (Fig. S4 and Fig. S5). The further exclusion of studies improved $\tau^2$ again, especially for the effect size risk difference (Table S8). Due to stable estimate of $\tau^2$ for both RR and RD, model 3 was selected for presenting the results.

**Figure S4: Hospitalisations. Model diagnostics - Standardised residuals, Cook's Distance, Hat value**

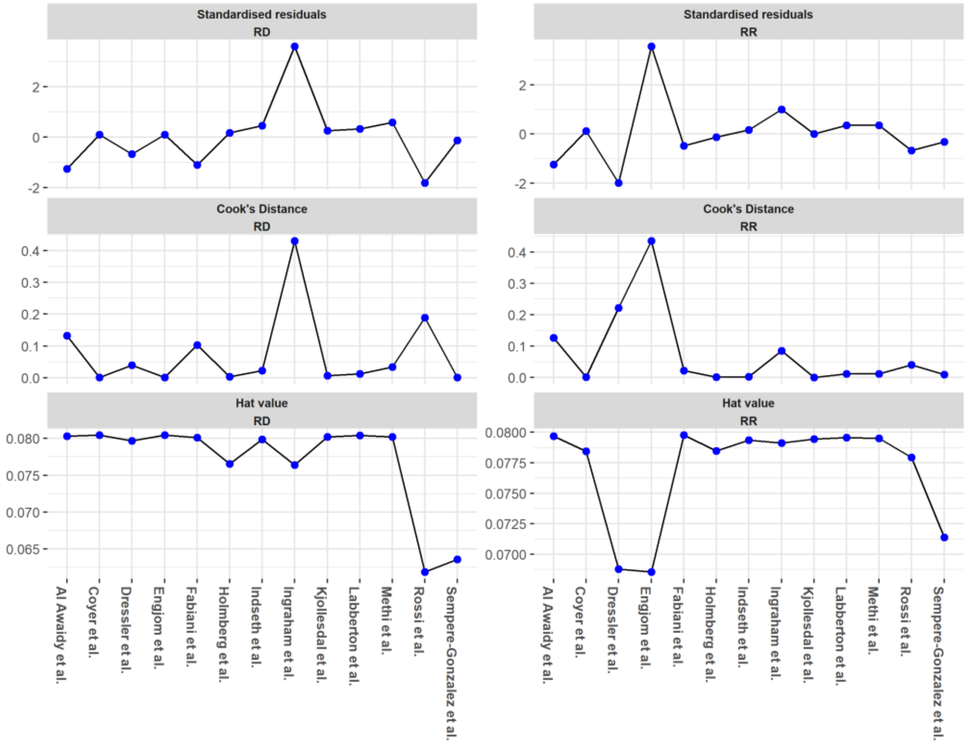

### Figure S5: Hospitalisations. Model diagnostics – LOO-CV

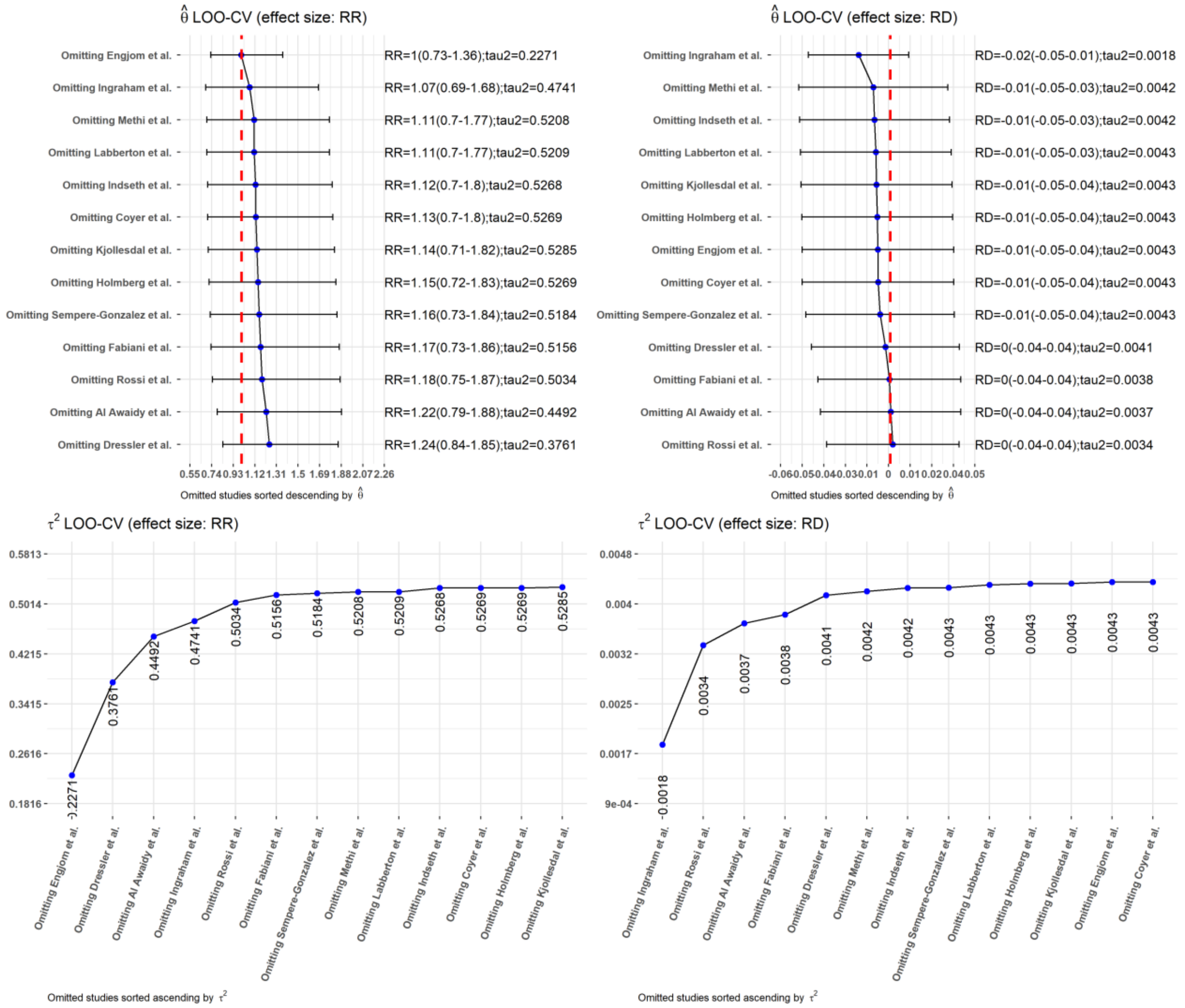

### Table S8: Hospitalisations. Modelling results

| Model-ID | Excluded studies | Risk Ratio (RR) | | | Risk Difference (RD) | | |
|---|---|---|---|---|---|---|---|
| | | $\hat{\theta}$ (95% $CI$) | $\tau^2$ (95% $CI$) | $\chi^2$ ($p$) | $\hat{\theta}$ (95% $CI$) | $\tau^2$ (95% $CI$) | $\chi^2$ ($p$) |
| 1 | | 1·14 (0·74-1·74) | 0·4748 (0·2390-1·5068) | 1356·94 (p < 0·01) | -0·01 (-0·05-0·03) | 0·0039 (0·0019-0·0117) | 1805·01 (p = 0) |
| 2 | Dressler et al. (North Europe), Engjom et al. (North Europe) | 1·09 (0·82-1·43) | 0·1584 (0·0737-0·4833) | 1282·14 (p < 0·01) | 0 (-0·05-0·04) | 0·0046 (0·0021-0·0155) | 1730·53 (p = 0) |
| 3 | Dressler et al. (North Europe), Engjom et al. (North Europe), Ingraham et al. (North America, Rossi et al. (South Europe) | 1·05 (0·8-1·37) | 0·1155 (0·0494-0·4094) | 1048·42 (p < 0·01) | -0·01 (-0·04-0·02) | 0·0017 (0·0008-0·0058) | 1607·91 (p = 0) |

The funnel plot for model 3 (Fig. S6) shows the relation of effects size (RR or RD) and precision of studies $(1/SE)$ between migrants and non-migrants for Covid-19 hospitalisations. Smaller studies are scattered at the bottom due to lower precision (in particular observable for risk difference), and studies with larger sample size are predominantly scattered within the 0·01 or 0·05 significance levels. Potential asymmetry patterns can be observed in the funnel plot. However, Peter's Regression Test resulted in no asymmetry, which indicates that publication bias can be expected to be unlikely for this outcome (RR: t = 0·30, p = 0·7712; RD: t = 0·71, p = 0·5006). However, as the number of studies is lower than recommended $n \geq 10$ (i.e. $n = 9$ studies included),[16] the test may have not detected real asymmetry due to lower statistical power. Consequently, it cannot completely ruled out if publication bias or other sources (e.g. functional differences between studies) are responsible for potential observed asymmetry irrespective of the test results.

**Figure S6: Hospitalisations. Funnel plot**

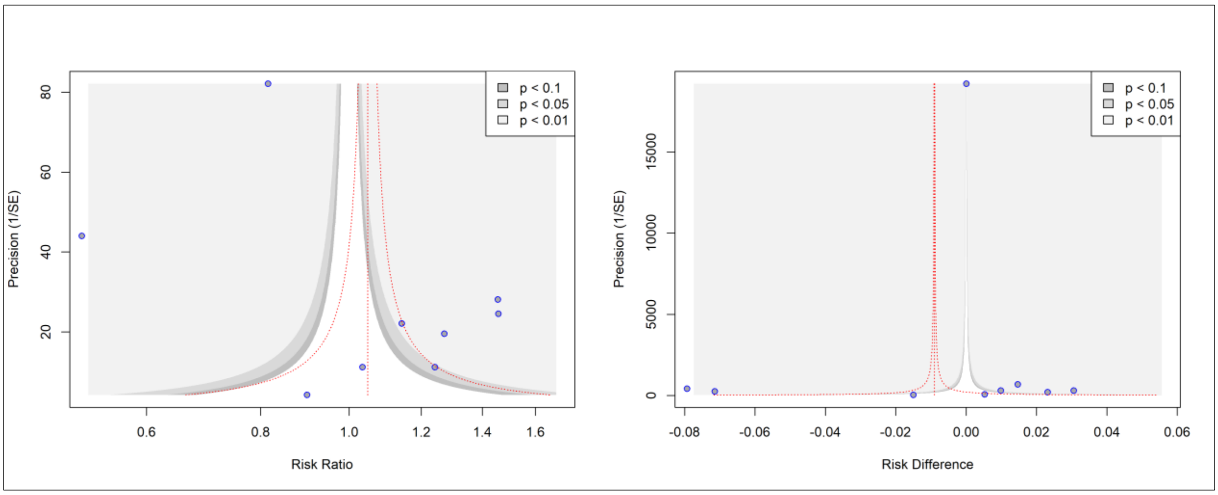

**Peter's Regression test of funnel plot asymmetry**

| | **RR** | | **RD** | |
|---|---|---|---|---|
| **Test results** | t, df | p-value | t; df | p-value |
| | *t = 0·30*<br>*df = 7* | *0·7712* | *t = 0·71*<br>*df = 7* | *0·5006* |
| **Sample test** | bias (se) | Intercept (se) | bias (se) | Intercept (se) |
| | *492·0052*<br>*(1627·6697)* | *-0·1987*<br>*(0·0943)* | *143·4257*<br>*(201·9862)* | *-0·0619*<br>*(0·0117)* |

#### 6.3 ICU admissions

In model 2, the study from Ngiam et al. was excluded due to outlying values in both effect sizes (RR and RD) for standardised residuals, Cook's Distance, as well as a considerable impact on $\hat{\theta}$ and $\tau^2$ (Fig. S7 and Fig. S8). The exclusion of the mentioned study improved $\tau^2$ (Table S9). In model 3, we further excluded the study from Martin-Sanchez et al. because of high direct impact on $\tau^2$ for the effect size RR (Fig. S8).[17] The exclusion of this study further improved $\tau^2$ (Table S9). Due to stable estimate of $\tau^2$ for both RR and RD, model 3 was selected for presenting the results.

**Figure S7: ICU admissions. Model diagnostics – Standardised residuals, Cook's Distance, Hat value**

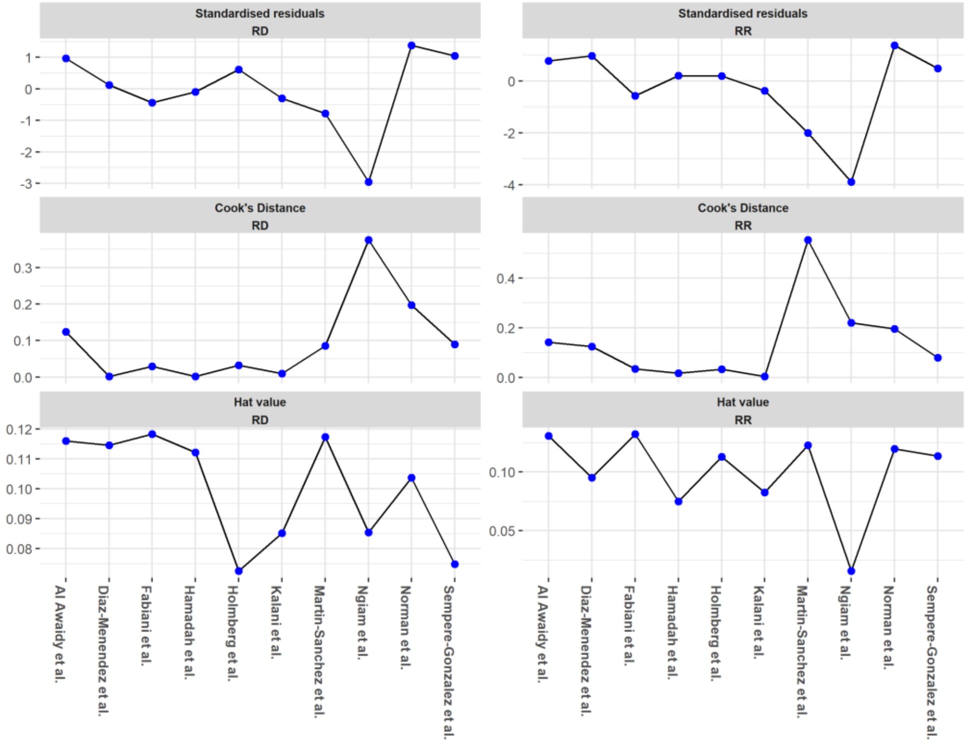

### Figure S8: ICU admissions. Model diagnostics – LOO-CV

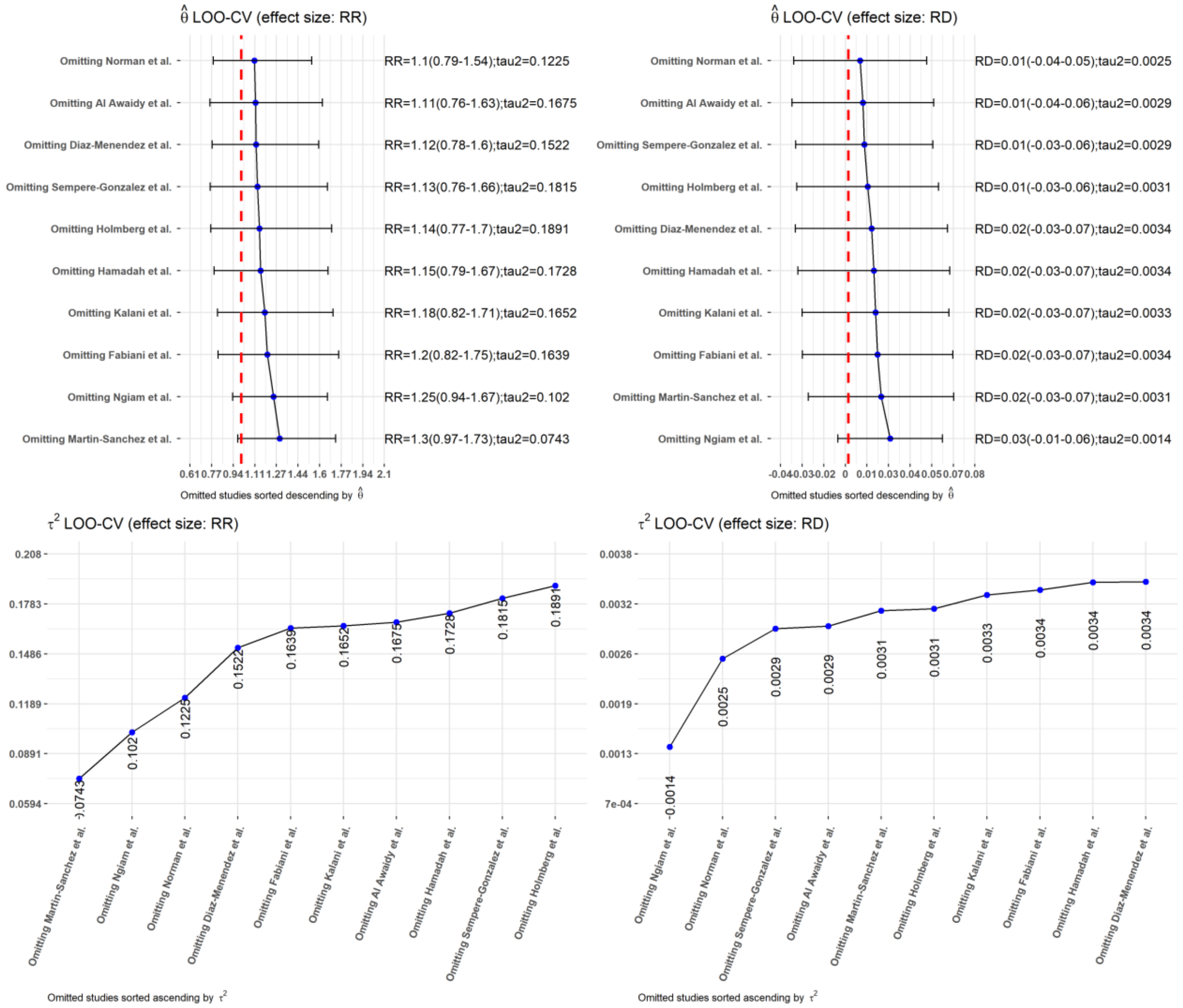

### Table S9: ICU admissions. Modelling results

| Model-ID | Excluded studies | Risk Ratio (RR) | | | Risk Difference (RD) | | |
|---|---|---|---|---|---|---|---|
| | | $\hat{\theta}$ (95% $CI$) | $\tau^2$ (95% $CI$) | $\chi^2$ ($p$) | $\hat{\theta}$ (95% $CI$) | $\tau^2$ (95% $CI$) | $\chi^2$ ($p$) |
| 1 | | 1·17 (0·85-1·62) | 0·1412 (0·1662-5·4130) | 114·80 (p < 0·01) | 0·02 (-0·03-0·06) | 0·0029 (0·0011-0·0128) | 114·27 (p < 0·01) |
| 2 | Ngiam et al. (Asia) | 1·25 (0·94-1·67) | 0·1020 (0·0329-0·3813) | 98·69 (p < 0·01) | 0·03 (-0·01-0·06) | 0·0014 (0·0004-0·0059) | 99·06 (p < 0·01) |
| 3 | Ngiam et al. (Asia), Martin-Sanchez et al. (South Europe) | 1·36 (1·04-1·78) | 0·0570 (0·0142-0·2486) | 77·22 (p < 0·01) | 0·04 (0-0·07) | 0·0011 (0·0003-0·0056) | 73·12 (p < 0·01) |

The funnel plot for model 3 (Fig. S9) shows the relation of effects size (RR or RD) and precision of studies ($1/SE$) between migrants and non-migrants for Covid-19 related ICU admissions. Smaller studies are scattered at the bottom due to lower precision, and studies with larger sample size are dispersed to the top of the graph. Potential asymmetry patterns can be observed in the funnel plot. However, Peter's Regression Test confirmed a symmetric pattern, which indicates that publication bias can be expected to be unlikely for this outcome (RR: t = 1·05, p = 0·3344; RD: t = 1·29, p = 0·2431). However, as the number of studies is lower than recommended $n \geq 10$ (i.e. $n = 8$ studies included),[16] the test may have not detected real asymmetry due to lower statistical power. Consequently, it cannot completely ruled out if publication bias or other sources (e.g. functional differences between studies) are responsible for potential observed asymmetry irrespective of the negative test results.

**Figure S9: ICU admissions. Funnel plot**

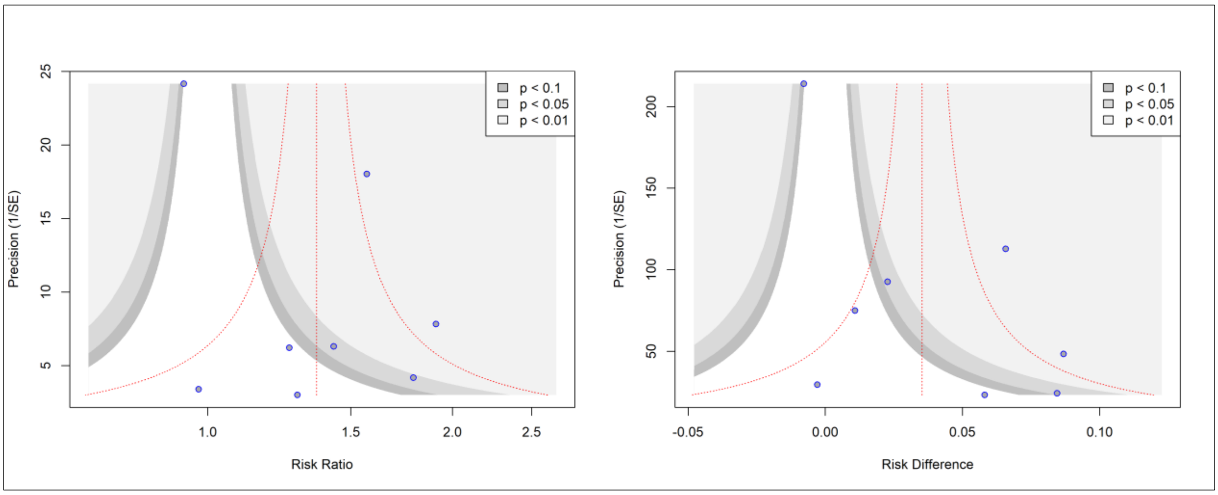

**Peter's Regression test of funnel plot asymmetry**

| | **RR** | | **RD** | |
|---|---|---|---|---|
| **Test results** | t, df | p-value | t; df | p-value |
| | *t = 1·05*<br>*df = 6* | *0·3344* | *t = 1·29*<br>*df = 6* | *0·2431* |
| **Sample test** | bias (se) | Intercept (se) | bias (se) | Intercept (se) |
| | *270·5329 (257·7745)* | *-0·0059 (0·0767)* | *45·4454 (35·1111)* | *0·0002 (0·0104)* |

#### 6.4 Mortality

For the outcome mortality, we conducted separate meta-analyses by different denominators for mortality, i.e. studies reporting on all deaths, and hospital associated deaths, respectively.

##### 6.4.1 Denominator: all deaths

The diagnostic statistics showed that individual studies did not take a considerable high impact either on $\hat{\theta}$ or $\tau^2$ (Fig. S11), as well as did not indicate extreme outliers (Fig. S10). Furthermore, due to a small number of studies reporting on denominator *all deaths* ($n = 6$), statistical power is expected to be comparably low. Therefore, no study was excluded from the model.

**Figure S10: Mortality – all deaths. Model diagnostics – Standardised residuals, Cook's Distance, Hat value**

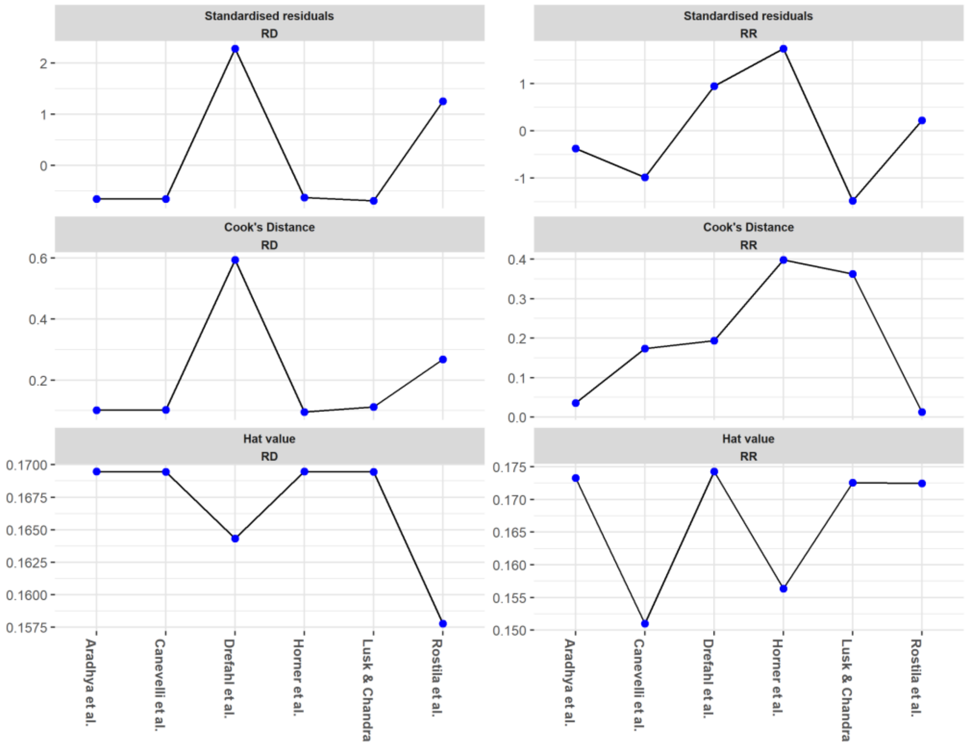

**Figure S11: Mortality – all deaths. Model diagnostics – LOO-CV**

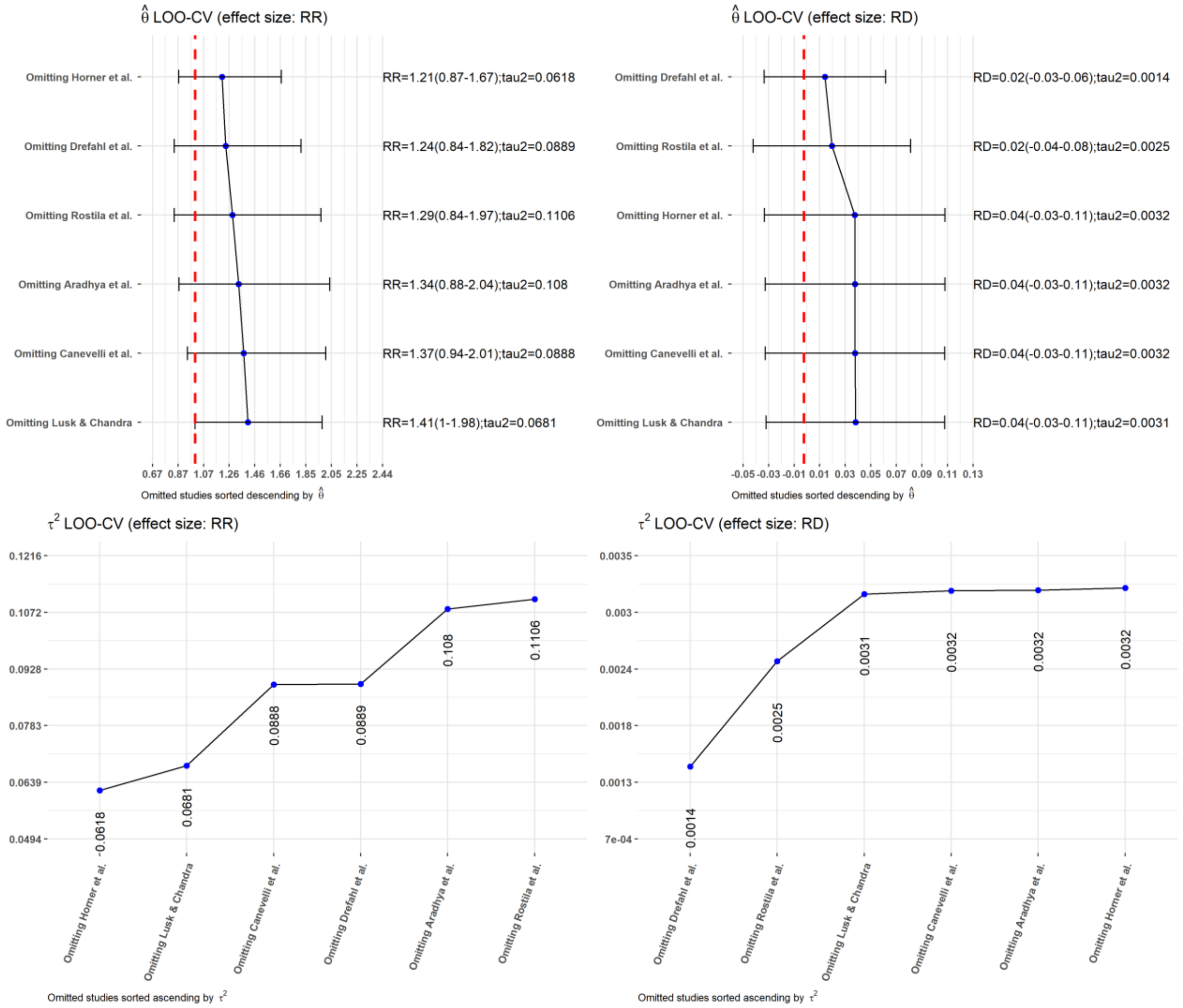

The funnel plot for model 3 (Fig. S12) shows the relation of effects size (RR or RD) and precision of studies ($1/SE$) between migrants and non-migrants for Covid-19 associated deaths based on denominator *all deaths*. As the number of studies is lower than recommended $n \geq 10$ (i.e. $n = 6$ studies included), which indicates lower statistical power,[16] plotted estimates and Peter's regression test may have not detected real asymmetry. Consequently, publication bias or other sources of asymmetry (e.g. functional differences in studies) cannot be excluded for the mortality denominator *all deaths*.

**Figure S12: Mortality – all deaths. Funnel plot**

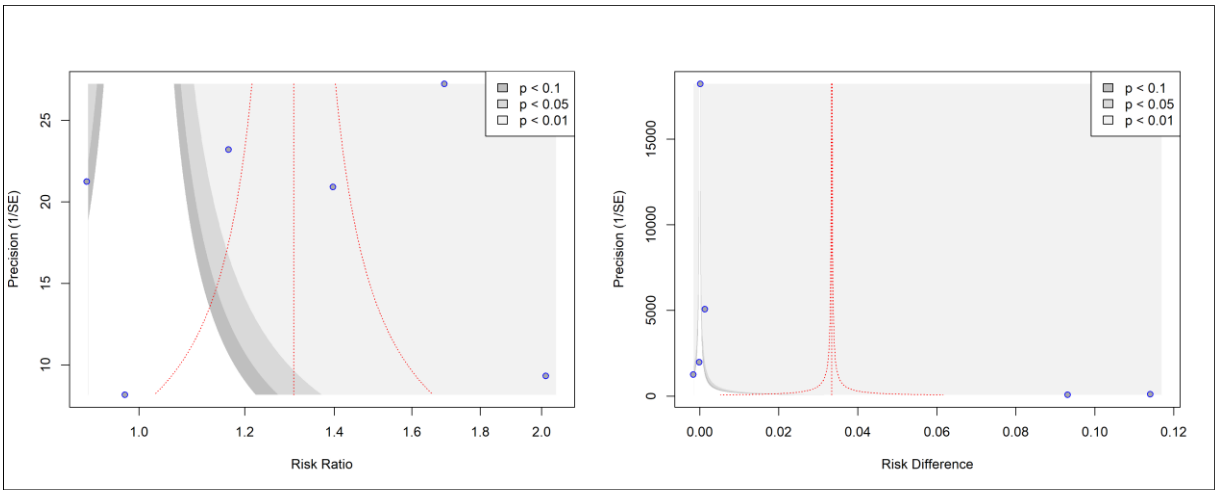

**Peter's Regression test of funnel plot asymmetry**

| | **RR** | | **RD** | |
|---|---|---|---|---|
| **Test results** | t, df | p-value | t; df | p-value |
| | *t = 1·46*<br>*df = 4* | *0·2171* | *t = 2·56*<br>*df = 4* | *0·0627* |
| **Sample test** | bias (se) | Intercept (se) | bias (se) | Intercept (se) |
| | *3294·8019 (2250·9047)* | *0·0061 (0·0949)* | *788·6064 (308·0857)* | *0·0036 (0·0130)* |

#### 6.4.2 Denominator: hospitalised cases

In model 2, the study from Al Awaidy was excluded because of high values in standardised residuals (for both effect sizes RD and RR) and Cook's Distance (for effect size RR), as well as considerable direct impact on $\tau^2$.[18] The exclusion of the study improved $\tau^2$ (Table S10). In model 3, we further excluded the model from Fabiani et al. due to high direct impact on $\tau^2$ and $\hat{\theta}$ in both effect sizes.[19] The exclusion of this study further improved $\tau^2$ (Table S10). Due to stable estimate of $\tau^2$ for both RR and RD, model 3 was selected for presenting the results.

**Figure S13: Mortality – hospitalised cases. Model diagnostics – Standardised residuals, Cook's Distance, Hat value**

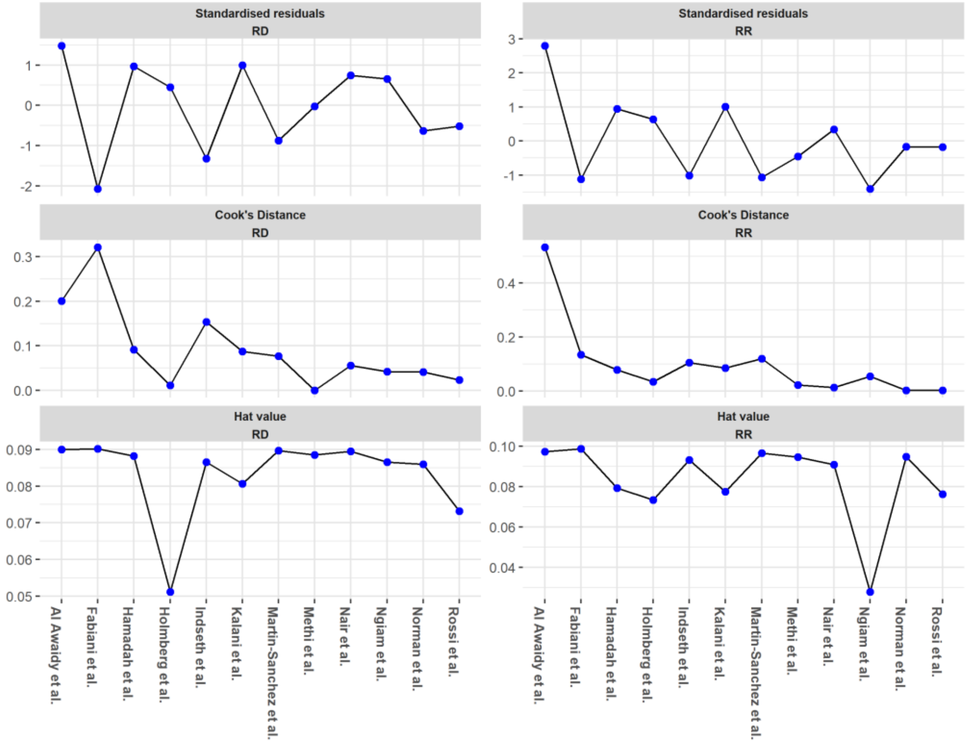

### Figure S14: Mortality – hospitalised cases. Model diagnostics – LOO-CV

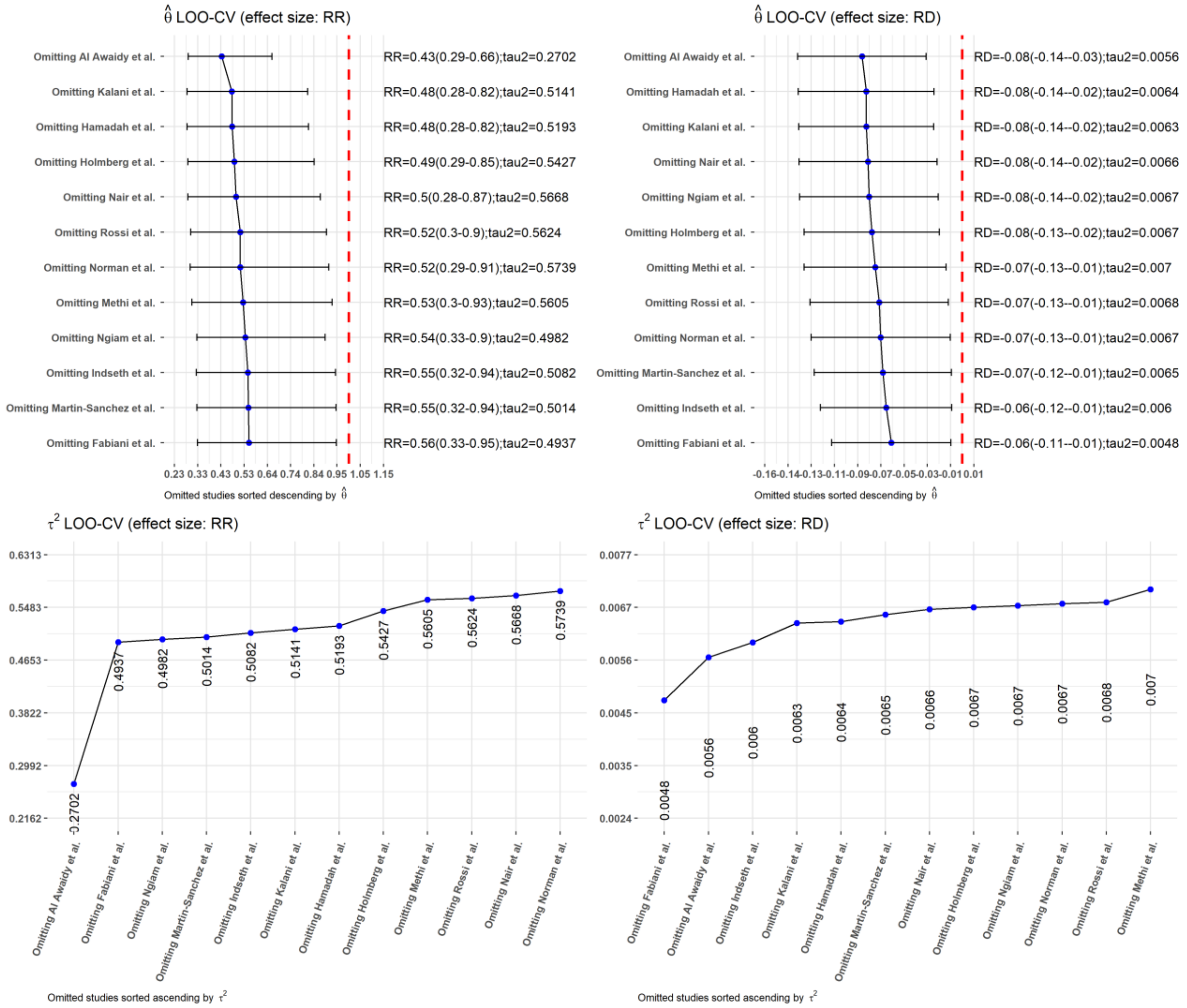

### Table S10: Mortality – hospitalised cases. Modelling results

| Model-ID | Excluded studies | Risk Ratio (RR) | | | Risk Difference (RD) | | |
|---|---|---|---|---|---|---|---|
| | | $\hat{\theta}$ (95% CI) | $\tau^2$ (95% CI) | $\chi^2$ (p) | $\hat{\theta}$ (95% CI) | $\tau^2$ (95% CI) | $\chi^2$ (p) |
| 1 | | 0·51 (0·31-0·85) | 0·5120 (0·2244-1·8593) | 386·58 (p < 0·01) | -0·07 (-0·13; -0·02) | 0·0064 (0·0029-0·0180) | 1631·70 (p = 0) |
| 2 | Al Awaidy et al. (Asia) | 0·43 (0·29-0·66) | 0·2702 (0·0994-1·4203) | 73·28 (p < 0·01) | -0·08 (-0·14; -0·03) | 0·0056 (0·0025-0·0174) | 794·97 (p < 0.01) |
| 3 | Al Awaidy et al. (Asia), Fabiani et al. (South Europe) | 0·47 (0·3-0·73) | 0·2527 (0·0823-1·6095) | 47·59 (p < 0·01) | -0·07 (-0·12; -0·02) | 0·0041 [0·0017-0·0141) | 244·63 (p < 0.01) |

The funnel plot for model 3 (Fig. S12) shows the relation of effects size (RR or RD) and precision of studies $(1/SE)$ between migrants and non-migrants for Covid-19 associated deaths based on denominator *hospitalised cases*. Studies predominantly scattered symmetrically in the funnel plot, which is also confirmed by Peter's Regression test indicating no asymmetry (RR: t = 1·83, p = 0·1054; RD: t = 1·30, p = 0·2305), i.e. publication bias is expected to be low for this outcome.

**Figure S15: Mortality – hospitalised cases. Funnel plot**

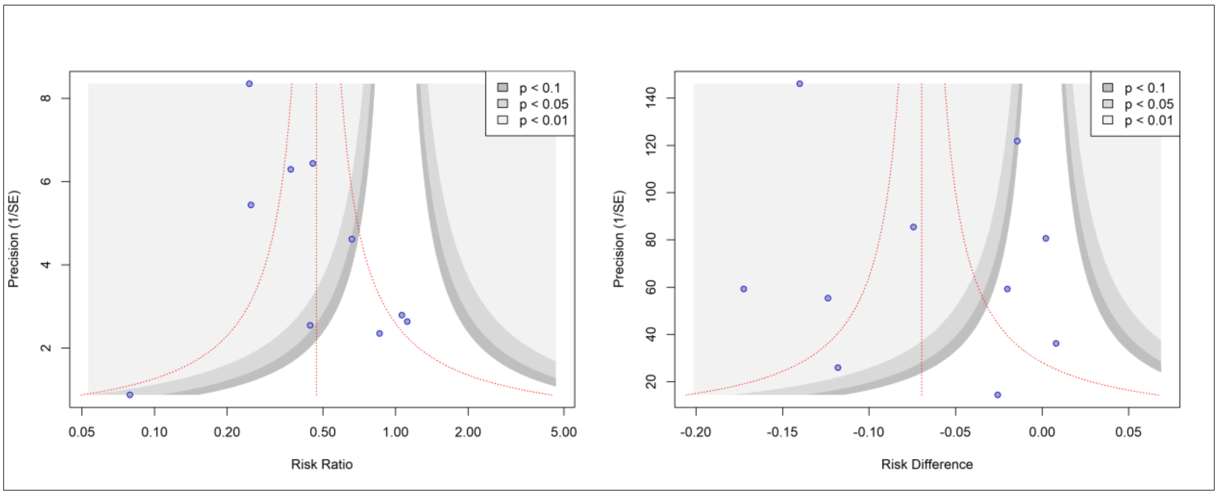

**Peter's Regression test of funnel plot asymmetry**

| | **RR** | | **RD** | |
|---|---|---|---|---|
| **Test results** | t, df | p-value | t; df | p-value |
| | *t = 1·83*<br>*df = 8* | *0·1054* | *t = 1·30*<br>*df = 8* | *0·2305* |
| **Sample test** | bias (se) | Intercept (se) | bias (se) | Intercept (se) |
| | *289·1607*<br>*(158·4411)* | *-1·2630*<br>*(0·1252)* | *22·9915*<br>*(17·7133)* | *-0·1339*<br>*(0·0140)* |

### 7 Subgroup analysis by migrant indicators for SARS-CoV-2 Cases

Figure S16 shows the subgroup analysis by migrant indicators for SARS-CoV-2 cases.

**Figure S16: SARS-CoV-2 Cases. Subgroup analysis by migrant indicator**

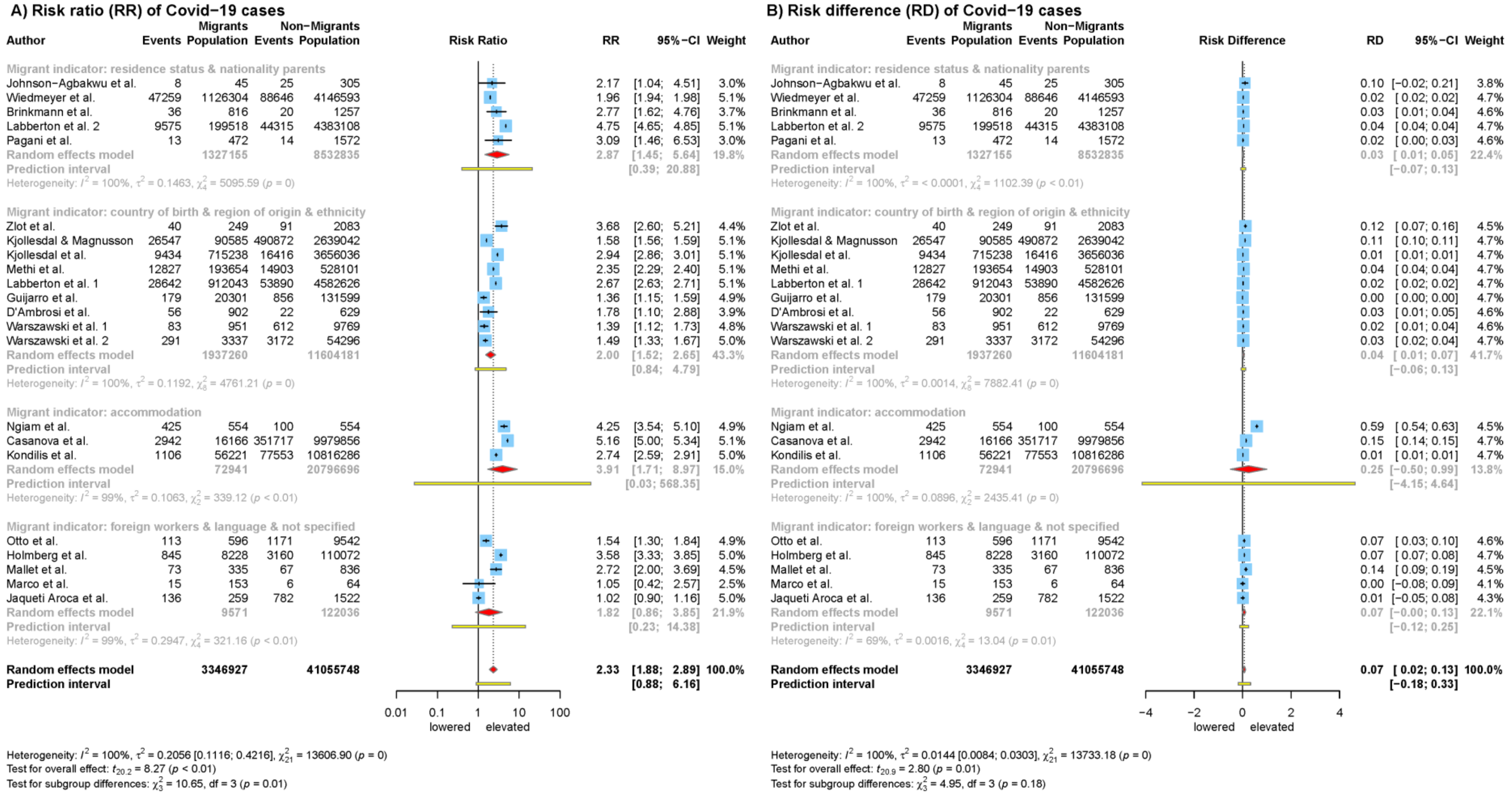

Note: "Ethnicity" refers to region of origin, the wording is taken the authors' formulation.[20]

### 8 PRISMA Checklist

Table S11a shows the PRISMA-Checklist for Abstracts.[21] Table S11b contains the PRISMA-Checklist for the main Article.[21]

#### Table S11a: PRISMA-Checklist for Abstracts

| Section and Topic | Item # | Checklist item | Reported (Yes/No) |
|---|---|---|---|
| **TITLE** | | | |
| Title | 1 | Identify the report as a systematic review. | yes |
| **BACKGROUND** | | | |
| Objectives | 2 | Provide an explicit statement of the main objective(s) or question(s) the review addresses. | yes |
| **METHODS** | | | |
| Eligibility criteria | 3 | Specify the inclusion and exclusion criteria for the review. | yes |
| Information sources | 4 | Specify the information sources (e.g. databases, registers) used to identify studies and the date when each was last searched. | yes |
| Risk of bias | 5 | Specify the methods used to assess risk of bias in the included studies. | no |
| Synthesis of results | 6 | Specify the methods used to present and synthesise results. | yes |
| **RESULTS** | | | |
| Included studies | 7 | Give the total number of included studies and participants and summarise relevant characteristics of studies. | yes |
| Synthesis of results | 8 | Present results for main outcomes, preferably indicating the number of included studies and participants for each. If meta-analysis was done, report the summary estimate and confidence/credible interval. If comparing groups, indicate the direction of the effect (i.e. which group is favoured). | yes |
| **DISCUSSION** | | | |
| Limitations of evidence | 9 | Provide a brief summary of the limitations of the evidence included in the review (e.g. study risk of bias, inconsistency and imprecision). | no |
| Interpretation | 10 | Provide a general interpretation of the results and important implications. | yes |
| **OTHER** | | | |
| Funding | 11 | Specify the primary source of funding for the review. | yes |
| Registration | 12 | Provide the register name and registration number. | yes |

#### Table S11b: PRISMA-Checklist

| Section and Topic | Item # | Checklist item | Location where item is reported |
|---|---|---|---|
| **TITLE** | | | |
| Title | 1 | Identify the report as a systematic review. | p.1 (manuscript) |
| **ABSTRACT** | | | |
| Abstract | 2 | See the PRISMA 2020 for Abstracts checklist. | p.119 (supplement) |
| **INTRODUCTION** | | | |
| Rationale | 3 | Describe the rationale for the review in the context of existing knowledge. | p. 4 (manuscript) |
| Objectives | 4 | Provide an explicit statement of the objective(s) or question(s) the review addresses. | p.4 (manuscript) |

| Section and Topic | Item # | Checklist item | Location where item is reported |
|---|---|---|---|
| **METHODS** | | | |
| Eligibility criteria | 5 | Specify the inclusion and exclusion criteria for the review and how studies were grouped for the syntheses. | p.4+5 (manuscript) |
| Information sources | 6 | Specify all databases, registers, websites, organisations, reference lists and other sources searched or consulted to identify studies. Specify the date when each source was last searched or consulted. | p. 4 (manuscript) |
| Search strategy | 7 | Present the full search strategies for all databases, registers and websites, including any filters and limits used. | Table S1 & S2 (supplement) |
| Selection process | 8 | Specify the methods used to decide whether a study met the inclusion criteria of the review, including how many reviewers screened each record and each report retrieved, whether they worked independently, and if applicable, details of automation tools used in the process. | p.4 (manuscript) |
| Data collection process | 9 | Specify the methods used to collect data from reports, including how many reviewers collected data from each report, whether they worked independently, any processes for obtaining or confirming data from study investigators, and if applicable, details of automation tools used in the process. | p. 4+5 (manuscript) |
| Data items | 10a | List and define all outcomes for which data were sought. Specify whether all results that were compatible with each outcome domain in each study were sought (e.g. for all measures, time points, analyses), and if not, the methods used to decide which results to collect. | p. 4+5 (manuscript) |
| | 10b | List and define all other variables for which data were sought (e.g. participant and intervention characteristics, funding sources). Describe any assumptions made about any missing or unclear information. | n.a. |
| Study risk of bias assessment | 11 | Specify the methods used to assess risk of bias in the included studies, including details of the tool(s) used, how many reviewers assessed each study and whether they worked independently, and if applicable, details of automation tools used in the process. | p. 5 (manuscript) |
| Effect measures | 12 | Specify for each outcome the effect measure(s) (e.g. risk ratio, mean difference) used in the synthesis or presentation of results. | p. 5 (manuscript) |
| Synthesis methods | 13a | Describe the processes used to decide which studies were eligible for each synthesis (e.g. tabulating the study intervention characteristics and comparing against the planned groups for each synthesis (item #5)). | p. 5 (manuscript); chapter 6 (supplement) |
| | 13b | Describe any methods required to prepare the data for presentation or synthesis, such as handling of missing summary statistics, or data conversions. | n.a. |
| | 13c | Describe any methods used to tabulate or visually display results of individual studies and syntheses. | p.5 & Panel 2 (manuscript) + Chapter 6 (Supplement) |
| | 13d | Describe any methods used to synthesize results and provide a rationale for the choice(s). If meta-analysis was performed, describe the model(s), method(s) to identify the presence and extent of statistical heterogeneity, and software package(s) used. | Panel 2 (manuscript)+ Chapter 6 (Supplement) |
| | 13e | Describe any methods used to explore possible causes of heterogeneity among study results (e.g. subgroup analysis, meta-regression). | Panel 2 (manuscript)+ Chapter 6 (Supplement) |
| | 13f | Describe any sensitivity analyses conducted to assess robustness of the synthesized results. | Chapter 6 (Supplement) |
| Reporting bias assessment | 14 | Describe any methods used to assess risk of bias due to missing results in a synthesis (arising from reporting biases). | n.a |
| Certainty assessment | 15 | Describe any methods used to assess certainty (or confidence) in the body of evidence for an outcome. | n.a. |
| **RESULTS** | | | |
| Study selection | 16a | Describe the results of the search and selection process, from the number of records identified in the search to the number of studies included in the review, ideally using a flow diagram. | Figure 1 (p.4) + p. 6 (manuscript) |
| | 16b | Cite studies that might appear to meet the inclusion criteria, but which were excluded, and explain why they were excluded. | n.a. |
| Study characteristics | 17 | Cite each included study and present its characteristics.Table S3 | Table S6 p.35 (Supplement) |
| Risk of bias in studies | 18 | Present assessments of risk of bias for each included study. | p. 20 ff (supplement) |

| Section and Topic | Item # | Checklist item | Location where item is reported |
|---|---|---|---|
| Results of individual studies | 19 | For all outcomes, present, for each study: (a) summary statistics for each group (where appropriate) and (b) an effect estimate and its precision (e.g. confidence/credible interval), ideally using structured tables or plots. | p. 6+7 (manuscript) |
| Results of syntheses | 20a | For each synthesis, briefly summarise the characteristics and risk of bias among contributing studies. | Table 1a & b (manuscript) |
| | 20b | Present results of all statistical syntheses conducted. If meta-analysis was done, present for each the summary estimate and its precision (e.g. confidence/credible interval) and measures of statistical heterogeneity. If comparing groups, describe the direction of the effect. | Fig 2a – Fig. 5 |
| | 20c | Present results of all investigations of possible causes of heterogeneity among study results. | n.a. |
| | 20d | Present results of all sensitivity analyses conducted to assess the robustness of the synthesized results. | Chapter 6 (Supplement) |
| Reporting biases | 21 | Present assessments of risk of bias due to missing results (arising from reporting biases) for each synthesis assessed. | n.a. |
| Certainty of evidence | 22 | Present assessments of certainty (or confidence) in the body of evidence for each outcome assessed. | p. 6+7 (manuscript) |
| **DISCUSSION** | | | |
| Discussion | 23a | Provide a general interpretation of the results in the context of other evidence. | p.9 (manuscript) |
| | 23b | Discuss any limitations of the evidence included in the review. | p. 9f (manuscript) |
| | 23c | Discuss any limitations of the review processes used. | p. 10 (manuscript) |
| | 23d | Discuss implications of the results for practice, policy, and future research. | p. 10 (manuscript) |
| **OTHER INFORMATION** | | | |
| Registration and protocol | 24a | Provide registration information for the review, including register name and registration number, or state that the review was not registered. | p. 4 (manuscript) |
| | 24b | Indicate where the review protocol can be accessed, or state that a protocol was not prepared. | P .4 (manuscript) |
| | 24c | Describe and explain any amendments to information provided at registration or in the protocol. | p. 4 (manuscript) |
| Support | 25 | Describe sources of financial or non-financial support for the review, and the role of the funders or sponsors in the review. | p. 5 (manuscript) |
| Competing interests | 26 | Declare any competing interests of review authors. | p. 11 (manuscript) |
| Availability of data, code and other materials | 27 | Report which of the following are publicly available and where they can be found: template data collection forms; data extracted from included studies; data used for all analyses; analytic code; any other materials used in the review. | p. 11 (manuscript) |
